# Molnupiravir or nirmatrelvir-ritonavir versus usual care in patients admitted to hospital with COVID-19 (RECOVERY): a randomised, controlled, open-label, platform trial

**DOI:** 10.1101/2024.05.23.24307731

**Authors:** RECOVERY Collaborative Group, Peter W Horby, Natalie Staplin, Leon Peto, Jonathan R Emberson, Mark Campbell, Guilherme Pessoa-Amorim, Buddha Basnyat, Louise Thwaites, Rogier Van Doorn, Raph L Hamers, Jeremy Nel, John Amuasi, Richard Stewart, Dipansu Ghosh, Fergus Hamilton, Purav Desai, Nicholas Easom, Jaydip Majumdar, Paul Hine, David Chadwick, Graham Cooke, Sara Sharp, Hanif Esmail, J Kenneth Baillie, Maya H Buch, Saul N Faust, Thomas Jaki, Katie Jeffery, Edmund Juszczak, Marian Knight, Wei Shen Lim, Alan Montgomery, Aparna Mukherjee, Andrew Mumford, Kathryn Rowan, Guy Thwaites, Marion Mafham, Richard Haynes, Martin J Landray

**Author notes:** The writing committee and trial steering committee are listed at the end of this manuscript and a complete list of collaborators in the Randomised Evaluation of COVID-19 Therapy (RECOVERY) trial is provided in the Supplementary Appendix. Correspondence to: Prof Peter W Horby and Prof Martin J Landray, RECOVERY Central Coordinating Office, Richard Doll Building, Old Road Campus, Roosevelt Drive, Oxford OX3 7LF, United Kingdom.

## Abstract

**Background:** Molnupiravir and nirmatrelvir-ritonavir (Paxlovid) are oral antivirals that have been proposed as treatments for patients admitted to hospital with COVID-19.

**Methods:** In this randomised, controlled, open-label, adaptive platform trial, several potential treatments for patients hospitalised with COVID-19 pneumonia were evaluated. Molnupiravir and nirmatrelvir-ritonavir were assessed in separate comparisons in RECOVERY, both of which are reported here. Eligible and consenting adults could join the molnupiravir comparison, the nirmatrelvir-ritonavir comparison, or both. For each comparison, participants were randomly allocated in a 1:1 ratio to the relevant antiviral (five days of molnupiravir 800mg twice daily or nirmatrelvir-ritonavir 300mg/100mg twice daily) or to usual care without the relevant antiviral drug, using web-based unstratified randomisation with allocation concealment. The primary outcome was 28-day mortality, and secondary outcomes were time to discharge alive from hospital, and among those not on invasive ventilation at baseline, progression to invasive ventilation or death. Analysis was by intention-to-treat. Both comparisons were stopped by the investigators because of low recruitment. ISRCTN (50189673) and clinicaltrials.gov (NCT04381936).

**Findings:** From 24 January 2022 to 24 May 2023, 923 patients were recruited to the molnupiravir comparison (445 allocated molnupiravir and 478 allocated usual care), and from 31 March 2022 to 24 May 2023, 137 patients were recruited to the nirmatrelvir-ritonavir comparison (68 allocated nirmatrelvir-ritonavir and 69 allocated usual care). More than three-quarters of the patients in both comparisons were vaccinated and had anti-spike antibodies at randomisation, and more than two-thirds were receiving other SARS-CoV-2 antivirals (including remdesivir or sotrovimab). In the molnupiravir comparison, 74 (17%) patients allocated to molnupiravir and 79 (17%) patients allocated usual care died within 28 days (hazard ratio [HR] 0.93; 95% confidence interval [CI] 0.68-1.28; p=0.66). In the nirmatrelvir-ritonavir comparison, 13 (19%) patients allocated nirmatrelvir-ritonavir and 13 (19%) patients allocated usual care died within 28 days (HR 1.02; 95% CI 0.47-2.23; p=0.96). In neither comparison was there evidence of a significant difference in the duration of hospitalisation or the proportion of patients progressing to invasive ventilation or death.

**Interpretation:** In adults hospitalised with COVID-19, neither molnupiravir nor nirmatrelvir-ritonavir were associated with reductions in 28-day mortality, duration of hospital stay, or risk of progressing to invasive mechanical ventilation or death although these comparisons had limited statistical power due to low recruitment.

**Funding:** UK Research and Innovation (Medical Research Council) and National Institute of Health and Care Research (Grant ref: MC_PC_19056), and Wellcome Trust (Grant Ref: 222406/Z/20/Z).

**Trial registration:** ClinicalTrials.gov NCT04381936 https://clinicaltrials.gov/ct2/show/NCT04381936

ISRCTN50189673 http://www.isrctn.com/ISRCTN50189673

## INTRODUCTION

Early antiviral treatment of unvaccinated patients at high risk of severe COVID-19 can substantially reduce the risk of subsequent hospitalisation or death.^1–3^ There is less evidence supporting antiviral treatment in people admitted to hospital, and in these patients it may be that immune-mediated lung damage, rather than ongoing viral replication, is primarily responsible for disease progression. Antiviral treatment with neutralising monoclonal antibodies (nMAb) has been shown to substantially reduce mortality in hospitalised patients, but only in those not yet producing their own anti-SARS-CoV-2 antibodies.^4^ However, most immunocompetent adults now have some SARS-COV-2 immunity following vaccination or previous infection, and the available nMAbs are now largely ineffective because of spike gene mutations in globally prevalent SARS-COV-2 variants.^5, 6^ Remdesivir, a nucleoside analogue inhibitor of the viral RNA-dependent RNA polymerase, reduces time-to-discharge by around one day in hospitalised patients and is associated with a moderate reduction in mortality, at least in non-ventilated patients.^7, 8^ Other potent SARS-CoV-2 antivirals, including molnupiravir and nirmatrelvir-ritonavir (Paxlovid), have not been adequately tested in randomised trials in hospitalised patients, and it could be that these drugs, given alone or in combination with other antivirals, would improve clinical outcomes.

Molnupiravir is an orally absorbed prodrug of N(4)-Hydroxycytidine, a nucleoside-analogue substrate of the viral RNA-dependent RNA polymerase. It has a broad spectrum of activity against RNA viruses, including coronaviruses, and a high barrier to the development of viral resistance.^9–11^ Its mechanism of action is distinct to remdesivir, impairing viral RNA replication by facilitating ambiguous base pairing, leading to an accumulation of transversion mutations. In the MOVe-OUT trial, early treatment of high-risk unvaccinated patients with COVID-19 reduced the risk of hospitalisation or death by 30% (risk ratio[RR] 0.70; 95% CI 0.49-0.99; p=0.045), but no significant benefit was shown in the subsequent PANORAMIC trial among lower-risk, vaccinated patients infected with Omicron variants (RR 1.07; 95% CI 0.81-1.41; p=0.5).^12, 13^ MOVe-IN is the only reported trial of molnupiravir in hospitalised patients, which included 304 unvaccinated individuals.^14^ This found no significant difference in the primary outcome of recovery by day 29 (84% molnupiravir group vs. 85% placebo group), or mortality (6% molnupiravir group vs. 3% placebo group), but was underpowered to rule out worthwhile improvements in either outcome.

Nirmatrelvir is an orally administered small-molecule inhibitor of the viral 3-chymotrypsin-like (3CL) protease, which is co-administered with ritonavir to enhance its pharmacokinetics.^15^ In the EPIC-HR trial of high-risk unvaccinated patients with early COVID-19 it reduced the risk of hospitalisation or death by 88% (RR 0.12; 95% CI 0.06-0.25; p<0.0001) although no significant benefit was present in the subsequent EPIC-SR trial of vaccinated and lower risk patients (RR 0.48; 95% CI 0.17-1.41; p=0.18).^3, 16^ Only one trial has reported nirmatrelvir-ritonavir use in hospitalised patients, which included 264 patients.^17^ In this trial there was no significant difference in the primary outcome of 28-day mortality (4% nirmatrelvir-ritonavir group vs. 6% standard treatment group), but it was underpowered to rule out a worthwhile benefit of treatment.

Here we report the results of independent evaluations of molnupiravir and nirmatrelvir-ritonavir versus usual care in the RECOVERY trial, a randomised, open-label platform trial evaluating treatments for patients hospitalised with COVID-19 pneumonia.

## METHODS

### Study design and participants

The Randomised Evaluation of COVID-19 therapy (RECOVERY) trial is an investigator-initiated, individually randomised, controlled, open-label, adaptive platform trial to evaluate the effects of potential treatments in patients hospitalised with COVID-19. Details of the trial design and results for other treatments have been published previously (dexamethasone, hydroxychloroquine, lopinavir-ritonavir, azithromycin, tocilizumab, convalescent plasma, colchicine, aspirin, casirivimab plus imdevimab, baricitinib, empagliflozin, dimethyl fumarate, and high-dose corticosteroids in hypoxic patients not requiring ventilatory support).^4, 18–29^ The trial was conducted at hospital organisations in the United Kingdom supported by the National Institute for Health and Care Research Clinical Research Network, as well as in South and Southeast Asia and Africa. Of these, 75 hospitals in the UK, 2 in Nepal, and 2 in Indonesia enrolled participants in the molnupiravir comparison, and 32 UK hospitals enrolled participants in the nirmatrelvir-ritonavir comparison (appendix pp 2-31). The trial is coordinated by the Nuffield Department of Population Health at University of Oxford (Oxford, UK), the trial sponsor. The trial is conducted in accordance with the principles of the International Conference on Harmonisation–Good Clinical Practice guidelines and approved by the UK Medicines and Healthcare products Regulatory Agency (MHRA) and the Cambridge East Research Ethics Committee (ref: 20/EE/0101). The protocol, statistical analysis plan, and additional information are available on the study website www.recoverytrial.net.

Patients admitted to hospital were eligible for the study if they had confirmed SARS-CoV-2 infection with a pneumonia syndrome thought to be related to COVID-19, and no medical history that might, in the opinion of the managing physician, put the patient at significant risk if they were to participate in the trial. Patients were excluded from the molnupiravir comparison if (i) they were pregnant or breastfeeding, or (ii) they had received molnupiravir during their current illness. Patients were excluded from the nirmatrelvir-ritonavir comparison if (i) they were in the first trimester of pregnancy, (ii) had severe liver impairment (Child-Pugh class C), (iii) had severe renal impairment (eGFR <30ml/min/1.73m^2^), (iv) had received nirmatrelvir-ritonavir during their current illness, or (v) were receiving a concomitant medication with CYP3A4 dependent metabolism that risked a severe drug-drug interaction with nirmatrelvir-ritonavir. Children (age <18 years) and those unable to take medication orally were excluded from both comparisons. If a study treatment was unavailable, or if the managing physician considered a study treatment to be either definitely indicated or definitely contraindicated, then patients were excluded from the relevant comparison. Written informed consent was obtained from all patients, or a legal representative if patients were too unwell or otherwise unable to provide informed consent.

### Randomisation and masking

Baseline data were collected using a web-based case report form that included demographics, level of respiratory support, major comorbidities, suitability of the study treatment for a particular patient, SARS-CoV-2 vaccination status, and study treatment availability at the study site (appendix pp 43-46). A serum sample and nose swab were collected at randomisation from UK patients and sent to central laboratories for testing. Serum was tested for anti-SARS-CoV-2 spike antibodies, anti-SARS-CoV-2 nucleocapsid antibodies, and SARS-CoV-2 nucleocapsid antigen using Roche Elecsys assays (Roche Diagnostics, Basel, Switzerland). Patients were classified as positive or negative for anti-spike and anti-nucleocapsid antibodies using manufacturer defined thresholds, and as positive or negative for serum nucleocapsid antigen using the study population median value (as this assay had not previously been validated on serum samples). Nose swabs were tested for SARS-CoV-2 RNA using TaqPath COVID-19 RT-PCR (Thermo Fisher Scientific, Massachusetts, US). Samples with sufficient concentration of viral RNA were sequenced using the ONT Midnight protocol (Oxford Nanopore Technologies, Oxford, UK).^30^ Sequence data were used to detect mutations associated with resistance to molnupiravir or nirmatrelvir-ritonavir identified from literature searches. Further details of laboratory analyses are in the appendix (pp 32-33).

Patients could enter either one or both of the comparisons provided they were eligible. For each comparison they entered, patients were randomly assigned in a 1:1 ratio to either usual standard of care plus the relevant treatment or usual standard of care without the relevant treatment, using web-based simple (unstratified) randomisation with allocation concealed until after randomisation (appendix pp 41-43). Patients allocated to molnupiravir were to receive 800mg orally twice daily for 5 days. Patients allocated to nirmatrelvir-ritonavir were to receive 300mg/100mg orally twice daily for 5 days, reduced to 150mg/100mg twice daily if they had moderate renal impairment (eGFR 30-59ml/min/1.73m^2^). In both comparisons the course was to be continued after discharge if required.

As a platform trial, and in a factorial design, patients could be simultaneously randomised to other concurrently evaluated treatment groups: (i) empagliflozin versus usual care, (ii) higher-dose corticosteroids versus usual care, (iii) sotrovimab versus usual care (appendix pp 41-42). Participants and local study staff were not masked to allocated treatment. Other than members of the Data Monitoring Committee, all individuals involved in the trial were masked to aggregated outcome data while recruitment and 28-day follow-up were ongoing.

### Procedures

Follow-up nose swabs were collected from UK patients on day 3 and day 5 (counting the day of randomisation as day 1). These were analysed in the same manner as the baseline swab described above.

A single online follow-up form was completed when participants were discharged, had died or at 28 days after randomisation, whichever occurred earliest (appendix pp 47-55). Information was recorded on adherence to allocated study treatment, receipt of other COVID-19 treatments, duration of admission, receipt of respiratory or renal support, and vital status (including cause of death). In addition, in the UK, routine healthcare and registry data were obtained, including information on vital status (with date and cause of death), discharge from hospital, receipt of respiratory support, or renal replacement therapy. For sites outside the UK a further case report form (appendix pp 56-57) collected vital status at day 28 (if not already reported on the initial follow-up form).

### Outcomes

Outcomes were assessed at 28 days after randomisation, with further analyses specified at 6 months. The primary outcome was all-cause mortality at 28 days. Secondary outcomes were time to discharge from hospital, and, among patients not on invasive mechanical ventilation at randomisation, invasive mechanical ventilation (including extra-corporal membrane oxygenation) or death. Prespecified subsidiary clinical outcomes were use of non-invasive respiratory support, time to successful cessation of invasive mechanical ventilation (defined as cessation of invasive mechanical ventilation within, and survival to, 28 days), use of renal dialysis or haemofiltration, cause-specific mortality, bleeding events, thrombotic events, major cardiac arrhythmias, non-SARS-CoV-2 infections, and metabolic complications (including ketoacidosis). Virological outcomes were viral RNA levels in nose swabs taken at day 3 and day 5, and the frequency of detection of resistance markers. Information on suspected serious adverse reactions was collected in an expedited fashion to comply with regulatory requirements.

### Sample size

The intention for this comparison was to continue recruitment until sufficient primary outcomes had accrued to have 90% power to detect a proportional risk reduction of 20% at a two-sided significance level of 0.01.

Following the initial wave of Omicron infection in the UK in early 2022, the number of patients hospitalised with COVID-19 pneumonia reduced substantially in the UK, as did recruitment to both comparisons. Because of persistently low recruitment, the RECOVERY Trial Steering Committee decided to close both comparisons on 24^th^ May 2023 whilst still blinded to the results.

### Statistical Analysis

The primary analysis for all outcomes was by intention-to-treat, comparing patients randomised to the study treatment with patients randomised to usual care but for whom that study treatment was both available and suitable as a treatment. For the primary outcome of 28-day mortality, the hazard ratio from an age- and respiratory status-adjusted Cox model was used to estimate the mortality rate ratio. We constructed Kaplan-Meier survival curves to display cumulative mortality over the 28-day period. We used the same Cox regression method to analyse time to hospital discharge and successful cessation of invasive mechanical ventilation, with patients who died in hospital right-censored on day 29. Median time to discharge was derived from Kaplan-Meier estimates. For the pre-specified composite secondary outcome of progression to invasive mechanical ventilation or death within 28 days (among those not receiving invasive mechanical ventilation at randomisation), and the subsidiary clinical outcomes of receipt of invasive or non-invasive ventilation, or use of haemodialysis or haemofiltration, the precise dates were not available and so a log-binomial regression model was used to estimate the risk ratio adjusted for age and respiratory status. SARS-CoV-2 viral RNA levels in nose-swabs were estimated with analysis of covariance (ANCOVA) using the log transformed values after adjustment for each participant’s baseline value, age and level of respiratory support at randomisation.

Prespecified subgroup analyses were performed for the primary outcome using the statistical test of interaction (test for heterogeneity or trend), in accordance with the prespecified analysis plan, defined by the following characteristics at randomisation: age, sex, ethnicity, level of respiratory support, days since symptom onset, and use of corticosteroids (appendix p 135). Exploratory sub-group analyses were also performed by SARS-COV-2 antibody status (anti-S and anti-N), serum nucleocapsid antigen status, and use of other antivirals.

Estimates of rate and risk ratios are shown with 95% confidence intervals. All p-values are 2-sided and are shown without adjustment for multiple testing. The full database is held by the study team, which collected the data from study sites and performed the analyses at the Nuffield Department of Population Health, University of Oxford (Oxford, UK).

Analyses were performed using SAS version 9.4 and R version 3.4. The trial is registered with ISRCTN (50189673) and clinicaltrials.gov (NCT04381936).

## RESULTS

### Molnupiravir comparison

Between 24 January 2022 and 24 May 2023, 923/1242 (74%) patients enrolled in RECOVERY at sites participating in the molnupiravir comparison were eligible to be randomly allocated to molnupiravir, of whom 445 were allocated molnupiravir and 478 were allocated usual care without molnupiravir (figure 1A). The 319 RECOVERY patients not included in the molnupiravir comparison had similar characteristics to those included (webtable 1). The mean age of study participants in this comparison was 71.4 years (SD 14.1), 767 (83%) had received a COVID-19 vaccine, and the median time since symptom onset was 5 days (IQR 3 to 9 days). 133/923 (14%) patients in the molnupiravir comparison simultaneously participated in the nirmatrelvir-ritonavir comparison. At randomisation, 809 (88%) patients were receiving corticosteroids, and 629 (68%) were receiving, or allocated to receive, a SARS-CoV-2 antiviral other than molnupiravir (including usual care remdesivir, and sotrovimab or nirmatrelvir-ritonavir allocated in another RECOVERY comparison). 227 (25%) patients were anti-N seropositive and 705 (76%) were anti-S seropositive.

**Figure 1A:**
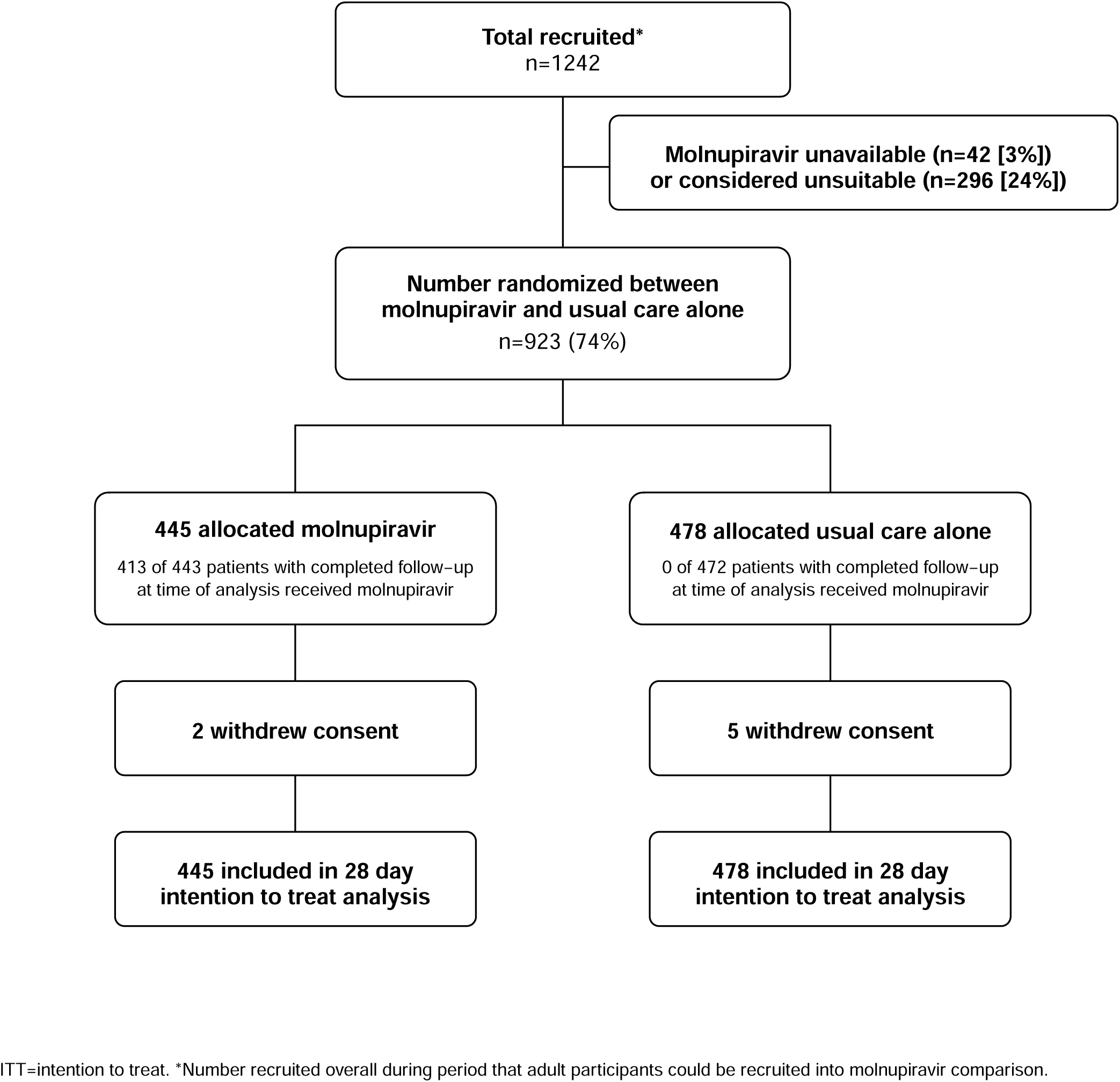
Trial profile for molnupiravir comparison.

**Figure 1B:**
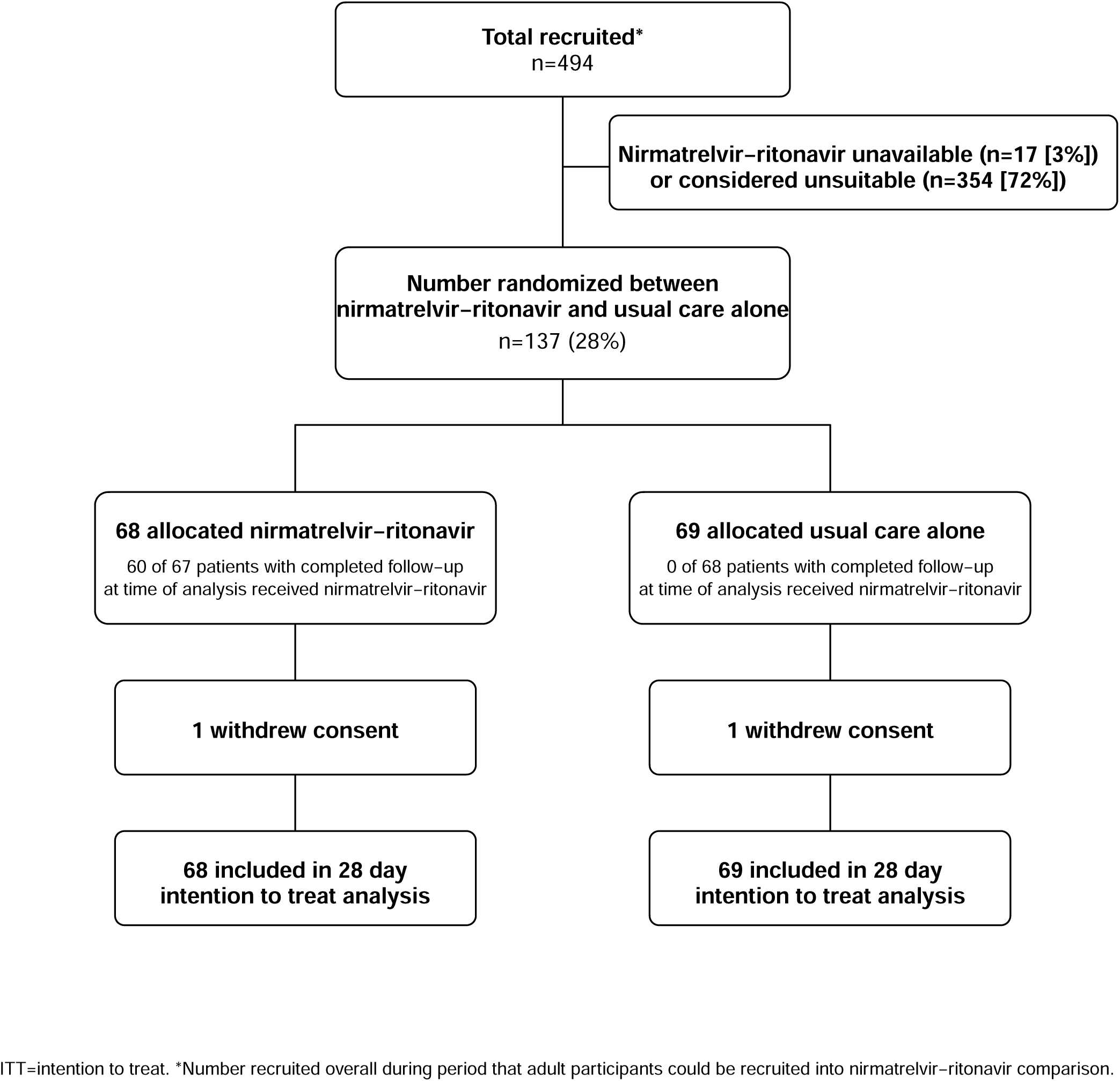
Trial profile for nirmatrelvir-ritonavir comparison.

**Table 1:**
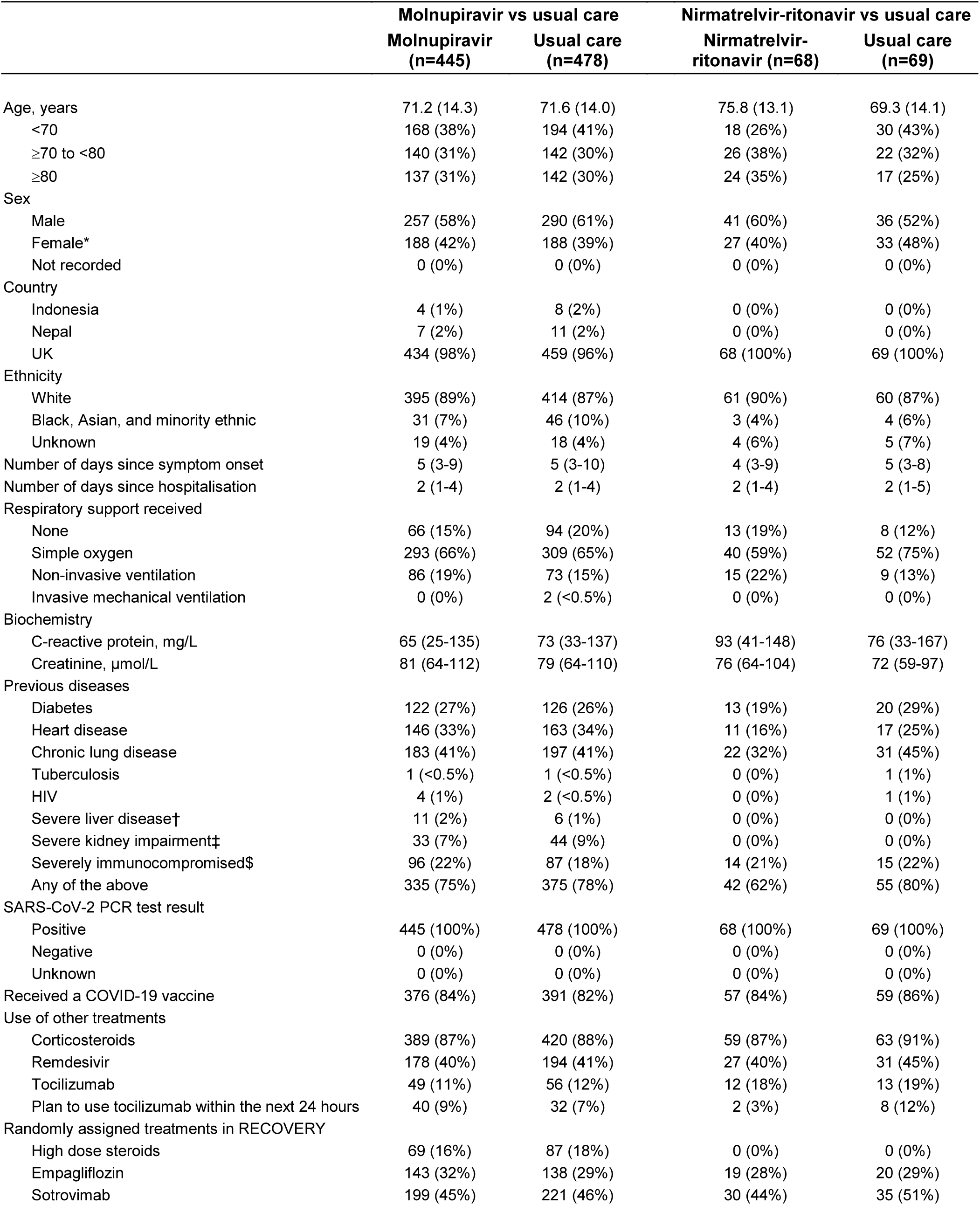
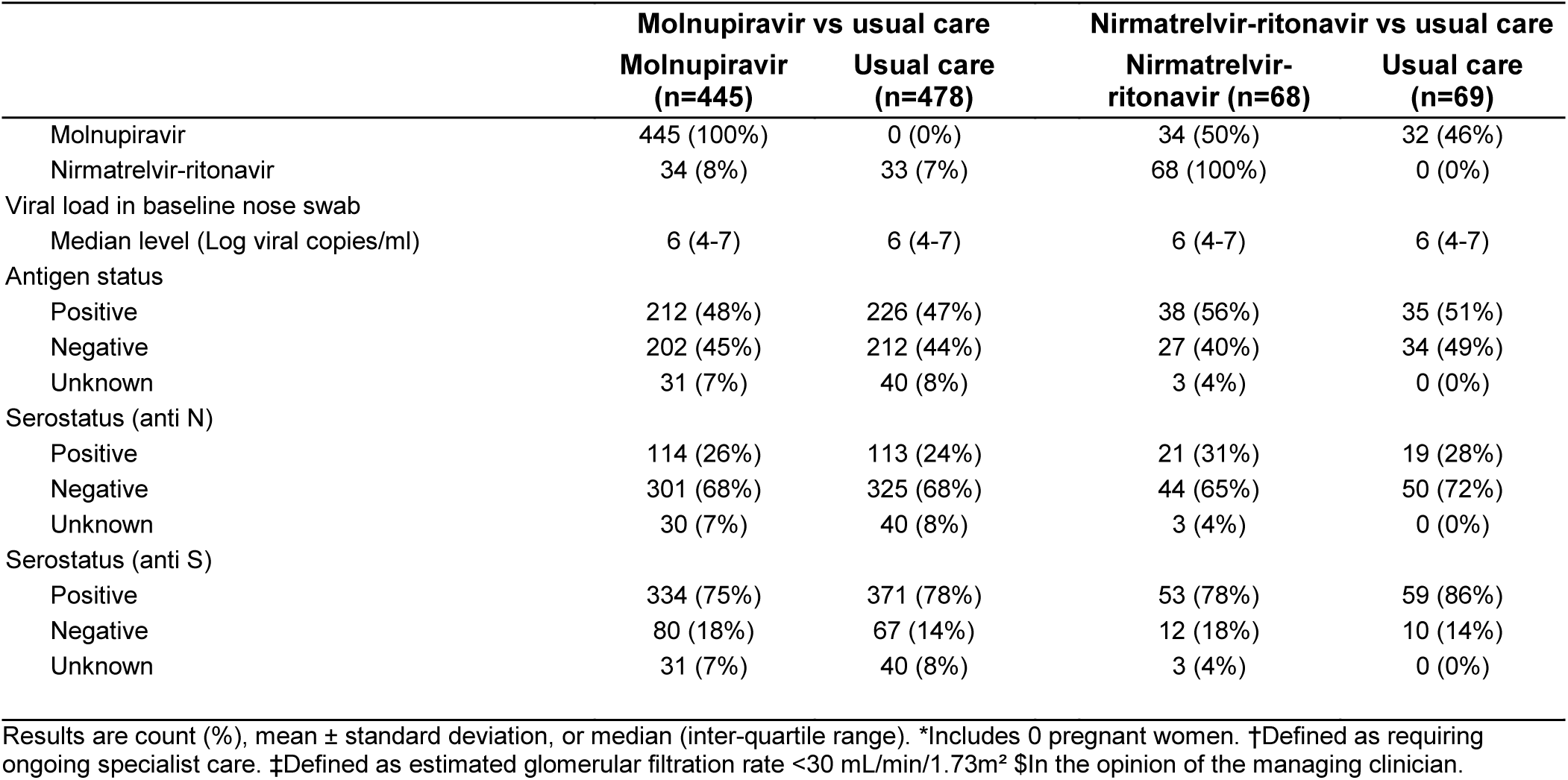
Baseline characteristics.

|  | Molnupiravir vs usual care |  | Nirmatrelvir-ritonavir vs usual care |  |
| --- | --- | --- | --- | --- |
|  | Molnupiravir<br>(n=445) | Usual care<br>(n=478) | Nirmatrelvir-ritonavir (n=68) | Usual care<br>(n=69) |
| Age, years | 71.2 (14.3) | 71.6 (14.0) | 75.8 (13.1) | 69.3 (14.1) |
| <70 | 168 (38%) | 194 (41%) | 18 (26%) | 30 (43%) |
| ≥70 to <80 | 140 (31%) | 142 (30%) | 26 (38%) | 22 (32%) |
| ≥80 | 137 (31%) | 142 (30%) | 24 (35%) | 17 (25%) |
| Sex |  |  |  |  |
| Male | 257 (58%) | 290 (61%) | 41 (60%) | 36 (52%) |
| Female* | 188 (42%) | 188 (39%) | 27 (40%) | 33 (48%) |
| Not recorded | 0 (0%) | 0 (0%) | 0 (0%) | 0 (0%) |
| Country |  |  |  |  |
| Indonesia | 4 (1%) | 8 (2%) | 0 (0%) | 0 (0%) |
| Nepal | 7 (2%) | 11 (2%) | 0 (0%) | 0 (0%) |
| UK | 434 (98%) | 459 (96%) | 68 (100%) | 69 (100%) |
| Ethnicity |  |  |  |  |
| White | 395 (89%) | 414 (87%) | 61 (90%) | 60 (87%) |
| Black, Asian, and minority ethnic | 31 (7%) | 46 (10%) | 3 (4%) | 4 (6%) |
| Unknown | 19 (4%) | 18 (4%) | 4 (6%) | 5 (7%) |
| Number of days since symptom onset | 5 (3-9) | 5 (3-10) | 4 (3-9) | 5 (3-8) |
| Number of days since hospitalisation | 2 (1-4) | 2 (1-4) | 2 (1-4) | 2 (1-5) |
| Respiratory support received |  |  |  |  |
| None | 66 (15%) | 94 (20%) | 13 (19%) | 8 (12%) |
| Simple oxygen | 293 (66%) | 309 (65%) | 40 (59%) | 52 (75%) |
| Non-invasive ventilation | 86 (19%) | 73 (15%) | 15 (22%) | 9 (13%) |
| Invasive mechanical ventilation | 0 (0%) | 2 (<0.5%) | 0 (0%) | 0 (0%) |
| Biochemistry |  |  |  |  |
| C-reactive protein, mg/L | 65 (25-135) | 73 (33-137) | 93 (41-148) | 76 (33-167) |
| Creatinine, µmol/L | 81 (64-112) | 79 (64-110) | 76 (64-104) | 72 (59-97) |
| Previous diseases |  |  |  |  |
| Diabetes | 122 (27%) | 126 (26%) | 13 (19%) | 20 (29%) |
| Heart disease | 146 (33%) | 163 (34%) | 11 (16%) | 17 (25%) |
| Chronic lung disease | 183 (41%) | 197 (41%) | 22 (32%) | 31 (45%) |
| Tuberculosis | 1 (<0.5%) | 1 (<0.5%) | 0 (0%) | 1 (1%) |
| HIV | 4 (1%) | 2 (<0.5%) | 0 (0%) | 1 (1%) |
| Severe liver disease† | 11 (2%) | 6 (1%) | 0 (0%) | 0 (0%) |
| Severe kidney impairment‡ | 33 (7%) | 44 (9%) | 0 (0%) | 0 (0%) |
| Severely immunocompromised\$ | 96 (22%) | 87 (18%) | 14 (21%) | 15 (22%) |
| Any of the above | 335 (75%) | 375 (78%) | 42 (62%) | 55 (80%) |
| SARS-CoV-2 PCR test result |  |  |  |  |
| Positive | 445 (100%) | 478 (100%) | 68 (100%) | 69 (100%) |
| Negative | 0 (0%) | 0 (0%) | 0 (0%) | 0 (0%) |
| Unknown | 0 (0%) | 0 (0%) | 0 (0%) | 0 (0%) |
| Received a COVID-19 vaccine | 376 (84%) | 391 (82%) | 57 (84%) | 59 (86%) |
| Use of other treatments |  |  |  |  |
| Corticosteroids | 389 (87%) | 420 (88%) | 59 (87%) | 63 (91%) |
| Remdesivir | 178 (40%) | 194 (41%) | 27 (40%) | 31 (45%) |
| Tocilizumab | 49 (11%) | 56 (12%) | 12 (18%) | 13 (19%) |
| Plan to use tocilizumab within the next 24 hours | 40 (9%) | 32 (7%) | 2 (3%) | 8 (12%) |
| Randomly assigned treatments in RECOVERY |  |  |  |  |
| High dose steroids | 69 (16%) | 87 (18%) | 0 (0%) | 0 (0%) |
| Empagliflozin | 143 (32%) | 138 (29%) | 19 (28%) | 20 (29%) |
| Sotrovimab | 199 (45%) | 221 (46%) | 30 (44%) | 35 (51%) |

Molnupiravir or nirmatrelvir-ritonavir for COVID-19
|  | Molnupiravir vs usual care |  | Nirmatrelvir-ritonavir vs usual care |  |
| --- | --- | --- | --- | --- |
|  | Molnupiravir<br>(n=445) | Usual care<br>(n=478) | Nirmatrelvir-ritonavir (n=68) | Usual care<br>(n=69) |
| Molnupiravir | 445 (100%) | 0 (0%) | 34 (50%) | 32 (46%) |
| Nirmatrelvir-ritonavir | 34 (8%) | 33 (7%) | 68 (100%) | 0 (0%) |
| Viral load in baseline nose swab |  |  |  |  |
| Median level (Log viral copies/ml) | 6 (4-7) | 6 (4-7) | 6 (4-7) | 6 (4-7) |
| Antigen status |  |  |  |  |
| Positive | 212 (48%) | 226 (47%) | 38 (56%) | 35 (51%) |
| Negative | 202 (45%) | 212 (44%) | 27 (40%) | 34 (49%) |
| Unknown | 31 (7%) | 40 (8%) | 3 (4%) | 0 (0%) |
| Serostatus (anti N) |  |  |  |  |
| Positive | 114 (26%) | 113 (24%) | 21 (31%) | 19 (28%) |
| Negative | 301 (68%) | 325 (68%) | 44 (65%) | 50 (72%) |
| Unknown | 30 (7%) | 40 (8%) | 3 (4%) | 0 (0%) |
| Serostatus (anti S) |  |  |  |  |
| Positive | 334 (75%) | 371 (78%) | 53 (78%) | 59 (86%) |
| Negative | 80 (18%) | 67 (14%) | 12 (18%) | 10 (14%) |
| Unknown | 31 (7%) | 40 (8%) | 3 (4%) | 0 (0%) |

Results are count (%), mean ± standard deviation, or median (inter-quartile range). \*Includes 0 pregnant women. †Defined as requiring ongoing specialist care. ‡Defined as estimated glomerular filtration rate <30 mL/min/1.73m<sup>2</sup> \$In the opinion of the managing clinician.

The follow-up form was completed for 915 (99%) patients, and among them, 413/443 (93%) in the molnupiravir group received at least one dose of molnupiravir, compared to 0/472 (0%) in the usual care group (webtable 3). Primary and secondary outcome data are known for >99% of randomly assigned patients. There was no evidence of a significant difference in the proportion of patients who met the primary outcome of 28-day mortality between the two randomised groups (74 [17%] patients in the molnupiravir group vs. 79 [17%] patients in the usual care group; hazard ratio 0.93; 95% confidence interval [CI], 0.68-1.28; p=0.66; table 2, figure 2A). We observed similar results in all pre-specified sub-groups, and in exploratory subgroups defined by serum SARS-CoV-2 antigen or antibody status, and use of other SARS-CoV-2 antiviral treatments (figure 3).

**Figure 2A:**
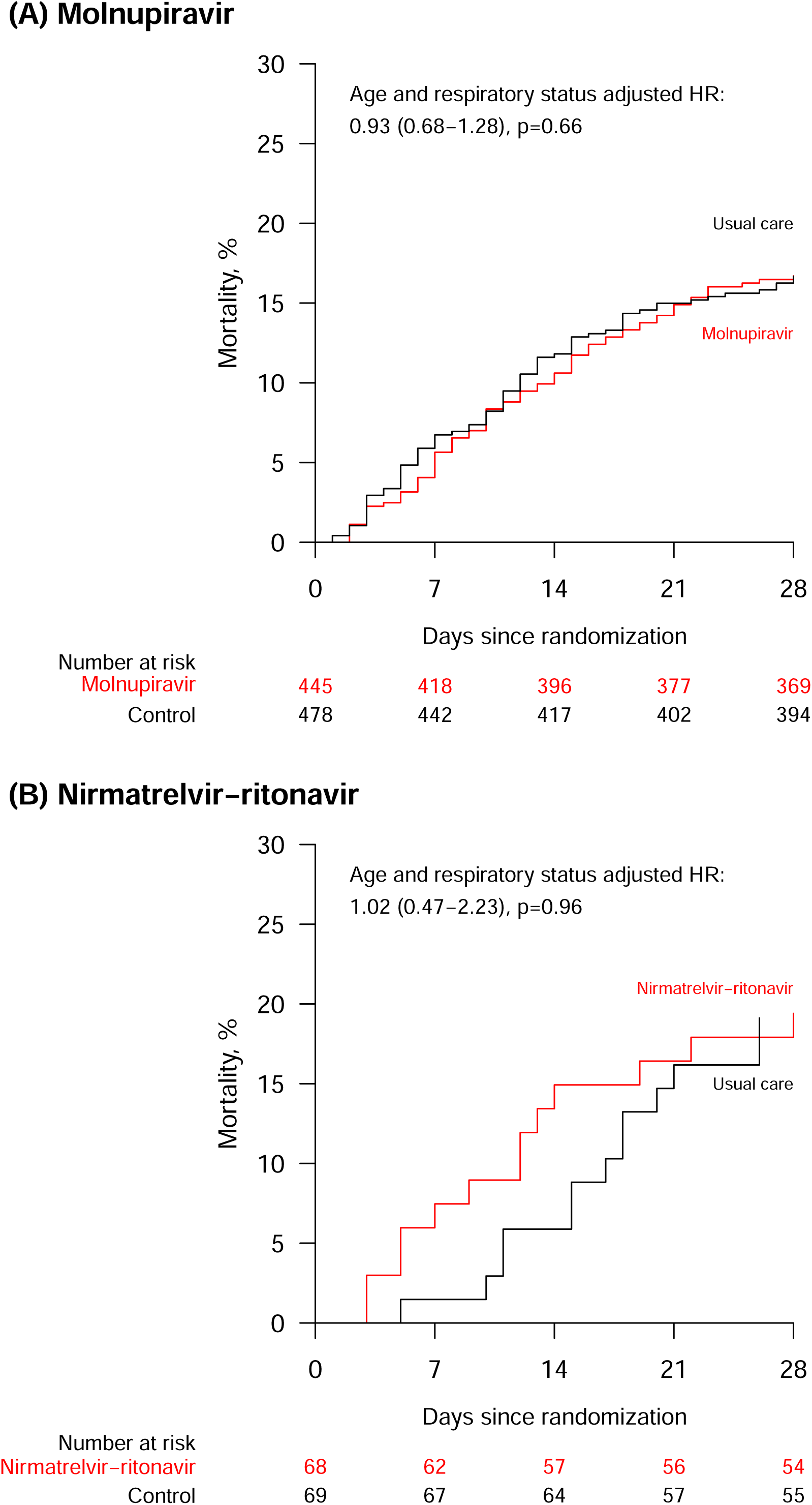
Effect of allocation to molnupiravir and nirmatrelvir-ritonavir on 28-day mortality.

**Figure 3:**
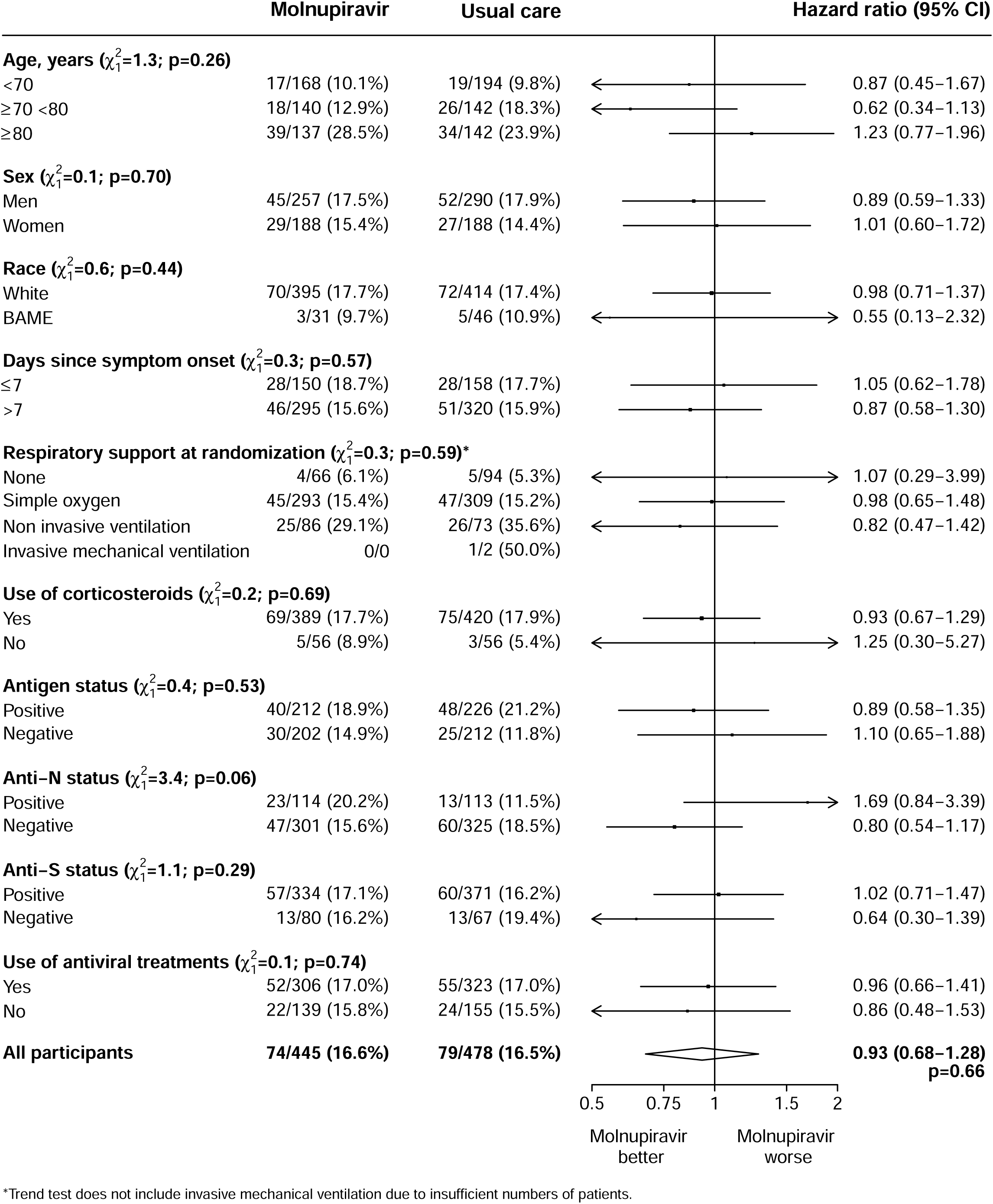
Effects of allocation to molnupiravir on 28-day mortality by baseline characteristics.

**Table 2:** Effect of allocation to molnupiravir and nirmatrelvir-ritonavir on key study outcomes.

|  | Molnupiravir vs Usual care |  |  |  | Nirmatrelvir-ritonavir vs Usual care |  |  |  |
| --- | --- | --- | --- | --- | --- | --- | --- | --- |
|  | Molnupiravir<br>(n=445) | Usual care<br>(n=478) | RR or mean<br>difference<br>(95% CI) | p value | Nirmatrelvir-ritonavir<br>(n=68) | Usual care<br>(n=69) | RR or mean<br>difference<br>(95% CI) | p value |
| **Primary outcome:** |  |  |  |  |  |  |  |  |
| 28-day mortality | 74 (17%) | 79 (17%) | 0.93 (0.68-1.28) | 0.66 | 13 (19%) | 13 (19%) | 1.02 (0.47-2.23) | 0.96 |
| **Secondary outcomes:** |  |  |  |  |  |  |  |  |
| Median time to being discharged alive, days | 10 (6 to >28) | 9 (5 to >28) |  |  | 10 (6 to >28) | 8 (5 to 21) |  |  |
| Discharged from hospital within 28 days | 319 (72%) | 354 (74%) | 0.96 (0.82-1.12) | 0.60 | 48 (71%) | 54 (78%) | 0.80 (0.54-1.20) | 0.29 |
| Receipt of invasive mechanical ventilation or death* | 77/445 (17%) | 81/476 (17%) | 0.96 (0.73-1.25) | 0.75 | 14/68 (21%) | 13/69 (19%) | 1.06 (0.54-2.08) | 0.86 |
| Invasive mechanical ventilation | 8/445 (2%) | 6/476 (1%) | 1.27 (0.45-3.60) | 0.65 | 1/68 (1%) | 1/69 (1%) | - | - |
| Death | 74/445 (17%) | 78/476 (16%) | 0.96 (0.73-1.26) | 0.77 | 13/68 (19%) | 13/69 (19%) | 0.98 (0.49-1.94) | 0.94 |
| **Subsidiary clinical outcomes** |  |  |  |  |  |  |  |  |
| Receipt of ventilation† | 38/359 (11%) | 34/403 (8%) | 1.24 (0.80-1.92) | 0.34 | 4/53 (8%) | 10/60 (17%) | 0.56 (0.18-1.73) | 0.31 |
| Non-invasive ventilation | 35/359 (10%) | 34/403 (8%) | 1.14 (0.73-1.78) | 0.58 | 4/53 (8%) | 10/60 (17%) | 0.56 (0.18-1.73) | 0.31 |
| Invasive mechanical ventilation | 5/359 (1%) | 1/403 (0%) | 5.66 (0.66-48.37) | 0.11 | 0/53 (0%) | 1/60 (2%) | - | - |
| Successful cessation of invasive mechanical ventilation‡ | 0/0 | 0/2 (0%) | - | - | 0/0 | 0/0 | - | - |
| Renal replacement therapy§ | 5/436 (1%) | 9/469 (2%) | 0.62 (0.20-1.86) | 0.39 | 0/68 (0%) | 1/69 (1%) | - | - |
| **Virological outcomes** |  |  |  |  |  |  |  |  |
| Baseline-adjusted viral load (log copies/ml) on day 3 | 4.36 (0.09) | 4.48 (0.09) | -0.13 (-0.38, 0.12) | 0.32 | 4.04 (0.24) | 4.41 (0.22) | -0.36 (-0.99, 0.26) | 0.26 |
| Baseline-adjusted viral load (log copies/ml) on day 5 | 3.51 (0.10) | 3.99 (0.13) | -0.48 (-0.80, -0.16) | 0.0037 | 2.90 (0.22) | 3.57 (0.22) | -0.68 (-1.29, -0.07) | 0.03 |

RR=Hazard ratio for the outcomes of 28-day mortality and hospital discharge, and risk ratio for the outcome of receipt of invasive mechanical ventilation or death (and its subcomponents).
CI=confidence interval. \*Analyses exclude those on invasive mechanical ventilation at randomization. †Analyses exclude those on any form of ventilation at randomisation. ‡Analyses restricted to those on invasive mechanical ventilation at randomisation. §Analyses exclude those on haemodialysis or haemofiltration at randomisation.

There was no evidence of a significant difference in the probability of being discharged alive within 28 days (72% vs. 74%, rate ratio 0.96, 95% CI 0.82 to 1.12, p=0.60) (table 2). Among those not on invasive mechanical ventilation at baseline, the number of patients progressing to the pre-specified composite secondary outcome of invasive mechanical ventilation or death was similar in both groups (17% vs. 17%, risk ratio 0.96, 95% CI 0.73 to 1.25, p=0.75). Similar results were seen in all pre-specified subgroups of patients (webfigures 1 and 2).

We found no evidence of significant differences in prespecified subsidiary clinical outcomes or cause-specific mortality between groups (table 2, webtable 4). There were more episodes of hyperglycaemia requiring insulin in patients allocated to molnupiravir versus usual care (7.4% vs 3.1%, absolute difference 4.3%, [95% CI 1.4-7.2] p=0.0038) (webtable 5). The rates of other safety outcomes were similar between groups, including new cardiac arrhythmia, thrombotic events, clinically significant bleeds, non-coronavirus infections, seizures, acute liver injury, and acute kidney injury (webtable 5). There were no reported suspected serious adverse reactions in patients allocated molnupiravir.

872/893 (98%) of UK patients had at least one nose swab available for analysis. Allocation to molnupiravir was associated with a lower baseline-adjusted viral load in nose swabs taken on day 5 (-0.48 log10 copies/ml; 95% CI -0.80 to -0.16; p=0.0037), but not on day 3 (table 2). 622 (67%) patients had at least one successfully sequenced sample with ≥90% genome coverage, and of these 620 (>99%) were Omicron variants (primarily BA.1, BA.2, BA.5, and XBB). No candidate molnupiravir resistance mutations were identified from literature searches, so we were not able evaluate baseline or follow-up nose swabs for mutations associated with resistance.

### Nirmatrelvir-ritonavir comparison

Between 31 March 2022 and 24 May 2023, 137/494 (28%) of patients recruited at sites participating in the nirmatrelvir-ritonavir comparison were eligible to be randomly allocated to nirmatrelvir-ritonavir, of whom 68 were allocated nirmatrelvir-ritonavir and 69 allocated to usual care without nirmatrelvir-ritonavir (figure 1B). The 357 RECOVERY patients not included in the nirmatrelvir-ritonavir comparison had similar characteristics to those included (webtable 2). The mean age of study participants in this comparison was 72.5 years (SD 13.9), 116 (85%) had received a COVID-19 vaccine, and the median time since symptom onset was 4 days (IQR 3 to 8 days). 133 (97%) patients participating in the nirmatrelvir-ritonavir comparison also participated in the molnupiravir comparison. At randomisation, 122 (89%) patients were receiving corticosteroids, and 111 (81%) were receiving, or allocated to receive, a SARS-CoV-2 antiviral other than nirmatrelvir-ritonavir (including usual care remdesivir, and sotrovimab or molnupiravir allocated in another RECOVERY comparison). 40 (29%) patients were anti-N seropositive and 112 (82%) were anti-S seropositive.

The follow-up form was completed for 135 (99%) patients, and among them, 60/67 (90%) in the nirmatrelvir-ritonavir group received at least one dose of nirmatrelvir-ritonavir, compared to 0/68 (0%) in the usual care group (webtable 3). Primary and secondary outcome data are known for >99% of randomly assigned patients. There was no evidence of a significant difference in the proportion of patients who met the primary outcome of 28-day mortality between the two randomised groups (13 [19%] patients in the nirmatrelvir-ritonavir group vs. 13 [19%] patients in the usual care group; hazard ratio 1.02; 95% CI, 0.47-2.23; p=0.96; table 2, figure 2B). Because of low recruitment to this comparison, no subgroup analyses were performed.

There was no evidence of a significant difference in the probability of being discharged alive within 28 days (71% vs. 78%, rate ratio 0.80, 95% CI 0.54 to 1.20, p=0.29) (table 2). Among those not on invasive mechanical ventilation at baseline, the number of patients progressing to the pre-specified composite secondary outcome of invasive mechanical ventilation or death was similar in both groups (21% vs. 19%, risk ratio 1.06, 95% CI 0.54 to 2.08, p=0.86).

We found no evidence of significant differences in prespecified subsidiary clinical outcomes or cause-specific mortality between groups (table 2, webtable 4). The rates of all safety outcomes were similar between groups (webtable 5). There were no reported suspected serious adverse reactions in patients allocated nirmatrelvir-ritonavir.

All patients had at least one nose swab available for analysis. Allocation to nirmatrelvir-ritonavir was associated with a significantly lower baseline-adjusted viral load in nose swabs taken on day 5 (-0.68 log10 copies/ml; 95% CI -1.29 to -0.07; p=0.03), but not on day 3 (table 2). 97 (71%) patients had at least one sample successfully sequenced with ≥90% genome coverage, and of these 96 (99%) were Omicron variants. No sequenced samples contained mutations at the 20 nucleotide positions in the 3CL protease that had previously been associated with >2.5 fold median reduction in inhibition by nirmatrelvir.

## DISCUSSION

In these two reported evaluations from the RECOVERY trial, among patients admitted to hospital for severe COVID-19, neither molnupiravir nor nirmatrelvir-ritonavir was found to reduce mortality, duration of hospitalisation, or the risk of being ventilated or dying for those not on ventilation at baseline. However, both comparisons lacked statistical power to exclude modest differences in these outcomes.

Previous trials have indicated the potential benefit of antiviral treatment with nMAbs or remdesivir in hospitalised patients, but randomised evidence has been inadequate for molnupiravir and nirmatrelvir-ritonavir, two widely available antivirals with efficacy in early infection. For each drug, only one other randomised trial in hospitalised COVID-19 patients has been reported to date, but neither were large enough to detect plausibly moderate benefits of treatment.^14, 17^ The present RECOVERY comparisons were both stopped because of low recruitment before they had reached the planned sample size, with 923 patients recruited to the molnupiravir comparison and 137 recruited to the nirmatrelvir-ritonavir comparison. Our results do not suggest any benefit in adding these antivirals to routine care, but limited recruitment means we cannot exclude a benefit.

The incidence of COVID-19 pneumonia has reduced substantially following widespread vaccination starting in 2021 and the global dominance of Omicron SARS-CoV-2 variants in 2022. In this context, infection with SARS-CoV-2 in hospitalised patients is often an incidental finding, or is associated with non-respiratory illness, and the benefits of antiviral therapy in this setting may be limited. By contrast, RECOVERY only included patients with pneumonia thought to be related to COVID-19. In over 80% of participants this had developed despite previous COVID-19 vaccination, and in keeping with this only around a quarter of participants were anti-spike antibody negative at baseline, but around three-quarters were anti-nucleocapsid antibody negative, indicating that this was their first SARS-CoV-2 infection.

The power to perform subgroup analyses was limited even in the molnupiravir comparison, and here there was no strong signal of a differential effect of treatment in patients by antibody status, level of serum viral antigen, use of other antiviral treatments, symptom duration, or severity of illness. In patients allocated molnupiravir there was an excess of hyperglycaemia requiring insulin compared to usual care, reported in 33 vs 15 patients. An excess of hyperglycaemia was also reported in the MOVe-IN trial (9 vs 1 events), but there is no apparent mechanism to explain this, and these may represent chance findings. The increased viral clearance in day 5 nose swabs seen in those allocated molnupiravir is in keeping with its known antiviral activity, and with results from trials in early infection, although this has not previously been demonstrated in hospitalised patients.^12, 14, 31, 32^ Nevertheless, this reduction in viral load was not shown in this trial to translate into clinical benefit.

Recruitment to the nirmatrelvir-ritonavir comparison was substantially lower than the molnupiravir comparison, reflecting its introduction just after the initial wave of Omicron in the UK in early 2022, the involvement of fewer hospital sites, and a high proportion of patients for whom it was considered unsuitable. Reasons for unsuitability were not systematically recorded, but this was frequently related to potential interactions between ritonavir and concomitant medications. We were therefore unable to reliably assess whether nirmatrelvir improves clinical outcomes, although a reduction in viral load among participants allocated nirmatrelvir was observed.

Strengths of this trial include that it was randomised, had broad eligibility criteria, baseline characterisation of markers of SARS-CoV-2 immune status and infection, and more than 99% of patients were followed up for the primary outcome. However, the limited sample size does not allow us to exclude modest benefits of the treatments tested. Also, use of other antiviral treatments was common in both comparisons, and it is possible that the treatments tested may have had a greater effect in the absence of other antivirals. Although this randomised trial is open label (i.e. participants and local hospital staff were aware of the assigned treatment), the primary and secondary outcomes are unambiguous and were ascertained without bias through linkage to routine health records in the large majority of patients. However, detailed information on radiological or physiological outcomes was not collected The RECOVERY trial only studied patients who had been hospitalised with COVID-19 and, therefore, is not able to provide any evidence on the safety and efficacy of these antivirals used in other patient groups. Due to the recommendation that both drugs be taken orally, and not via a gastric feeding tube, there were few patients recruited requiring invasive mechanical ventilation.

In summary, the results of this randomised trial do not support the use of molnupiravir or nirmatrelvir-ritonavir as a treatment for adults hospitalised with COVID-19.

## Contributors

This manuscript was initially drafted by LP, RH, PWH and MJL, further developed by the Writing Committee, and approved by all members of the trial steering committee. PWH and MJL vouch for the data and analyses, and for the fidelity of this report to the study protocol and data analysis plan. PWH, JKB, MB, SNF, TJ, EJ, KJ, MK, WSL, AMo, AMuk, AMum, JN, KR, GT, MM, RH, and MJL designed the trial and study protocol. MM, MC, G P-A, LP, RS, DG, FH, PD, NE, JM, PH, DC, GC, SS, HE, the Data Linkage team at the RECOVERY Coordinating Centre, and the Health Records and Local Clinical Centre staff listed in the appendix collected the data. NS did the statistical analysis. All authors contributed to data interpretation and critical review and revision of the manuscript. PWH and MJL had access to the study data and had final responsibility for the decision to submit for publication.

## Writing Committee (on behalf of the RECOVERY Collaborative Group)

Peter W Horby*, Natalie Staplin*, Leon Peto*, Jonathan R Emberson, Mark Campbell, Guilherme Pessoa-Amorim, Buddha Basnyat, Louise Thwaites, Rogier van Doorn, Raph L Hamers, Jeremy Nel, John Amuasi, Richard Stewart, Dipansu Ghosh, Fergus Hamilton, Purav Desai, Nicholas Easom, Jaydip Majumdar, Paul Hine, David Chadwick, Graham Cooke, Sara Sharp, Hanif Esmail, J Kenneth Baillie, Maya Buch, Saul N Faust, Thomas Jaki, Edmund Juszczak, Katie Jeffery, Marian Knight, Wei Shen Lim, Alan Montgomery, Aparna Mukherjee, Andrew Mumford, Kathryn Rowan, Guy Thwaites, Marion Mafham^†^, Richard Haynes^†^, Martin J Landray^†^.

*,^†^ equal contribution

## Data Monitoring Committee

Peter Sandercock, Janet Darbyshire, David DeMets, Robert Fowler, David Lalloo, Mohammed Munavvar, Janet Wittes.

## Data sharing

The protocol, consent form, statistical analysis plan, definition & derivation of clinical characteristics & outcomes, training materials, regulatory documents, and other relevant study materials are available online at www.recoverytrial.net. As described in the protocol, the Trial Steering Committee will facilitate the use of the study data and approval will not be unreasonably withheld. Deidentified participant data will be made available to bona fide researchers registered with an appropriate institution within 3 months of publication. However, the Steering Committee will need to be satisfied that any proposed publication is of high quality, honours the commitments made to the study participants in the consent documentation and ethical approvals, and is compliant with relevant legal and regulatory requirements (e.g. relating to data protection and privacy). The Steering Committee will have the right to review and comment on any draft manuscripts prior to publication. Data will be made available in line with the policy and procedures described at: https://www.ndph.ox.ac.uk/data-access. Those wishing to request access should complete the form at https://www.ndph.ox.ac.uk/files/about/data_access_enquiry_form_13_6_2019.docx and e-mailed to

## Data Availability

The protocol, consent form, statistical analysis plan, definition & derivation of clinical characteristics & outcomes, training materials, regulatory documents, and other relevant study materials are available online at www.recoverytrial.net. As described in the protocol, the Trial Steering Committee will facilitate the use of the study data and approval will not be unreasonably withheld. Deidentified participant data will be made available to bona fide researchers registered with an appropriate institution within 3 months of publication. However, the Steering Committee will need to be satisfied that any proposed publication is of high quality, honours the commitments made to the study participants in the consent documentation and ethical approvals, and is compliant with relevant legal and regulatory requirements (e.g. relating to data protection and privacy). The Steering Committee will have the right to review and comment on any draft manuscripts prior to publication. Data will be made available in line with the policy and procedures described at: https://www.ndph.ox.ac.uk/data-access. Those wishing to request access should complete the form at
https://www.ndph.ox.ac.uk/files/about/data_access_enquiry_form_13_6_2019.docx
and e-mailed to

## Acknowledgements

Above all, we would like to thank the patients who participated in this trial. We would also like to thank the many doctors, nurses, pharmacists, other allied health professionals, and research administrators at NHS hospital organisations across the whole of the UK, supported by staff at the National Institute of Health and Care Research (NIHR) Clinical Research Network, NHS DigiTrials, UK Health Security Agency, Department of Health & Social Care, the Intensive Care National Audit & Research Centre, Public Health Scotland, National Records Service of Scotland, the Secure Anonymised Information Linkage (SAIL) at University of Swansea, and the NHS in England, Scotland, Wales and Northern Ireland.

The RECOVERY trial was supported by grants to the University of Oxford from UK Research and Innovation (UKRI) and NIHR (MC_PC_19056), the Wellcome Trust (Grant Ref: 222406/Z/20/Z) through the COVID-19 Therapeutics Accelerator, and by core funding provided by the NIHR Oxford Biomedical Research Centre, the Wellcome Trust, the Bill and Melinda Gates Foundation, the Foreign, Commonwealth and Development Office, Health Data Research UK, the Medical Research Council, the NIHR Health Protection Unit in Emerging and Zoonotic Infections, and NIHR Clinical Trials Unit Support Funding. TJ is supported by grants from UK Medical Research Council (MC_UU_0002/14 and MC_UU_00040/03). WSL is supported by core funding provided by NIHR Nottingham Biomedical Research Centre.

The views expressed in this publication are those of the authors and not necessarily those of the NHS, the NIHR, or the UK Department of Health and Social Care.

## References

1 Weinreich DM, Sivapalasingam S, Norton T, et al. REGN-COV2, a Neutralizing Antibody Cocktail, in Outpatients with Covid-19. N Engl J Med 2021; 384: 238–51.

2 Gottlieb RL, Vaca CE, Paredes R, et al. Early Remdesivir to Prevent Progression to Severe Covid-19 in Outpatients. N Engl J Med 2022; 386: 305–15.

3 Hammond J, Leister-Tebbe H, Gardner A, et al. Oral Nirmatrelvir for High-Risk, Nonhospitalized Adults with Covid-19. N Engl J Med 2022; published online Feb 16. DOI:10.1056/NEJMoa2118542.

4 RECOVERY Collaborative Group. Casirivimab and imdevimab in patients admitted to hospital with COVID-19 (RECOVERY): a randomised, controlled, open-label, platform trial. Lancet Lond Engl 2022; 399: 665–76.

5 Bergeri I, Whelan MG, Ware H, et al. Global SARS-CoV-2 seroprevalence from January 2020 to April 2022: A systematic review and meta-analysis of standardized population-based studies. PLoS Med 2022; 19: e1004107.

6 COVID-19 vaccine quarterly surveillance reports (September 2021 to April 2024). GOV.UK. 2024; published online April 25. https://www.gov.uk/government/publications/covid-19-vaccine-weekly-surveillance-reports (accessed May 10, 2024).

7 WHO Solidarity Trial Consortium. Remdesivir and three other drugs for hospitalised patients with COVID-19: final results of the WHO Solidarity randomised trial and updated meta-analyses. Lancet Lond Engl 2022; 399: 1941–53.

8 Amstutz A, Speich B, Mentré F, et al. Effects of remdesivir in patients hospitalised with COVID-19: a systematic review and individual patient data meta-analysis of randomised controlled trials. Lancet Respir Med 2023; 11: 453–64.

9 Yoon J-J, Toots M, Lee S, et al. Orally Efficacious Broad-Spectrum Ribonucleoside Analog Inhibitor of Influenza and Respiratory Syncytial Viruses. Antimicrob Agents Chemother 2018; 62: e00766–18.

10 Sheahan TP, Sims AC, Zhou S, et al. An orally bioavailable broad-spectrum antiviral inhibits SARS-CoV-2 in human airway epithelial cell cultures and multiple coronaviruses in mice. Sci Transl Med 2020; 12: eabb5883.

11 Strizki JM, Gaspar JM, Howe JA, et al. Molnupiravir maintains antiviral activity against SARS-CoV-2 variants and exhibits a high barrier to the development of resistance. Antimicrob Agents Chemother 2024; 68: e0095323.

12 Jayk Bernal A, Gomes da Silva MM, Musungaie DB, et al. Molnupiravir for Oral Treatment of Covid-19 in Nonhospitalized Patients. N Engl J Med 2022; 386: 509–20.

13 Butler CC, Hobbs FDR, Gbinigie OA, et al. Molnupiravir plus usual care versus usual care alone as early treatment for adults with COVID-19 at increased risk of adverse outcomes (PANORAMIC): an open-label, platform-adaptive randomised controlled trial. Lancet Lond Engl 2023; 401: 281–93.

14 Arribas JR, Bhagani S, Lobo SM, et al. Randomized Trial of Molnupiravir or Placebo in Patients Hospitalized with Covid-19. NEJM Evid 2022; 1: EVIDoa2100044.

15 Owen DR, Allerton CMN, Anderson AS, et al. An oral SARS-CoV-2 Mpro inhibitor clinical candidate for the treatment of COVID-19. Science 2021; 374: 1586–93.

16 Hammond J, Fountaine RJ, Yunis C, et al. Nirmatrelvir for Vaccinated or Unvaccinated Adult Outpatients with Covid-19. N Engl J Med 2024; 390: 1186–95.

17 Liu J, Pan X, Zhang S, et al. Efficacy and safety of Paxlovid in severe adult patients with SARS-Cov-2 infection: a multicenter randomized controlled study. Lancet Reg Health West Pac 2023; 33: 100694.

18 RECOVERY Collaborative Group, Horby P, Lim WS, et al. Dexamethasone in Hospitalized Patients with Covid-19. N Engl J Med 2021; 384: 693–704.

19 RECOVERY Collaborative Group, Horby P, Mafham M, et al. Effect of Hydroxychloroquine in Hospitalized Patients with Covid-19. N Engl J Med 2020; 383: 2030–40.

20 RECOVERY Collaborative Group. Lopinavir-ritonavir in patients admitted to hospital with COVID-19 (RECOVERY): a randomised, controlled, open-label, platform trial. Lancet Lond Engl 2020; 396: 1345–52.

21 RECOVERY Collaborative Group. Azithromycin in patients admitted to hospital with COVID-19 (RECOVERY): a randomised, controlled, open-label, platform trial. Lancet Lond Engl 2021; 397: 605–12.

22 RECOVERY Collaborative Group. Tocilizumab in patients admitted to hospital with COVID-19 (RECOVERY): a randomised, controlled, open-label, platform trial. Lancet Lond Engl 2021; 397: 1637–45.

23 RECOVERY Collaborative Group. Convalescent plasma in patients admitted to hospital with COVID-19 (RECOVERY): a randomised controlled, open-label, platform trial. Lancet Lond Engl 2021; 397: 2049–59.

24 RECOVERY Collaborative Group. Colchicine in patients admitted to hospital with COVID-19 (RECOVERY): a randomised, controlled, open-label, platform trial. Lancet Respir Med 2021; 9: 1419–26.

25 RECOVERY Collaborative Group. Aspirin in patients admitted to hospital with COVID-19 (RECOVERY): a randomised, controlled, open-label, platform trial. Lancet Lond Engl 2022; 399: 143–51.

26 RECOVERY Collaborative Group. Baricitinib in patients admitted to hospital with COVID-19 (RECOVERY): a randomised, controlled, open-label, platform trial and updated meta-analysis. Lancet Lond Engl 2022; 400: 359–68.

27 RECOVERY Collaborative Group. Empagliflozin in patients admitted to hospital with COVID-19 (RECOVERY): a randomised, controlled, open-label, platform trial. Lancet Diabetes Endocrinol 2023; 11: 905–14.

28 RECOVERY Collaborative Group, Horby PW, Peto L, et al. Dimethyl fumarate in patients admitted to hospital with COVID-19 (RECOVERY): a randomised, controlled, open-label, platform trial. Nat Commun 2024; 15: 924.

29 RECOVERY Collaborative Group. Higher dose corticosteroids in patients admitted to hospital with COVID-19 who are hypoxic but not requiring ventilatory support (RECOVERY): a randomised, controlled, open-label, platform trial. Lancet Lond Engl 2023; : S0140–6736(23)00510-X.

30 Constantinides B, Webster H, Gentry J, et al. Rapid turnaround multiplex sequencing of SARS-CoV-2: comparing tiling amplicon protocol performance. medRxiv 2022; : 2021.12.28.21268461.

31 Khoo SH, FitzGerald R, Saunders G, et al. Molnupiravir versus placebo in unvaccinated and vaccinated patients with early SARS-CoV-2 infection in the UK (AGILE CST-2): a randomised, placebo-controlled, double-blind, phase 2 trial. Lancet Infect Dis 2023; 23: 183–95.

32 Fischer W, Eron JJ, Holman W, et al. Molnupiravir, an Oral Antiviral Treatment for COVID-19. 2021; : 2021.06.17.21258639.

