## Appendix for "Molnupiravir or nirmatrelvir-ritonavir versus usual care in patients admitted to hospital with COVID-19 (RECOVERY): a randomised, controlled, open-label, platform trial"

#### SUPPLEMENTARY APPENDIX

#### RECOVERY Collaborative Group

Webtable 1: Baseline characteristics of patients considered unsuitable for randomization to molnupiravir compared with those randomized to molnupiravir versus usual care 60
Webtable 2: Baseline characteristics of patients considered unsuitable for randomization to nirmatrelvir-ritonavir compared with those randomized to nirmatrelvir-ritonavir versus usual care 62
Webtable 3: Treatments given during the follow-up period, by randomized allocation 64
Webtable 4: Effect of allocation to molnupiravir and nirmatrelvir-ritonavir on cause-specific 28-day mortality 65
Webtable 5: Effect of allocation to molnupiravir and nirmatrelvir-ritonavir on new cardiac arrhythmia, thrombotic events, clinically significant bleeds, non-coronavirus infections and metabolic complications 66
**Supplementary Figures** ........ **67**
Webfigure 1: Effects of allocation to molnupiravir on hospital discharge 68
by baseline characteristics 68
Webfigure 2: Effects of allocation to molnupiravir on invasive mechanical ventilation or death in those not on invasive mechanical ventilation at randomisation, by baseline characteristics 69
**Appendices** ........ **70**
Appendix 1: RECOVERY Trial Protocol V25.0 71
Appendix 2: RECOVERY Trial Statistical Analysis Plan V3.2 114
Appendix 3: Definition and Derivation of Baseline Characteristics and Outcomes 153

### Details of the RECOVERY Collaborative Group

#### Writing Committee

Peter W Horby[a,b,c]*, Natalie Staplin[d]*, Leon Peto[d,e]*, Jonathan R Emberson[d], Mark Campbell[d,e], Guilherme Pessoa-Amorim[d], Buddha Basnyat[f], Louise Thwaites[a,g],Rogier van Doorn[a,g], Raph L Hamers[a,h], Jeremy Nel[i], John Amuasi[j], Richard Stewart[k], Dipansu Ghosh[l], Fergus Hamilton[m], Purav Desai[n], Nicholas Easom[o,p], Jaydip Majumdar[q], Paul Hine[r], David Chadwick[s], Graham Cooke[t], Sara Sharp[u], Hanif Esmail[v], J Kenneth Baillie[w], Maya H Buch[x], Saul N Faust[y], Thomas Jaki[z,za], Katie Jeffery[g,zb], Edmund Juszczak[zc], Marian Knight[zd], Wei Shen Lim[ze], Alan Montgomery[zc], Aparna Mukherjee,[zf] Andrew Mumford[zg], Kathryn Rowan[zh], Guy Thwaites[a,h], Marion Mafham[d†], Richard Haynes[d,e†], Martin J Landray[d,zi†]

[a] Centre for Tropical Medicine and Global Health, Nuffield Department of Medicine, University of Oxford, Oxford, United Kingdom

[b] International Severe Acute Respiratory and emerging Infections Consortium (ISARIC), University of Oxford, Oxford, United Kingdom

[c] Pandemic Sciences Centre, University of Oxford, Oxford, United Kingdom

[d] Nuffield Department of Population Health, University of Oxford, Oxford, United Kingdom

[e] Oxford University Hospitals NHS Foundation Trust, Oxford, United Kingdom

[f] Oxford University Clinical Research Unit Nepal, Patan Academy of Health Sciences, Kathmandu, Nepal

[g] Oxford University Clinical Research Unit, Ho Chi Minh City, Viet Nam

[h] Oxford University Clinical Research Unit Indonesia, Faculty of Medicine, Universitas Indonesia, Jakarta, Indonesia

[i] Division of Infectious Diseases, Department of Medicine, University of the Witwatersrand, Johannesburg, South Africa

[j] Kumasi Center for Collaborative Research in Tropic Medicine, Kumasi, Ghana

[k] Milton Keynes University Hospital, Milton Keynes, United Kingdom

[l] Leeds Teaching Hospitals NHS Foundation Trust, Leeds, United Kingdom

[m] University Hospital of Bristol and Weston NHS Foundation Trust and University of Bristol, Bristol, United Kingdom

[n] Calderdale and Huddersfield NHS Foundation Trust, Huddersfield, United Kingdom

[o] Hull York Medical School, Hull, UK

[p] Hull University Teaching Hospitals NHS Trust, Hull, United Kingdom

[q] Mid Cheshire Hospitals NHS Foundation Trust, United Kingdom

[r] Liverpool University Hospitals NHS Foundation Trust, Liverpool, United Kingdom

[s] South Tees Hospitals NHS Foundation Trust, United Kingdom

[t] Imperial College Healthcare NHS Trust, London, United Kingdom

[u] Medway NHS Foundation Trust, United Kingdom

[v] University College London Hospitals NHS Foundation Trust, London, United Kingdom

[w] Roslin Institute, University of Edinburgh, Edinburgh, United Kingdom

[x] Centre for Musculoskeletal Research, University of Manchester, Manchester, United Kingdom

[y] NIHR Southampton Clinical Research Facility and Biomedical Research Centre, University Hospital Southampton NHS Foundation Trust and University of Southampton, Southampton, United Kingdom

[z] Faculty for Informatics and Data Science, University of Regensburg, Germany

[za] MRC Biostatistics Unit, University of Cambridge, UK

[zb] Radcliffe Deparment of Medicine, University of Oxford, United Kingdom

[zc] School of Medicine, University of Nottingham, Nottingham, United Kingdom

[zd] National Perinatal Epidemiology Unit, University of Oxford, United Kingdom

[ze] Respiratory Medicine Department, Nottingham University Hospitals NHS Trust, Nottingham, United Kingdom

[zf] Indian Council of Medical Research, New Delhi, India

[zg] School of Cellular and Molecular Medicine, University of Bristol, Bristol, United Kingdom

[zh] Intensive Care National Audit & Research Centre, London, United Kingdom

[zi] NIHR Oxford Biomedical Research Centre, Oxford University Hospitals NHS Foundation Trust, Oxford, United Kingdom

*,[†] equal contribution

**Steering Committee**

*Co-Chief Investigators* PW Horby, MJ Landray, *Members* JK Baillie, M Buch, L Chappell (until August 2021), J Day, SN Faust, R Haynes, T Jaki, K Jeffery, E Juszczak, M Knight (from August 2021), WS Lim, M Mafham, A Montgomery, A Mukherjee, A Mumford, K Rowan, G Thwaites.

**International Committee**

*Chair* DV Dung *Regional Lead Investigators* G Thwaites, J Day *Independent members* NN Quang, Prof. Binh, E Burhan, B Alisjahbana, J Koirala, S Basnet *Other members* E Kestelyn, B Basnyat, P Gyanwali, RL Hamers, P Horby

**Data Monitoring Committee**

P Sandercock (chair), J Darbyshire, D DeMets, R Fowler, D Lalloo, M Munavvar (from January 2021), I Roberts (until December 2020), A Warris (from March 2021), J Wittes *Non-voting statisticians* J Emberson, N Staplin.

**RECOVERY Trial Central Coordinating Office**

*Co-Chief Investigators* P Horby, MJ Landray; *Clinical Trial Unit Lead* R Haynes; *Trial management* A Cradduck-Bamford (coordinator), J Barton, A Basoglu, R Brown, W Brudlo, E Denis, L Fletcher, S Howard, S Musini, K Taylor; *Programming and validation* G Cui, B Goodenough, A King, M Lay, D Murray, W Stevens, K Wallendszus, R Welsh; *Data linkage* C Crichton, J Davies, R Goldacre, C Harper, F Knight, M Mafham, M Nunn, H Salih, J Welch; *Clinical support* M Campbell, G Pessoa-Amorim, L Peto, M Zayed; *Quality assurance* J Wiles; *Statistics* J Emberson, E Juszczak, E Spata, N Staplin; *Communications* G Bagley, S Cameron, S Chamberlain, B Farrell, H Freeman, A Kennedy, A Whitehouse, S Wilkinson, C Wood; *Evidence synthesis (Vascular Overviews Group)* C Reith (coordinator) K Davies, H Halls, L Holland, A Roddick, R Truell, K Wilson; *Administrative support* L Howie, M Lunn, P Rodgers

**RECOVERY Trial Regional Coordinating Centres**

Vietnam: *Oxford University Clinical Research Unit* G Thwaites, C Thwaites, E Kestelyn, NTH Thuong, TB Huyen, NT Thao, NB Tran, NTT Nguyen, NTP Dung, NTH Quyen, VTK Thi, NTH Trang, R van Doorn, HX Nam, PTT Hien, NY Nhi, NH Khanh

Nepal: *Nepal Health Research Council* P Gyanwali, M Dhimal, S Pant *Oxford University Clinical Research Unit* B Basnyat, A Karkey, S Rijal, S Shrestha

Indonesia: *Oxford University Clinical Research Unit Indonesia* RL Hamers, K Baird, K Puspatriani, M Rahardjani, A Rimainar, F Wulandari *Faculty of Medicine Universitas Indonesia* EJ Nelwan

India: *Indian Council for Medical Research* A Mukherjee, JJ Cherian, G Kumar

South Africa: *Wits Health Consortium* J Nel, H Rees

Ghana: *Kumasi Center for Collaborative Research in Tropic Medicine* J Amuasi, O Maiga, J Bonney, E Matey, A Afum-Adjei Awuah, H Hasford, A Amoako Adusei

Gambia: *MRC Unit The Gambia at LSHTM* B Nadjm, A Rocca, U D'Alessandro, E Usuf, M Bittaye, C Roberts, A Jagne

**UK National Institute for Health Research Clinical Research Network**

*Coordinating Centre* A Barnard, J Beety, C Birch, M Brend, E Chambers, L Chappell, S Crawshaw, C Drake, H Duckles-Leech, J Graham, T Harman, H Harper, S Lock, K Lomme, N McMillan, I Nickson, U Ohia, E OKell, V Poustie**,** S Sam, P Sharratt, J Sheffield, H Slade, W Van't Hoff, S Walker, J Williamson; *Urgent Public Health Clinical Links* A De Soyza, P Dimitri, SN Faust, N Lemoine, J Minton; *East Midlands* K Gilmour, K Pearson *Eastern* C Armah, D Campbell, H Cate, A Priest, E Thomas, R Usher; *North East & North Cumbria* G Johnson, M Logan, S Pratt, A Price, K Shirley, E Walton, P Williams, F Yelnoorkar; *Kent, Surrey & Sussex* J Hanson, H Membrey, L Gill, A Oliver; *North West London* S Das, S Murphy, M Sutu; *Greater Manchester* J Collins, H Monaghan, A Unsworth, S Beddows; *North West Coast* K Barker-Williams, S Dowling, K Gibbons, K Pine; *North Thames* A Asghar, P Aubrey, D Beaumont-Jewell, K Donaldson, T Skinner; *South London* J Luo, N Mguni, N Muzengi, R Pleass, E Wayman; *South West Peninsula* A Coe, J Hicks, M Hough, C Levett, A Potter, J Taylor; *Thames Valley and South Midlands* M Dolman, L Gerdes, C Hall, T Lockett, D Porter *Wessex* J Bartholomew, L Dowden, C Rook, J Walters; *West of England* E Denton, H Tinkler; *Yorkshire & Humber* A Alexander, H Campbell, K Chapman, A Hall, A Rodgers; *West Midlands* P Boyle, M Brookes, C Callens, H Duffy, C Green, K Hampshire, S Harrison, J Kirk, M Naz, L Porter, P Ryan, J Shenton, J Warmington; *Devolved nations* M Amezaga, P Dicks, J Goodwin, H Hodgson, S Jackson, M Odam, D Williamson.

**Paediatric working group**

SN Faust (coordinator), A Bamford, S Bandi, J Bernatoniene, K Cathie, P Dmitri, S Drysdale, M Emonts, J Evans, A Finn, P Fleming, J Furness, C Gale, R Haynes, CE Jones, E Juszczak, D Jyotish, D Kelly, C Murray, N Pathan, L Pollock, A Ramanan, A Riordan, C Roehr, M Wan, E Whittaker.

**Obstetric working group**

M Knight (coordinator), K Hodson, S Pavord, C Williamson.

#### Clinical support

*Clinical Trial Service Unit Out of Hours clinical support* L Bowman, F Chen, R Clarke, M Goonasekara, R Haynes, W Herrington, P Judge, M Mafham, K Mayne, S Ng, D Preiss, C Reith, E Sammons, D Zhu.

#### Health records

*NHS DigiTrials, Southport* H Pinches, P Bowker, V Byrne-Watts, G Chapman, G Coleman, J Gray, A Rees, MJ Landray, M Mafham, N Mather, T Denwood; *Intensive Care National Audit & Research Centre, London* D Harrison; *National Records of Scotland* G Turner; *Public Health Scotland* J Bruce; *SAIL Databank, University of Swansea* C Arkley, S Rees.

#### Local Clinical Centre RECOVERY trial staff

(listed in descending order of the number of patients randomised per site)

**Milton Keynes University Hospital NHS Foundation Trust** R Stewart (PI), J Alin, L Anguvaa, J Bae, G Bega, S Bowman, A Chakraborty, E Clare, S Fox, S Franklin, S George, L How, M Kennedy, J Mead, L Mew, D Mital, L Moran, E Mwaura, M Nathvani, A Rose, D Scaletta, S Shah, L Siamia, O Spring, S Sutherland, F Teasdale, S Velankar, L Wren, F Wright.

**Leeds Teaching Hospitals NHS Trust** D Ghosh (PI), S Ahmed, A Ashworth, N Balatoni, M Baum, L Bonney, J Calderwood, E Carter, S Charlton, J Clarke, C Coupland, M Crow, C Favager, J Glossop, J Hemingway, S Hemphill, K Holliday, A Humphries, S James, K Johnson, A Jones, M Kacar, K Khokhar, P Lewthwaite, A Marcyniuk, G Martin, F McGill, J Minton, D Mistry, J Murira, Z Mustufvi, S O'Riordan, K Robinson, G Saalmink, R Saman (Associate PI), D Singh, B Staniforth, S Straw, A Westwood.

**University Hospitals Bristol and Weston NHS Foundation Trust** G Hamilton (PI), N Blencowe (Co-PI), E Stratton (Co-PI), M Abraham, D Adams, B Al-Ramadhani, B Amit, A Archer (Associate PI), G Asher (Associate PI), G Aziz, A Balcombe, K Bateman, M Baxter, L Beacham, K Beard, K Belfield (Associate PI), N Bell, M Beresford, J Bernatoniene, A Bhat, D Bhojwani, S Biggs, C Blair, J Blazeby, K Bobruk, S Brooks, N Brown, L Buckley, P Butler, A Cannon, C Caws, E Chakkarapani, K Chatar, D Chatterton, B Chivima, E Clark, C Clemente de la Torre, K Cobain, H Cooke, D Cotterill, E Courtney, S Cowman, K Coy, H Crosby, K Curtis, P Davis, O Drewett, K Druryk, R Duncan, H Dymond, K Edgerley, M Ekoi, M Elokl, B Evans, T Farmery, N Fineman, A Finn, L Gamble, F Garty, B Gibbison, L Gourbault, D Grant, K Gregory, M Griffin, R Groome, L Gurung, V Haile, M Hamdollah-Zadeh, A Hannington, R Harrison, J Heywood, A Hindmarsh, N Holling, C Horrobin, R Houlihan, J Hrycaiczuk, H Hudson, K Hurley, J Iqbal, R Jarvis, B Jeffs, A Jones, R E Jones, E King-Oakley, E Kirkham, L Kirkpatrick, R Kumar, M Kurdy, A Lagnado, S Lang, L Leandro, H Legge, F Loro, A Low, H Martin, J Mayer, T Mayo, L McCullagh, G McMahon, L Millett, K Millington, J Mok, J Moon, L Morgan, S Mulligan, L Murray, T Nandwani, C O'Donovan, E Payne, C Penman, M Pezard-Snell, J Pickard, M Pitchford, C Plumptre, D Putensen, A Ramanan, J Ramirez, S Ratcliffe, N Redman, E Robbins, V Roberts, J Robinson, M Roderick, S Scattergood, A Schadenberg, E Schofield, R Sheppeard, C Shioi, J Shurlock, D Simpson, P Singhal, A Skorko, B Smart, N Smith, R Squires (Associate PI), V Stefania, C Stewart, M Stuttard, P Sugden, S Sundar, C Swanson-Low, T Swart, E Swift, A Tate, M Thake, K Thompson, M Trevelyan, K Turner, S Turner, A Tyer, S Vergnano, R Vincent, R Ward, A White, S Wilkinson, J Williams, S Williams, J Willis, H Winter, Z Woodward, L Woollen, R Wright, A Younes Ibrahim.

**Calderdale and Huddersfield NHS Foundation Trust** P Desai (PI), A Abbott, K Abouelela, U Akudi, D Appleyard, L-A Bayo, D Bromley, N Chambers, M Collins, S Dale, L Gledhill, J Goddard, J Greig, K Hallas, K Hanson, K Holroyd, M Home, D Kelly, A Maharajh, L Matapure, S Mellor, E Merwaha (Associate PI), M Prior-Ong, K Rajalingam, H Riley, M Robinson, C Rourke, K Sandhu, K Schwarz, N Scriven, L Shaw, L Terrett, M Thompson (Associate PI), G Turner, M Usher, A Wilson, T Wood.

**Hull University Teaching Hospitals NHS Trust** N Easom (PI), K Adams, K Ahmed, D Allsup, G Barlow, R Barton, J Batac, D Clark, L Cullen, K Drury, A Frygier, S Gribben, P Gunasekera, A Harvey, M Hayes, M Ivan, S Khan, M Kolodziej, P Lillie, V Lowthorpe, V Mathew, S Mongolu, A Morrow, I Muazzam, D Norton, K Nu, R Patmore, T Perinpanathan, C Philbey, B Pickwell-Smith, D Purchase, S Rastogi, A Richards, L Rollins, A Samson, T Sathyapalan, Y See (Associate PI), L Sherris, K Sivakumar, D Smith, T Taynton (Associate PI), G Walker, A Wolstencroft, H Yates.

**Mid Cheshire Hospitals NHS Foundation Trust** J Majumdar (PI), T Adeyemo, K Best, G Bridgwood, R Broadhurst, C Brockelsby, T Brockley, J Brown, R Bujazia, A Burton, S Clarke, J Cremona, C Dixon, S Dowson, H Drogan, F Duncan, M Emms, H Farooq, D Fullerton, N G, C Gabriel, S Hammersley, R Hum, T Jones, S Kay, E Kelly, M Kidd, D Lees, R Lowsby, D Maren, D Maseda, E Matovu, K McIntyre, H Moulton, K Nourein, K Pagett, A Ritchings, S Smith, J Taylor, K Thomas, K Turbitt, M Williams, S Yasmin.

**Liverpool University Hospitals NHS Foundation Trust** P Hine (PI), P Albert (Co-PI), S Todd (Co-PI), I Welters (Co-PI), D Wootton (Co-PI), M Ahmed, R Ahmed, A Al Balushi, M Anderson, R Anderson, Z Ashfak, A Atomode, R Ball, P Banks, D Barr, J Bassett, A Bennett, H Bond, A Bracken, T Brankin-Frisby (Associate PI), G Bretland, P Brinksman, M Brodsky, J Brown, H Burhan, C Burston, J Byrne, F Carlin, S Casey, L Chambers, D Coey, T Cross, J Cruise, J Currie, S D'Souza, L Dobbie, R Downey, A Du Thinh, G Duncan, I Duru, J Early, T Evans, K Fenlon, J Fernandez Roman, I Fordham, H Frankland, S Glynn, J Goodall, S Gould, A Gureviciute, J Hackett, K Haigh, M Hamilton, L Hampson, A Hanson, M Harrison, L Hawker, P Hazenberg, D Heath, S Hicks, S Hope, M Howard, K Hunter, T Ingram, A Islim, K Janes, B Johnston, S Karmali, S Kavanagh, L Keogan, W Khan, S King, K Krasauskas, J Lewis, M Lofthouse, P Lopez, C Lowe, Z Mahmood, F Malein, K Martin, A Mediana, L Melling, Z Mellor, P Merron, B Metcalfe, M Middleton, K Monsell, A Morgan, H Murphy, N Nicholas, A Nuttall, L Oliver, R Osanlou, J Parsons, L Pauls, L Pilling, R Price-Eland, C Prince, S Pringle, E Richardson, L Rigby, M Riley, A Rowe, E Rybka, M Samuel, D Scanlon, J Sedano, D Shaw, F Shiham, C Smith (Associate PI), S Stevenson, A Stockdale, J Tempany, P Thu-Ta, C Toohey, I Turner-bone, S Victoria, A Waite, E Wasson, A Watkin, R Watson, V Waugh, R Westhead, L Wilding, K Williams, A Wood, A Yeoh, D Zeinali.

**South Tees Hospitals NHS Foundation Trust** D Chadwick (PI), A Aboagye-Odei, S Armstrong, D Athorne, A Awadelkareem, M Branch, J Brolly, S Brown, J Cheaveau, H Chen, Y Chua, N Cunningham, M Dafalla, S Davies, J Dodds, S Dorgan, D Dunn, P Dunn, M Elsayed, E Hammond, P Harper, H Harwood, K Hebbron, F Hunt, A Kala Bhushan, P Lambert, C Lawrence, D Leaning, T Linn, T Manders, B McCarron, N Miller-Biot, C Milne, W Mohammad, M Mollet, J Mulcahy, A Murad, S Ooi, M Owston, J Potts, C Proctor, S Puliyakkadi, B Puvaneswaran, S Rao, R Raw (Associate PI), M Seelarbokus, P Singh, V Srirathan, L Swithenbank, A Szekeres, L Thompson, H Wardy, L Wiblin, J Widdrington, J Williams, P Winder, C Wroe.

**Imperial College Healthcare NHS Trust** G Cooke (PI), L Young (Co-PI), O Adedeji, E Adewunmi, Z Al-Saadi, R Ashworth, J Barnacle, N Bohnacker, A Cann, F Cheng, J Clark, S Cooray, S Darnell, A Daunt (Associate PI), V Dave, A D'Mello, L Evison, S Fernandez Lopez, F Fitzgerald, C Gale, M Gibani, S Hamilton, S Hunter, A Jimenez Gil, S Johal, B Jones, A Kountourgioti, J Labao, V Latham, N Madeja, S Mashate, C Matthews, H McLachlan, A Mehar,

J Millard (Associate PI), M Molina, A Perry, H Rafferty, S Rey, S Ryder, R Shah, E Sidebotham, R Thomas, D Thornton, J Tuff, E Whittaker, C Wignall, P Wilding, C Wong, T Yates (Associate PI), C Yu.

**Medway NHS Foundation Trust** R Sarkar (PI), K Abernethy (Associate PI), C Adams, L Adams, A Addo, F Aliyuda, S Archer, A Arya, E Attubato, F Babatunde, M Bachour, P Balasingam, A Bhandari, F Brokke, R Chauhan, V Chawla, R Chineka, A Davis, N Edmond, M Elbeshy, C Ezenduka, S Ferron, C Gnanalingam, D Gotham, M Hollands, M Iqbal, A Jamal, B Josiah, S Kidney, M Kim, K Koukou, T Kyere-Diabour, L Leach, A Liao, A Maheswaran, M Mansour, N Miah, J Morilla, L Naglik, K Naicker, Z Nurgat, S Rai, I Redknap, Z Rehman, A Ryan, Y Samuel, A Shaibu, P Soor, R Squires, W Stagg, W Ul Hassan, P Vankayalapati, E Vyras, A Williams, J Wood, N Zuhra (Associate PI).

**University College London Hospitals NHS Foundation Trust** H Esmail (PI), R Heyderman (Co-PI), V Johnston (Co-PI), D Moore (Co-PI), A Andrews, F Beynon, P Bodalia, I Bokobza, X Chan, Z Chaudhry, C Chung, J Cohen, D Crilly, S Eisen, N Fard, J Gahir, L Germain, J Glanville, R Gupta, N Lack, A Last, A Luintel, M Merida Morillas, B Nadjm, N Platt, T Rampling, H Rickman, S Rokadiya, S Roy, A Samson, R Shortman, I Skorupinska, M Skorupinska, J Spillane, C Thakker, L Wellings.

**Portsmouth Hospitals NHS Trust** T Brown (PI), J Andrews, R Baker, M Baker-Moffatt, A Bamgboye, D Barnes, S Baryschpolec, V Bataduwaarachchi, S Begum, L Bell, H Blackman, J Borbone, N Borman, P Braga Sardo, L Brimfield, M Broadway, F Brogan, R Bungue-Tuble, K Burrows, A Chauhan, E Cowan, M Czekaj, Z Daly, A Darbyshire, A Das, M Davey, M David, J Denham, R Duhoky, C Edwards, S Elliott, H Evison, L Fox, Z Garner, I Gedge, B Giles, S Glaysher, S Gosling, A Gribbin, J Hale, Y Harrington-Davies, L Hawes, J Hayward, A Hicks, A Holmes, S Howe, H Htet, B Jones, C Lameirinhas, E Lavington, B Longhurst, L Lopes, M Mamman, J Marshall, S McCready, L Milne, C Minnis, M Moon, J Mouland, L Murray, J Ouyang, C Pereira Dias Alves, M Rason, L Richmond, D Rodgers, S Rose, M Rowley, M Rutgers, T Scorrer, L Shayler, R Sievers, K Siggens, S Smith, C Stemp, N Szarazova, E Taylor, R Thornton, A Tiller, C Turner, L Vinall, E Walmsley, M Wands, J Warren, L Watkins, M White, L Wiffen (Associate PI), J Winter, C Wong, K Wren, Y Yang.

**Sheffield Teaching Hospitals NHS Foundation Trust** P Collini (PI), A Ali, A Alli, S Anderson, A Ang, I Aslam, A Basran, J Belcher, G Benoy, S Bird, M Boothroyd, H Bowler, A Card, K Cawthorne, L Chapman, K Chin, A Clarke, S Clenton, V Clubb, L Cockayne, D Cohen, J Cole, H Colton, A Condliffe, R Condliffe, A Creaser-Myers, M Cribb, S Curran, S Darby, T Darton, D de Fonseka, T de Silva, A Dunn, C Durojaiye, P Easton, S Eggleston, A Ellwood, A Emery, E Ferriman, H Foot, A Ford, R Foster, Z Gabriel, V Goodall, J Greig, N Guthrine, J Hall, H Hanratty, S Hardman, K Harrington, J Hau, E Headon, C Holden, B Holroyd-Hind, K Housely, A Howell, L Hunt, E Hurditch, J Hurdman, Z Hussein, F Ilyas, J Jackman, Y Jackson, C Jarman, F Kibutu, B Kilner, T Kitching, S Lassa, R Lawson, C Lee, R Lenagh, L Lewis, N Lewis, T Locke, H Luke, A Lye, I Macharia, L Mair, G Margabanthu, P May, A Mbuyisha, J McNeill, S Megson, J Meirill, J Meiring, J Middle, J Middleton, L Milner, P Morris, D Mosby, A Nazir, H Newell, T Newman, L Nwafor, V Palissery, V Parkash, L Passby, R Payne, I Pernicova, P Phillips, M Plowright, A Raithatha, G Rana, P Ravencroft, S Renshaw, A Rothman, M Roycroft, D Sammut, S Sammut, R Sellars, S Sherwin, J Sidebottom, P Simpson, L Smart, M Sterrenburg, B Stone, M Surtees, A Telfer, B Thamu, R Thompson, Y Thompson, N Tinker, H Trower, K Turner, M Ul Haq, N Vethanayagam, P Wade, J Wadsley, S Walker, J G R Watson, J Wesonga, R West, R Whitham, T Williams, J Willson, A Wilson, I Wilson, T Zak, E Zincone, STH CRF COVID Nursing team.

**Stockport NHS Foundation Trust** R Stanciu (PI), M Afridi, M Al Dakhola, M Alafifi, S Bennett, L Brown, C Cooper, A Davison, D Eleanor, J Farthing, A Ferrera, S Gandhi, L Gomez, Z Hassan, P Haywood, C Heal, H Jackson, J Johnston, A Lloyd, S McCaughey, R Mills, R Owen,

A Pemberton, F Rahim, H Robinson, N Sadiq, R Samlal, V Subramanian, D Suresh, D Tudor, H Wieringa, I Wright, B Xia.

**NHS Fife** D Dhasmana (PI), F Adam, K Aniruddhan, J Boyd, N Bulteel, P Cochrane, I Fairbairn, S Finch, K Gray, L Hogg, S Iwanikiw, M Macmahon, P Marks, A McGregor, A Morrow, J Penman, J Pickles, J Ramsay, A Scullion, H Sheridan, M Simpson, D Sloan, C Stewart, J Tait, A Timmins, M Topping.

**Sukraraj Tropical and Infectious Disease Hospital, Nepal** M Rawal (PI), B Chalise, S Gyawali, A Gupta, Y Sapkota, S Paudel, J Khatri, S Pant, P Gyanwali, U Devkota, S Mandal, L Bhandari, K Paudel, N K.C., S Karki, A Chaudhary, L Chaulagain, R Kumar, R Jaiswal, M Bajracharya, S Gyawali, S Magar, J Neupane, G Shrestha, R Jaiswal, K Poudel.

**University Hospitals Birmingham NHS Foundation Trust** C Green (PI), T Whitehouse (Co-PI), I Ahmed, N Anderson, C Armstrong, A Bamford, H Bancroft, M Bates, M Bellamy, T Bellamy, C Bergin, K Bhandal, E Butler, M Carmody, N Cianci (Associate PI), K Clay (Associate PI), L Cooper, J Daglish, J Dasgin, A Desai, S Dhani, D Dosanjh, E Forster, J Gresty, E Grobovaite, N Haider, B Hopkins, D Hull, Y Hussain, A Kailey, M Lacson, M Lovell, D Lynch, C McGhee, C McNeill, F Moore, A Nilsson, J Nunnick, W Osborne (Associate PI), S Page, D Parekh, C Prest, K Price, V Price, M Sangombe, H Smith, I Storey, L Thrasyvoulou, K Tsakiridou, D Walsh, S Welch, H Willis, L Wood, J Woodford, G Wooldridge, C Zullo.

**Blackpool Teaching Hospitals NHS Foundation Trust** G Hardy (PI), A Abdullah (Associate PI), N Ahmed, O Assaf, A Barnett, A Beasley, L Benham, D Bennett, P Bradley, Z Bradshaw, M Brunton, T Capstick, M Caswell, V Cunliffe, J Cupitt, R Downes, L Elawamy, J Howard, C Jeffs, N John, N Latt, J Mason, R McDonald, A Mulla, E Mutema, J Navin, A Parker, A Potter, S Preston, N Slawson, E Stoddard, S Traynor, V Vasudevan, E Ward, S Warden, A Wignall, J Wilson, A Zmierczak.

**Cambridge University Hospitals NHS Foundation Trust** M Knolle (PI), E Gkrania-Klotsas (Co-PI), O Abani, R Bousfield (Associate PI), T Dymond, L Eadie, K Gajewska-Knapik, J Galloway, R Gore, K Leonard, T Mikolasch, H-P Mok, N Pathan, P Polgarova, J Sahota, K Sharrocks, R Swain.

**East Suffolk and North Essex NHS Foundation Trust** J Douse (PI), M Ramali (Co-PI), K Ahmed, S Alam, A Arumaithurai, B Atraskiewicz, J Bailey, I Balluz, D Beeby, S Bell, J Bloomfield, S Blows, N Broughton, C Buckman, M Burton, C Calver, J Campbell, P Carroll, C Chabo, R Chalmers, K Cheung, M Chowdhury, G Christoforou, K Cooke, N Deole, T H Dinh (Associate PI), C Driscoll, J Dulay, S Finbow, I Floodgate, R Francis, C Galloway, E Galloway, M Garfield, A Ghosh, G Gray, P Greenfield, A Gribble, M Gunawardena, M Hadjiandreou, H Hewer, M Hossain, R Howard-Griffin, K Howlett, C Huah, N Innes, V Inpadhas, A Islam, L James, Z Jiao, K Johannessen, J Kathirgamachelvam, M Khan Tharin, V Kushakovsky, S Lee, R Lewis, R Lloyd, LH Lui, L Mabelin, P Mallett, D Morris, S Nallapareddy, S Nishat, H C Ooi (Associate PI), R Osagie, H Patel, AK Phyo, M Pretorius, B Purewal, F Ramali, H Rawlins, P Ridley, V Rivers, J Rosier, E Rushforth, S Sethi, A Sharma, S Sharma, A Sheik, J Shoote, M Shuvo, R Skelly, R Smith, R Smith, R Sreenivasan, P Tovey, A Turner, K Turner, K Vithian, I Weichert, W Win.

**Cardiff & Vale University LHB** M Wise (PI), J Underwood (Co-PI), A Balan, B Basker, S Bird, Z Boult, V Britten, L Broad, H Cendl, M Chakraborty, J Cole, E Davies, M Edgar, M Edwards, N Elashhar, S Elliott, J Evans, J Evans, M Evans, E Evans, C Fegan, M Forester, J Forton, S Frayling, S Gage, F Greaves, S Harrhy, M Haynes, A Heavens, H Hill, Z Hilton, L Jones, A Kelly, L Knibbs, N Krishnapalli, D Lau, J May, E McGough, A McQueen, J Milner, M Morgan, F Muhammad, R Norman, K Nyland, C Oliver, K Paradowski, M Patal, B Phillips, M Potdar, K

Rahilly, D Rigler, C Robinson, S Scourfield, A Semmens, M Starr, J Stevens, S Struik, E Thomas, R Thomas-Turner, G Thueux, G Williams, J Williams, M Williams, S Zaher.

**West Suffolk NHS Foundation Trust** M Moody (PI), S Barkha, H Cockerill, K Durrant, J Godden, J Kellett, T Murray, A Saraswatula, A Williams, L Wood.

**Great Western Hospitals NHS Foundation Trust** A Kerry (PI), A Aldesouki, A Azeem, V Barlow, A Beale, T Benn, S Bhandari, A Brooks, C Browne, J Butler (Associate PI), J Callaghan, B Chandrasekaran, N Clark, L Davies, R Davies, T Elias, J Evans, D Finch, S Flockhart, P Foley, E Fowler, E Fraile, G Gowda, J Gregory, C Hunt, A Ipe, A Jaffery, M Juniper, S Khan, I Laing-Faiers, H Langton, G Laura, C Lewis-Clarke, J Lodge, C Mackinlay, P Mappa, A Maxwell, L McCafferty, W Mears, E Mousley, T Novak, C Novis, L Pannell, S Peglar, A Pereira, I Ponte Bettencourt dos Reis, E Price, A Quayle, Q Qurratulain, M Ryder, S Small, H Smith, C Strait, E Stratton, M Tinkler, J Ugoji, A van der Meer, L von Oven, A Waldron, R Waller, M Watters, S White, L Whittam, T Wiliams, Z Xia, K Yein, V Zinyemba, G Zubikarai.

**Dorset County Hospital NHS Foundation Trust** J Chambers (PI), F Best, J Birch, L Bough, S Caddy, A Cave, J Chambers, J Colton, J Graves, S Horton, J Rees, R Thomas, W Verling, P Williams, S Williams, B Winter-Goodwin, S Wiseman, D Wixted.

**North Middlesex University Hospital NHS Trust** J Moreno-Cuesta (PI), A Dhariwal (Co-PI), S Jain (Co-PI), T Amin, D Bakthavatsalam, P Brooks, M Farah, A Govind, A Haldeos, K Leigh-Ellis, T Light, S Rokadiya, C Vansomeren, R Vincent, L Walker.

**Buckinghamshire Healthcare NHS Trust** N Wong (PI), J Abrams, A Alkhudhayri, N Aung, A Baldwin, O Bannister, J Barker, H Beddall, H Blamey, E Chan, J Chaplin, B Chisnall, C Cleaver, M Corredera, S Crotty, H Cui, B Davies, P Dey, L Downs, S Gettings, B Hammans, S Jackman, P Jenkins, M Kononen, S Kudsk-Iversen, A Kudzinskas, M Laurenson, R Mancinelli, J Mandeville, K Manso, B Marks, S McLure, O Michalec, E Morgan-Smith, A Ngumo, H Noe (Associate PI), R Oxlade, A Parekh (Associate PI), V Pradhan, M Rahman, C Robertson, R Rule, S Shah, H Smith, J Tebbutt, N Vella, M Veres, A Watson, R West, L Western, M Zammit-Mangion, M Zia.

**Countess Of Chester Hospital NHS Foundation Trust** S Scott (PI), A Abdelaziz, A Abdulakeem, M Abouibrahim, A Adeyanju, A Aditya, M Ahmed, S Ahmed, S H Ahmed, A Ajibode, L Alomari, F Ashworth, A Asrar, E Austin, A Awan, P Bamford, A Banerjee, A Barclay, L Barker, K Barker-Williams, T Barnes, W Barnsley, H Batty, F Bawa, G Beech, J Bellamu, K Bennion, I Benton, J Bhanich Supapol, S Billingham, K Blythe, T Bowker, S Brearey, S Brigham, V Brooker, A Brown, C Burchett, M Burgess, R Cade, F Cameron, R Cannan, M Cardwell, K Cawley, N Chavasse, L Cheater, Z Cheng, K Clark, R Clarke, R Clifford, E Cole, C Cook, C Cotton, S Cremona, D Curran, H Dallow, J Dangerfield, A Davidson, A Davies, A Davies, K Davis, M Devonport, Y Doi, E Doke, C Dragos, H Duffy, L Eagles, M Edwards, L Ellerton, R Ellis, H Elmasry, E Entwistle, A Fallow, M Faulkner, E Ferrelly, K Flewitt, C Foden, D Font, L Fuller, R Gale, L Gamble, M Gardiner, K Gillespie, L Goodfellow, M Grant, S Griffiths, Y Griffiths, J Grounds, V Guratsky, L Hartley, P Hayle, C Haylett, B Hemmings, N Henry, P Hettiarachchi, L Hill, H Hodgkins, S Holt, S Hunt, M Ibraheim, M Irshad, M Iyer, Z Jamal, R James, A Jayachandran, R Jayaram, H Jeffrey, A Johari, O Johnson, C Jones, C Jones, E Jones, R Jones, A Kazeem, N Kearsley, K Kouranloo, A Kumar, S Leason, N Lightfoot, B Lim, D Llanera, R Llewellyn, S Loh, E London, C Lonsdale, T Maheswaran, G Markham, E Martin, P Maskell, A Mather, A Mathew, I Mazen, M McBuigan, J McBurney, M McCarthy, R McEwen, M McGuigan, E Meeks, G Metcalf-Cuenca, L Michalca-Mason, S Middleton, L Mihalca-Mason, D Mock Font, A Moore, L Morris, R Mufti, K Murray, I Nagra, A Nallasivan, M Nayyar, F Naz, R Nelson, R Nicholas, C Oakley, E Okpo (Associate PI), F Olonipile, C O'Neill, C Ord, J Osman, E Owens, S Pajak, S Pal, J Parker, H Parry, S Pearson, K Penney, E Perritt, D

Phillips, C Pickering, A Ponnuswamy, S Powell, V Prescott, J Prince, E Pye, S Rahman, P Rath, S Read, J Reed, A Reid (Associate PI), J Roberts, N Robin, R Robinson, M Saladi, D Scholes, A Scott, S Scott, H Shabbir, M Shah, G Sheppard, F Shiham, T Spencer, J Spriggs, C Steele, Z Syrimi, L Theocharidou, M Thevendra, C Thorne, A Timmis, S Tomlin, T Trussell, O Vass, B Vlies, A Walters, L Walton, K Webb, T Webster, S Wheaver, D Whitmore, A Williams, C Wyatt, W Yap, J Ye, C Yeung, E Young, L Zammit, E Zin.

**Epsom and St Helier University Hospitals NHS Trust** S Winn (PI), J Evans (Co-PI), F Abbas, S Acheampong, S Ahamed Sadiq, A Aldana, B Al-Hakim, A Alipustain, P Allen, G Azzopardi, E Balogh, Z Baxter, C Bond, C Cheeld, R Chicano, I Chukwulobelu, N Colbeck, R Dogra, E Doherty, A Elradi, J Emberton, L Evans, R Ganapathy, N Hamilton, J Handford, M Haque, J Hayes, R Hayre, L Ilves, C Jagadish, S Jain, K Jian, A Johnson, L Johnson, B Kim, J Kotecha, A Kundu, D Langer, E Lester, A Lunia, Y Mashhoudi, K Mathias, T Medveczky, F Mellor, G Mic, S Moore, M Morgan, O Mushtaq, S Nafees, A Oluwole-Ojo, V Palagiri Sai, M Phanish, H Rahimi, S Ramanna, A Ramnarine, J Ratoff, M Ridha, S Rozewicz, T Samuel, G Saxena, S Selvendran, B Shah, S Shahnazari, R Shail, A Sharif, S Singh, S Somalanka, R Suckling, P Swift, L Tarrant, V Tyagi, N Vilimiene, P Virdee, R Wake, L Walsh.

**RS UNAIR, Surabaya** N Nasronudin (PI), C Nugroho, R Prananingtias, E Puji Lestari, B Rachman, A Rosyid, F Utomo, A Veterini.

**Royal Free London NHS Foundation Trust** T Mahungu (PI), H Tahir (Co-PI), A Abdul, R Abdul-Kadir, H Aboelela, M Al-Khalil, N Allan, I Alshaer, M Anderson, M Araujo, G Badhan, A Bakhai, S Bhagani, B Bobie, A Brraka, B Caplin, A Carroll, A Carroll, H Century, E Cheung, D Cohen (Associate PI), O Coker, D Collier, V Conteh, N Cooper, J Crause, N Davies, R Davies, V Deelchand, M Dosani, L Ehiorobo, C Ellis, G Ferenando, J Franklin, P Gardiner, F Geele, J Gosai, N Handzewniak, E Hanison, S Hanson, N Holdhof, H Hughes, C Jack, C Jarvis, V Jennings, H Koo, V Krishnamurthy, A Kurani (Associate PI), Z Ladan, L Lamb, A Lang, V Le, S Lee, S Lo, A Luintel, A Maharajh, H Mahdi, T Majekdunmi, D Matila, S Melander, F Mellor, A Molloy, R Moores, J Morales, G Moray, A Nandani, S Nasa, S O'Farrell, A Oomatia, A Osadcow, J Osei-Bobie, G Pakou, P Patel, C Patterson (Associate PI), E Pyart, E Quek, S Rabinowicz, T Rampling, R Rankhelawon, A Rodger, A Scobie, S Sharma, C Singh, S Sithiravel, T Sobande, P Talbot, P Taribagil, S Veerasamy, G Wallis, J Whiteley, E Witele, A Wong, E Woodford, N Yaqoob.

**Wirral University Teaching Hospital NHS Foundation Trust** A Wight (PI), F Adeoye, L Bailey, E Barker, S Bokhari, S Brownlee, A Bull, J Corless, C Denmade, N Ellard, A Farrell, A Hufton, R Jacob, A Jones, K Jones, H Kerss, J McEntee, N Morris, R Myagerimath, T Newcombe, M Parsonage, H Peake, D Pearson, R Penfold, S Rath, A Reddington, R Saunders, A Sharp, B Spencer, A Suliman, S Sutton, H Tan, D Tarpey, L Thompson, T Thornton, E Twohey, D Wagstaff, Z Wahbi, G Wardere, S Williams.

**NHS Lothian: Western General Hospital** O Koch (PI), A Abu-Arafeh, E Allen, L Bagshaw, C Balmforth, R Barnes, A Barnett-Vanes, R Baruah, S Begg, S Blackley, M Braithwaite, G Clark, S Clifford, D Dockrell, M Evans, V Fancois, C Ferguson, S Ferguson, N Freeman, E Gaughan, E Godden, S Hainey, R Harrison, B Hastings, S Htwe, A Ju Wen Kwek, M Ke, O Lloyd, C Mackintosh, A MacRaild, W Mahmood, E Mahony, J McCrae, S Morris, C Mutch, S Nelson, K Nunn, D O Shea, I Page, M Perry, J Rhodes, N Rodgers, J Schafers (Associate PI), A Shepherd, G Soothill, S Stock, R Sutherland, A Tasiou, A Tufail, D Waters, R Weerakoon, T Wilkinson, R Woodfield (Associate PI), J Wubetu.

**University Hospitals Of Derby and Burton NHS Foundation Trust** T Bewick (PI), P Daniel (Co-PI), U Nanda (Co-PI), A Anada, P Basvi, G Bell, H Cox, C Downes, K English, A Fletcher, J Hampson, S Hathaway-Lees, M Hayman, N Jackson, A Matthews, C McDonald, S Ojha, L

Prince, J Radford, K Riches, G Robinson, A Sathyanarayanan, C Smith, E Starkey, J Usher, L Wilcox, L Wright, M Zawadzka.

**University Hospitals Of Morecambe Bay NHS Foundation Trust** S Bari (PI), S Achieng, M Al-Hayali, M Al-Jibury, K Allison, V Anu, C Bartlett, L Bartlett, M S Bhuiyan, S Bhuiyan, L Bishop, M Bukhari, K Burns, J Craig, A Davies, A Dent, J Dodd, A Dow, M El-Naggar, A Fielding, M Glover, M Gorst, J Han, C Hay, A Higham, O Igwe, I Irabor, S Kaprapina, J Keating, H Khalid, T Khan, R Kubaisi, L Lobosco, M Lwin, F Mahmood, P Mallinder, B Marwan, L McDougall, S Moorby, H Morris, K Ngwenya, O Onuoha, Z Patel, S Peters, D Power, J Ritchie, P Sarmiento, A Shams, S Sharma, K Simpson, H Spickett, C Stokes, M Tai, H Thatcher, C Till, A Varghese, H Williams, M Williams, T Win, F Wood, S Woodmansey.

**Wye Valley NHS Trust** I DuRand (PI), J Al-Fori, A Almahroos, J Annett, A Barclay, J Bartlett, J Birch, N Bray, A Carrasco, M Cohn, E Collins, K Crowley, A Davies, M Forkan, A Fratila, H Gashau, S Gayle, K Hammerton, A Hassan, A Hedges, R Homewood, Z Khan, S Maryosh, S Meyrick, L Moseley, B Mwale, L Myslivecek, A Phillips, J Porteous, M Qayum, P Ryan, A Salam, C Seagrave, F Suliman, A Talbot -Smith, S Turner, S Vaughan, E Wales, H Walker, J Woolley.

**Bedford Hospital NHS Trust** T Chapman (PI), I Nadeem (Co-PI), H Ahmed, A Amjad, N Amjad, A Anthony-Pillai, I Armata, R Arora, A Ashour, D Bagmane, V Bastion, R Bhanot, D Callum, K Chan, P Chrysostomou, C Currie, F De Santana Miranda, B Djeugam, C Donaldson, P Eskander, S Farnworth, F Farrukh, N Fatimah, C Fernando, R Field, K Gelly, L Grosu, A Haddad, M Hannun, T Hashimoto, K Hawes, E Hawley-Jones, M Hayathu, M Hikmat, B Jallow, C Jenkins, U Keke, U Khatana, I Koopmans, T Large, P Lemessy, H Lindsay-Clarke, E Lister, R Lorusso, G Lubimbi, W-O Makinde, A Mian, F Miranda, J Morris, N Nasheed, N Nathaniel, M Negmeldin, K Ng, C Ong, K Pandya, S Peerbhoy, M Penacerrada, J Prieto, Q Quratulain, S Rahama, M Revels, L Salih, J Sarella, Z Shaida, V Simbi, N Simon, B Small, M Smith, N Suresh, W Tan, E Thomas, S Trussell, U Vaghela, A Vaidya, J Valentine, O Walker, F Wang, J Wischhusen, R Wulandari, Y Yanagisawa, L Ylquimiche Melly.

**Hampshire Hospitals NHS Foundation Trust** A Goldsmith (PI), R Partridge (Co-PI), Y Abed El Khaleq, M Adamczyk, M Alvarez Corral, A Arias, E Bevan, J Chan, M Chapman, J Conyngham, E Cox, E Defever, A Edwards, C Fitton, D Griffin, A Heath, M Jordan, B King, E Levell, X Liu, J Martin, N Muchenje, M Mupudzi, A Nejad, A Prabhu, S Senra, F Stourton, R Thomas, B White (Associate PI), G Whitlingum, L Winckworth, C Wrey Brown, S Zagalo.

**St George's University Hospitals NHS Foundation Trust** T Bicanic (PI), T Harrison (Co-PI), Y Aceampong, A Adebiyi, M Ali, D Baramova, M Caudwell, J De Sousa, P Diwan, S Drysdale, L Hamzah, O Harrison, J Hayat, A Janmohamed, H Ju, A Khalil, A Lisboa, E Marler, M Mencias, J Millard, C Page, J Pang, K Patel, A Perry, A Rana, V Raspa, P Ribeiro, N Said, T Samakomva, A Seward, O Skelton, J Sousa, K Spears, A Sturdy, V Tavoukjian, J Texeira, S Tinashe, O Toffoletti, C Ward.

**The Newcastle Upon Tyne Hospitals NHS Foundation Trust** E Hunter (PI), R Agbeko, A Bailey, K Baker, A Barr, J Brolly, E Cameron, Q Campbell Hewson, R Capstick, J Cheaveau, I Clement, A De Soyza, C Duncan, M Emonts, A Fenn, S Francis, L Gardner, B Ghavami Kia, J Glover Bengtsson, A Greenhalgh, A Hanrath, H Hanson, C Hays, K Houghton, D Jerry, G Jones, S Kelly, A Kimber, N Lane, J Macfarlane, P McAlinden, I McCullagh, S McDonald, O Mohammed, P Nwajiugo, R Obukofe, J Parker, B Patel, A Patience, B Payne, R Percival, D Price, Z Razvi, N Rice, S Robson, A Sanchez Gonzalez, B Shillitoe, A Stanton, E Stephenson, N Trewick, S Tucker, G Waring, R Welton, S West, E Williams, E Wong, F Yelnoorkar.

**Maidstone and Tunbridge Wells NHS Trust** A Ross-Parker (PI), A Abbott, L Adams, S Anandappa, B Babiker, M Barbosa, G Chamberlain, K Cox, D Datta, M Davey, N Earl, F

Gallam, R Gowda, S Goyal, A Gupta, E Harlock, C Hart, E Hughes, S Husaini, E Hutchinson, A Keough, A Khan, S Kumar, R Nemane, L Orekoya, I Pamphlett, T Patel (Associate PI), C Pegg, P Rajagopalan, A Richards, S Sathianandan, R Seaman, S Siddavaram, O Solademi, M Szeto, P Tsang, V Uppal, A Waller, H Winwood.

**Mid Yorkshire Hospitals NHS Trust** B Sloan (PI), J Ashcroft, R Beckitt, P Blaxill, S Bond, S Boot, S Buckley, G Castle, E Clayton, N de Vere, A Dwarakanath, J Ellam, D Gomersall, S Gordon, C Hettiarachchi, C Hutsby, R Kousar´┐¢, K Lindley, A Major, A Metcalfe, S Oddy, A Poole, U Prasad, J Quinn, K Rajalingam, A Rose, L Slater, B Taylor, S Taylor, M Thirumaran.

**Salisbury NHS Foundation Trust** M Sinha (PI), K Ames, A Anthony, J Barr, A Barton, L Bell, G Chaplin, M Cook, S Diment, P Donnison, B Eapen, S Evans, R Fennelly, S Gray, H Harcourt, A Hawkins, I Jenkins, M Johns, T Jones, L Joy, V King, I Leadbitter, W Matimba-Mupaya, L Mattocks, R Mehta, H Morgan, A Rand, S Salisbury, K Seymour, S Strong-Sheldrake, C Thompson, F Trim, E Underhill.

**Sherwood Forest Hospitals NHS Foundation Trust** M Roberts (PI), L Allsop, K Amsha, K Bennett, T Brear, P Buckley, G Cox, N Downer, L Dunn, M Flynn, M Gill, C Goodwin, C Heeley, D Hodgson, L Holloway, M Holmes, R Holmes, J Hutchinson, R Johnson, S Kalsoom, J Kirk, E Langthorne, W Lovegrove, G Mabeza, A Molyneux, C Moulds, D Nash, Z Noor, J Rajeswary, T Sewell, S Shelton, H Shirt, K Slack, S Smith, N Thorpe, S Turner, B Valeria, I Wynter, M Yanney, I Ynter.

**Shrewsbury and Telford Hospital NHS Trust** J Moon (PI), R Baldwin-Jones, N Biswas, A Bowes, H Button, E Cale, M Carnahan, E Crawford, E Damm (Associate PI), S Deshpande, D Donaldson, C Fenton, R Heinink, S Hester, Y Hussain, K Ibison, M Ibrahim, S Islam (Associate PI), J Jones, S Jose, A Makan, H Millward, J Moorcroft, N Motherwell, H Moudgil, C Mowatt, J Nixon, S Pajak, S Passey (Associate PI), S Pickstock, L Price, M Rees, E Roddy, N Rowe, N Schunke, K Srinivasan, A Stephens, J Stickley, M Tadros, H Tivenan, G Wood.

**University Hospital Southampton NHS Foundation Trust** S Fletcher (PI), M Avery, M Bader, A Baltmr, S Bartolmeu-Pires, S Beeby, J Bigg, C Bolger, D Brooks, H Burke, K Cathie, S Chabane, B Clancy, M Coleman, A Collier, A Cook, M Dibas, R Digpal, S N Faust, A Fazleen, M Felongo, S Fletcher, J Forbes, T Francis-Bacon, A Freeman, J Halliday, M Hardwick, E Holliday, M Johnson, A Jones, C E Jones, T Jones, A Koutalopoula, J Law, E Marouzet, C Mc Cague, S Michael, M Nelson, J Nolasco, A Pambouka, S Patel, M Petrova, T Phillips, L Presland, A Procter, S Rahmany, N Rayner, J Rojkova, R Samuel, L Sanga, T Sass, M Shaji, C Silva Moniz, A Tanner, T Thomas, K Tluchowska, A Torokwa, S Triggs, N Tucker, C Watkins, B Welham, S Wellstead, H Wheeler.

**Western Sussex Hospitals NHS Foundation Trust** L Hodgson (PI), M Margarson (Co-PI), L Albon, A Alcala, K Amin, M Bailey, Y Baird, S Beckley, N Botting, A Butler, V Cannons, C Chandler, A Colino-Acevedo, N Falcone, A Fletcher, S Floyd, L Folkes, H Fox, S Funnell, N Gent, J Gibson, J Gilbert, E Glenday, R Gomez-Marcos, C Gonzalez, M Gorniok, C Goumalatsou, D Green, D Heasman, K Hedges, G Hobden, A Hunter, M Iftikhar, P Jane, R Khan, S Kimber, K King, M King, C Lau, M Linney, L Lipskis, J Margalef, T Martindale, A Mathew, P McGlone, E Meadows, D Melville, S Moore, S Murphy, Q Nguyen, R Njafuh, L Norman, N Numbere, D O Rinn, J Park, M Perera, E Petrovics, D Ravaccia, D Raynard, D Reynish, J Richardson, C Ridley, A Roskilly, T Shafi, T Shaw, S Sinha, J Smith, T Standley, F Stourton, S Sweetman, D Szabó, P Tate, Y Thirlwall, P Thorburn, S Troedson, T Tsawayo, E Vamvakiti, R Venn, N White, J Wileman, E Yates.

**Guy's and St Thomas' NHS Foundation Trust** H Winslow (PI), M Abeywickrema, L Aguilar Jimenez, M Ahmad, N Amutio Martin, S Aslam, L Aslett, T Bawa, K Bisnauthsing, L Brace, L Bremner, K Brooks, B Browne, K Burns, J Butler, M Carter, B Castles , K Chan, L Chappell,

A Chiapparino, P Cinardo, J Cohen, D Cooke, J Cordle, T Crowley, C D'aloia, P Dargan, A Davies, T Doudouliaki, D Finucane, M Flanagan, O Fox, D Hake, B Hamilton, D Hydes, B Jackson, T Jayatilleke, L Jimenez, L Jose, A Kasiappan Balasubramanian, J Kenny, H Kerslake, E Lee, G Lee, A Lewin, C Marsh, N Martin, L Martinez, M Mathew, A Mazzella, L McCabe, B Merrick, S Mieres, N Mlambo, F Morselli, L Nel, M Ng, Y Ng, S Nizamis, M O Toole, M Opena, M Ostermann, A Packham, D Pamela, T Rajeswaran, A Raynsford, A Rose, J Ross, K Rutkowski, K Satchithananthasivam, T Serafimova, C Singh, L Snell, N Spence, D Symington, H Tarft, H Thompson, S Tohill, E Wayman, C Williamson, K Wilson, A Xhikola, C Yearwood Martin, G Zindoga.

**Royal Devon and Exeter NHS Foundation Trust** R Sheridan (PI), M Masoli (Co-PI), H Bakere, A Bowring, T Burden, A Corr, P Czylok, L Dobson, A Forrest, E Goodwin, H Gower, A Hall, S Heddon, G Joseph, L Knowles, H Mabb, A Mackey, V Mariano, E Matkins, E McEvoy, L Mckie, P Mitchelmore, L Morgan, T Nightingale, R Oram, C Oreilly, N Osborne, H Palfrey, S Patten, J Pearce (Associate PI), I Seaton, A Smallridge, P Smith, M Steward (Associate PI), D Sykes, J Tipper, S Todd, C Webb, S Whiteley, S Wilkins, N Withers, K Zaki, L Zitter.

**Southern HSC Trust** R Convery (PI), J Acheson, J Brannigan, D Cosgrove, C McCullough, D McFarland, R McNulty, S Sands, O Thompson.

**The Rotherham NHS Foundation Trust** A Hormis (PI), C Brown, D Collier, J Field, C Graham, J Ingham, V Maynard, J McCormick, S Oakley, S Poku, S Sampath, R Walker, L Zeidan.

**The Royal Wolverhampton NHS Trust** S Gopal (PI), R Barlow, S Bhasin, A Bland, C Chacko, C Cheong, D Churchill, K Davies, S Ganguly, M Green, N Harris, W Hawkins, S Kaur, A Kumar, A Macduff, S Metherell, S Milgate, R Pearse, E Qureshi (Associate PI), L Radford, J Rogers, I Sayers, A Smallwood, K Vassell, D Warrender, L Wild.

**Yeovil District Hospital NHS Foundation Trust** A Broadley (PI), S Abdul, N Beer, S Board, C Buckley, A Daxter, I Doig, L Every, A Getachew, B Giri, i Hamed-Adekale, E Hindley, L Howard, R Jonnalagadda, A Kubisz-Pudelko, A Lewis, K Mansi, R Mason, A Melinte, B Mulhearn, Z N Oo, S Pallipparambil Antony, J Reid, A Shah, R Smith, A Uddin, Z Ul-Haq, D Wood.

**Chesterfield Royal Hospital NHS Foundation Trust** N Spittle (PI), S Beavis, S Beghini, A Blowers, L Blundell, J Bradder, S Broadhead, K Campbell, C Cart, J Cooke, J Cort, J Cresswell, A Crowder, K Dale, S Davies, A Foo, A Forsyth, C Foster, J Gardner, R Gascoyne, P Hickey, M Hollowday, T Horne, M Kelly-Baxter, L Lawson, J Lee, R Lewis (Associate PI), L Lowe, E Mackay, A Mendelski, N Minskip, L Mitchell, E Moakes, L Morgan, K Moxham, I Ogunjembola, A Padmakumar, L Peters, J Pobjoy (Associate PI), K Pritchard, J Salmon, C Sampson, K Sharp, A Smith, R Smith, L Stevenson, S Vittoria, N Weatherly, E Welch, A Whileman, S Williams, E Wolodimeroff, S Wright.

**County Durham and Darlington NHS Foundation Trust** J Limb (PI), V Atkinson, M Birt, C Brady, E Brown, A Cowton, V Craig, P De, D Egginton, D Fernandes, A Hallman, N Hewitson, A Ivy, D Jayachandran, J Jennings, A Kay, M Kent, T Khalifa, S Manning, S McAuliffe, S Naylor, G Nyamugunduru, J O'Brien, M Omar, K Postlethwaite, K Potts, P Ranka, G Rogers, M Sen, S Sen, J Temple, I Tzinieris, S Wadd, H Walters, M Wayman, T Wong, J Yorke.

**Gloucestershire Hospitals NHS Foundation Trust** S Message (PI), F Ahmed, O Barker, O Bintcliffe, P Brown, R Bulbulia, S Bullock, A Chavasse, K Collins, J Collinson, T Cope, A Creamer, C Davies, W Doherty, E Eldridge, I Evans, H Farr, M Fredlund, L Gale, J Glass, S Harrington, A Hill, H Iftikhar (Associate PI), M James, S Jones, C Lim, J Mellersh, H Munby, D Nakiboneka-Ssenabulya, J Noble, M O'Flaherty, J Ord, J Patel, R Peek, T Pickett, N

Robinson, C Sharp, A Simpson, M Slade, H Steer, C Thompson, A Tyler, H Uru, H Uzu, N Vallotton, J Waldron, D Ward, R Woolf.

**Kettering General Hospital NHS Foundation Trust** N Siddique (PI), M Abedalla, A Abeer Ul Amna, K Adcock, S Adenwalla, O Adesemoye, R Ahmed Ali, A Ali, S Aransiola, M Ashraf, M Ashraq, S Ashton, H Asogan, A Aung, H Aung, A Baral, F Bawani, S Beyatli, H Britton, S Chowdhury, M Curtis, J Dales, R Deylami, A Elliott, S Gali, A Gkioni, L Hollos, N Hollos, M Hussam El-Din, A Ibrahim, Y Jameel, G Keyte, J Khatri, S Kiran, R Kusangaya, S Little, K Lwin, E Lyka, A Madu, G Margabanthu, M Mustafa, H Naeem, D Ncomanzi, G Nikonovich (Associate PI), A Nisha James, C Oo, Y Owoseni, N Pandian, D Patel, D Patel, I Petras, M Raceala, D Ramdin, G Ramnarain, S Saunders, A Shaikh, M Shakeel, S Sharma, S Stapley, S Sudershan, P Swift, N Veli, A Virk, T Ward, A Wazir, S White, J Wood, N Zakir.

**Luton and Dunstable University Hospital NHS Foundation Trust** M Nisar (PI), S Tariq (Co-PI), N Ahmed (Associate PI), A Alabi, S Ali, S Allen, M Alzetani, C Ambrose, K Aneke, T Angel, Z Aung, A Aziz, R Banerjee, T Baqai, A Batla, M Bergstrom, S Bhakta, N Bibi, K Bull, T Chapman, O Choudhury, S Dipro, L Dirmantaite, M Edmondson, E Elfar, M Elgamal, H El-Sbahi, D Fishman, C Fornolles, T Forshall, A Francioni, S Gent, A Ibrahim, A Ingram, R James, D Joshi, K Kabiru Dawa, F Khan, M Khan, S Lee, C Lingam, C Luximon, N Mahdi, A Malicka, N Marcus, M Masood, R Mejri, A Moharram, C Moss, S Mottershaw, G Naik, A Ng, L Nicholls, V Parmar, I Perera, F Prasanth Raj, V Puisa, V Quick, B Ramabhadran, A Reddy, A Reka, N Riaz, B Rudran, S Sabaretnam, H Sagoo, D Salim, S Sarma, K Savlani, P Shah, D Shaw, S Siddique, S Soo, P Sothirajah, I Southern, M Tate, C Travill, V Uppal, D Vayapooree, B Wali, W Wakeford.

**NHS Lothian: Royal Infirmary of Edinburgh** A Gray (PI), J Dear (Co-PI), M Adam, R Al-Shahi Salman, A Anand, R Anderson, J Baillie, D Baird, T Balaskas, J Balfour, C Barclay, P Black, C Blackstock, S Brady, R Buchan, R Campbell, J Carter, P Chapman, M Cherrie, C Cheyne, C Chiswick, A Christides, D Christmas, A Clarke, M Coakley, A Corbishley, A Coull, A Crawford, L Crisp, C Cruickshank, D Cryans, M Dalton, K Dhaliwal, M Docherty, R Dodds, L Donald, S Dummer, M Eddleston, S Ferguson, N Fethers, E Foster, R Frake, N Freeman, B Gallagher, E Gaughan, D Gilliland, E Godden, E Godson, J Grahamslaw, A Grant, A Grant, N Grubb, S Hainey, Z Harding, M Harris, M Harvey, D Henshall (Associate PI), S Hobson, N Hunter, Y Jaly, J Jameson, D Japp, R Kay, H Khin, L Kitto, S Krupej, C Langoya, R Lawrie, A Levynska, M Lindsay, A Lloyd, S Low, B Lyell, D Lynch, J Macfarlane, L MacInnes, I MacIntyre, A MacRaild, M Marecka, A Marshall, M Martin, E McBride, C McCann, F McCurrach, M McLeish, R Medine, H Milligan, E Moatt, W Morley, S Morrison (Associate PI), M Morrissey, K Murray, S Nelson, D Newby, K Nizam Ud Din, R O'Brien, M Odam, E O'Sullivan, R Penman, A Peterson, P Phelan, G Pickering, T Quinn, N Robertson, L Rooney, N Rowan, M Rowley, R Salman, A Saunderson, J Schafers (Associate PI), C Scott, L Sharp, A Shepherd, J Simpson, E Small, P Stefanowska, A Stevenson, S Stock, J Teasdale, E Thompson, J Thompson, I Walker, K Walker, A Williams.

**Royal Surrey County Hospital NHS Foundation Trust** K McCullough (PI), H Abu, C Beazley, H Blackman, P Bradley, D Burda, B Creagh-Brown, J de Vos, S Donlon, C Everden, J Fisher, H Gale, D Greene, O Hanci, L Harden, E Harrod, N Jeffreys, E Jones, J Jones, R Jordache, C Marriott, I Mayanagao, R Mehra, N Michalak, O Mohamed, S Mtuwa, K Odedra, C Piercy, V Pristopan, A Salberg, M Sanju, E Smith, S Stone, E Tarr, J Verula.

**Royal United Hospitals Bath NHS Foundation Trust** J Suntharalingam (PI), R Anstey, J Avis, C Bressington, C Broughton, A Budds, S Burnard, H Burton, R Carver, D Catibog, T Clark, T Cooke, T Costa, C Demetriou, G Dixon (Associate PI), H Duncan, F Easton, J Evans, J Fiquet, J Ford, J Goodlife, O Griffiths, R Hamlin, J Harper, T Hartley, S Jones, I Kerslake, A Kirby, J Macaro, R MacKenzie Ross, C Marchand, V Masani, Z Maseko, R Mason, H McDill, C McKerr, S Mitchard, J Noble, J Nolan, A Palmer, C Parrish, J Pullen, L Ramos, M Rich, J

Rossdale (Associate PI), A Seatter, S Sturney, G Towersey, J Tyler, J Walters, K White, T Williams, S Winearl, S Zulfikar.

**South Eastern HSC Trust** F McElwaine (PI), V Adell, D Alderdice, H Allsop, J Baker, A Campbell, J Courtney, R Eadie, A Eccles, J Elder, J Foreman, G Gamble, P Gillen, S Graham, S Hagan, L Hammond, D Hart, K Henry, R Hewitt, K Jones, S Kelly, A Kerr, D Kinnear, C Loughlin, J MacIntyre, C Madden-McKee, K McCollum, J McFlynn, J McKeever, K McMillen, L Moore, C Mulligan, C O'Gorman, F O'kane, J Patterson, S Regan, S Rowan, A Smith, T Trinick, B Valecka, P Yew, G Young.

**Walsall Healthcare NHS Trust** K Shalan (PI), M Akram, S Al-Hity, M Ali, A Alina, B Allman, J Bearpark, S Bibi, A Bland, A Bowes, L Boyd, W Campbell, Z Chandler, J Collins, L Dwarakanath, A Farg, A Foot, S Gaffarena, A Garg, A Gondal, T Gupta, A Hassan, M Islam, T Jemima, B Jones, P Joseph, O Khan, J Khatri, R Krishnamurthy, H Mahmoud, R Marsh, R Mason, M Matonhodze, S Misra, A Mohammad, O Mostafa, J Muhammad, A Naqvi, S Nortcliffe, S Odelberg, M Phipps, K Punia, H Qureshi, G Rajmohan, P Ranga, A Raymond-White, N Richardson, L Rogers, A Sheikh, P Sinha, A Srirajamadhuveeti, N Sunni, R Turel, E Virgilio, H Willis, F Wyn-Griffiths.

**Bolton NHS Foundation Trust** M Balasubramaniam (PI), C Acton, S Ahmad, R Ahmed, A Ajmi, A Al-Asadi, S Altaf, A Amin, A Bajandouh, R Carey, Z Carrington, J Chadwick, S Cocks, C Dawe, S Farzana, O Froud, A Gibson, A Green, P Hill, A Hindle, R Holmes, G Hughes, R Hull, M Ijaz, R Kalayi, M Khan, A Koirata, S Latham, G Lipscomb, K Lipscomb, A McCorkindale, M McNulty, O Meakin, N Meghani, N Natarajan, D Nethercott, P Nicholas, T Pandya, A Parkinson, V Priyash, L Pugh, J Rafique, J Robertson, M Saleh, W Schneblen, B Sharma, O Sharma, D Shaw, Z Shehata, J Shurmer (Associate PI), R Sime, S Singh, R Smith, C Subudhi, R Tallent, E Tanton, K Teasdale, D Tewkesbury, P Thet, S Thornton, J Timerick, C Underwood, N Wang, M Watts, I Webster, B Wilson.

**Doncaster and Bassetlaw Teaching Hospitals NHS Foundation Trust** C H Wong (PI), S Allen, Y Aung, R Chadwick, R Codling, V Cooke, M Cox (Associate PI), F Dunning, M Fairweather, G Herdman, M Highcock, J Kay, N Khoja, G Kirkman, C Knapp (Associate PI), M Kyi, M Luscombe, C Marsden, V Maxwell, A McGeachan, D Murray, E Mustafa, A Nasimudeen, A Natarajan, P Prentice, D Pryor, F Shafique, L Thompson (Associate PI), M Wallis, D Walstow, L Warren, N Wilkinson.

**Frimley Health NHS Foundation Trust** M Meda (PI), J Democratis (Co-PI), Y Abusamra, J Al-fori, N Barnes, N Brooks, E Bryden, L Chapman, R Conway, J da Rocha, R Dolman, A Edwards, T Foster, F Fowe, S Gee, H Gunasekara, R Ho, C Hodge, S Holland, C Hussain, S Jaiswal, A Jayadev, A Jones, N Kader, M Kain, L Kavanagh, S Lee, Y Madani, H Mahmood, A Maqsood, L McAllister, S Menzies, M Molloholli, T Nisar, J Norcliffe, S Power, A Raguro, H Rayner, F Regan, G Roberts, L Rowe-Leete, A Shaw, V Singler, C Smith, T Sonoiki, D Stewart, H Taylor, O Touma, M van de Venne, T Weerasinghe.

**Homerton University Hospital NHS Foundation Trust** S Capocci (PI), A Claxton (Co-PI), Y Akinfenwa, N Aladangady, M Avari, A Begum, H Bouattia, A Boyd, R Brady, L Cabrero, L Canclini, A Chiapparino, R Corser, A Drexel, T Fong, R Frowd, C Gorman, C Holbrook, M Jagpal, S Jain, M James, J Kaur, R Leary, J Miah, R Mullett, C Nimmo (Associate PI), I Noakes, C Quah, P Reynolds, T Tanqueray, I Teeluck, L Terry, E Timlick, K Woods.

**London North West University Healthcare NHS Trust** A Whittington (PI), B Akinbiyi, R Ali, J Barnacle, J Barrett, Z Cahilog, F Cawa, S Chhabra, S Chita, M Colin, A Dagens, E Davenport, E Dhillon, S Filson, J Goodall, R Gravell, A Gupta-Wright, S Gurram, H Houston, G Hulston (Associate PI), S Isralls, N Khan, C Kukadiya, J Low, K Man, B Nayar, O Ojo, P

Papineni, V Parris, M Patel, S Quaid, H Rafferty, A Sturdy (Associate PI), B Tyagi, L Vaccari, N van der Stelt, K Vutipongsatorn, G Wallis, E Watson.

**Manchester University NHS Foundation Trust** T Felton (PI), S Carley (Co-PI), R Lord (Co-PI), A Ustianowski (Co-PI), M Abbas, A Abdul Rasheed, T Abraham, S Aggarwal, A Ahmed, A Ahmed, S Akili, P Alexander, A Allanson, B Al-Sheklly, D Arora, M Avery, C Avram, A Aya, J Banda, H Banks, M Baptist, M Barrera, E Barrow, R Bazaz, R Behrouzi, M Bennett, V Benson, A Bentley, A Bhadi, A Biju, A Bikov, K Birchall, S Blane, S Bokhari, P Bradley, J Bradley-Potts, J Bright, R Brown, S Burgess, M Butt, G Calisti, C Carey, N Chaudhuri, S Chilcott, C Chmiel, A Chrisopoulou, E Church, R Clark, J Clayton-Smith, R Conway, E Cook, S Crasta, G Cummings-Fosong, S Currie, H Dalgleish, C Davies, K Dean, A Desai, R Dhillon, J Digby, D Dolan, G Donohoe, A Duggan, B Duran, H Durrington, C Eades, R Eatough, S Elyoussfi, F Essa, G Evans, A Fairclough, D Faluyi, S Ferguson, J Fielding, S Fiouni, J Flaherty, G Fogarty, S Fowler, A Fox, C Fox, B George, V George, S Giannopoulou, R Gillott, A Gipson, S Glasgow, T Gorsuch, G Grana, G Gray, A Grayson, G Grey, B Griffin, J Guerin, P Hackney, B Hameed, I Hamid, S Hammond, S Handrean, A Harvey, J Henry, S Hey, L Higgins, L Holt, A Horsley, L Howard, S Hughes, A Hulme, P Hulme, A Hussain, M Hyslop, J Ingham, O Ismail, A Jafar, R Jama, S Jamal, L James, F Jennings, A John, M John, E Johnstone, D Kanabar, N Karunaratne, Z Kausar, J Kayappurathu, R Kelly, A Khan, W Khan, J King, S Knight, E Kolakaluri, C Kosmidis, E Kothandaraman, S Krizak, K Kuriakose, N Kyi, F Lalloo, G Lawrence, G Lindergard, C A Logue (Associate PI), L Macfarlane, A Madden, A Mahaveer, L Manderson, G Margaritopoulos, P Marsden, J Mathews, A Mathioudakis, E McCarthy, J McDermott, B McGrath, P McMaster, H McMullen, C Mendonca, A Metryka, D Micallef, A Mishra, H Mistry, S Mitra, S Moss, A Muazzam, D Mudawi, C Murray, M Naguib, S Naveed, P Ninan, M Nirmalan, R Norton, N Odell, R Osborne, G Padden, A Palacios, A Panes, C Pantin, B Parker, L Peacock, A Peasley, N Phillips, M PI, F Pomery, J Potts, N Power, M Pursell, A Ramchandani, A Rasheed, S Ratcliffe, M Reilly, C Reynard, E Rice, M Rice, P Riley, P Rivera Ortega, J Rogers, T Rogers, R Santosh, T Scoones, A Scott, K Sellers, N Sen, T Shanahan (Associate PI), A Shawcross, S Shibly, C Shilladay, A Simpson, S Sivanadarajah, N Skehan, C Smith, J Smith, L Smith, J Soren, W Spiller, K Stewart, J Stratton, A Stubbs, A Sukumaran, K Swist-Szulik, B Tallon, C Taylor, R Tereszkowski-Kaminski, S Thomas, S Thorpe, M Tohfa, R Tousis, T Turgut, A Uriel, M Varghese, G Varnier, I Venables, S Vinay, R Wang, L Ward, C Warner, E Watson, D Watterson, L Wentworth, C Whitehead, D Wilcock, J Williams, E Willis, L Woodhead, S Worton, B Xavier.

**NHS Forth Valley: Forth Valley Royal Hospital** J Selwyn (PI), A Baggott, K Bohmova, G Clark, J Donnachie, E Henderson, S Huda, G Jayasekera, I Macpherson, M Maycock, E McCann, L McGenily, N McInnes, J McMinn, D Morrison, A Pearson, L Prentice, C Rafique, D Salutous, M Spears, M Stewart, R Thompson, A Todd, P Turner.

**NHS Greater Glasgow and Clyde: Queen Elizabeth University Hospital** M Sim (PI), K Blyth (Co-PI), L Abel, A Arnott, H Bannister, N Baxter, S Beattie, C Doherty, J Ferguson, K Ferguson, S Finan, L Gilmour, R Hague, S Henderson, P Holland, L Jawaheer, L Kelly, S Kennedy-Hay, A Kidd, H King, M Ledingham, K Longbottom, F Lowe, M Lowe, G McCreath, R McDougall, M McFadden, M McGettrick, N McGlinchey, L McKay, B McLaren, F Mcmeeken, C McParland, T McSorley, J McTaggart, J Millar, B Milligan, S Moore, L O'Donohoe, I Orlikowska, N Parker, K Paterson, T Pettigrew, A Phillips, L Pollock, M Ralston, K Robertson, J Rollo, S Silva, H Stubbs, M Tate, L Walker, F Watson, M Wilson, S Wishart.

**NHS Lanarkshire: University Hospital Hairmyres** M Patel (PI), T Baird, D Bell, R Boyle, F Burton, L Clark, K Douglas, L Glass, R Hamill, L Jamieson, E Lee, L Lennon, S Marshall, J McKeane, T McLennan, C Meney, R Tejwani, B Welsh.

**NHS Lanarkshire: University Hospital Wishaw** M Patel (PI), N Ali, K Black, R Boyle, S Clements, J Fleming, A Fyfe, L Glass, R Hamill, L Hamilton, C Hughes, E Jarvie, L Lennon, A

Lynas, C Macdonald, S Marshall, M Maycock, C McEwan, E McGarry, K Moar, N Moody (Associate PI), S Naidoo, Z Puyrigaud, A Smith, C Stewart, B Welsh, J West, P Wu.

**Armed Police Force Hospital, Nepal** R Jha (PI), B Adhikari, R Ale, A Gupta, A Gurung, S Gyawali, J Khatri, S Paudel, Y Sapkota, A Shrestha.

**North Cumbria Integrated Care NHS Foundation Trust** C Graham (PI), A Abdelaziz, P Adair, O Ali, W Armstrong, J Atkinson, G Bell, C Brewer, E Carver, M Clapham, R Cleeton, S Cook, H Craig, J Craig, M Elsebaei, R Graham, J Gregory, S Hanif, R Harper, M Hodson, M Holliday, P Jaques, M Lane, J Masters, E Mawson, A McSkeane, P Mead, R Mutch, G Mynott, A Nelson, M Phipps, U Poultney, K Poulton, S Pritchard, S Shah, J Shawcross, G Simmons, C Smit, S Trous, P Tzavaras, V Vasadi, A Wilson, T Wilson, M Wood, D Zehnder.

**Oxford University Hospitals NHS Foundation Trust** K Jeffery (PI), M Ainsworth, C Arnison-Newgass, A Bashyal, K Beadon, S Beer, S Black, A Bloss, S Blrd, L Buck, D Buttress, W Byrne, A Capp, P Carter, L Carty, P Cicconi, R Corrigan, C Coston, L Cowen, N Davidson, K Dixon, L Downs, J Edwards, R Evans, S Gardiner, D Georgiou, A Gillesen, A Harin, M Havinden-Williams, R Haynes, C Hird, A Hudak, P Hutton, R Irons, P Jastrzebska, S Johnston, M Kamfose, K Lewis, T Lockett, F Maria del Rocio, J Martinez Garrido, S Masih, A Mentzer, S Morris, G Mounce, C O'Callaghan, Z Oliver, J Patachako, S Paulus, E Perez, L Periyasamy, L Peto, D Porter, S Prasath, C Purdue, M Ramasamy, C Roehr, A Rudenko, V Sanchez, A Sarfatti, M Segovia, T Sewdin, J Seymour, V Skinner, L Smith, A Sobrino Diaz, G Soni, M Taylor-Siddons, H Thraves, C Tsang, M Vatish, Y Warren, E Wilcock, T WIshlade.

**RSUP Dr Hasan Sadikin, Bandung** U Sumardi (PI), A Albertus, A Bachti, A Amelia Ganefianty, EN Lestari, TT Maulidya, W Naftalia, CB Prayuda, I Silanas, RF Sobarna, T Tulus Satriasih, W Rina, H Yovita.

**Royal Cornwall Hospitals NHS Trust** D Browne (PI), Z Berry, H Chenoweth, A Collinson, F Hammonds, C James, L Jones, E Laity, K Morgan, C Murphy, T Nisbett, R Sargent, L Trethowan, K Watkins, L Welch.

**Torbay and South Devon NHS Foundation Trust** L Anning (PI), P Acheampong, I Akinpelu, T Allen, S Atkins, N Aveyard (Associate PI), M Bailey, E Baird, M Ball (Associate PI), J Blackler, C Carver, M Chirgwin, T Clark, J Clouston, G Curnow, M Dawson, Z Dyar, L Ellis, C Fearnley, A Foulds, S Godlee, J Graham, A Green, C Grondin, E Hale, S Howlett, C Huggins, A Hutchinson (Associate PI), L Kyle (Associate PI), C Lam, S Maddison, R Mankiewitz, S Martin, P Mercer, B Murphy, W O’Rourke, J Palmer, M Parkinson, A Redome, J Redome, A Revill, K Subramaniam, T Tanzil-Al-Imran, R Tozer, J Turvey, C Webb, S Wright.

**Airedale NHS Foundation Trust** T Gregory (PI), M Babirecki, H Bates, E Dooks, F Farquhar, B Hairsine, S Marsden (Associate PI), S Nallapeta, S Packham.

**Alder Hey Children's NHS Foundation Trust** D Hawcutt (PI), K Allison, R Dore, J Moss, L O'Malley, L Rad, G Seddon.

**All India Institute Of Medical Sciences, Rishikesh, Uttarakhand** P Kumar Panda (PI), V Pai (co-PI), Y Bahurupi, G Chikara, P Dhangar, D Kalita, R Kumar, V Saini, N Sarkar.

**Aneurin Bevan University LHB** S Fairbairn, (PI), L Aitken, K Arora, E Baker, M Brouns, O Burbidge, J Cann, S Cherian, S Cutler, A Dell, C Dunn (Associate PI), N Duric (Associate PI), M Edwards, S El Behery, S Goyal, J Grenville, A Griffiths, J Harris, N Hawkins, E Heron, J Hickey, M Hoare, S Hodge, G Hodkinson, A Ionescu, C Ivenso, C James, P James, T James, M Jones, S Jones, N Lawson, A Lucey, G Mallison, R Manikonda, G Marshall, S Nicholls, J Northfield, A Paracha, L Parfitt, A Pennington, L Peter, C Price, M Pynn, D Ritchie, E

Rowlands, A Roynon-Reed, M Scott, C Sing, J Singh, M Singh, C Somashekar, T Szakmany, J Tuffney, R Venkataramakrishnan, G Warwick, A Waters, G Williams, S Williams, K Zalewska.

**Ashford and St Peter's Hospitals NHS Foundation Trust** C Russo (PI), M Aquino, M Croft, S Fathima, V Frost, M Gavrila, K Gibson, A Glennon, C Gray, N Holland, K Jabbar, N Johnson, J Law, F McGee, R Pereira, L Renouf, P Reynolds, H Tarft, J Thomas, L Walding, A Williams.

**Barking, Havering and Redbridge University Hospitals NHS Trust** M Phull (PI), A Umaipalan (Co-PI), N Adams, M Awaly, C Baldwin, A Basumatary, S Bates, H Chandler, A Davison, G De-La-Cedra, P Dugh, K Dunne, K Fielder, A George, P Greaves, W Halder, J Hastings, H Hlaing, A Holman, S Huang, K Hunt, V Karunanithi, V Katsande, M Khalid, R Khan, P Knopp, A Loverdou, A McGregor, A Misbahuddin, A Mohamed, H Neils, D Nicholls, N O'Brien, J Olatujoye, L Parker, T Pogreban, L Rosaroso, S Sagrir, E Salciute, P Saravanamuthu, M Sharma, G Siame, H Smith, R Suthar, E Visentin, H Ward, E Yacoba.

**Barnsley Hospital NHS Foundation Trust** K Inweregbu (PI), R Bowmer, S Cutts, A Daniels, J Emberey (Associate PI), A Galvin, C Green, R Gupta, L Harrison, J Hartley, A Hassan, C Hirst, S Hope, M Hussain, S Hussein, A Khalil, M Longshaw, S Meghjee, A Nicholson, A Sanderson, E-J Stoner, C Swales, D Webster.

**Barts Health NHS Trust** H Kunst (PI), H Abbass, A Aboaba, A Alam, F Ali, R Allen, V Amosun, C Ardley, R Astin-Chamberlain, G Bacon, H Baillie, V Baker, S Begum, F Bibi, B Bloom, M Browne, R Buchanan, M Buckland, N Caponi, C Chan, S Chandler, B Cipriano, H Dawe, M Dawkins, P Dias, S Elia, K El-Shakankery, D Fedorova (Associate PI), M Fernandez, A Fikree, R Flynn (Associate PI), R Goiriz, P Goldsmith, M Griffiths, S Grigoriadou, K Gunganah, B Gurung, B Hack (Associate PI), P Hartridge, C Harwood, K Hashem, C Heckman, U Hemmila, D Ho, D Hobden, L Howaniec, G Huntington, I Hussain, S Iliodromiti, M Jawad, A Jayakumar, V Kapil, S Kelly, M Khan, H Khatun, J Lai (Associate PI), I Lancona-Malcolm, I Lee, D Lieberman, S Liebeschuetz, C Lindsay, O Lucey (Associate PI), K Majid, H Malcolm, C Maniero, H Marshall (Associate PI), T Martin, J Martin-Lazaro, N Matin, L Mayola, E McAleese, R McDermott, B McDonald, A Miah, I Milligan (Associate PI), A Mohammed, N Moramorell, S Naeem, T Newman, C Nic Fhogartaigh, L Noba, M Omar, N Omer, G Osoata, A Pakozdi, K Parkin, R Pearse, P Pfeffer, N Plaatjies, J Pott, J Powell, R Rachman (Associate PI), M Ramirez, K Rathod, W Ricketts, A Riddell, P Rughani, N Sahdev, K Samuels, F Santos, K Sarkar, G Sedgwick, F Seidu, B Selvarajah, M Shaikh, T Simangan, I Skene, K Smallshaw, A So, C Suarez, T Swaine (Associate PI), S Thomas, N Thorn, S Tiberi, C Tierney, G Tunesi, C Tweed, R Uddin, S Ullah, L Velauthar, K Wiles, T Wodehouse, P Woodland, H Yong, S Youssouf.

**Basildon and Thurrock University Hospitals NHS Foundation Trust** K Thomas (PI), T George (Co-PI), E Cannon, J Cartwright, M Forsey, A Ikomi, L Kittridge, G Maloney, D Mukherjee, M Mushabe, A Nicholson, K Pannu, M Pittman, J Riches, J Samuel, N Setty, A Solesbury, D Southam, M Vertue, K Wadsworth, B Yung.

**Belfast HSC Trust** D Downey (PI), A Blythe, S Carr, A Cassells, D Comer, D Dawson, K Hayes, R Ingham, L Keith, J Kidney, E Killen, J Leggett, D Linden, N Magee, R Marshall, D McClintock, M McFarland, L McGarvey, C McGettigan, S McGinnity, S McMahon, A Nugent, M Paul, A Redfern-Walsh, J Richardson, L Speirs, M Spence, R Stone, D Tweed, A Usher-Rea, B Wells.

**Betsi Cadwaladr LHB: Glan Clwyd Hospital** D Menzies (PI), N Abdallah, A Abou-Haggar, R Al-Sammarraie, S Ambalavanan, K Darlington, F Davies, G Davies, J Davies, L Davies, H Duvnjak, J Easton, T Grenier, M Joishy, R Lean, J Lewis, C Mackay, B Parry, K Parvin, R Poyner, R Pugh, X Qui, S Rees, J Scanlon, N Sengupta, S Ullah, H Williams.

**Betsi Cadwaladr LHB: Wrexham Maelor Hospital** D Southern (PI), S Ahmer, R Bachar, G Bennett, L Brohan, D Counsell, R Cowell, S Davies, D Dhiru, J Harris, E Heselden, M Howells, R Hughes, S Kelly, J Kilbane, A Lloyd, H Maraj, A Mashta, G Mayers, R Petersen, H Reddy, L Richards, S Robertson, S Sandow, O Smith, S Smuts, G Spencer, G Szabo, S Tomlins.

**Betsi Cadwaladr LHB: Ysbyty Gwynedd** C Subbe (PI), N Abdallah, C Bishop, N Boyer, N Boyle, C Butterworth, T Collingwood, S Evans, T Grenier, J Healy, M Joishy, E Knights, A Kutera, C Lewis, M Payne, S Rawashdeh, G Rieck, W Scrase, C Speare, E Stenson, A Thomas, C Thorpe, A Van Loggerenberg.

**Birmingham Women's and Children's NHS Foundation Trust** K Morris (PI), D Jyothish (Co-PI), H Krishnan (Co-PI), E Al-Abadi, J Groves, S Hartshorn, K Hong, R Horner (Associate PI), R Howman, S Jordan, C O'Hara, S Samar (Associate PI), H Sohal, S Sultan, H Williamson, H Winmill.

**Bradford Teaching Hospitals NHS Foundation Trust** D Saralaya (PI), N Akhtar, V Beckett, L Brear, J Butler, V Drew, J Eedle, N Hawes, S Kmachia, S Moss, S Oddie, J Paget, K Regan, D Ryan-Wakeling, A Shenoy, K Storton, R Swingler, J Syson, J Todd, R Wane, A Wilson.

**Brighton and Sussex University Hospitals NHS Trust** M Llewelyn (PI), R Ahmed, F Baldwin, E Barbon, H Brown, G Bassett, L Bennett, A Bexley, P Bhat, C Bresges, M Bridgett, H Brown, M Campbell, Z Cipinova, A Crepet, E Cross (Associate PI), S Dhillon, A Elkins, L Evans, S Filipa, M Flowerdew, J Gaylard, R Govindan, D Hansen, A Hassan, Z He, C Hunter, O Jones, S Jujjavarapu, C Laycock, E Mekonnen, J Messenger, D Mullan, J Myerson, J Newman, J Nowak, M Pavitt, C Richardson, R Robinson, V Sellick, D Skinner, M Smith, S Sobowiec Kouman, K Stewart, K Trivedi, D Yusef.

**Charlotte Maxeke Johannesburg Academic Hospital** I Kalla (PI), L Labuschagné, ON Makoetlane, J Mhlongo, L Murray, V Naidoo, J Nel.

**Chelsea and Westminster Hospital NHS Foundation Trust** P Shah (PI), B Mann (Co-PI), S Ahmed, K Alatzoglou, K Alizaedeh, M Al-Obaidi, K Ballard, A Barker, C Bautista, M Boffito, M Bourke, D Buche, R Bull, C Caneja, J Carungcong, F Conway, P Costa, E Dwyer, A Farah, C Fernandez, C Fung, J Garner, J Girling, E Hamlyn, A Holyome, L Horsford, N Hynes, J Irisari, M Johnson, U Kirwan, S Maheswaran, M Martineau, T Ngan, K Nundlall, C Orton, T Peters, A Sayan, A Schoolmeesters, M Svensson, A Tana, A Thayanandan, J Tonkin, B Vijayakumar, C Winpenny, F Yang, O Zibdeh.

**Chris Hani Baragwanath Academic Hospital, Soweto** M Tsitsi (PI), A Badat, C Menezes, J Robinson, S Van Blydenstein, M Venter.

**Clairwood Hospital, Mobeni** N P Magula (PI), S Pillay (Co-PI), SA Bengu, C Dipheko, S Kubheka, L Labuschagne, S Matibela, J Nel.

**Croydon Health Services NHS Trust** T Castiello (PI), J Adabie-Ankrah, G Adkins, B Ajay, A Ameen, S Ashok, A Dean, S Dillane, F Fedel, V Florence, D Griffiths, I Griffiths, J Hajnik, J Hetherington, C Jones, A Latheef, S Lee, J McCammon, S Patel, A Raghunathan, P Shah, J Talbot-Ponsonby, G Tsinaslanidis, G Upson.

**Cwm Taf Morgannwg University LHB** C Lynch (PI), Z Auer, E Davies, B Deacon, S Eccles, A Gaurav, B Gibson, M Hibbert, C Lai, L Margarit, D Nair, S Owen, C Pothecary, J Pugh, L Roche, S Sathe, J Singh, J Smeaton, D Tetla, K-A Wilson, C Woodford.

**Dartford and Gravesham NHS Trust** B Khan (PI), D Ail, R Aldouri, K Aung, G Awadzi, B Bassoy, R Bhalla, J Billings, S Bokhari, G Boniface, J Cernova, T Chen, P Chimbo, N Chitalia, S Danso-Bamfo, A Davis-Cook, D Depala, A Dhanoa, T Edmunds, E Fernandez, T Ferrari, B Fuller, Z Galani, A Gherman, P Grist, R Heire, L Ilves, C Kamundi, B Khan, L Lacey, E Lawrence, M Lewis, A Maric, W Martin, Z Min, C Newman, R Nicholas, N Oakley, O Olufuwa, N Pieniazek, M Protopapas, T Qadeer, S Rathore, S Sathianandan, C Scott, A Shonubi (Associate PI), S Siddique, G Sisson, M Soan, D Streit, C Stuart, M Szekely, W Umeojiako, S Urruela, B Warner, M Waterstone, S White, A-M Zafar, S Zaman.

**Dr D Y Patil Medical College, Hospital And Research Centre, Pune** S Kanitkar (PI), P Athavale, MS Barthwal, J Jadhav, P Jali, D Kadad, A Kakrani, R Kothavale, S Mahajan, B Patil, N Sairam, S Sarawade, P Sharma, BG Thomson, S Tripathy.

**East and North Hertfordshire NHS Trust** P Ferranti (PI), F Alsheikh, M Boampoaa, M Chaudhury, C Cruz, M Ebon, M Erotocritou, A Fajardo, A Frosh, S Gelves-Zapata, D Gorog, T Ingle, J Kefas, C Matei, J Mathers, L Miller (Associate PI), I Nadeem (Associate PI), Y Odedina, D Palit, L Peacock, S Tewari (Associate PI), L Ventilacion, R Zill-E-Huma, E Zinkin (Associate PI).

**East Cheshire NHS Trust** T Nagarajan (PI), X Lee (PI), S Shashaa (PI), L Alghazawi, M Babores, C Gorman, A Henderson, M Holland, R Hughes, L Huhn, M Husain, N Keenan, L Paisley, M Porteous, H Wassall, L Wilkinson, K Wolffsohn.

**East Kent Hospitals University NHS Foundation Trust** G Boehmer (PI), A Alegria (Co-PI), R Kapoor (Co-PI), N Richardson (Co-PI), K Adegoke, L Allen, S Anantapatnaikuni, D Baker, E Beranova, H Blackgrove, T Boumrah, P Christian, T Cosier, N Crisp, T Curtis, J Davis, J Deery, A Elgohary, T Elsefi, A Gillian, C Hargreaves, T Hazelton, G Hector, R Hulbert, A Ionita, A Knight, C Linares, S Liu, D Loader, K Lodhia, S Mandal, E Matisa, J McAndrew, K Mears, S Millington (Associate PI), M Montasser, A Moon, C Oboh, P Offord, S Parashar (Associate PI), M Patel, C Pelham, C Price, J Quindoyos, A Rajasri, J Rand, S Rogers, S Saminathan, N Schumacher, A Skaria, R Solly, D Starnes, D Stephensen, S Stirrup, L Tague, S Tilbey, S Turney, V Vasu, A Velugupati, M Venditti, R Vernall, H Weston, Z Woodward.

**East Lancashire Hospitals NHS Trust** S Chukkambotla (PI), A Batista, H Collier, S Duberley, K Geerthan, W Goddard, B Hammond, L Hoole, R Hussain, A Konstantinidis, K Marsden, A Mulla, A Newby, J Nugent, F Pickering, D Rusk, C Spalding, A Sur, D Sutton, V Taylor, N Truman, J Umeadi.

**East Sussex Healthcare NHS Trust** A Marshall (PI), A Abousamra, S Ahmed (Associate PI), S Blankley, H Brooke-Ball, T Christopherson, M Clark, T de Freitas, E de Sausmarez, D Dharmasena, I Doss, A Eggink, A Ekunola, S Ghazal, D Hemsley, J Highgate, A Iakovou, J Jeater, T Kalmus Eliasz, O Kankam, J Khoo, A Lowe, T Mak, S Merritt, Y Mohammed, T Morley, A Newby, S Panthakalam, S Qutab (Associate PI), A Rajagopal, N Roberts, M Simon, K Subba, S Tieger, A Trimmings, R Venn, F Willson, T Win, M Yakubi, A Zubir.

**Father Muller Medical College Hospital, Mangalore** A Bhat (PI), P Dsouza, DK Joy, V KN, Prathima.

**Gateshead Health NHS Foundation Trust** D Mansour (PI), R Allcock, M Armstrong, J Barbour, M Bokhar, J Curtis, A Dale, V Deshpande, I Hashmi, I Helgesen, E Johns, L Jones, R King, V Kumar, E Ladlow (Associate PI), R Mackie, D Mansour, C Maughan, B McClelland, W McCormick, C McDonald, M Mehmood, C Moller-Christensen, A Neal, R Palmer, R Petch, F Rasul, S Razvi, R Sharma, L Southern, G Stiller, E Watkins, H Wilkins.

**George Eliot Hospital NHS Trust** S George (PI), V Bains, M Bryant, S Bukhari, Y Chang, S Dean, N Elndari, H Gachi, J Glasgow, K Ellis, V Gulia, J Gunn, J Heys, C Ison, A Javier, T Kannan, D McDonald, J Mellor, R Musanhu, A Naeem, R Nair, N Navaneetham, S Nawaz, B Nazari, N Patel, E Sung, D Suter, M Tahir, P Vanmali (Associate PI), B Vilcinskaite, E Vlad, R Walker, I Yusuf.

**Ghana Infectious Disease Centre** C Owoo (PI), J Oliver-Commey (Co-PI), AS Adam, PN Agasiya, E K Ahadome, C Aidoo, A Amoako-Adusei, G Arhin-Sam, A Arthur, GA Asare, HA Asiamah, YE Atabudzi, S Awisi, E Boatemah, GS Dadzie, IK Duah, ED Dzidzornu, C Enimpah, S Gyambrah, E Matey, JE Mensshan, IV Omale, L Quarm, R Quartey, S Quaye, LJ Quist, SK Siabi, SK Ssiabi.

**Great Ormond Street Hospital For Children NHS Foundation Trust** M Peters (PI), L Grandjean (Co-PI), O Akinkugbe, J Andrews, A Bamford, H Belfield, S Benkenstein, C Chisholm, L Chiverton, L Dawson, J Hassell, G Jones, K Kupiec, K Leigh-Ellis, I Manjra, T McHugh, C Mellish, K Moshal, L O'Neill, J Penner, R Shamsah, D Shingadia, S Tingley, A Tomas.

**Groote Schuur Hospital, Cape Town** M Mendelson (PI), LE Hermans (Co-PI), M Adriaanse, C Bonaconsa, S Dlamini, N Hickman, M Kati, H Kyepa, G Meintjes, NS Papavarnavas, K Stoffberg, S Wasserman.

**Harrogate and District NHS Foundation Trust** A Kant (PI), A Amin, C Bennett, O Cohen, S Foxton, E Lau, S Longstaffe, C Morgan, M O'Kane, A Royson, N Singh, S Vivekananthan, A Williamson, L Wills.

**Hospital for Advanced Medicine &Surgery (HAMS)** HR Paneru (PI), D Aryal (Co-PI), S Kayastha, D Khadka, R Pangeni, R Shakya, R Shrestha.

**Hospital for Tropical Diseases, Ho Chi Minh City** N Phong (PI), TD Khoa, NT Nguyen, VT Khuong, NN Thao, TH Nguyen, DG Bao, NTM Tuyen, NV Khai, NM Thu, CTC Van, HP Thao, NVH Linh, DTN Quyen, HTC Tu, NQ Vinh, DH Le, HM Phuong, NTT Huyen

**Hywel Dda LHB** S Ghosh (PI), K Lewis (Co-PI), S Coetzee, K Davies, L Hill, B Icke, S Jenkins, S Griffiths, L O'Brien, Z Omar, S Peebles, E Perkins, C Williams, J Williams, C Woollard.

**Hywel Dda LHB: Bronglais General Hospital** M Hobrok (PI), D Asandei, B Atkins, S Jenkins, K Khan, R Loosley, H McGuinness, D McKeogh, A Mohamed, L Raisova, A Snell, H Tench, W Wolf, R Wolf-Roberts.

**Hywel Dda LHB: Withybush Hospital** J Green (PI), Y Adegeye, J Brooks, R Hughes, C Macphee, A Puffett, M Rafiq, G Ross, H Thomas.

**Isle Of Wight NHS Trust** A R Naqvi (PI), A Abbas, B Ahmed Mohamud, A Attia, A Azkoul, A Bester, A Brown, C Caraenache, G Debreceni, S Din, R Fasina, P Firi, A Gardener (Associate PI), S Grevatt, S Hinch, O Hudson, E Jenkins, N Kadam, E Karbasi, A Khan, S Knight, X Liu, C Magier, J Mattappillil, N Mburu, B Mohamud, S Mukhtar, R Naik, P Nathwani, H Ng, E Nicol, E Norris, E O'Bryan, B Osman, N Otey, M Pugh, B Rizvi, P Rogers, J Selley, I Smith, M Stevens, L Sylvester, O Tarin, C Taylor, A Tirumalai Adisesh, J Wilkins, M Woodman.

**James Paget University Hospitals NHS Foundation Trust** J Patrick (PI), B Burton (Co-PI), A Ayers, R Brooks, H Bye, J Chapman, V Choudhary, S Cotgrove, G Darylile, L Felton, D Griffiths, C Hacon, H Hall, W Harrison, P Hassell, A Hearn, F Iqbal Sait, K Mackintosh, J North, S Parslow-Williams, J Sanghera, H Sutherland, M Whelband (Associate PI), C Whitehouse, E Wilhelmsen, J Wong, J Woods.

**Jawaharlal Nehru Medical College, Aligarh Muslim University, Aligarh, Uttar Pradesh** M Shameem (PI), S Anwar, A Fawad, M Tabish.

**King's College Hospital NHS Foundation Trust** J Galloway (PI), M McPhail (Co-PI), J Adeyemi, J Aeron-Thomas, M Aissa, E Alveyn, A Buazon , T Buttle, S Candido, E Canonizado, A Cavazza , M Cesay, K Clark, M Cockerell , A Comerford, M Depante, G Dimitriadis, C Donaldson, K El-Bouzidi, S Finch, C Finney, K Garrero, G Godwin, S Gogoi, N Griffiths, A Gupta, L Hall, J Hannah, Y Hu, E Jerome (Associate PI), N Jones, N Kametas, M Kumar, L Linkson, N Long, M Magtalas, E Makanju, H Martin, D Nagra, H Noble, K O'Reilly, V Patel, T Pirani, A Posada, N Powell, J Prince, D Rao, S Ratcliff, S Saha, B Sari, N Sikondari, J Smith, B Solis, A Te, C Tey, M Tran, R Uddin, A Varouxaki, J Vidler (Associate PI), J Ward, M West, C Williamson, M Yates, A Zamalloa.

**Kingston Hospital NHS Foundation Trust** S Mahendran (PI), A Ali (Co-PI), A Joseph (Co-PI), M Agarwal, S Bacciarelli, A Baggaley, G Bambridge, S Barrett, F Bazari, P Beak, S Bidgood, L Boustred, D Caldow, T Carlin, S Cavinato, A Conroy, J Crooks, E Donnelly, A Edwards, L Elliot, S El-Sayeh, A Feben, J Fox, R Gisby, M Grout, S Hassasing, R Heath, R Herdman-Grant, E Jackson, D Jajbhay, O James, S Jones, J Khera, T Leahy, S Luck, R Maamari, M Madden, H Matthews, J May, M McCullagh, G McKnight, L Mumelj, K Muralidhara, G Natarajan, D Newman, A Nicholson, T O'Brien, J Odone, R Patterson, R Patterson, A Pavely, J Poxon, C Quamina, G Quartermaine, A Ratnakumar, R Rodriguez-Belmonte, T Sanderson, S Sharma, R Simms, D Sivakumran, A Sothinathan, A Swain, S Swinhoe, M Taylor, M Trowsdale Stannard, B Ummat, J Vance-Daniel, H Warren-Miell, R Williams, T Woodhead, D Zheng.

**Komfo Anokye Teaching Hospital** Y Hardy (PI), C Aboah, F Acheampong, L Addai-Boampong, P Adjepong, P Afrim, FA Afriyie, JA Agyekum, K Agyen, J Amoah-Dankwa, M Ansah, JM Ayerh, PG Barning, KA Boahen, J Bonney, KG Danso, M Donkor, D Dwomoh Nkrumah, H Haywood Hasford, M Kuma-Mintah, O Lambert, LN Lartey, L Micah-Amuah, PAB Minkah, L Nyanor, E Ofori, PK Omane-Donkor, A Opata, MA Oteng, C Poku, M Siaw-Frimpong, AB Wongkyezeng.

**Lancashire Teaching Hospitals NHS Foundation Trust** S Laha (PI), A Alty, A Ashfaq, J Beishon (Associate PI), A Bellis, D Cameron, M Chiu, W Choon Kon Yune, M Deeley, W Flesher, K Gandhi, S Gudur, R Gupta, A Huckle, G Long, P Mannion (Associate PI), A McCarrick, J Mills, P Mulgrew, S Nawaz, L Nelson, J Nixon, O Parikh, A Peer, S Punnilath Abdulsamad, D Rengan, E Seager, S Sherridan, B So, R Sonia, T Southworth, S Sowden, K Spinks, A Timoroksa, B Vernon, A Williams, K Williams, H Wu.

**Lewisham and Greenwich NHS Trust** S Kegg (PI), A Aghababaie, H Azzoug, E Bates, M Chakravorty, K Chan, C Chin-Saad, P Coakley, R Dodds, E Gardiner, A Hastings, D Jegede, J Juhl, H Kelly, K Law, M Lewis, M Magriplis, A Mascagni, C Milliken, J Muglu, D Mukimbiri, M Nadheem, T Nair, M Nyirenda, T OConnor, T Ogbara, L Olaivar, R Olaiya, E Omoregie, C Onyeagor, V Palaniappan, A Pieris, S Pilgrim, T Simpson, A Taylor, E Treus Gude, K Wesseldine, R Williams, M Woodman, E Woolley.

**Liverpool Women's NHS Foundation Trust** R McFarland (PI), M Dower, S Holt, A Mahdi, C Morgan, E Neary, A Smith, M Turner, E Willis.

**Medical College Kolkata, Kolkata** A Talukdar (PI), K Chaudhuri, D Sarkar.

**Mid Essex Hospital Services NHS Trust** A Hughes (PI), J Radhakrishnan (Co-PI), R Arnold, T Camburn, E Cannon, C Catley, E Dawson, L Durdle, N Fox, H Gerrish, S Gibson, H Guth,

M Iman, L James, F McNeela, C Mitchell, A Rao, S Reid, B Singizi, S Smolen, M Vitaglione, S Williams, L Willsher, J Wootton.

**National Hospital for Tropical Diseases, Hanoi** PN Thach (PI), NT Cap, NT Ha, LNM Hoa, DTT Thuy, DQ Tung, DP Khiem, PV Phuc, TV Kien, VD Linh, DTT Hang, TV Giang, TT Hien, HTK Oanh

**Nepal Police Hospital** D Paudel (PI), B Rajbhandari (Co-PI), NB Chaudhary, B Gajmer, M Gartaula, J Khatri, SS Kunwar, M Maharjan, P Maskey, K Neupane, R Niroula, A Regmi, S Singh, S Singh, BJ Thakuri, S Yadav.

**NHS Ayrshire and Arran: University Hospital Ayr** K Walker (PI), M Boden, C Burns, D Callaghan, R Cuthbertson, K Gibson, D Gilmour, M Henry, L Kelly, J Locke, L McNeil, S Meehan, A Murphy, K Naismith, K Prasad, M Rodger, R Slingsby, C Turley, S Walton, M Wilson, S Wood.

**NHS Ayrshire and Arran: University Hospital Crosshouse** A Clark (PI), T Adams, S Allen, E Anderson, L Barker, C Burns, D Callaghan, N Connell, V Dey, A Dunleavy, F Elliott, K Gibson, D Gilmour, K Grant, M Henry, J Locke, T Massa, L McNeil, S Meehan, A Murphy, E Peggie, M Rodger, S Smith, C Turley, S Walton, D Wilkin, M Wilson, S Wood.

**NHS Borders: Borders General Hospital** A Scott (PI), G Alcorn, S Alcorn, J Aldridge, J Bain, C Barnetson, P Bell, A Campbell, B Chow, S Corbet, F Craighead, J Dawson, E Dearden, S Dickson, G Donaldson, T Downes, A Duncan, H El-Taweel, C Evans, T Fairbairn, L Finlayson, C Flanders, J Fletcher, J Foo, R Gallagher, N Hafiz, F Hall, M Herkes, M Hume, J Inglis, L Knox, J Lonnen, J Manning, H Matthews, A McLaren, C Murton, R Murton, B Muthukrishnan, E Nicol, F North, C Osbourne, R Porter, K Ralston, R Richmond, F Rodger, H Sakuri, B Soleimani, R Stewart, R Sutherland, A Tan, M Tolson, J Wilkie, R Williamson.

**NHS Dumfries and Galloway: Dumfries & Galloway Royal Infirmary** A W Hay (PI), M McMahon (Co-PI), J Candlish, P Cannon (Associate PI), J Duignan, C Jardine (Associate PI), N Lungu (Associate PI), A Mitra, P Neill (Associate PI), S Svirpliene, J Um (Associate PI), D Williams, S Wisdom.

**NHS Golden Jubilee National Hospital** B Shelley (PI), V Irvine, F Thompson.

**NHS Grampian: Aberdeen Royal Infirmary** J Cooper (PI), R Soiza (Co-PI), V Bateman, M Black, R Brittain-Long, K Collie, K Colville, D Counter, A Coutts, S Devkota, P Dospinescu, F Flett, P Ganley, L Hakeem, R Hunter, J Irvine, L Kane (Associate PI), C Kaye, A Khan, R Laing, A Mackenzie, M MacLeod, G Malik, J McLay, D Miller, J Noble, K Norris, Y Pang, V Taylor, K Thomasson, I Tonna, A Vileito, L Walker.

**NHS Greater Glasgow and Clyde: Glasgow Royal Infirmary** K Puxty (PI), O Adeagbo, C Aiken, J Alexander, L Bailey, H Bayes, A Begg, M Broom, A Brown, S Carmichael, S Cathcart, F Christie (Associate PI), C Clayton, R Colbert, R Cowan, H Cranston, I Crawford, S Currie, J Cuthill, A Dougherty, K Duffy, A Duncan, C Dunne, T French, A Goodfellow, T Grandison, S Griffiths, P Grose, Z Harzeli, J Hawkins, N Hickey, M Hughes, J Ireland, A Jamison, D Jenkins, J Johnstone, R Kearns, D Kernaghan, K Lake, L Latif, L Littlejohn, J Luveta, L Martin, M McCloskey, C McCue, C McGovern, M McIntyre, A Munro, E Murphy, S Nelson, C Ogilvie, L Paton, H Peddie, G Piper, L Pollock, A Puxty, T Quasim, D Rimmer, K Scott, G Semple, M Shaw, S Speirs, F Steffensen, J Tait, S Thornton, L Turner.

**NHS Greater Glasgow and Clyde: Inverclyde Royal Hospital** M Azharuddin (PI), H Papaconstantinou (Co-PI), D Cartwright, W Gallagher, J Hampson, T McClay, E Murray, O Olukoya.

**NHS Greater Glasgow and Clyde: Royal Alexandra Hospital** A Corfield (PI), L Abel, S Brattan (Associate PI), A Brunton, C Clark, P Clark, S Currie, D Grieve, M Hair, M Heydtmann, E Hughes, L Imam-Gutierrez, R Keen, D McGlynn, B McLaren, A Rankin, G Ray, J Robertson, N Rodden, K Rooney, R Sundaram, N Thomson, L Walker.

**NHS Highland** B Sage (PI), R Acquah, L Adamu-Ikeme, C Barr, F Barrett, A Batty, W Beadles, C Bradley, R Campbell, A Cochrane, R Cooper, I Dawson, K Donald, J Finlayson, A Goh, L Heron, G Jervis, P Laidler, S Macleod, S Makin, J Matheson, D McDonald, M McKenzie, C Millar, K Monaghan, L Murray, M O'Hara, L O'Keefe, D Patience, M Peirse, S Reilly, M Robertson, N Shahzad, G Simpson, K Watson, J Wilson.

**NHS Lanarkshire: University Hospital Monklands** M Patel (PI), C McGoldrick (Co-PI), P Anstey, T Baird, C Beith, A Blunsum, D Cairney, A Crothers, S Dundas, F Farquhar, G Fleming, D Garner, L Glass, P Goyal, P Grant, R Hamill, S Howard, T Jones, E Lee, J Lees, L Lennon, C Macrae, S Marshall, A McAlpine, M McFadden, R McGovern, C McInnes, C McKeag, A McKie, M McLaughlin, R Millington, J Neil, N Quail, M Ralston, J Reid, J Robb, M Rodger, S Rundell, T Sammut, M Smith, A Stark, C Sykes, M Taylor, B Welsh, J West, R Williams.

**NHS Lothian: Royal Hospital for Children and Young People** C McDougall (PI).

**NHS Lothian: St John's Hospital** S Lynch (PI), R Anderson, S Begg, J Bentley, S Brady, M Colmar, R Frake, A Gatenby, C Geddie, F Guarino, S Guettari, C Hurley, C Kuronen-Stewart, A MacRaild, M Mancuso-Marcello, M Marecka, G McAlpine, N McCullough, A Megan, T North, O Otite, L Primrose, C Rees, L Rooney, A Saunders, A Saunderson, A Stevenson, S Stock, A Wakefield, E Walsh, J Wraight, T Wright, S Yusef.

**NHS Tayside: Ninewells Hospital** J Chalmers (PI), H Abo-Leyah, C Deas, H Loftus, A Nicoll, A Strachan, J Taylor, C Tee.

**NHS Western Isles** M Murdoch (PI), A Apostolopoulos (Co-PI), G Stanczuk (Co-PI), S Klaczek.

**Norfolk and Norwich University Hospitals NHS Foundation Trust** E Mishra (PI), D Archer, C Atkins, S Bailey, K Bohanan, M Cambell-Kelly, E Chiang, P Clarke, L Coke, B Collinson, J Cook, A Cooper, M Cornwell, A Dann, P De Souza, M Del Forno, A Elsheikh, S Fletcher, C Fraser, S Gajebasia, H Gorick, N Gray, A Haestier, S Hand, M Harmer, L Harris, L Hudig, M Ilsley, K Jethwa, L Jones, A Kamath, D Kelly (Associate PI), J Kennedy, J Keshet-Price, J Knights, E Kolokouri, V Licence, E Lowe, E Malone, M Marks, G Maryan, K Metcalfe, A Miller-Fik, P Moondi, M Morris, K Myint, K Nagumantry, W Ng, J Nortje, M Oliver, M Patel, T Potter, R Rallan, S Rehman, J Roberts, A Sanda-Gomez, A Sanz-Cepero, D Sethi, J Staines, K Stammers, E Tropman, M Ur Rasool, S Walton, N Ward, D Watts, C Websdale, R Wiseman, C Wright.

**North Bristol NHS Trust** N Maskell (PI), H Adamali, R Adhikary (Associate PI), M Alvarez, D Arnold, M Attwood, P Bailey, N Bale, G Bardsley, S Barratt, S Bevins, R Bhatnagar, A Bibby, L Bradshaw, A Brown, C Burden, H Cheshire, S Clarke, A Clive, R Cousins, S Dawson, A Dipper, J Dodd, F Easton, K Farmer, L Fox, L Gethin, P Halford, F Hamilton, J Hardy, D Higbee, S Holloway, A Jayebalan, A Jenkins, L Jennings, S Jones, C Kilby, H Lee, M Martin, H McDill, H McNally, S Merritt, A Milne, K Minou, E Morab, A Morley, G Nava, N Novas Duarte, S Patole, A Pereira, E Perry, S Rafiq, M Rigby, N Rippon, V Sandrey, C Sellar, M Slowinska, K Smith, L Solomon, L Stadon, M Tout, J Townley, R Wach, D Warbrick, C Watkins, H Welch, C Woods.

**North Tees and Hartlepool NHS Foundation Trust** B Prudon (PI), M Abouzaid, C Adams, A Al Aaraj (Associate PI), O Alhabsha, M Ali, E Aliberti, D Ashley, D Barker, H Bashir, B Campbell, A Chilvers, E Chinonso, V Collins, E Connell, K Conroy, E Cox, J Deane, J Dunleavy, I Fenner, C Gan, I Garg, C Gibb, S Gowans, W Hartrey, F Hernandez, J Jacob, V Jagannathan, V Jeebun, S Jones, M Khan, Y Koe, D Leitch, L Magnaye, T Mane, T Mazhani, N McDonnell, M Nafei, B Nelson, L O'Rourke, L Poole, E Poyner, S Purvis, J Quigley, A Ramshaw, H Reynolds, L Robinson, I Ross, R Salmon, L Shepherd, E Siddle, S Sinclair, M Smith, R Srinivasan, K Stewart, R Taylor, G Wallace, S Wang, L Watson, M Weetman, B Wetherill, S Wild, K Win.

**North West Anglia NHS Foundation Trust** K Rege (PI), K Abbas, B Abdelqader, R Abdul-Raheem, A Achara, K Adatia (Associate PI), Z Aftab, C Agbo, A Ahonia, O Akindolie, O Aktinade, A Al Swaifi, S Al-Juboori, F Allan, A Al-Rabahi, R Ambrogetti, S Amin, D Arcoria, K Aung, A Auwal, G Aviss, A Azman Shah, A Babs-Osibodu, K Bahadori, J Baker, T-A Baker, N Bakhtiar, K Bandaru, A Bhanot, K Bhatt, J Bhayani, K Blaylock, T Bodenham, T Bond, H Boughton, H Bowyer, D Braganza, S Brooks, D Butcher, M Butt, N Butterworth-Cowin, R Buttery, A Cabandugama, J Carpenter, P Carter, A Catana, L Cave, R Chaube, M Cheung, Z Chikwanha, C Chisenga, S Choi, S Choudhury, V Christenssen, M Chukwu, B Clearyb, H Cooper, C Cordell, D Corogeanu, A Cowan, R Croysdill, S Dahiya, M Davies, R De Pretto, J de Souza, S Diaz, B Donohue, O Drosos, N Duff, L Dufour, O Ebigbola, C Eddings, M El-Din, I Eletu, N Enachi, J Faccenda, A Feroz, L Fieldhouse, L Finch, N Flaris, C Freer, P Gerard, E Gillham, A Gkoritsa, P Goodyear, R Gooentilleke, R Gosling, G Gosney, V Goss, S Goyal, J Hafsa, W Halford, S Havlik, D Hornan, Z Horne, T Hoskins, M Hunt, C Huson, A Idowu, S Inglis-Hawkes, I Iordanov, M Ishak, M Iyer, D Jafferji, E Jarnell, H Javed, E Johnson, A Jones, D Jones, J Jones, T Jones, E Kallistrou, J Kamara, R Kamath, S Kar, R Kark, N Kasianczuk, R Kassam, D Kaur, A Kerr, S Khalid, A Khan, A Khan, M Kheia, F Kleemann, E Kopyj, G Koshy, A Kozak Eskenazia, R Kurian, S Lahane, M Lami, D Lamrani, K Leng, D Limbu, C Lippold, F Magezi, M Mahmud, A Malik, J Marshall, R Matewe, F Maxton, N McCammon, K McDevitt, S McGuire, C McMurran, W Mensah, P Mitchell, L Molloy, P Morzaria, Q Moyo, I Mugal, O Muraina, N Muru, A Mustafa, Y Myint, S Nazir, O Oderinde, S Ojha, I Okpala, C Oladipo, T Old, G Oleszkiewicz, N Olokoto, H Orme, S O'Sullivan, P Paczko, S Pal, A Pandey, N Pao, N Parker, S Parker, A Patel, S Pathak, S Peacock, B Peirce, T Perumpral, S Ponnusamy, S Pooboni, S Poon, J Porter, A Prasad, A Rahimi, J Rajkanna, M Rather, R Renu Vattekkat, Z Rimba, S Rizvi, P Roberts, S Sahedra, S Saliu, M Samyraju, J Sanyal, V Saulite, R Schofield, S Schofield, K Scholes, S Shah, W Shah, S Shahad, P Sharma, L Simon, L Singh, P Sivasothy, E Smith, S Stacpoole, O Syed, B Tan, K Tan, J Taylor, N Temple, K Thazhatheyil, M Uddin, S Vardy, N Veale, A Waheed Adigun, D Walter, M Walton, B Whelan, A White, G Williams, C Willshire, M Wong, H Wroe.

**North West Boroughs Healthcare NHS Foundation Trust** A Baldwin (PI)

**Northampton General Hospital NHS Trust** M Zaman (PI), B Abdul, A Abdulmumeen, N Abdulshukkoor, A Adnan, M Ahammed Nazeer, B Ahmed, M Ashab, A Bazli, N Benesh, W Chin, N Cunningham, H Daggett, E Davies, E Elmahi, H Enyi, S Fawohunre, J Fernando, N Geoghegan, J Glover, J Hague, K Hall, C Hallett, K Haresh, J Hewertson, J Hosea, N Htoon, H Huynh, M Idrees, C Igwe, H Imtiaz, M Irshad, A Ismail, R Jeffrey, J Jith, P Joshi, R Kaliannan Periyasami, A Khalid, M Khalid, R Kodituwakku, R Kontogonis, P Lopez, A Mahmood, M Malanca, I Mapfunde, V Maruthamuthu, S Masood, M Matharu, A Merchant, F Merchant, S Naqvi, N Natarajan, R Natarajan, K Nawaz, O Ndefo, O Ndefo, O Ogunkeye, S Paranamana, N Pugh, A Raj, K Rashid, M Rogers, M Saad, A Sajid, G Selvadurai, A Shah, M Shahzeb, U Shamji, N Shrestha, A Singh, K Smith, B Sohail, M Spinks, L Stockham, A Takyi, Y Teoh, W Ul Hassan, S Ullah, H Vayalaman, S Wafa, T Ward, R Watson, D White, L Ylquimiche Melly, R Zulaikha.

**Northern Devon Healthcare NHS Trust** R Manhas (PI), C Alexander, K Allen, F Bellis, H Black, S Bosence, A Brunchi, D Dalton, D Davies, J Gilham, N Hadfield, A Hanson, R Hartley, R Holbrook, J Hunt, G Isitt, M Jeelani, M Lamparski, E Lekoudis, S Ley, B Rowlands, C Santos Ferreira De Almeida, A Umeh, L van Koutrik, C Vooght, C White, E Willis, H Wright.

**Northern HSC Trust** P Minnis (PI), W Anderson, S Baird, M BinRofaie, J Burns, C Cruickshank, L Davidson (Associate PI), L Denley, R Donnelly, G Doran, M Drain, A Fryatt, J Gallagher, M Hollyer, M Kawser, L Kingsmore, O Lepiarczyk, I Masih, C McGoldrick, S McKenna, M McMaster, S McNeill, E Murtagh, M Nugdallah, T Scullion, M Stewart, M Wray.

**Northern Lincolnshire and Goole NHS Foundation Trust** A Mitra (PI), R Abrams, N Akhtar, H Al-Moasseb, S Amamou, T Behan, S Biuk, M Brazil, M Brocken, C Burnett, C Chatha, M Cheeseman, L Cottam, T Cruz Cervera, O Davies, K Dent, C Downing, C Dyball, K Edwards, M Elhadi, R Elmahdi, Q Farah, S Farooq, S Gooseman, J Hargraves, M Haroon, J Hatton, E Heeney, J Hill, E Horsley, R Hossain, S Hudson, A Hussien, D Hutchinson, J Hyde-Wyatt, A Ibrahim, M Iqbal, N James, S Khalil, M Madhusudhana, A Marriott, M Masood, G McTaggart, K Mellows, R Miller, U Nasir, M Newton, GCE Ngui, S Pearson, C Pendlebury, R Pollard, N Pothina, D Potoczna, S Raha, A Rehan, SAS Rizvi, A Saffy, J Sanders, H Sangha, K Shams, C Shaw, A Shirgaonkar, S Spencer, R Stead, R Sundhar, D Taylor, E Thein, E Waldeck, L Warnock, K Wong.

**Northumbria Healthcare NHS Foundation Trust** B Yates (PI), C Ashbrook-Raby, A Aujayeb, L Barton, J Bell, S Bourke, H Campbell, D Charlton, K Connelly, D Cooper, A Dawson, L Dismore, S Dodds, C Edwards, S Fearby, V Ferguson, A Green (Associate PI), N Green, H Grover, E Hall, I Hamoodi, S Haney, P Heslop, P Jones, H Lewis, J Luke, L Mackay, C McBrearty, G McCafferty, I McEleavy, H Mckie, N McLarty, U McNelis, A Melville, J Miller, A Morgan, S Parker, L Patterson, H Peggie, S Pick, H Rank (Associate PI), D Ripley, S Robinson, E Rosby, J Rushmer, H Shah, T Smith, V Smith, D Snell, J Steer, E Sykes, A Syndercombe, C Tanney, L Taylor, J Ward, R Warren, M Weatherhead, R Whittle, L Winder.

**Nottingham University Hospitals NHS Trust** W S Lim (PI), M Ali, L Anderson, A Andrews, S Ashraf, D Ashton, G Babington, G Bartlett, D Batra, L Bendall, N Benetti, T Brear, A Buck, G Bugg, J Butler, R Cammack, J Cantliff, L Clark, E Connor, P Davies, M Dent, C Dobson, A Fatemi, M Fatemi, L Fleming, J Grundy, J Hallas, L Hodgen, S Hodgkinson, S Hodgson, L Howard, C Hutchinson, B Jackson, J Kaur, E Keddie-Gray, E Kendall, C Khurana, M Langley, L Lawless, L Looby, M Meredith, L Morris, H Navarra, R Nicol, J Oliver, C Peters, B Petrova, R Purdy, Z Rose, L Ryan, J Sampson, G Squires, J Squires, R Taylor, A Thomas, J Thornton, K Topham, O Vincent, S Warburton, S Wardle, H Waterfall, S Wei, T Wildsmith, L Wilson.

**Pennine Acute Hospitals NHS Trust** J Raw (PI), R Tully (Co-PI), K Abdusamad, Z Antonina, E Ayaz, B Blackledge, P Bradley, F Bray, M Bruce, E Bullock, C Carty, B Charles, G Connolly, C Corbett, J Cornwell, S Dermody, L Durrans, U Elenwa, E Falconer, J Flaherty, C Fox, J Guerin, D Hadfield, J Harris, J Haslam, S Hey (Associate PI), L Hoggett, A Horsley, C Houghton, L Howard-Sandy, S Hussain, R Irving, P Jacob, D Johnstone, R Joseph, P Kamath, T Khatun, T Lamb, H Law, M Lazo, G Lindergard, S Lokanathan, L Macfarlane, S Mathen, S McCullough, P McMaster, D McSorland, J Melville, B Mishra, G Moth, M Mulcahy, S Munt, J Naisbitt, A Neal, R Newport, G O'Connor, D O'Riordan, I Page, V Parambil, J Philbin, M Pinjala, C Rishton, M Riste, J Rothwell, M Sam, Z Sarwar, L Scarratt, A Sengupta, H Sharaf, J Shaw, J Shaw (Associate PI), K Shepherd, A Slack, D Symon, H T-Michael, A Ustianowski, O Walton, S Warran, S Williams.

**Pokhara Academy of Health Sciences** S Baniya (PI), SA Adhikari, SC Baral, A Karki, P KC, S Pathak, KK Roka, A Thapa.

**Poole Hospital NHS Foundation Trust** H Reschreiter (PI), S Bokhandi, J Camsooksai, C Colvin, J Dube, S Grigsby, C Humphrey, S Jenkins, E Langridge, S Patch, M Tighe, L Vinayakarao, B Wadams, M Woolcock.

**Royal Berkshire NHS Foundation Trust** M Frise (PI), R Arimoto, J Armistead, A Aslam, A Barrett, S Bartley, P Bhuie, K Bostock, A Burman, C Camm, R Carson, H Coles, J D'Costa, A Donohoe, E Duffield, F Emond, S Everden, E Gabbitas, E Garden, N Gould, S Gurung Rai, S Hadfield, A Hayat, S Haysom, J Hilton, N Jacques, L Keating, C Knowles, H Lawrence, K Lennon, A McGown, B Mitchell, L Mokogwu, T Okeke, G Parsons, S Rai, L Sathyanarayanan, F Selby, M Thakker, S Vettikumaran, A Walden.

**Royal Brompton & Harefield NHS Foundation Trust** A Shah (PI), A Reed (Co-PI), A Angela, B Araba, L Banton, A Catelan Zborowski, M Damani, P De Sousa, V Jardim, K Mahay, T Maria Pfyl, H Middleton, H Passmore, T Poonian, C Prendergast, P Rogers, G Sloane, N Soussi, J Tan, V Teli, V Thwaiotes, L Tous Sampol, J Wallen, A Watson.

**Royal National Orthopaedic Hospital NHS Trust** R Baumber (PI), D Brooking, F Fitzgerald, E Hanison, J Hunt.

**Royal Papworth Hospital NHS Foundation Trust** R Rintoul (PI), H Barker, F Bottrill, H Brzezicki, G Couch, K Dorey, R Druyeh, M Earwaker, C East, S Fielding, D Finnerty, K Fitzjohn, A Fofana, C Freeman, C Galloway, L Garner, A Gladwell, D Grogono, U Hill, V Hughes, R Hussey, N Jones, T Keady, J Kelliher, C Kosztolanyi, J Krzowski, J Mackie, P Madhivathanan, S Mepham, A Michael, B Moshy, H Munday, M Nizami, J Pack, K Paques, H Parfrey, G Polwarth, J Quijano-Campos, R Staples, S Victor, A Vuylsteke, S Webb, K Woodall, S Woods, J Zamikula.

**RSU Martha Friska** F Ginting (PI), M Barimbing, I Rambe, D Saragih, A Tantri

**RSU Puri Raharja, Denpasar** M Sri (PI), NLPE Arisanti (Co-PI), NKN Andari, NM Artini, P Khairunnisa, R Nurpeni, K Puspatriani, SI Putu, A Rimainar, T Valleri.

**Rumah Sakit Metropolitan Medical Centre, Jakarta** E Nelwan (PI), A Abidin, M Butar, A Cahyareny, L Handayani, H Istiqomah, S Kumala Dewi, T Monika, S Prabowo, N Pratiwi, K Puspatriani, N Rika, A Saraswati, D Sari, P Suherman, T Tesha, J Wanda, K Wattimena, N Yufaniaputri.

**Salford Royal NHS Foundation Trust** P Dark (PI), J Allen, E Apetri, N Bakerly, B Blackledge, L Catlow, B Charles, M Collis, R Doonan, J Harris, A Harvey, K Knowles, S Lee, T Marsden, E Mclaughlan, L McMorrow, A Michael, J Pendlebury, J Perez, S Simpkins, G Squires, R Sukla, M Taylor, V Thomas, D Walker, S Warran, O Wickens (Associate PI).

**Sandwell and West Birmingham Hospitals NHS Trust** S Clare (PI), M Ahmed, Y Beuvink, K Blachford, S Clamp, J Colley, P De, D Devonport, L Fares (Associate PI), M Fenton, B Gammon, M Gohel, R Grenfell, A Hayes, L Henry, S Hussain, S Joseph, F Kinney, T Knight, R Kumar, W Leong, T Lim, B Mahay, M Nelson, Y Nupa, A Orme, W Osborne, Z Pilsworth, S Potter, S Prew, N Rajaiah, A Rajasekaran, H Senya, N Shah, N Shamim, S Sivakumar, L Smith, S Spray, P Thozthumparambil, N Trudgill, A Turner, L Wagstaff, S Willetts, H Willis, M Yan.

**Sheffield Children's NHS Foundation Trust** P Avram (PI), A Bellini, F Blakemore, H Chisem, J Clements, H Cook, S Gormely, D Hawley, C Kerrison, N Lawrence, G Margabanthu, A McMahon, N Roe, F Shackley, J Sowter, T Williams.

**Somerset NHS Foundation Trust** J Pepperell (PI), J Ashcroft, R Burgess, G Chilcott, S Crouch, I Cruickshank, T Dean, J Dryburgh-Jones (Associate PI), J Foot, L Graham, N Grieg, K James, S James, K Johnson, C Lanaghan, D Lewis, A Locke, C Lorimer, J Lucas, H Mills, G Modgil, C Morgan (Associate PI), A Moss, M Nixon, S Northover, K O'Brien, J Page (Associate PI), I Parberry, R Purnell, K Roberts, A Robinson (Associate PI), J Rogers, C Shovelton, C Susan, C Thompson, N Thorne, R Twemlow, C Vickers, R Wallbutton, A Whitcher, J Youens, E Zebracki.

**South Tyneside and Sunderland NHS Foundation Trust** H Grover (PI) (Associate PI), A MacNair (Co-PI), C Brown, A Burns, C Caroline, M Chopra, R Davidson, M Dickson, J Doughty, N Elkaram, I Emmerson, L Fairlie, L Fuller, M Hashimm, J Henderson, K Hinshaw, J Holden, R Hovvels, S Laybourne, K Martin, M McKee, J McKenna, J Moore, N Mullen, P Murphy, L Palmer, G Parish, M Rangar, M Richardson, A Rostron, A Smith, L Smith, L Terry, A Trotter, F Wakinshaw, E Walton, M Walton.

**South Warwickshire NHS Foundation Trust** S Tso (PI), P Parsons (Co-PI), S Bird, B Campbell, G Kakoullis, F Mackie, C O'Brien, P Rai (Associate PI), A Smith, K Webb.

**Southend University Hospital NHS Foundation Trust** G Koduri (PI), F Hayes (Co-PI), V Vijayaraghavan Nalini (Co-PI), S Badhrinarayanan, N Chandran, J Galliford, L Ginn, S Gokaraju V Gupta, P Harman, M Mercioniu, D Qureshi, M S Rabbani (Associate PI).

**Southport and Ormskirk Hospital NHS Trust** S Pintus (PI), A Nune (Co-PI), A Ahmed, H Ahmed, L Bishop, D Dickerson, Z Haslam, E Isherwood, M Jackson, A Morris, M Morrison, R Purves (Associate PI), V Subramanian, A Tageldin (Associate PI).

**St Helens and Knowsley Teaching Hospitals NHS Trust** G Barton (PI), A Tridente (Co-PI), S Burnett, N Collins, S Dealing, R Garr, S Greer, N Hornby, J Keating, J Kirk, S Mayor, A McCairn, S Paul (Associate PI), S Rao, K Shuker, P Stockton, S A I Tay, T Thornton, J Williamson (Associate PI).

**Surrey and Sussex Healthcare NHS Trust** E Potton (PI), S Abbasi, D Acharya, A Acosta, L Ahmed, S Ali, M Alkhusheh, V Amosun, A Arter, M Babi, J Bacon, K Bailey, N Balachandran, S Bandyopadhyam, L Banks, J Barla, T Batty, S Bax, A Belgaumkar, G Benison-Horner, A Boles, N Broomhead, E Cetti, C Chan, I Chaudhry, D Chudgar, J Clark, S Clueit, L Clutterbuck, S Collins, E Combes, G Conway, O Curtis, M Das, M Daschel, S Davies, A Day, M Dhar, K Diaz-Pratt, C Dragan, H Dube, V Duraiswamy, S Edwards, J Elias, A Ellis, T-Y Ellis, J Emmanuel, A Engden, Y Fahmay, B Field, K Fishwick, U Ganesh, C Gilbert, T Giokanini-Royal, E Goudie, S Griffith, S Gurung, R Habibi, C Halevy, A Haqiqi, R Hartley, A Hayman, J Hives, S Holden, M Horsford, S Hughes, C Hui, F Huq, R Hussain, C Iles, L Jackson, N Jain, A James, D Jayaram, E Jessup-Dunton, T Joefield, A Khadar, N Khan, W Kieffer, E Knox, R Kumar, V Kumar, V Kurmars, H Lafferty, F Lamb, R Layug, N Leitch, W Lim, U Limbu, R Loveless, M Mackenzie, N Maghsoodi, S Maher, M Maljk, I Man, N McCarthy, B Mearns, C Mearns, P Morgan, K Morgan-Jones, G Mortem, G Morton, B Moya, G Murphy, S Mutton, A Myers, T Nasser, J Navaratnam, S Nazir, S Nepal, K Nimako, L Nimako, C O'Connor, I Odysseos-Beaumont, A Patel, K Patel, J Penny, V Phongsathorn, P Pillai, M Poole, N Qureshi, I Rajkumar, S Ranjan, A Rehman, T Samuels, S Sathianandan, E Scott, G Sekadde, A Sharma, G Sharp, S Shotton, O Simmons, P Singh, S Smith, K Sri Paranthamen, S Suresh, B Tejero Moya, K Thevarajah, L Thomas, H Timms, N Tomasova, S Tucker, I van Bruggen, S Vara, C Vaz, S Weller, J White, M Wilde, I Wilkinson, C Williams, M Win, D Woosey, D Wright.

**Swansea Bay University Local Health Board** B Healy (PI), M Baker, S Bareford, I Blyth, A Bone, E Brinkworth, R Chudleigh, A Cook, C Davies, Y Ellis, D Evans, E Evans, S Georges, S Green, R Harford, A Holborow, C Johnston, P Jones, M Krishnan, N Leopold, F Morris, A

Mughal, C Murphy, L O'Connell, T Rees, S Richards, M Ryan, G Saleeb, R Stacey, C Thomas, J Travers, P Vallabhaneni, J Watts, M Williams.

**Tameside and Glossop Integrated Care NHS Foundation Trust** B Ryan (PI), J Abbas, A Abraheem, C Afnan, B Ahmed, O Ahmed, R Alhameed, N Amar, M Amsal, M Anim-Somuah, A Armitage, P Arora, N Aziz, C Bann, M Beecroft, P Buckley, T Bull, K Burke, A Butt, N Carter, P Chalakova, S Chandra, M Clark, M Coulding, T Dhorajiwala, S Durogbola, D Enenche, J Fallon, A Fazal, J Foster, I Foulds, V Galvis, N Garlick, H Ghanayem, R Gray, S Gulati, R Hafiz-Ur-Rehman, A Halim, M Hameed, M Hamie, T Heslop, A Hewetson, B Ho, G Hodgetts, V Horsham, W Hughes, W Hulse, A Humphries, M Hussain, N Johal, T Joshi, E Jude, M Kelly, A Kendall-Smith, M Khan, S Kilroy, A Kirk, R Law, C Loan, J Majumdar, A Masud, J McCormick, J McIntosh, V Melnic, O Mercer, T Mirza, J Mukhtar, R Murphy, F Nikita, A Nisar, B Obale, M Pattrick, P Potla, S Pudi, K Qureshi, A Rachid, M Rafique, R Rana, E Rees-Jones, H Rehman, S Ridgway, R Roberts, J Roddy, C Rolls, S Saini, M Sammut, H Savill, M Saxton, M Soliman, S Sommerfield, C Stebbing, J Thomas, A Thornton, V Turner, A Tyzack, A Umate, J Vere, L Whiteley, M Zainab.

**Teku Hospital (Shukraraaj Tropical & Infectious Disease Hospital)** A Bastola (PI), B Chalise (Co-PI), K Maharjan (Co-PI), L Bhandari, U Devkota, A Gupta, S Gyawali, J Khatri, S Mandal, S Pant, K Paudel, M Paudel, S Paudel, A Phuyal, B Poudyal, G Pradip, S Rajbhandari, D Rawal, Y Sapkota.

**The Christie NHS Foundation Trust** V Kasipandian (PI), A Binns, J King, P Mahjoob-Afag, R Mary-Genetu, P Nicola, A Patel, R Shotton, D Sutinyte.

**The Dudley Group NHS Foundation Trust** H Ashby (PI), P Amy, S Ashman-Flavell, S de Silva, J Dean, N Fisher, E Forsey, J Frost, S Jenkins, A John, D Kaur, A Lubina Solomon, S Mahadevan-Bava, T Mahendiran, V Moore, M O'Toole, S Pinches, D Rattehalli, M Reay, U Sinha, L Stanton, M Subramanian, J Vamvakopoulos, S Waidyanatha, S Westwood.

**The Hillingdon Hospitals NHS Foundation Trust** S Kon (PI), T Bate, A Chan, W Chia, A Danga, J Ganapathi, N Hadjisavvas, B Haselden, M Holden, M Ibrahim, E Kam, J Korolewicz, M Kovac, A Lam, H Lamont, G Landers, P Law, N Mahabir, M Majumder, N Malhan, T Nishiyama, P Palanivelu, J Potter, S Ramraj, A Seckington, S Vandeyoon, W Varney, D Wahab, J Winterton, C Woollard.

**The Princess Alexandra Hospital NHS Trust** U Ekeowa (PI), S Sakthi (Co-PI), Q Shah (Co-PI), M Anwar, B Badal, K Bumunarachchi, G Cook, A Daniel, K Dyer, A Easthope, J Finn, C Freer, A Gani, S Harris, E Haworth, L Hughes, K Ixer, N Konar, S Kuckreja, G Lucas, C Muir, P Nabayego, S Naik, O Newman, F Ozdes, R Ragatha, P Russell, R Saha, L Sandhu, E Shpuza, N Staines, S Waring, L Wee, N White, T White, A Zahoor.

**The Queen Elizabeth Hospital, King's Lynn, NHS Foundation Trust** M Blunt (PI), S Abubacker, J Ali, O Aribike, K Beaumont, K Bishop, H Bloxham, P Chan, Z Coton, H Curgenven, M Elsaadany, T Fuller, T Ganeshanathan, M Gigi, G Hasnip, M Iqbal, M Israa, S Jeddi, S Kamerkar, E Lim, E Nadar, K Naguleswaran, O Poluyi, H Rangarajan, G Rewitzky, F Richardson, S Ruff, S Shedwell, E Ukaegbu, A Velusamy, H Webb.

**The Royal Bournemouth and Christchurch Hospitals NHS Foundation Trust** M Schuster Bruce (PI), D Baldwin, N Barratt, Z Clark, M Dale, D Griffiths, E Gunter, A Hogan, S Horler, T Joyce, M Keltos, S Kennard, N Lakeman, R Lee (Associate PI), L Mallon, R Miln, M Mirela, N Moore, S Nix, S Orr, S Pitts, L Purandare, L Rogers, J Samways, L Sankaran (Associate PI), E Stride, H Tiller, L Vamplew, L Wallis, A Wilce.

**The Royal Marsden NHS Foundation Trust** K Tatham (PI), P Angelini, E Bancroft, E Black, A Dela Rosa, E Durie, M Hogben, S Jhanji, I Leslie, A Okines, I Sana, S Shepherd, N Taylor, S Wong.

**The Walton Centre NHS Foundation Trust** R Davies (PI), H Arndt, A Clyne, E Hetherington, G Hull.

**United Lincolnshire Hospitals NHS Trust** M Chablani (PI), R Barber (Co-PI), C Abhinaya, S Archer, M Asghar, S Beck, S Butler, A Chingale, K Dos Santos, P Duckenfield, C Flood, O Francis, C Hewitt, A Hilldrith, K Hubbard, M Hussain, A Judd, S Karelia, A Kirkby, M Kocsor, R Mishra, P Nath, N Ndoumbe, K Netherton, M Okubanjo, L Osborne, H Palmer, G Phalod, N Rana, A Reddy, A Rond-Alliston, B Saint, S Shephardson, A Sloan, R Spencer, K Szymiczek, J Tan, S Tavares.

**University Hospitals Coventry and Warwickshire NHS Trust** K Patel (PI), C Imray (Co-PI), N Aldridge, A Ali, C Bassford, T Brigstock, A Campbell, S Cliff, S Dale, D Davies, G Evans, E French, R Grenfell, S Hewins, D Hewitt, M Hussain, J Jones, A Jose, R Kumar, B Lara, E Mshengu, S Quenby, H Randheva, K Read, P Satodia, E Sear, R Srikantaiah, C Stokes, M Truslove, K Westwood, S Wurie, J Zhang.

**University Hospitals Of Leicester NHS Trust** C Brightling (PI), N Brunskill (Co-PI), M Wiselka (Co-PI), S Adenwalla, P Andreou, H Aung, P Bakoulas, S Bandi, S Batham, T Beaver, K Bhandal, M Bourne, L Boyles, M Cannon, A Charalambou, C Cheung, A Christou, R Cotter, S Debbie, S Diver, A Dunphy, O Elneima, J Fawke, J Finch, N Fowkes, C Gardiner-Hill, G Genato, M Graham-Brown, J Hailstone, C Haines, B Hargadon, H Holdsworth, W Ibrahim, L Ingram, J Jesus Silva, M Joshi, S Kapoor, K Kaul, J Kim, A Kuverji, K Kyriaki, A Lea, T Lee, B Leonard, A Lewszuk, L Lock, K Macconaill, R Major, Y Makkeyah, H McAuley, P McCourt, D Mullasseril Kutten, A Palfreeman, D Pan (Associate PI), E Parker, K Patel, M Patterson, R Phillips, L Plummer, I Qureshi, M Raval, R Russell, H Selvaskandan, S Southin, C Torrance, K Tsilimpari, D Vail, V Wenn, C Wiesender, D Yewatkar, A Yousuf.

**University Hospitals Of North Midlands NHS Trust** T Kemp (PI), J Alexander, W Al-Shamkhani, L Bailey, N Bandla, O Bani-Saad, A Bland, N Bodasing, S Brammer, A Cadwgan, M Chaudhary, L Cheng, M Chikungwa, S Church, F Clark, N Coleman, M Davies, L Diwakar, A Farmer, F Farook, M Ganaie, U Garcia, M Gellamucho, K Glover, M Haris, J Humphries, I Hussain, J Ibrahim, N Idrees (Associate PI), S Khan, L Korcierz, S Krueper, J Lee, S Lord, J Marshall, I Massey, H McCreedy, A McGowan, G Muddegowda, I Mustapha, N Mustfa, K Nettleton, Z Noori, W Osman, H Parker, N Patel, I Ponce, A Quinn, M Ram, A Remegoso, E Sadler, L Scott, T Scott, E Sernicola, N Sheikh, R Shorrocks, R Swift, C Thompson, J Tomlinson, K Tomlinson, R Varquez, L Verueco, L Walker, J Weeratunga, C White, J White, P Wu.

**University Hospitals Plymouth NHS Trust** D Lewis (PI), D Affleck, O Anichtchik, K Bennett, M Chopra, J Corcoran, M Cramp, H Davies, J Day, M Dobranszky Oroian, E Freeman, C Gordon, L Jose, L Madziva, R Mansour, G Marsh, E May, H McDill, P Moodley (Associate PI), C Morton, M Mwadeyi, H Newman, H Notman, C Orr, A Patrick, L Pritchard, G Selby, J Shawe, H Tan, N Thiri Phoo (Associate PI), K Whitehorn.

**Velindre NHS Trust** J Powell (PI), R Adams, A Jackson.

**Warrington and Halton Teaching Hospitals NHS Foundation Trust** M Murthy (PI), R Arya, A Baluwala, T Blunt, R Chan, L Connell, M Davey, L Ditchfield, G Drummond, A Ibrahim, J Little, N Marriott, B Mathew, M Moonan, T Nagarajan, S Patel, H Prady, L Roughley, S Sharma, H Whittle.

**West Hertfordshire Hospitals NHS Trust** R Vancheeswaran (PI), L Norris, V Page, J Palman, A Yousafzar, X Zhao.

**Western HSC Trust** M Kelly (PI), D Concannon, P Corry, K Ferguson, L Gelmon, D Glowski, J Kara, B Keegan, D McClintock, P McDermott, T McManus, V Mortland, F Okpoko, J Pastrana, K Ryan, D Smalls, N Smyth, S Sreenivasan, J Wieboldt.

**Whittington Health NHS Trust** C Parmar (PI), S Ahmed, W Ang, S Ashcroft, H Bateman, L Booker, N Brown, D Burrage, M Christy, P Dlouhy, J Flor, R Fromson, G Fung, K Gilbert, F Green, M Horsford, L Howard, N Ivin, I Jenkin, C Jetha, J Kibaru, M Kousteni, N Kulkarni, A Kyei-Mensah, A Lillis, I Lim, S Lock, C Macfadyen, C Mason, P McCormack, J Merritt, S Mindel, H Montgomery, S Myers, F Nur, L Parker, B Pattenden, A Pender, A Prately, N Prevatt, A Reid, S Rudrakumar, J Sabale, S Sharma, P Sharratt, K Simpson, J Southwell, S Taylor, L Veys, J Webster, N Wolff, D Worley, J Yuan, A Zuriaga-Alvaro.

**Worcestershire Acute Hospitals NHS Trust** C Hooper (PI), K Austin, T Dawson, A Durie, C Hillman-Cooper, O Kelsall, M Ling, Z Parvez, D Stocker, S Stringer, J Thakrar, H Tranter, J Tyler, P Watson, B Wild, D Wilson, H Wood.

**Wrightington, Wigan and Leigh NHS Foundation Trust** A Ashish (PI), I Aziz, C Brown, L Claxton, D Heaton, L Hirst, S Hough, J Jeganathan Ponraj, C Moore, V Parkinson, M Regan, E Robinson, T Taylor, C Tierney, A Verma, N Waddington, J Wedlin, C Williams, C Zipitis.

**York and Scarborough Teaching Hospitals NHS Foundation Trust** D Yates (PI), M Abdelfattah, A Abung, J Anderson, P Antill, S Appleby, P Armtrong, J Azam, T Berriman, D Bull, K Chandler, O Clayton, A Corlett, D Crocombe, S Davies, K Elliott, L Fahel, K Freshwater, J Ghosh, N Gott, D Greenwood, T Holder, K Howard, J Ingham, P Inns, W Lea, E Lindsay, K Mack, N Marshall, S McMeekin, R Miller, R Molyneux, T Momoniat, P Nikolaos, M O'Kane, G Patrick, H Pearson, P Ponnusamy, A Poole, N Price, R Proudfoot, G Purssord, H Redfearn, S Roche, Z Scott, C Sefton, S Shahi, D Smith, B Sohail, E Suleiman, S Sutton, R Thomas, A Turnbull, L Turner, H Watchorn, E Wiafe, J Wilson.

### Supplementary Methods

#### Study organization

The RECOVERY trial is an investigator-initiated, individually randomised, open-label, controlled trial to evaluate the efficacy and safety of a range of putative treatments in patients hospitalized with COVID-19. The protocol is available at www.recoverytrial.net. The trial is being conducted at 177 National Health Service (NHS) hospital organizations in the United Kingdom and hospitals in Vietnam, Nepal, Indonesia, South Africa, India, and Ghana. The trial is coordinated by a team drawn from the Clinical Trial Service Unit and the National Perinatal Epidemiology Clinical Trials Unit within the Nuffield Department of Population Health at University of Oxford, the trial sponsor. Support for local site activities is provided by the National Institute for Health Research Clinical Research Network.

Access to relevant routine health care and registry data is supported by NHS DigiTrials, the Intensive Care National Audit and Research Centre, Public Health Scotland, National Records Service of Scotland, and the Secure Anonymised Information Linkage (SAIL) at University of Swansea.

*Regulatory and ethics approvals*

| Country | Regulatory body | Ethics |
|---|---|---|
| UK | Medicines and Healthcare products Regulatory Agency (MHRA) | Cambridge East Research Ethics Committee (ref: 20/EE/0101) |
| Vietnam | Vietnam Ministry of Health | Hospital for Tropical Diseases Ethics Committee* |
| Nepal | Government of Nepal Department of Drug Administration | Ethical Review Board, Nepal Health Research Council (NHRC) |
| Indonesia | Badan Pengawas Obat Dan Makanan (BPOM) | Ethics Committee of the Faculty of Medicine, University of Indonesia* |
| Ghana | Food and Drugs Authority | Ghana Health Service Ethics Review Committee |
| South Africa | South African Health Products Regulatory Authority (SAHPRA) | The University of the Witwatersrand, Human Research Ethics Committee* |
| India | Not required | ICMR Central Ethics Committee on Human Research (CECHR) |

* For countries without a national ethics committee the name of the committee approving the first site is listed.

#### Laboratory analysis

*Serum anti-SARS-CoV-2 antibodies and nucleocapsid antigen*

Serum samples were collected at randomisation from all UK participants and analysed at the UK Health Security Agency SARS-CoV-2 Serology laboratory, Porton Down, UK. Total antibodies to SARS-CoV-2 spike protein and nucleocapsid protein were measured using the Roche Elecsys Anti-SARS-CoV-2 S and Elecsys Anti-SARS-CoV-2 assays, respectively. Patients were classified as positive or negative using manufacturer defined thresholds. Serum nucleocapsid antigen was measured using the Roche Elecsys SARS-CoV-2 Antigen assay. This assay had previously been validated only on respiratory specimens, so additional validation was performed to evaluate performance on serum samples, including repeatability, linearity, and cross reactivity. Samples were classified as positive or negative using the median value for participants across all antiviral comparisons (COI 0.626).

*Nasal SARS-CoV-2 viral PCR and sequencing*

Nose (mid-turbinate) swabs were collected at randomisation (day 1), on day 3, and on day 5 from all UK patients and analysed at the Oxford University Hospitals microbiology laboratory, Oxford, UK. Swabs had RNA extraction using the Kingfisher Flex system, and SARS-CoV-2 PCR using the TaqPath COVID-19 RT-PCR assay (Thermo Fisher Scientific, Massachusetts, US). A standard curve was produced using a panel of whole virus SARS-CoV-2 standards (Qnostics SARS-CoV-2 Analytical Q Panel), which was used to quantify viral genome copy concentration in samples.

Samples with sufficient viral concentration (2.6 log viral copies/ml) were sequenced using the Oxford Nanopore Technologies Midnight and tiling amplicon protocol, using a GridION instrument (Oxford Nanopore Technologies, Oxford, UK).[1] Sequence data was processed using the Global Pathogen Analysis System (https://gpas.global), a web-based sequence analysis pipeline that uses the Viridian assembly system and assigns viral lineage using Pangolin. Sequence data were evaluated for the presence of mutations associated with molnupiravir or nirmatrelvir resistance, which were identified from literature searches and the Stanford University Coronavirus antiviral & resistance database (https://covdb.stanford.edu/).[2] No mutations associated with molnupiravir resistance were identified, so it was not possible to evaluate samples in the molnupiravir comparison for the presence of resistance. Mutations at 20 nucleotide positions in the 3CL protease gene were identified as being associated with >2.5 fold median reduction in inhibition by nirmatrelvir (positions 25, 49, 138, 140, 142, 143, 144, 165, 166, 167, 168, 172, 173, 188, 189, 192, 194, 252, 297, and 304). No mutations at these positions were identified in any RECOVERY samples in the nirmatrelvir-ritonavir comparison.

1 Constantinides B, Webster H, Gentry J, et al. Rapid turnaround multiplex sequencing of SARS-CoV-2: comparing tiling amplicon protocol performance. medRxiv 2022; : 2021.12.28.21268461.

2 Tzou PL, Tao K, Pond SLK, Shafer RW. Coronavirus Resistance Database (CoV-RDB): SARS-CoV-2 susceptibility to monoclonal antibodies, convalescent plasma, and plasma from vaccinated persons. PLoS One 2022; 17: e0261045.

#### Systematic review methods

*Search Strategy*

We searched Medline, Embase, MedRxiv and the WHO International Clinical Trials Registry Platform between 1 September 2019 and 24 April 2024 for randomised controlled trials comparing the effect of molnupiravir or nirmatrelvir and usual care or placebo in patients hospitalised with COVID-19 using the search terms (Coronavirus infection or COVID or COVID-19 or COVID19 or 2019n-CoV or SARS-COV-2 or SARSCoV2 or SARS-Cov2) AND (molnupiravir or Lagevrio or nirmatrelvir or Paxlovid) and using validated filters to select for randomised controlled trials. No language restrictions were applied.

*Search Results Processing*

Results were screened by researchers experienced in carrying out large-scale systematic reviews and meta-analyses of randomised trials. A trial research clinician reviewed the full texts of shortlisted studies to finalise the list of included studies. The research clinician then performed quality assessment of the included study using the Cochrane Risk of Bias 2 tool.

*Risk of Bias Assessment*

Performed for published studies only, for the outcome of 28-day (or similar) mortality.

| **Study** | **Randomisation process** | **Deviations from intended interventions** | **Missing outcome data** | **Measurement of outcome** | **Selection of reported result** | **Overall** |
|---|---|---|---|---|---|---|
| MOVE-IN[1] | Low | Low | Low | Low | Low | Low |
| Liu et al[2] | Low | Low | Low | Low | Low | Low |

1. Arribas JR, Bhagani S, Lobo SM, et al. Randomized Trial of Molnupiravir or Placebo in Patients Hospitalized with Covid-19. NEJM Evid. 2022 Feb;1(2):EVIDoa2100044. Epub 2021 Dec 16. PMID: 38319178.
2. Liu J, Pan X, Zhang S, et al. Efficacy and safety of Paxlovid in severe adult patients with SARS-Cov-2 infection: a multicenter randomized controlled study. Lancet Reg Health West Pac. 2023 Apr;33:100694. Epub 2023 Feb 6. PMID: 36777445

#### Protocol changes

RECOVERY is a randomised trial among patients hospitalized for COVID-19. All eligible patients receive usual standard of care in the participating hospital and are randomly allocated between no additional treatment and one of several active treatment arms. Over time, additional treatment arms have been added (see Table).

The final protocol relevant to high dose corticosteroids are included in the supplementary material to this publication, together with summaries of the changes made.

##### Table. Protocol changes to COVID-19 treatment comparisons

| Protocol version | Date | Randomisation | Treatment arms |
|---|---|---|---|
| 1.0 | 13-Mar-2020 | Main (part A) | No additional treatment<br>Lopinavir-ritonavir[a]<br>Low-dose corticosteroid[b]<br>Nebulised Interferon-ß-1a (never activated) |
| 2.0 | 23-Mar-2020 | Main (part A) | No additional treatment<br>Lopinavir-ritonavir[a]<br>Low-dose corticosteroid[b]<br>Hydroxychloroquine |
| 3.0 | 07-Apr-2020 | Main (part A) | No additional treatment<br>Lopinavir-ritonavir[a]<br>Low-dose corticosteroid[b]<br>Hydroxychloroquine[c]<br>Azithromycin[d] |
| 4.0 | 14-Apr-2020 | Main (part A)<br><br><br><br><br><br>Second[e,f] | No additional treatment<br>Lopinavir-ritonavir[a]<br>Low-dose corticosteroid[b]<br>Hydroxychloroquine[c]<br>Azithromycin[d]<br><br>No additional treatment<br>Tocilizumab[f] |
| 5.0 | 24-Apr-2020 | - | (no change – extension to children <18 years old) |
| 6.0 | 14-May-2020 | Main (part A)<br><br><br><br><br><br>Main (part B factorial)<br><br><br>Second[e,f] | No additional treatment<br>Lopinavir-ritonavir[a]<br>Low-dose corticosteroid[b]<br>Hydroxychloroquine[c]<br>Azithromycin[d]<br><br>No additional treatment<br>Convalescent plasma<br><br>No additional treatment<br>Tocilizumab[f] |
| 7.0 | 18-Jun-2020 | Main (part A)<br><br><br><br><br>Main (part B factorial)<br><br><br>Second[e,f] | No additional treatment<br>Lopinavir-ritonavir[a]<br>Low-dose corticosteroid[b]<br>Azithromycin[d]<br><br>No additional treatment<br>Convalescent plasma<br><br>No additional treatment<br>Tocilizumab[f] |

| **Protocol version** | **Date** | **Randomisation** | **Treatment arms** |
|---|---|---|---|
| 8.0 | 03-Jul-2020 | Main (part A) | No additional treatment<br>Low-dose corticosteroid[b]<br>Intravenous immunoglobulin[g]<br>High-dose corticosteroid[g]<br>Azithromycin[d] |
| | | Main (part B factorial) | No additional treatment<br>Convalescent plasma |
| | | Second[e,f] | No additional treatment<br>Tocilizumab[f] |
| 9.1 | 18-Sep-2020 | Main (part A) | No additional treatment<br>Low-dose corticosteroid[b]<br>Intravenous immunoglobulin[g]<br>High-dose corticosteroid[g]<br>Azithromycin[d] |
| | | Main (part B factorial) | No additional treatment<br>Convalescent plasma<br>Casirivimab and imdevimab |
| | | Second[e,f] | No additional treatment<br>Tocilizumab[f] |
| 10.1 | 01-Nov-2020 | Main (part A) | No additional treatment<br>Low-dose corticosteroid[b]<br>Intravenous immunoglobulin[g]<br>High-dose corticosteroid[g]<br>Azithromycin[d] |
| | | Main (part B factorial) | No additional treatment<br>Convalescent plasma<br>Casirivimab and imdevimab |
| | | Main (part C factorial) | No additional treatment<br>Aspirin |
| | | Second[e,f] | No additional treatment<br>Tocilizumab[f] |
| 11.1 | 27-Nov-2020 | Main (part A) | No additional treatment<br>Low-dose corticosteroid[b]<br>Intravenous immunoglobulin[g]<br>High-dose corticosteroid[g]<br>Colchicine |
| | | Main (part B factorial) | No additional treatment<br>Convalescent plasma<br>Casirivimab and imdevimab |
| | | Main (part C factorial) | No additional treatment<br>Aspirin |
| | | Second[e,f] | No additional treatment<br>Tocilizumab[f] |

| Protocol version | Date | Randomisation | Treatment arms |
|---|---|---|---|
| 12.1 | 16-Dec-2020 | Main (part A)[h] | No additional treatment<br>Low-dose corticosteroid[b]<br>Intravenous immunoglobulin[g]<br>High-dose corticosteroid[g]<br>Colchicine |
| | | Main (part B factorial)[h] | No additional treatment<br>Convalescent plasma<br>Casirivimab and imdevimab |
| | | Main (part C factorial)[h] | No additional treatment<br>Aspirin |
| | | Second[e,f] | No additional treatment<br>Tocilizumab[f] |
| 13.0 | 26-Jan-2021 | Main (part A)[h] | No additional treatment<br>Low-dose corticosteroid[b]<br>Intravenous immunoglobulin[g]<br>High-dose corticosteroid[g]<br>Colchicine |
| | | Main (part B factorial)[h] | No additional treatment<br>Casirivimab and imdevimab |
| | | Main (part C factorial)[h] | No additional treatment<br>Aspirin |
| | | Main (part D factorial) | No additional treatment<br>Baricitinib |
| | | Second[e,f] | No additional treatment<br>Tocilizumab[f]<br>Anakinra |
| 14.0 | 15-Feb-2021 | Main (part A)[h] | No additional treatment<br>Low-dose corticosteroid[b]<br>Intravenous immunoglobulin[g]<br>High-dose corticosteroid[g]<br>Colchicine<br>Dimethyl fumarate |
| | | Main (part B factorial)[h] | No additional treatment<br>Casirivimab and imdevimab |
| | | Main (part C factorial)[h] | No additional treatment<br>Aspirin |
| | | Main (part D factorial) | No additional treatment<br>Baricitinib |
| | | Second[e,f] | No additional treatment<br>Tocilizumab[f]<br>Anakinra |

| Protocol version | Date | Randomisation | Treatment arms |
|---|---|---|---|
| 15.0 | 12-Apr-2021 | Main (part A)[h] | No additional treatment<br>Low-dose corticosteroid[b]<br>Intravenous immunoglobulin[g]<br>High-dose corticosteroid[g]<br>Dimethyl fumarate |
| | | Main (part B factorial)[h] | No additional treatment<br>Casirivimab and imdevimab |
| | | Main (part D factorial) | No additional treatment<br>Baricitinib<br>Infliximab[j] |
| | | Main (part E factorial)[i] | High-dose dexamethasone[j] |
| | | Second[e,f] | No additional treatment<br>Tocilizumab[f]<br>Anakinra |
| 16.1 | 08-Jul-2021 | Main (part A)[h] | No additional treatment<br>Low-dose corticosteroid[b]<br>Intravenous immunoglobulin[g]<br>High-dose corticosteroid[g]<br>Dimethyl fumarate |
| | | Main (part D factorial) | No additional treatment<br>Baricitinib |
| | | Main (part E factorial)[i] | High-dose dexamethasone[j] |
| | | Main (part F factorial)[l] | Empagliflozin[l] |
| | | Second[e,f] | No additional treatment<br>Tocilizumab[f]<br>Anakinra |
| 17.1 | 10-Aug-2021 | Main (part A)[h] | No additional treatment<br>Dimethyl fumarate |
| | | Main (part D factorial) | No additional treatment<br>Baricitinib |
| | | Main (part E factorial)[i] | High-dose dexamethasone[j] |
| | | Main (part F factorial)[l] | Empagliflozin [l] |
| | | Second[e,f] | No additional treatment<br>Tocilizumab[f]<br>Anakinra |

| **Protocol version** | **Date** | **Randomisation** | **Treatment arms** |
|---|---|---|---|
| 18.1 | 24-Oct-2021 | Main (part A)[h]<br><br><br>Main (part D factorial)<br><br><br>Main (part E factorial)[i]<br><br>Main (part F factorial)[l]<br><br>Second[e,f] | No additional treatment<br>Dimethyl fumarate<br><br>No additional treatment<br>Baricitinib<br><br>High-dose dexamethasone[j]<br><br>Empagliflozin[l]<br><br>No additional treatment<br>Tocilizumab[f]<br>Anakinra |
| 19.1 | 16-Nov-2021 | Main (part D factorial)[k]<br><br><br>Main (part E factorial)[i]<br><br>Main (part F factorial)[l]<br><br>Second[e,f,h] | No additional treatment<br>Baricitinib<br><br>High-dose dexamethasone[j]<br><br>Empagliflozin[l]<br><br>No additional treatment<br>Tocilizumab[f]<br>Anakinra |
| 20.0 | 29-Nov-2021 | Main (part E factorial)[i]<br><br>Main (part F factorial)[l]<br><br>Second[e,f,h] | High-dose dexamethasone[j]<br><br>Empagliflozin[l]<br><br>No additional treatment<br>Tocilizumab[f]<br>Anakinra |
| 21.0 | 17-Dec-2021 | Main (part E factorial)[i]<br><br>Main (part F factorial)[l]<br><br>Main (part J factorial)<br><br>Main (part K factorial)<br><br>Second[e,f,h] | High-dose dexamethasone[j]<br><br>Empagliflozin[l]<br><br>Sotrovimab<br><br>Molnupiravir<br><br>No additional treatment<br>Tocilizumab[f]<br>Anakinra |
| 22.0 | 19-Jan-2022 | Not implemented | |
| 23.0 | 08-Mar-2022 | Main (part E factorial)[i]<br><br>Main (part F factorial)[l]<br><br>Main (part J factorial)<br><br>Main (part K factorial)<br><br>Main (part L factorial) | High-dose dexamethasone[j]<br><br>Empagliflozin[l]<br><br>Sotrovimab<br><br>Molnupiravir<br><br>Nirmatrelvir-ritonavir |
| 24.0 | 13-May-2022 | Not implemented | |

| Protocol version | Date | Randomisation | Treatment arms |
|---|---|---|---|
| 25.0 | 23-May-2022 | Main (part E factorial)[i]<br><br>Main (part F factorial)[l]<br><br>Main (part J factorial)<br><br>Main (part K factorial)<br><br>Main (part L factorial) | High-dose dexamethasone[j]<br><br>Empagliflozin[l]<br><br>Sotrovimab<br><br>Molnupiravir[m]<br><br>Nirmatrelvir-ritonavir[m] |

[a] enrolment ceased 29 June 2020 when the Data Monitoring Committee advised that the Chief Investigators should review the unblinded data.
[b] enrolment of adults ceased 8 June 2020 as more than 2,000 patients had been recruited to the active arm. Enrolment of children ceased on 8 July 2021.
[c] enrolment ceased 5 June 2020 when the Data Monitoring Committee advised that the Chief Investigators should review the unblinded data.
[d] enrolment of adults ceased 27 November 2020 as more than 2,500 patients had been recruited to the active arm
[e] for patients with (a) oxygen saturation <92% on air or requiring oxygen or children with significant systemic disease with persistent pyrexia; and (b) C-reactive protein ≥75 md/L)
[f] enrolment of adults ceased 24 January 2021 as more than 2,000 patients had been recruited to the active arm.
[g] for children only. Enrolment ceased 8 July 2021.
[h] from protocol version 12.1, children could enter the second randomisation regardless of whether they were included in the main randomisation. Enrolment ceased 8 March 2022.
[i] for patients with (a) oxygen saturation <92% on air or requiring oxygen. Enrolment of patients receiving no or simple oxygen ceased on 13 May 2022.
[j] for patients outside UK (until protocol V20.0 when extended to UK)
[k] enrolment ceased 29 December 2021
[l] enrolment ceased on 6 March 2023
[m] enrolment ceased 24 May 2023

#### Main and second randomisation for adults

All RECOVERY trial participants received usual standard of care. On study entry, adult participants initially underwent the Main Randomisation. Trial participants with clinical evidence of progressive COVID-19 (defined as oxygen saturation <92% on room air or requiring oxygen therapy, and C-reactive protein ≥75 mg/L) could be considered for the Second Randomisation at any time up to 21 days after the initial randomisation, and regardless of initial treatment allocation(s). A web-system was used to provide simple randomisation (without stratification or minimisation) with allocation concealment until randomisation had been completed.

Over time, treatment arms were added and removed from the protocol, factorial randomisations were introduced (see below), and not all treatments were available at every hospital. Similarly, not all treatments were deemed by the attending clinician to be suitable for some patients (e.g. due to comorbid conditions or concomitant medication). In any of these cases, randomisation involved fewer arms (and/or fewer factorial elements).

*Main randomisation for adults*

A single participant could be randomised at most to 1 arm from each of part A, B, C, D and E of the factorial randomisations (depending on location), and thus receive between 0 and 4 treatments on top of usual standard of care.

**Part A** (from 19 March 2020)

Eligible participants could be randomised to one of the following arms:

| Treatment arm | Arm opened | Arm closed |
|---|---|---|
| No additional treatment | 19 March 2020 | 12 November 2021 |
| Dexamethasone | 19 March 2020 | 8 June 2020 |
| Lopinavir-ritonavir | 19 March 2020 | 29 June 2020 |
| Hydroxychloroquine | 23 March 2020 | 5 June 2020 |
| Azithromycin | 7 April 2020 | 27 November 2020 |
| Colchicine | 27 November 2020 | 5 March 2021 |
| Dimethyl fumarate | 15 February 2021 | 12 November 2021 |

**Part B** (from 14 May 2020)

Eligible participants could be randomised to one of the following arms:

| Treatment arm | Arm opened | Arm closed |
|---|---|---|
| No additional treatment | 14 May 2020 | 21 May 2021 |
| Convalescent plasma | 14 May 2020 | 15 January 2021 |
| Casirivimab and imdevimab * | 18 September 2020 | 21 May 2021 |

* monoclonal neutralising antibody cocktail

**Part C** (from 1 November 2020)

Eligible participants could be randomised to one of the following arms:

| Treatment arm | Arm opened | Arm closed |
|---|---|---|
| No additional treatment | 1 November 2020 | 21 March 2021 |
| Aspirin | 1 November 2020 | 21 March 2021 |

**Part D** (from 1 November 2020)

Eligible participants could be randomised to one of the following arms (UK only):

| Treatment arm | Arm opened | Arm closed |
|---|---|---|
| No additional treatment | 2 February 2021 | 29 December 2021 |
| Baricitinib | 2 February 2021 | 29 December 2021 |

**Part E** (from 25 May 2021)

Eligible participants could be randomised to one of the following arms:

| Treatment arm | Arm opened | Arm closed |
|---|---|---|
| No additional treatment | 25 May 2021 | Ongoing |
| High-dose dexamethasone | 25 May 2021 | Ongoing (Enrolment of patients receiving no or simple oxygen ceased on 13 May 2022) |

**Part F** (from 8 July 2021)

Eligible participants could be randomised to one of the following arms:

| Treatment arm | Arm opened | Arm closed |
|---|---|---|
| No additional treatment | 8 July 2021 | 6 March 2023 |
| Empagliflozin | 8 July 2021 | 6 March 2023 |

**Part J** (from 29 December 2021)

Eligible participants could be randomised to one of the following arms (UK only):

| Treatment arm | Arm opened | Arm closed |
|---|---|---|
| No additional treatment | 30 December 2021 | Ongoing |
| Sotrovimab | 30 December 2021 | Ongoing |

**Part K** (from 30 December 2021)

Eligible participants could be randomised to one of the following arms:

| Treatment arm | Arm opened | Arm closed |
|---|---|---|
| No additional treatment | 30 December 2021 | 24 May 2023 |
| Molnupiravir | 30 December 2021 | 24 May 2023 |

**Part L** (from 8 March 2022)

Eligible participants could be randomised to one of the following arms (UK only):

| Treatment arm | Arm opened | Arm closed |
|---|---|---|
| No additional treatment | 8 March 2022 | 24 May 2023 |
| Nirmatrelvir-ritonavir | 8 March 2022 | 24 May 2023 |

*Second randomisation for adults (from 14 April 2020)*

From 14 April 2020, a participant could be randomised to one of the following arms and thus receive 0 or 1 treatment on top of those allocated in the initial randomisation and usual standard of care:

| Treatment arm | Arm opened | Arm closed |
|---|---|---|
| No additional treatment | 14 April 2020 | 24 January 2021 |
| Tocilizumab | 14 April 2020 | 24 January 2021 |

**Ascertainment and classification of study outcomes**

Information on baseline characteristics and study outcomes was collected through a combination of electronic case report forms (see below) completed by members of the local research team at each participating hospital and (in the UK) linkage to National Health Service, clinical audit, and other relevant health records. Full details are provided in the RECOVERY Definition and Derivation of Baseline Characteristics and Outcomes Document (see Appendix 3).

*Randomisation form*

The (main) Randomisation form (shown below) was completed by trained study staff. It collected baseline information about the participant (including demographics, COVID-19 history, comorbidities and suitability for the study treatments) and availability of the study treatments. Once completed and electronically signed, the treatment allocation was displayed.

The following modifications were made to the Randomisation form during the trial:

| Randomisation form version | Date of release | Major modifications from previous version |
|---|---|---|
| 1.0 | 19-Mar-20 | Initial version (protocol V1.0) |
| 2.0 | 25-Mar-20 | For protocol V2.0<br>• Hydroxycholoroquine added as treatment<br>• Known long QT syndrome added to comorbidities<br>• Severe depression removed from comorbidities |
| 3.0 | 09-Apr-20 | For protocol V3.0<br>• Azithromycin added as treatment<br>• Suspected SARS-CoV-2 infection included in eligibility criteria |
| [Second randomisation form introduced] | 23-Apr-20 | For protocol 4.0<br>• Eligibility criteria for second randomisation<br>• Tocilizumab vs control as treatment allocations |
| 5.0 | 09-May-20 | For protocol V5.0<br>• Age ≥18 years removed from eligibility criteria<br>• Additional questions on child's age and weight added |
| 6.0 | 21-May-20 | For protocol V6.0<br>• Convalescent plasma added as treatment<br>• Baseline use of remdesivir |
| 7.0 | 01-Jul-20 | For protocol V7.0<br>• Participants eligible if convalescent plasma is only available and suitable treatment |
| 8.0 | 13-Aug-20 | For protocol V8.0<br>• Addition of low-dose and high-dose corticosteroids and intravenous immunoglobulin for children (and removal of dexamethasone for children) |
| 9.0 | 24-Sep-20 | For protocol V9.0<br>• Casirivimab and imdevimab added as treatment<br>• Additional baseline information |
| 10.0 | 06-Nov-20 | For protocol V10.1<br>• Aspirin added as treatment |
| 11.0 | 27-Nov-20 | For protocol V11.1<br>• Colchicine added as treatment |
| 12.0 | 22-Dec-20 | For protocol V12.1<br>• Allow children to enter trial without entering main randomisation |
| 13.0 | 02-Feb-21 | For protocol V13.0<br>• Baricitinib added as treatment |
| 14.0 | 24-Feb-21 | For protocol V14.0<br>• Dimethyl fumarate added as treatment |
| 15.0 | 11-May-21 | For protocol V15.0<br>• High-dose dexamethasone added as treatment |
| 16.0 | 28-Jul-21 | For protocol V16.1<br>• Addition of empagliflozin as treatment |
| 17.0 | 20-Aug-21 | For protocol V17.1<br>• Additional warnings about eligibility for empagliflozin |

| Randomisation form version | Date of release | Major modifications from previous version |
|---|---|---|
| 18.0 | 30-Dec-21 | For protocol V21.1<br>• Sotrovimab and molnupiravir added as treatments<br>• Inclusion of UK participants in high-dose dexamethasone comparison |
| 19.0 | 28-Mar-22 | For protocol V23.0<br>• Nirmatrelvir-ritonavir added as treatment |

RECOVERY
Randomised Evaluation of COVID-19 Therapy

#### Sample Form (v19.00 - 28/03/22)

#### Randomisation Program

Call Freefone **0800 138 5451** to contact the RECOVERY team for **URGENT** problems using the Randomisation Program or for medical advice. All **NON-URGENT queries** should be emailed to ****

Logged in as: **RECOVERY Site**

##### Section A: Baseline and Eligibility

Date and time of randomisation: 27 Mar 2022 14:00

**Treating clinician**

**A1.** Name of treating clinician

**Patient details**

**A2.** Patient surname

Patient forename

**A3.** NHS number — ☐ Tick if not available

**A4.** What is the patient's date of birth? — 01 / January / 2000 Age: 22y 2m

**A5.** What is the patient's sex?

**Inclusion criteria**

**A6.** Has consent been taken in line with the protocol?
If answer is No patient cannot be enrolled in the study
NB current PIS/ICF version is V22.0 (adults) or V14.0 (children)

**A6.0.1** How was consent obtained?

**A6.5** Does this patient have viral pneumonia? — Yes
See protocol for typical features. If answer is No patient cannot be enrolled in the study

**A7.0** Does the patient have proven SARS-CoV-2 infection? — Yes

**A7.0.1** What was lateral flow test result?

**A7.0.2** What was PCR test result?

**A7.1** Does the patient have proven influenza infection?

**A8.** Does the patient have any medical history that might, in the opinion of the attending clinician, put the patient at significant risk if they were to participate in the trial?

**A9.** Symptom onset date: / /

**A10.** Date of hospitalisation: / /

**A11.** Does the patient require oxygen?

**A12.** Please select one of the following to describe the current level of ventilation support

**A12.1** Enter latest oxygen saturation measurement (%)

**A12.2** Enter latest CRP measurement since admission to hospital (mg/L) — ☐ Tick if not measured ☐ Tick if greater than limit of measurement
Enter 0 if below the limit of measurement

**A12.3** Enter latest creatinine measurement since admission to hospital — μmol/L ☐ Tick if not measured

**A12.4** Enter latest D-dimer measurement since admission to hospital — ng/mL ☐ Tick if not measured ☐ Tick if greater than limit of measurement
Enter 0 if below the limit of measurement

**A12.5** Has the patient received a COVID-19 vaccine?

**A12.6** Has the patient received an influenza vaccine in the last 12 months?

**Does the patient have any CURRENT comorbidities or other medical problems or treatments?**

**A13.1** Diabetes

**A13.2** Heart disease

**A13.3** Chronic lung disease

**A13.4** Tuberculosis

**A13.5** HIV

**A13.6** Severe liver disease

**A13.7** Severe kidney impairment (eGFR<30 or on dialysis)

**A13.9.0** Does their clinician consider the patient to be severely immunocompromised?

**A13.12** Has the patient received tocilizumab or sarilumab therapy during this admission?

**A13.14** Current or planned treatment with neuraminidase inhibitor
eg, oseltamivir, zanamivir

**A13.15** Has the patient received casirivimab+imdevimab (Ronapreve) during this illness?

**A13.16** Has the patient received sotrovimab during this illness?

**A13.17** Has the patient received molnupiravir during this illness?

**A13.18** Has the patient received Paxlovid during this illness?

**Are the following treatments UNSUITABLE for the patient?**
**If you answer Yes it means you think this patient should NOT receive this drug.**

**A14E.1** High-dose corticosteroids
NB Please carefully consider suitability of patients already on higher doses (>=7 mg/day dexamethasone or equivalent). Patients eligible for the Paxlovid comparison will be automatically marked as unsuitable for this comparison.

**A14F.1** Empagliflozin
Empagliflozin is NOT suitable if patient (i) has type 1 or post-pancreatectomy diabetes mellitus; or (ii) has a history of ketoacidosis; or (iii) has blood ketones ≥1.5 mmol/L or urine ketones ≥2+; or (iv) is pregnant or breastfeeding
Empagliflozin cannot be given via an enteral feeding tube.

**A14J.1** Sotrovimab

**A14K.1** Molnupiravir
NB Molnupiravir is NOT suitable if patient cannot swallow capsules.

**A14L.1** Paxlovid — No
NB Paxlovid contains ritonavir and has many drug-drug interactions (see protocol and SmPC). Please ensure these have been checked. Paxlovid is NOT suitable if patient cannot swallow tablets. Paxlovid is not suitable for pregnant women in the first trimester.

**Are the following treatments available?**

**A15E.1** High-dose corticosteroids

**A15F.1** Empagliflozin

**A15J.1** Sotrovimab

**A15K.1** Molnupiravir

**A15L.1** Paxlovid — Yes

**Current medication**

**A16.1** Is the patient currently prescribed remdesivir?

**A16.2** Is the patient currently prescribed systemic corticosteroids (dexamethasone, prednisolone, hydrocortisone, methylprednisolone)?
Please do not include topical or inhaled treatments

**A16.5** What venous thromboembolism prophylaxis is the patient receiving?
Standard = usual for hospitalised patients (not increased due to COVID-19); Higher dose = treatment dose or increased prophylaxis due to COVID-19, or oral anticoagulation (eg, warfarin/DOAC).

**A16.6** Is the patient currently prescribed baricitinib (or other JAK inhibitor)?

**Serum sample collection**

**A17.0** Please confirm that patient has had a baseline serum sample collected according to the protocol

**A17.1** Please confirm that patient has had a baseline nasal swab collected according to the protocol

**Please sign off this form once complete**

Surname:

Forename:

Professional email:

Continue

Cancel

Home

*Follow-up form*

The Follow-up form (shown on the next page) collected information on study treatment adherence (including both the randomised allocation and use of other study treatments), vital status (including date and provisional cause of death if available), hospitalisation status (including date of discharge), respiratory support received during the hospitalisation, occurrence of any major cardiac arrhythmias and renal replacement therapy received. Questions on metabolic complications were added in version 16.0, after the high-dose corticosteroid comparison started.

The following modifications were made to the Follow-up form during the trial:

| **Follow-up form version** | **Date of release** | **Modifications from previous version** |
|---|---|---|
| 1.0 | 30-Mar-20 | Initial version |
| 2.0 | 09-Apr-20 | Information on other treatments used during admission:<br>• Azithromycin, IL-6 receptor antagonist<br>Fact and result of SARS-CoV-2 PCR test |
| 3.0 | 09-Apr-20 | Update to functionality; no changes to questions |
| 4.0 | 23-Apr-20 | Duration of treatments added |
| 5.0 | 12-May-20 | Capture of major cardiac arrhythmias added |
| 6.0 | 28-May-20 | Updates to wording of questions.<br>Information on other treatments used during admission:<br>• Remdesivir, convalescent plasma |
| 7.0 | 18-Jun-20 | Clarification of question wording |
| 8.0 | 10-Jul-20 | Information on new treatments for children adherence |
| 9.0 | 24-Sep-20 | Information on casirivimab and imdevimab adherence |
| 10.0 | 06-Nov-20 | Information on aspirin adherence<br>Capture of thrombotic and bleeding events added<br>Information of enrolment into other studies added |
| 11.0 | 16-Nov-20 | Minor changes to in-form validation |
| 12.0 | 27-Nov-20 | Information on colchicine adherence |
| 13.0 | 02-Feb-21 | Information on baricitinib adherence |
| 14.0 | 24-Feb-21 | Additional information on infections |
| 15.0 | 11-May-21 | Information on corticosteroid dosing |
| 16.0 | 28-Jul-21 | Information on empagliflozin adherence<br>Capture of metabolic complications |
| 17.0 | 20-Aug-21 | Additional information on metabolic complications |
| 18.0 | 30-Dec-21 | Information on sotrovimab and molnupiravir adherence<br>Information on sample collection |
| 19.0 | 25-Feb-22 | Information on nirmatrelvir-ritonavir adherence |
| 20.0 | 28-Mar-22 | Information on liver function tests and seizures |
| 21.0 | 31-Mar-22 | Translations added for V19.0 and V20.0 |

### Follow-up

#### Date of randomisation

**Please only report events that occurred from first randomisation until 28 days later on this form (except for Q2).**

**Patient's date of birth** *

yyyy-mm-dd

##### » Vital Status

**0. What is the patient's vital status?** *

- ○ Alive
- ○ Dead

**0.1 What is the patient's current hospitalisation status?** *

- ○ Inpatient
- ○ Discharged

The patient has been enrolled in the trial for **NaN** days

**0.1.1 Date follow-up form completed**

yyyy-mm-dd

**0.1.1 What was the date of discharge?** *

yyyy-mm-dd

**0.1 What was the date of death?** *

yyyy-mm-dd

**0.2 What was the underlying cause of death?** *

*This can be obtained from the last entry in part 1 of the death certificate*

- ○ COVID-19
- ○ Other infection
- ○ Cardiovascular
- ○ Other

**Please give details**

##### » Treatments

**1. Which of the following treatment(s) did the patient definitely receive as part of their hospital admission after randomisation?** *

*(NB Include RECOVERY study-allocated drug, only if given, PLUS any of the other treatments if given as standard hospital care)*

- ☐ No additional treatment
- ☐ Lopinavir-ritonavir

- [ ] Corticosteroid (dexamethasone, prednisolone, hydrocortisone or methylprednisolone)
- [ ] Hydroxychloroquine
- [ ] Azithromycin or other macrolide (eg, clarithromycin, erythromycin)
- [ ] Tocilizumab or sarilumab
- [ ] Remdesivir
- [ ] Intravenous immunoglobulin
- [ ] Synthetic monoclonal antibodies (REGN10933+REGN10987)
- [ ] Aspirin
- [ ] Colchicine
- [ ] Baricitinib
- [ ] Anakinra
- [ ] Favipiravir
- [ ] Empagliflozin
- [ ] Ivermectin
- [ ] Oseltamivir
- [ ] Other neuraminidase inhibitor (e.g. zanamivir, laninamivir)
- [ ] Baloxavir
- [ ] Sotrovimab
- [ ] Molnupiravir
- [ ] Paxlovid

**Please select number of days the patient received corticosteroid (dexamethasone, prednisolone, hydrocortisone or methylprednisolone) (of any dose)**

○ 1 ○ 2 ○ 3 ○ 4 ○ 5 ○ 6 ○ 7 ○ 8 ○ 9 ○ 10

**Dosing information:**

**6 mg dexamethasone is equivalent to 40 mg prednisolone or 160 mg hydrocortisone or 32 mg methylprednisolone.**

**10 mg dexamethasone is equivalent to 67 mg prednisolone or 267 mg hydrocortisone or 53 mg methylprednisolone**

**20 mg dexamethasone is equivalent to 133 mg prednisolone or 534 mg hydrocortisone or 106 mg methylprednisolone**

**Please indicate the highest dose received on a single day during the 10 days after randomisation**

○ <6 mg dexamethasone
○ 6 mg dexamethasone
○ >6 mg and <=10 mg dexamethasone
○ >10 mg and <20 mg dexamethasone
○ 20 mg dexamethasone
○ >20 mg dexamethasone

**Please select number of doses of tocilizumab or sarilumab the patient received**

○ 1 ○ >1

**Please select number of days the patient received remdesivir**

○ 1 ○ 2 ○ 3 ○ 4 ○ 5 ○ 6 ○ 7 ○ 8 ○ 9 ○ 10

**Please select number of days the patient received baricitinib**

1 2 3 4 5 6 7 8 9 10

**Please select number of days the patient received anakinra**

1 2 3 4 5 6 7

**Please select the proportion of days the patient received empagliflozin during the first 28 days after randomisation (or from randomisation to date of discharge if this is sooner)**

Most days (≥90%) Some days (≥50% <90%) Few days (<50% of days, but not zero) None

**Please select number of days the patient received oseltamivir**

1 2 3 4 5 6 7 8 9 10

**Please select number of doses of baloxavir the patient received**

1 2

**Did the participant experience an infusion reaction during or within 2 hours after the sotrovimab infusion?** *

- Yes
- No

**How severe was the reaction?** *

- Mild (no intervention required)
- Moderate (eg, antihistamines or steroids required)
- Severe (adrenaline required)

**Was the infusion completed?** *

- Yes
- No

**Please select the number of days the patient received molnupiravir**

1 2 3 4 5 6

**Was the participant provided with treatment to complete the course at home?**

- Yes
- No

**Please select the number of days the patient received Paxlovid**

1 2 3 4 5 6

**Was the participant provided with treatment to complete the course at home?**

- Yes
- No

**Only required if Q17.0 and or Q17.1 on the Randomisation form were answered Yes**

**Was the baseline serum sample collected?**

- Yes

No

Was the baseline swab samples collected? *

Yes

No

Was the DAY 3 follow-up swab sample collected? *

Yes

No

Swab sent home with patient

Was the DAY 5 follow-up swab sample collected? *

Yes

No

Swab sent home with patient

#### » Ventilation

**4. Did the patient require any form of assisted ventilation (ie, more than just supplementary oxygen) from day of randomisation until 28 days later?** *

Yes

No

**Please answer the following questions:**

**4.1 For how many days did the patient require assisted ventilation?** *

**4.2 What type of ventilation did the patient receive?**

| | Yes | No | Unknown |
|---|---|---|---|
| CPAP alone | ○ | ○ | ○ |
| Non-invasive ventilation (eg, BiPAP) | ○ | ○ | ○ |
| High-flow nasal oxygen (eg, AIRVO) | ○ | ○ | ○ |
| Mechanical ventilation (intubation/tracheostomy) | ○ | ○ | ○ |
| ECMO | ○ | ○ | ○ |

Total number of days the patient received invasive mechanical ventilation (intubation/tracheostomy) from randomisation until discharge/death/28 days after randomisation

#### » Cardiac arrhythmia

**5. Has the patient been documented to have a NEW cardiac arrhythmia at any point since the main randomisation until 28 days later?** *

- Yes
- No
- Unknown

**5.1 Please select all of the following which apply**

- Atrial flutter or atrial fibrillation
- Supraventricular tachycardia
- Ventricular tachycardia (including torsades de pointes)
- Ventricular fibrillation
- Atrioventricular block requiring intervention (eg, cardiac pacing)

#### » Renal outcomes

**6. Did the patient require use of renal dialysis or haemofiltration from main randomisation until 28 days later?** *

- Yes
- No

| 6.1 Please enter the highest creatinine level recorded after randomisation until 28 days later. * | Unit * | Date recorded * | Select if creatinine level not available * |
|---|---|---|---|
| | μmol/L<br>mg/dL | yyyy-mm-dd | Not available |

#### » Thrombosis and bleeding

**7. During the first 28 days after randomisation (or until discharge if sooner), did the participant have a thrombotic event?** *

- Yes
- No
- Unknown

**7.1 Please indicate the type of thrombotic event**

*Select all that apply*

- Pulmonary embolism
- Deep-vein thrombosis
- Ischaemic stroke
- Myocardial infarction
- Systemic arterial embolism
- Other

**8. During the first 28 days after randomisation (or until discharge if sooner), did the participant experience clinically-significant bleeding ie, intra-cranial bleeding or bleeding that required intervention (eg, surgery, endoscopy or vasoactive drugs) or a blood transfusion?** *

- Yes
- No

◯ Unknown

**8.1 Please indicate the site(s) of bleeding** *

*Select all that apply*

☐ Intra-cranial

☐ Gastrointestinal

☐ Other

**8.2 Please indicate which interventions were required to manage the bleed** *

*Select all that apply*

☐ Blood transfusion

☐ Surgery

☐ Endoscopy

☐ Vasoactive drugs (e.g. inotropes on ICU)

☐ None of the above

#### » Other infections

**9. During the first 28 days after randomisation (or until discharge if sooner), did the participant develop another infection?** *

◯ Yes

◯ No

◯ Unknown

**9.1 Please indicate the type of infection**

*Select all that apply*

☐ Pneumonia

☐ Urinary tract

☐ Biliary

☐ Other intra-abdominal

☐ Blood stream

☐ Skin

☐ Other

**Pneumonia - please indicate the putative organism**

◯ Bacterial ◯ Fungal ◯ Viral ◯ Other ◯ Unknown

**Please indicate the virus**

*NB do not record the virus leading to study entry*

☐ SARS-CoV-2 ☐ Influenza ☐ Other/unknown

**Urinary tract - please indicate the putative organism**

◯ Bacterial ◯ Fungal ◯ Other ◯ Unknown

[illegible]

Bacterial Fungal Other Unknown

**Intra-abdominal - please indicate the putative organism**

Bacterial Fungal Other Unknown

**Blood stream - please indicate the putative organism**

*Please only select this if positive blood culture but no known anatomical site found*

Bacterial Fungal Other Unknown

**Skin - please indicate the putative organism**

Bacterial Fungal Viral Other Unknown

**Other - please indicate the putative organism**

Bacterial Fungal Other

Unknown

**Please describe the anatomical site**

#### » Metabolic complications

**10. During the first 28 days after randomisation (or until discharge if sooner), did the participant have any of the following?**

| | Yes | No | Unknown |
|---|---|---|---|
| **Ketoacidosis** * <br> *Ketoacidosis is defined as (i) ketosis (blood ketones ≥1.5 mmol/L or urine ketones ≥2+) AND (ii) metabolic acidosis (eg, bicarbonate <15 mmol/L) AND (iii) no obvious alternative cause of acidosis* | | | |
| **Hyperglycaemic hyperosmolar state** * | | | |
| **Other hyperglycaemia requiring new use of insulin** * | | | |
| **Severe hypoglycaemia** * <br> *Hypoglycaemia causing reduced conscious level requiring another person to help recover.* | | | |

#### » Other safety outcomes

**11. Did the participant experience a seizure after randomisation?** *

Yes

No

Unknown

**11.1 Does the patient have a history of seizures or epilepsy?**

Yes

No

Unknown

**11.2 Please enter the highest ALT (or AST) level recorded after randomisation until 28 days later. If below the limit of detection, enter 0**

| Date * | Result * | Upper limit of normal * | Units |
|---|---|---|---|

| | | | |
|---|---|---|---|
| yyyy-mm-dd | | | IU/L or U/L<br>µmol/L<br>µkat/L |

**11.3 Please enter the highest bilirubin level recorded after randomisation until 28 days later. If below the limit of detection, enter 0**

| Date * | Result * | Upper limit of normal * | Units |
|---|---|---|---|
| yyyy-mm-dd | | | µmol/L<br>mg/dL |

#### » Other trials

**12. Please indicate if the participant participated in any other COVID-19 or influenza trials**

*Select all that apply*

- [ ] PRINCIPLE
- [ ] REMAP-CAP
- [ ] Other treatment trial(s)
- [ ] COVID-19 vaccine trial(s)

**Please give name of other treatment trial(s)**

**Please give name of COVID-19 vaccine trial(s)**

#### » Pregnancy

**13. If this woman was pregnant at randomisation (or had recently delivered), please enter UKOSS ID here.**

*Enter the full UKOSS case ID eg, COR_123*

*28 vital status form*

For sites outside the UK a further case report form collected vital status at day 28 (if not already reported on follow-up form).

### 28 Day Vital Status

**The vital status of this patient has been recorderd as died on the Follow-up form. Please do not enter any data and close the form**

**1. Please confirm participant's date of birth** *

yyyy-mm-dd

**2. Please indicate the participant's current vital status** *

◯ Alive ◯ Dead ◯ Unknown

**This form should be completed when participant is known to have died or after . Please re-try then**

**Are you sure you are unable to establish the vital status for this participant?**

**2.1 Please enter the date on which they were last known to be alive**

yyyy-mm-dd

**2.1. What was the date of death?**

yyyy-mm-dd

**2.2. Please select likely cause of death**

◯ COVID-19 ◯ Other infection ◯ Cardiovascular ◯ Other

**Interim analyses: role of the Data Monitoring Committee**

The independent Data Monitoring Committee reviewed unblinded analyses of the study data and any other information considered relevant at intervals of around 2-3 months. The committee was charged with determining if, in their view, the randomised comparisons in the study provide evidence on mortality that is strong enough (with a range of uncertainty around the results that was narrow enough) to affect national and global treatment strategies. In such a circumstance, the Committee would inform the Steering Committee who would make the results available to the public and amend the trial arms accordingly. Unless that happened, the Steering Committee, investigators, and all others involved in the trial would remain blind to the interim results until 28 days after the last patient had been randomised to a particular intervention arm. Further details about the role and membership of the independent Data Monitoring Committee are provided in the protocol.

The Data Monitoring Committee determined that to consider recommending stopping a treatment early for benefit would require at least a 3 to 3·5 standard error reduction in mortality. The Committee concluded that examinations of the data at every 10% (or even 5%) of the total data would lead to only a marginal increase in the overall type I error rate.

Between 30 December 2021 and 24 May 2023 the DMC reviewed data on molnupiravir and nirmatrelvir-ritonavir 7 times.

### Supplementary Tables

#### Webtable 1: Baseline characteristics of patients considered unsuitable for randomization to molnupiravir compared with those randomized to molnupiravir versus usual care

| | Randomized (n=923) | Considered unsuitable or not available (n=319) |
|---|---|---|
| Age, years | 71.4 (14.1) | 72.1 (16.1) |
| <70 | 362 (39%) | 121 (38%) |
| ≥70 to <80 | 282 (31%) | 83 (26%) |
| ≥80 | 279 (30%) | 115 (36%) |
| Sex | | |
| Male | 547 (59%) | 188 (59%) |
| Female* | 376 (41%) | 131 (41%) |
| Not recorded | 0 (0%) | 0 (0%) |
| Country | | |
| Ghana | 0 (0%) | 0 (0%) |
| India | 0 (0%) | 0 (0%) |
| Indonesia | 12 (1%) | 0 (0%) |
| Nepal | 18 (2%) | 2 (1%) |
| South Africa | 0 (0%) | 0 (0%) |
| Vietnam | 0 (0%) | 0 (0%) |
| UK | 893 (97%) | 317 (99%) |
| Ethnicity | | |
| White | 809 (88%) | 277 (87%) |
| Black, Asian, and minority ethnic | 77 (8%) | 23 (7%) |
| Unknown | 37 (4%) | 19 (6%) |
| Number of days since symptom onset | 5 (3-9) | 5 (3-10) |
| Number of days since hospitalisation | 2 (1-4) | 3 (1-7) |
| Respiratory support received | | |
| None | 160 (17%) | 48 (15%) |
| Simple oxygen | 602 (65%) | 180 (56%) |
| Non invasive ventilation | 159 (17%) | 64 (20%) |
| Invasive mechanical ventilation | 2 (<0.5%) | 27 (8%) |
| Biochemistry | | |
| C-reactive protein, mg/L | 69 (28-136) | 79 (29-150) |
| Creatinine, umol/L | 80 (64-112) | 78 (59-110) |
| Previous diseases | | |
| Diabetes | 248 (27%) | 77 (24%) |
| Heart disease | 309 (33%) | 106 (33%) |
| Chronic lung disease | 380 (41%) | 102 (32%) |
| Tuberculosis | 2 (<0.5%) | 1 (<0.5%) |
| HIV | 6 (1%) | 0 (0%) |
| Severe liver disease† | 17 (2%) | 7 (2%) |
| Severe kidney impairment‡ | 77 (8%) | 35 (11%) |
| Severely immunocompromised$ | 183 (20%) | 96 (30%) |
| Any of the above | 710 (77%) | 242 (76%) |
| SARS-CoV-2 PCR test result | | |
| Positive | 923 (100%) | 319 (100%) |
| Negative | 0 (0%) | 0 (0%) |
| Unknown | 0 (0%) | 0 (0%) |
| Received a COVID-19 vaccine | 767 (83%) | 266 (83%) |
| Use of other treatments | | |
| Corticosteroids | 809 (88%) | 285 (89%) |
| Remdesivir | 372 (40%) | 132 (41%) |
| Tocilizumab | 105 (11%) | 49 (15%) |
| Plan to use tocilizumab within the next 24 hours | 72 (8%) | 13 (4%) |
| Other randomly assigned treatments | | |
| High dose steriods | 156 (17%) | 40 (13%) |
| Empagliflozin | 281 (30%) | 36 (11%) |
| Sotrovimab | 420 (46%) | 139 (44%) |
| Molnupiravir | 445 (48%) | 0 (0%) |

| | Randomized (n=923) | Considered unsuitable or not available (n=319) |
|---|---|---|
| Nirmatrelvir-ritonavir | 67 (7%) | 1 (<0.5%) |
| Viral load | | |
| Median level (Log viral copies/ml) | 6 (4-7) | 6 (4-7) |
| Antigen status | | |
| Positive | 438 (47%) | 3 (1%) |
| Negative | 414 (45%) | 1 (<0.5%) |
| Unknown | 71 (8%) | 315 (99%) |
| Serostatus (anti N) | | |
| Positive | 227 (25%) | 0 (0%) |
| Negative | 626 (68%) | 4 (1%) |
| Unknown | 70 (8%) | 315 (99%) |
| Serostatus (anti S) | | |
| Positive | 705 (76%) | 3 (1%) |
| Negative | 147 (16%) | 1 (<0.5%) |
| Unknown | 71 (8%) | 315 (99%) |

Results are count (%), mean ± standard deviation, or median (inter-quartile range). †Defined as requiring ongoing specialist care. ‡Defined as estimated glomerular filtration rate <30 mL/min/1.73m² $In the opinion of the managing clinician.

**Webtable 2: Baseline characteristics of patients considered unsuitable for randomization to nirmatrelvir-ritonavir compared with those randomized to nirmatrelvir-ritonavir versus usual care**

| | Randomized (n=137) | Considered unsuitable or not available (n=357) |
|---|---|---|
| Age, years | 72.5 (13.9) | 72.8 (14.9) |
| <70 | 48 (35%) | 128 (36%) |
| ≥70 to <80 | 48 (35%) | 98 (27%) |
| ≥80 | 41 (30%) | 131 (37%) |
| Sex | | |
| Male | 77 (56%) | 200 (56%) |
| Female* | 60 (44%) | 157 (44%) |
| Not recorded | 0 (0%) | 0 (0%) |
| Country | | |
| Ghana | 0 (0%) | 0 (0%) |
| India | 0 (0%) | 0 (0%) |
| Indonesia | 0 (0%) | 0 (0%) |
| Nepal | 0 (0%) | 0 (0%) |
| South Africa | 0 (0%) | 0 (0%) |
| Vietnam | 0 (0%) | 0 (0%) |
| UK | 137 (100%) | 357 (100%) |
| Ethnicity | | |
| White | 121 (88%) | 315 (88%) |
| Black, Asian, and minority ethnic | 7 (5%) | 25 (7%) |
| Unknown | 9 (7%) | 17 (5%) |
| Number of days since symptom onset | 4 (3-8) | 4 (2-8) |
| Number of days since hospitalisation | 2 (1-4) | 3 (1-8) |
| Respiratory support received | | |
| None | 21 (15%) | 46 (13%) |
| Simple oxygen | 92 (67%) | 234 (66%) |
| Non invasive ventilation | 24 (18%) | 67 (19%) |
| Invasive mechanical ventilation | 0 (0%) | 10 (3%) |
| Biochemistry | | |
| C-reactive protein, mg/L | 86 (37-157) | 78 (31-140) |
| Creatinine, umol/L | 74 (62-99) | 81 (61-116) |
| Previous diseases | | |
| Diabetes | 33 (24%) | 95 (27%) |
| Heart disease | 28 (20%) | 134 (38%) |
| Chronic lung disease | 53 (39%) | 140 (39%) |
| Tuberculosis | 1 (1%) | 0 (0%) |
| HIV | 1 (1%) | 3 (1%) |
| Severe liver disease† | 0 (0%) | 14 (4%) |
| Severe kidney impairment‡ | 0 (0%) | 50 (14%) |
| Severely immunocompromised$ | 29 (21%) | 90 (25%) |
| Any of the above | 97 (71%) | 284 (80%) |
| SARS-CoV-2 PCR test result | | |
| Positive | 137 (100%) | 357 (100%) |
| Negative | 0 (0%) | 0 (0%) |
| Unknown | 0 (0%) | 0 (0%) |
| Received a COVID-19 vaccine | 116 (85%) | 292 (82%) |
| Use of other treatments | | |
| Corticosteroids | 122 (89%) | 311 (87%) |
| Remdesivir | 58 (42%) | 160 (45%) |
| Tocilizumab | 25 (18%) | 38 (11%) |
| Plan to use tocilizumab within the next 24 hours | 10 (7%) | 18 (5%) |
| Other randomly assigned treatments | | |
| High dose steriods | 0 (0%) | 25 (7%) |
| Empagliflozin | 39 (28%) | 60 (17%) |
| Sotrovimab | 65 (47%) | 159 (45%) |
| Molnupiravir | 66 (48%) | 106 (30%) |

| | Randomized (n=137) | Considered unsuitable or not available (n=357) |
|---|---|---|
| Nirmatrelvir-ritonavir | 68 (50%) | 0 (0%) |
| Viral load | | |
| Median level (Log viral copies/ml) | 6 (4-7) | 5 (3-7) |
| Antigen status | | |
| Positive | 73 (53%) | 99 (28%) |
| Negative | 61 (45%) | 111 (31%) |
| Unknown | 3 (2%) | 147 (41%) |
| Serostatus (anti N) | | |
| Positive | 40 (29%) | 66 (18%) |
| Negative | 94 (69%) | 144 (40%) |
| Unknown | 3 (2%) | 147 (41%) |
| Serostatus (anti S) | | |
| Positive | 112 (82%) | 186 (52%) |
| Negative | 22 (16%) | 24 (7%) |
| Unknown | 3 (2%) | 147 (41%) |

Results are count (%), mean ± standard deviation, or median (inter-quartile range). †Defined as requiring ongoing specialist care. ‡Defined as estimated glomerular filtration rate <30 mL/min/1.73m² $In the opinion of the managing clinician.

#### Webtable 3: Treatments given during the follow-up period, by randomized allocation

| | Molnupiravir vs usual care | | Nirmatrelvir-ritonavir vs usual care | |
|---|---|---|---|---|
| | Molnupiravir (n=445) | Usual care (n=478) | Nirmatrelvir-ritonavir (n=68) | Usual care (n=69) |
| Compliance data available | 443 | 472 | 67 | 68 |
| Received molnupiravir | 413 (93%) | 0 (0%) | 31 (46%) | 30 (44%) |
| Received nirmatrelvir-ritonavir | 35 (8%) | 35 (7%) | 60 (90%) | 0 (0%) |
| Other treatments received | | | | |
| Dexamethasone | 366 (83%) | 382 (81%) | 55 (82%) | 51 (75%) |
| Azithromycin or other macrolide | 64 (14%) | 64 (14%) | 8 (12%) | 10 (15%) |
| Tocilizumab or sarilumab | 47 (11%) | 51 (11%) | 5 (7%) | 15 (22%) |
| Remdesivir | 153 (35%) | 174 (37%) | 25 (37%) | 28 (41%) |
| Casirivimab+imdevimab | 0 (0%) | 0 (0%) | 0 (0%) | 0 (0%) |
| Aspirin | 49 (11%) | 49 (10%) | 4 (6%) | 6 (9%) |
| Colchicine | 1 (<1%) | 0 (0%) | 0 (0%) | 0 (0%) |
| Baricitinib | 25 (6%) | 22 (5%) | 11 (16%) | 13 (19%) |
| Empagliflozin | 132 (30%) | 122 (26%) | 16 (24%) | 19 (28%) |
| Sotrovimab | 191 (43%) | 199 (42%) | 26 (39%) | 33 (49%) |

Percentages are of those with a completed follow-up form. 60/70 (86%) of participants in the molnupiravir comparison who received nirmatrelvir-ritonavir had been randomly allocated to the nirmatrelvir-ritonavir arm. 61/61 (100%) of the participants in the nirmatrelvir-ritonavir comparison who received molnupiravir had been randomly allocated to the molnupiravir arm.

#### Webtable 4: Effect of allocation to molnupiravir and nirmatrelvir-ritonavir on cause-specific 28-day mortality

| | Molnupiravir vs usual care | | | Nirmatrelvir-ritonavir vs usual care | | |
|---|---|---|---|---|---|---|
| | Molnupiravir (n=445) | Usual care (n=478) | Absolute percent difference (95% CI) | Nirmatrelvir-ritonavir (n=68) | Usual care (n=69) | Absolute percent difference (95% CI) |
| COVID | 57 (12.8%) | 55 (11.5%) | 1.3 (-2.9,5.5) | 9 (13.2%) | 9 (13.0%) | 0.2 (-11.1,11.5) |
| Other infection | 0 (0.0%) | 2 (0.4%) | -0.4 (-1.0,0.2) | 1 (1.5%) | 0 (0.0%) | 1.5 (-1.4,4.3) |
| Cardiac | 1 (0.2%) | 5 (1.0%) | -0.8 (-1.8,0.2) | 0 (0.0%) | 1 (1.4%) | -1.4 (-4.3,1.4) |
| Stroke | 1 (0.2%) | 0 (0.0%) | 0.2 (-0.2,0.7) | 0 (0.0%) | 0 (0.0%) | 0.0 (0.0,0.0) |
| Other vascular | 0 (0.0%) | 1 (0.2%) | -0.2 (-0.6,0.2) | 0 (0.0%) | 0 (0.0%) | 0.0 (0.0,0.0) |
| Cancer | 4 (0.9%) | 5 (1.0%) | -0.1 (-1.4,1.1) | 0 (0.0%) | 2 (2.9%) | -2.9 (-6.9,1.1) |
| Other medical | 10 (2.2%) | 10 (2.1%) | 0.2 (-1.7,2.0) | 2 (2.9%) | 1 (1.4%) | 1.5 (-3.4,6.4) |
| External | 1 (0.2%) | 1 (0.2%) | 0.0 (-0.6,0.6) | 1 (1.5%) | 0 (0.0%) | 1.5 (-1.4,4.3) |
| Unknown cause | 0 (0.0%) | 0 (0.0%) | 0.0 (0.0,0.0) | 0 (0.0%) | 0 (0.0%) | 0.0 (0.0,0.0) |
| All-cause | 74 (16.6%) | 79 (16.5%) | 0.1 (-4.7,4.9) | 13 (19.1%) | 13 (18.8%) | 0.3 (-12.9,13.4) |

**Webtable 5: Effect of allocation to molnupiravir and nirmatrelvir-ritonavir on new cardiac arrhythmia, thrombotic events, clinically significant bleeds, non-coronavirus infections and metabolic complications**

| | Molnupiravir vs usual care | | | Nirmatrelvir-ritonavir vs usual care | | |
|---|---|---|---|---|---|---|
| | **Molnupiravir (n=445)** | **Usual care (n=478)** | **Absolute percent difference (95% CI)** | **Nirmatrelvir-ritonavir (n=68)** | **Usual care (n=69)** | **Absolute percent difference (95% CI)** |
| Number with follow-up form | 443 | 472 | | 67 | 68 | |
| New cardiac arrhythmia | | | | | | |
| Atrial flutter or fibrillation | 11 (2.5%) | 12 (2.5%) | -0.0 (-2.1,2.0) | 3 (4.4%) | 1 (1.4%) | 3.0 (-2.7,8.6) |
| Other supraventricular tachycardia | 1 (0.2%) | 0 (0.0%) | 0.2 (-0.2,0.7) | 0 (0.0%) | 0 (0.0%) | 0.0 (0.0,0.0) |
| Subtotal: Supraventricular tachycardia | 12 (2.7%) | 12 (2.5%) | 0.2 (-1.9,2.2) | 3 (4.4%) | 1 (1.4%) | 3.0 (-2.7,8.6) |
| Ventricular tachycardia | 1 (0.2%) | 1 (0.2%) | 0.0 (-0.6,0.6) | 1 (1.5%) | 0 (0.0%) | 1.5 (-1.4,4.3) |
| Ventricular fibrillation | 0 (0.0%) | 1 (0.2%) | -0.2 (-0.6,0.2) | 0 (0.0%) | 0 (0.0%) | 0.0 (0.0,0.0) |
| Subtotal: Ventricular tachycardia or fibrillation | 1 (0.2%) | 2 (0.4%) | -0.2 (-0.9,0.5) | 1 (1.5%) | 0 (0.0%) | 1.5 (-1.4,4.3) |
| Atrioventricular block requiring intervention | 0 (0.0%) | 1 (0.2%) | -0.2 (-0.6,0.2) | 1 (1.5%) | 0 (0.0%) | 1.5 (-1.4,4.3) |
| Total: Any major cardiac arrhythmia | 13 (2.9%) | 15 (3.1%) | -0.2 (-2.4,2.0) | 5 (7.4%) | 1 (1.4%) | 5.9 (-0.9,12.7) |
| Thrombotic events | | | | | | |
| Pulmonary embolism | 6 (1.3%) | 6 (1.3%) | 0.1 (-1.4,1.6) | 1 (1.5%) | 1 (1.4%) | 0.0 (-4.0,4.0) |
| Deep-vein thrombosis | 2 (0.4%) | 2 (0.4%) | 0.0 (-0.8,0.9) | 0 (0.0%) | 0 (0.0%) | 0.0 (0.0,0.0) |
| Ischaemic stroke | 1 (0.2%) | 1 (0.2%) | 0.0 (-0.6,0.6) | 0 (0.0%) | 0 (0.0%) | 0.0 (0.0,0.0) |
| Myocardial infarction | 0 (0.0%) | 2 (0.4%) | -0.4 (-1.0,0.2) | 1 (1.5%) | 0 (0.0%) | 1.5 (-1.4,4.3) |
| Systemic arterial embolism | 0 (0.0%) | 0 (0.0%) | 0.0 (0.0,0.0) | 0 (0.0%) | 0 (0.0%) | 0.0 (0.0,0.0) |
| Subtotal: Any thrombotic event | 9 (2.0%) | 10 (2.1%) | -0.1 (-1.9,1.8) | 2 (2.9%) | 1 (1.4%) | 1.5 (-3.4,6.4) |
| Clinically significant bleeds | | | | | | |
| Intra-cranial | 0 (0.0%) | 1 (0.2%) | -0.2 (-0.6,0.2) | 1 (1.5%) | 0 (0.0%) | 1.5 (-1.4,4.3) |
| Gastrointestinal | 3 (0.7%) | 5 (1.0%) | -0.4 (-1.6,0.8) | 1 (1.5%) | 1 (1.4%) | 0.0 (-4.0,4.0) |
| Other/unrecorded site | 1 (0.2%) | 5 (1.0%) | -0.8 (-1.8,0.2) | 0 (0.0%) | 2 (2.9%) | -2.9 (-6.9,1.1) |
| Requiring blood transfusion | 4 (0.9%) | 6 (1.3%) | -0.4 (-1.7,1.0) | 1 (1.5%) | 3 (4.3%) | -2.9 (-8.5,2.7) |
| Requiring surgery | 0 (0.0%) | 2 (0.4%) | -0.4 (-1.0,0.2) | 0 (0.0%) | 0 (0.0%) | 0.0 (0.0,0.0) |
| Requiring endoscopy | 2 (0.4%) | 1 (0.2%) | 0.2 (-0.5,1.0) | 0 (0.0%) | 1 (1.4%) | -1.4 (-4.3,1.4) |
| Requiring vasoactive drugs | 0 (0.0%) | 1 (0.2%) | -0.2 (-0.6,0.2) | 0 (0.0%) | 0 (0.0%) | 0.0 (0.0,0.0) |
| Subtotal: Any clinically significant bleeding | 4 (0.9%) | 11 (2.3%) | -1.4 (-3.0,0.2) | 2 (2.9%) | 3 (4.3%) | -1.4 (-7.7,4.9) |
| Non-coronavirus infection | | | | | | |
| Pneumonia | 42 (9.4%) | 40 (8.4%) | 1.1 (-2.6,4.7) | 5 (7.4%) | 2 (2.9%) | 4.5 (-2.9,11.8) |
| Urinary tract | 9 (2.0%) | 8 (1.7%) | 0.3 (-1.4,2.1) | 1 (1.5%) | 0 (0.0%) | 1.5 (-1.4,4.3) |
| Biliary | 0 (0.0%) | 0 (0.0%) | 0.0 (0.0,0.0) | 0 (0.0%) | 0 (0.0%) | 0.0 (0.0,0.0) |
| Other intra-abdominal | 2 (0.4%) | 3 (0.6%) | -0.2 (-1.1,0.8) | 0 (0.0%) | 0 (0.0%) | 0.0 (0.0,0.0) |
| Blood stream | 3 (0.7%) | 0 (0.0%) | 0.7 (-0.1,1.4) | 0 (0.0%) | 0 (0.0%) | 0.0 (0.0,0.0) |
| Skin | 2 (0.4%) | 2 (0.4%) | 0.0 (-0.8,0.9) | 0 (0.0%) | 0 (0.0%) | 0.0 (0.0,0.0) |
| Other | 11 (2.5%) | 13 (2.7%) | -0.2 (-2.3,1.8) | 1 (1.5%) | 3 (4.3%) | -2.9 (-8.5,2.7) |
| Subtotal: Any non-coronavirus infection | 63 (14.2%) | 62 (13.0%) | 1.2 (-3.2,5.6) | 7 (10.3%) | 5 (7.2%) | 3.0 (-6.4,12.5) |
| Metabolic complications | | | | | | |
| Ketoacidosis | 1 (0.2%) | 0 (0.0%) | 0.2 (-0.2,0.7) | 1 (1.5%) | 0 (0.0%) | 1.5 (-1.4,4.3) |
| Hyperglycaemic hyperosmolar state | 6 (1.3%) | 2 (0.4%) | 0.9 (-0.3,2.1) | 2 (2.9%) | 1 (1.4%) | 1.5 (-3.4,6.4) |
| Other hyperglycaemia requiring new use of insulin | 28 (6.3%) | 13 (2.7%) | 3.6 (0.9,6.3) | 3 (4.4%) | 3 (4.3%) | 0.1 (-6.8,6.9) |
| Subtotal: Any hyperglycaemia requiring insulin | 33 (7.4%) | 15 (3.1%) | 4.3 (1.4,7.2) | 6 (8.8%) | 3 (4.3%) | 4.5 (-3.8,12.8) |
| Severe hypoglycaemia | 6 (1.3%) | 1 (0.2%) | 1.1 (-0.0,2.3) | 0 (0.0%) | 1 (1.4%) | -1.4 (-4.3,1.4) |
| Acute kidney injury* | | | | | | |
| Stage 1 | 17/436 (3.9%) | 15/469 (3.2%) | 0.7 (-1.7,3.1) | 3/68 (4.4%) | 1/69 (1.4%) | 3.0 (-2.7,8.6) |
| Stage 2 | 4/436 (0.9%) | 11/469 (2.3%) | -1.4 (-3.1,0.2) | 0/68 (0.0%) | 1/69 (1.4%) | -1.4 (-4.3,1.4) |
| Stage 3 | 12/436 (2.8%) | 16/469 (3.4%) | -0.7 (-2.9,1.6) | 2/68 (2.9%) | 1/69 (1.4%) | 1.5 (-3.4,6.4) |
| Subtotal: Any AKI | 33/436 (7.6%) | 42/469 (9.0%) | -1.4 (-5.0,2.2) | 5/68 (7.4%) | 3/69 (4.3%) | 3.0 (-4.8,10.9) |
| Other outcomes | | | | | | |
| Seizures | 2 (0.4%) | 1 (0.2%) | 0.2 (-0.5,1.0) | 0 (0.0%) | 0 (0.0%) | 0.0 (0.0,0.0) |
| ALT >3x ULN | 18 (6.1%) | 25 (8.4%) | -2.3 (-6.4,1.9) | 6 (10.3%) | 4 (7.1%) | 3.2 (-7.1,13.5) |
| ALT >3x ULN and bilirubin >2x ULN | 0 (0.0%) | 4 (1.4%) | -1.4 (-2.7,-0.0) | 0 (0.0%) | 0 (0.0%) | 0.0 (0.0,0.0) |

*Analyses exclude those on haemodialysis or haemofiltration at randomisation.

### Supplementary Figures

### Webfigure 1: Effects of allocation to molnupiravir on hospital discharge by baseline characteristics

| | Molnupiravir | Usual care | Hazard ratio (95% CI) |
|---|---|---|---|
| **Age, years ($\chi^2_1$=0.6; p=0.42)** | | | |
| <70 | 137/168 (81.5%) | 159/194 (82.0%) | 0.99 (0.79−1.25) |
| ≥70 <80 | 102/140 (72.9%) | 104/142 (73.2%) | 1.03 (0.78−1.35) |
| ≥80 | 80/137 (58.4%) | 91/142 (64.1%) | 0.83 (0.62−1.13) |
| **Sex ($\chi^2_1$=0.2; p=0.65)** | | | |
| Men | 177/257 (68.9%) | 210/290 (72.4%) | 0.98 (0.81−1.20) |
| Women | 142/188 (75.5%) | 144/188 (76.6%) | 0.92 (0.73−1.16) |
| **Race ($\chi^2_1$=1.5; p=0.21)** | | | |
| White | 283/395 (71.6%) | 306/414 (73.9%) | 0.98 (0.83−1.15) |
| BAME | 23/31 (74.2%) | 40/46 (87.0%) | 0.70 (0.42−1.17) |
| **Days since symptom onset ($\chi^2_1$=0.5; p=0.47)** | | | |
| ≤7 | 105/150 (70.0%) | 113/158 (71.5%) | 1.04 (0.80−1.35) |
| >7 | 214/295 (72.5%) | 241/320 (75.3%) | 0.92 (0.77−1.11) |
| **Respiratory support at randomization ($\chi^2_1$=0.2; p=0.70)*** | | | |
| None | 54/66 (81.8%) | 81/94 (86.2%) | 0.87 (0.61−1.22) |
| Simple oxygen | 223/293 (76.1%) | 234/309 (75.7%) | 0.99 (0.83−1.19) |
| Non invasive ventilation | 42/86 (48.8%) | 39/73 (53.4%) | 0.94 (0.60−1.45) |
| Invasive mechanical ventilation | 0/0 | 0/2 (0.0%) | |
| **Use of corticosteroids ($\chi^2_1$=1.1; p=0.29)** | | | |
| Yes | 278/389 (71.5%) | 305/420 (72.6%) | 1.00 (0.85−1.18) |
| No | 41/56 (73.2%) | 47/56 (83.9%) | 0.79 (0.52−1.20) |
| **Antigen status ($\chi^2_1$=0.0; p=0.82)** | | | |
| Positive | 144/212 (67.9%) | 155/226 (68.6%) | 0.97 (0.78−1.22) |
| Negative | 150/202 (74.3%) | 170/212 (80.2%) | 0.94 (0.75−1.17) |
| **Anti−N status ($\chi^2_1$=0.0; p=0.85)** | | | |
| Positive | 80/114 (70.2%) | 87/113 (77.0%) | 0.93 (0.69−1.26) |
| Negative | 215/301 (71.4%) | 238/325 (73.2%) | 0.96 (0.80−1.16) |
| **Anti−S status ($\chi^2_1$=0.7; p=0.41)** | | | |
| Positive | 237/334 (71.0%) | 278/371 (74.9%) | 0.94 (0.79−1.11) |
| Negative | 57/80 (71.2%) | 47/67 (70.1%) | 1.12 (0.76−1.65) |
| **Use of antiviral treatments ($\chi^2_1$=0.6; p=0.44)** | | | |
| Yes | 218/306 (71.2%) | 232/323 (71.8%) | 1.00 (0.83−1.20) |
| No | 101/139 (72.7%) | 122/155 (78.7%) | 0.88 (0.68−1.15) |
| **All participants** | **319/445 (71.7%)** | **354/478 (74.1%)** | **0.96 (0.82−1.12) p=0.60** |

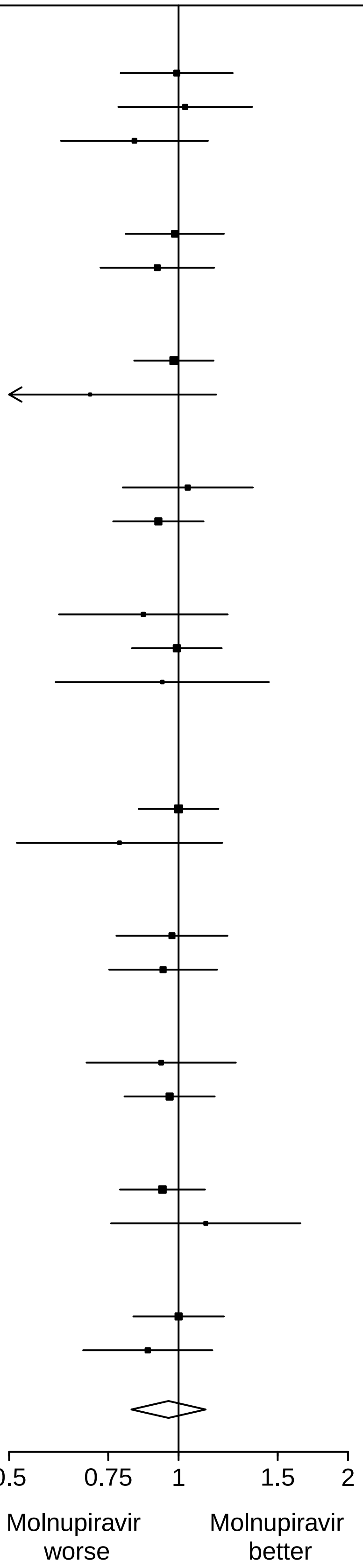

*Trend test does not include invasive mechanical ventilation due to insufficient numbers of patients.

### Webfigure 2: Effects of allocation to molnupiravir on invasive mechanical ventilation or death in those not on invasive mechanical ventilation at randomisation, by baseline characteristics

| | Molnupiravir | Usual care | RR (95% CI) |
|---|---|---|---|
| **Age, years ($\chi^2_1$=1.0; p=0.32)** | | | |
| <70 | 20/168 (11.9%) | 21/193 (10.9%) | 0.96 (0.55−1.69) |
| ≥70 <80 | 18/140 (12.9%) | 27/142 (19.0%) | 0.62 (0.36−1.06) |
| ≥80 | 39/137 (28.5%) | 33/141 (23.4%) | 1.19 (0.82−1.72) |
| **Sex ($\chi^2_1$=0.4; p=0.50)** | | | |
| Men | 46/257 (17.9%) | 54/288 (18.8%) | 0.90 (0.64−1.25) |
| Women | 31/188 (16.5%) | 27/188 (14.4%) | 1.08 (0.69−1.71) |
| **Race ($\chi^2_1$=0.7; p=0.42)** | | | |
| White | 73/395 (18.5%) | 73/413 (17.7%) | 1.01 (0.77−1.33) |
| BAME | 3/31 (9.7%) | 5/46 (10.9%) | 0.58 (0.15−2.18) |
| **Days since symptom onset ($\chi^2_1$=1.3; p=0.26)** | | | |
| ≤7 | 31/150 (20.7%) | 30/158 (19.0%) | 1.17 (0.77−1.76) |
| >7 | 46/295 (15.6%) | 51/318 (16.0%) | 0.85 (0.60−1.21) |
| **Respiratory support at randomization ($\chi^2_1$=0.4; p=0.55)*** | | | |
| None | 5/66 (7.6%) | 5/94 (5.3%) | 1.38 (0.42−4.56) |
| Simple oxygen | 45/293 (15.4%) | 48/309 (15.5%) | 0.98 (0.68−1.41) |
| Non invasive ventilation | 27/86 (31.4%) | 28/73 (38.4%) | 0.89 (0.60−1.34) |
| **Use of corticosteroids ($\chi^2_1$=0.1; p=0.78)** | | | |
| Yes | 71/389 (18.3%) | 76/418 (18.2%) | 0.95 (0.73−1.26) |
| No | 6/56 (10.7%) | 4/56 (7.1%) | 1.13 (0.34−3.74) |
| **Antigen status ($\chi^2_1$=0.4; p=0.53)** | | | |
| Positive | 40/212 (18.9%) | 48/225 (21.3%) | 0.92 (0.65−1.30) |
| Negative | 32/202 (15.8%) | 26/212 (12.3%) | 1.11 (0.70−1.76) |
| **Anti−N status ($\chi^2_1$=1.8; p=0.18)** | | | |
| Positive | 24/114 (21.1%) | 14/112 (12.5%) | 1.37 (0.77−2.46) |
| Negative | 48/301 (15.9%) | 60/325 (18.5%) | 0.87 (0.63−1.20) |
| **Anti−S status ($\chi^2_1$=1.4; p=0.24)** | | | |
| Positive | 59/334 (17.7%) | 60/370 (16.2%) | 1.04 (0.76−1.41) |
| Negative | 13/80 (16.2%) | 14/67 (20.9%) | 0.69 (0.38−1.27) |
| **Use of antiviral treatments ($\chi^2_1$=0.0; p=0.94)** | | | |
| Yes | 53/306 (17.3%) | 56/321 (17.4%) | 0.96 (0.70−1.33) |
| No | 24/139 (17.3%) | 25/155 (16.1%) | 0.94 (0.59−1.52) |
| **All participants** | **77/445 (17.3%)** | **81/476 (17.0%)** | **0.96 (0.73−1.25)** **p=0.75** |

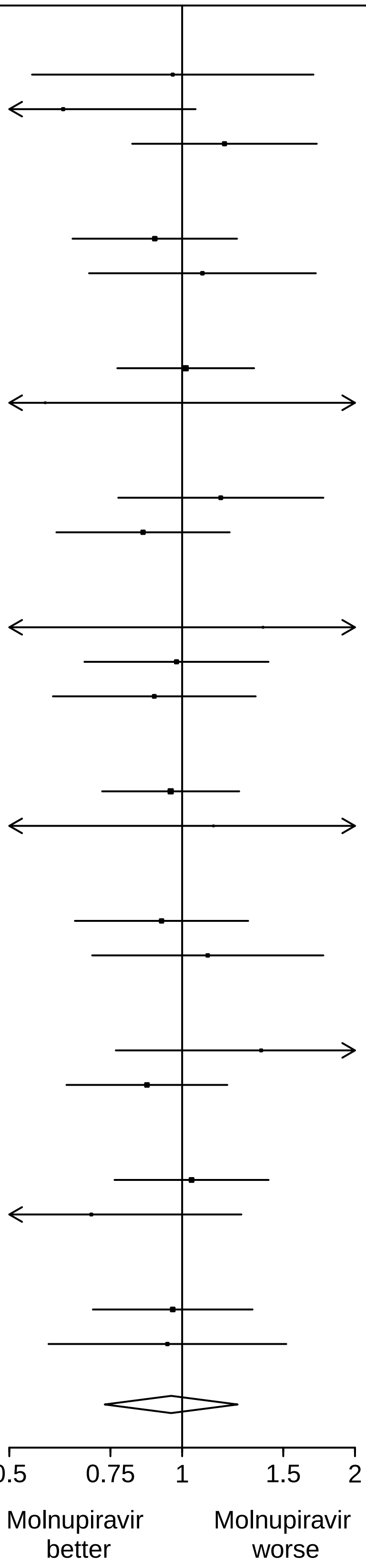

### Appendices

#### Appendix 1: RECOVERY Trial Protocol V25.0

RECOVERY
Randomised Evaluation of COVID-19 Therapy

### RANDOMISED EVALUATION OF COVID-19 THERAPY (RECOVERY)

**Background:** In early 2020, as this protocol was being developed, there were no approved treatments for COVID-19, a disease induced by the novel coronavirus SARS-CoV-2 that emerged in China in late 2019. The UK New and Emerging Respiratory Virus Threats Advisory Group (NERVTAG) advised that several possible treatments should be evaluated, including Lopinavir-Ritonavir, low-dose corticosteroids, and Hydroxychloroquine (which has now been done). A World Health Organization (WHO) expert group issued broadly similar advice. These groups also advised that other treatments would soon emerge that require evaluation. In addition, due to lack of community transmission due to COVID-19 control measures, a more severe influenza season is expected when these ease.

**Eligibility and randomisation:** This protocol describes a randomised trial among patients hospitalised for COVID-19 and/or influenza. (Treatments for influenza are only being assessed in the UK.) Eligible patients are randomly allocated between one or more treatment arms, each to be given in addition to the usual standard of care in the participating hospital. The study is dynamic, and treatments are added and removed as results and suitable treatments become available. The randomised treatment comparisons in this version of the protocol (which should be checked and confirmed as the current version) are shown in Table 1. For patients for whom not all the trial arms are appropriate or at locations where not all are available, randomisation will be between fewer arms..

| Condition | Randomised comparisons, each vs. usual care alone | UK | Other countries |
|---|---|---|---|
| COVID-19 | High-dose corticosteroids | ✓ (age ≥18 years with hypoxia)[a] | ✓ (age ≥18 years with hypoxia)[a] |
| | Empagliflozin | ✓ (age ≥18 years) | ✓ (age ≥18 years) |
| | Sotrovimab | ✓ (age ≥12 years) | × |
| | Molnupiravir | ✓ (age ≥18 years) | ✓ (age ≥18 years) |
| | Paxlovid | ✓ (age ≥18 years) | × |
| Influenza | Baloxavir | ✓ (age ≥12 years) | × |
| | Oseltamivir | ✓ (any age) | × |
| | Low-dose corticosteroids | ✓ (any age with hypoxia)[b] | × |

[a] without suspected or confirmed influenza infection; [b] without suspected or confirmed SARS-CoV-2 infection. Information on completed arms is available in Section 7.

**Table 1: Current comparisons**

In a partial factorial design, participants may be entered into one or more randomised comparisons of active treatment plus usual care vs. usual care alone, simultaneously. This allows the effects of one treatment to be assessed in the presence or absence of another which generates useful information for clinicians and health policy-makers. In particular, this allows antiviral therapies to be assessed as monotherapy and in combination, which will provide important information on the efficacy, safety and the development of resistance. This protocol indicates clearly where specific combinations are not desirable.

**Adaptive design:** The interim trial results will be monitored by an independent Data Monitoring Committee (DMC). The most important task for the DMC will be to assess whether the randomised comparisons in the study have provided evidence on mortality that is strong enough (with a range of uncertainty around the results that is narrow enough) to affect national and global treatment strategies. In such a circumstance, the DMC will inform the Trial Steering Committee who will make the results available to the public and amend the trial arms accordingly. Regardless, follow-up will continue for all randomised participants, including those previously assigned to trial arms that are modified or ceased. New trial arms can be added as evidence emerges that other candidate therapeutics should be evaluated.

**Outcomes:** The main outcomes will be death, discharge, need for ventilation and need for renal replacement therapy. For the main analyses, follow-up will be censored at 28 days after randomisation. Additional information on longer term outcomes may be collected through review of medical records or linkage to medical databases where available (such as those managed by NHS Digital and equivalent organisations in the devolved nations).

**Simplicity of procedures:** To facilitate collaboration, even in hospitals that suddenly become overloaded, patient enrolment (via the internet) and all other trial procedures are greatly streamlined. Informed consent is simple and data entry is minimal. Randomisation via the internet is simple and quick, at the end of which the allocated treatment is displayed on the screen and can be printed or downloaded. Key follow-up information is recorded at a single timepoint and may be ascertained by contacting participants in person, by phone or electronically, or by review of medical records and databases.

**Data to be recorded:** At randomisation, information will be collected on the identity of the randomising clinician and of the patient, age, sex, major co-morbidity, pregnancy, illness onset date and severity, and any contraindications to the study treatments. The main outcomes will be death (with date and probable cause), discharge (with date), need for ventilation (with number of days recorded) and need for renal replacement therapy. Reminders will be sent if outcome data have not been recorded by 28 days after randomisation. Suspected Unexpected Serious Adverse Reactions (SUSARs) to one of the study medications (e.g., Stevens-Johnson syndrome, anaphylaxis, aplastic anaemia) will be collected and reported in an expedited fashion. Other adverse events will not be recorded but may be available through linkage to medical databases.

**Numbers to be randomised:** The larger the number randomised the more accurate the results will be, but the numbers that can be randomised will depend critically on how large the epidemic becomes. If substantial numbers are hospitalised in the participating centres then it may be possible to randomise several thousand with mild disease and a few thousand with severe disease, but realistic, appropriate sample sizes could not be estimated at the start of the trial.

**Heterogeneity between populations:** If sufficient numbers are studied, it may be possible to generate reliable evidence in certain patient groups (e.g. those with major co-morbidity or who are older). To this end, data from this study may be combined with data from other trials of treatments for COVID-19 or influenza.

**Add-on studies:** Particular countries or groups of hospitals, may well want to collaborate in adding further measurements or observations, such as serial blood gases or chemistry, serial lung imaging, or serial documentation of other aspects of disease status. While well-organised additional research studies of the natural history of the disease or of the effects of the trial treatments could well be valuable (although the lack of placebo control may bias the assessment of subjective side-effects, such as gastro-intestinal problems), they are not core requirements.

**To enquire about the trial, contact the RECOVERY Central Coordinating Office**
Nuffield Department of Population Health, Richard Doll Building, Old Road Campus, Roosevelt Drive, Oxford OX3 7LF, United Kingdom
**To enquire about the trial outside of the UK, contact the relevant Clinical Trial Units**
**To RANDOMISE a patient, visit: www.recoverytrial.net**

### 1 BACKGROUND AND RATIONALE

#### 1.1 Setting

In 2019 a novel coronavirus-disease (COVID-19) emerged in Wuhan, China. A month later the Chinese Center for Disease Control and Prevention identified a new beta-coronavirus (SARS coronavirus 2, or SARS-CoV-2) as the aetiological agent.[1] The clinical manifestations of COVID-19 range from asymptomatic infection or mild, transient symptoms to severe viral pneumonia with respiratory failure. As many patients do not progress to severe disease the overall case fatality rate per infected individual is low, but hospitals in areas with significant community transmission have experienced a major increase in the number of hospitalised pneumonia patients, and the frequency of severe disease in hospitalised patients can be as high as 30%.[2-4] The progression from prodrome (usually fever, fatigue and cough) to severe pneumonia requiring oxygen support or mechanical ventilation often takes one to two weeks after the onset of symptoms.[2] The kinetics of viral replication in the respiratory tract are not well characterized, but this relatively slow progression provides a potential time window in which antiviral therapies could influence the course of disease. In May 2020 a new COVID-associated inflammatory syndrome in children was identified, Paediatric Inflammatory Multisystem Syndrome - Temporally associated with SARS-CoV-2 (PIMS-TS).[5] A rapid NHS England-led consensus process identified the need to evaluate corticosteroids and intravenous immunoglobulin (IVIg) as initial therapies in PIMS-TS, and confirmed tocilizumab as one of the biological anti-inflammatory agents to be evaluated as a second line therapy.

The COVID-19 control measures in place in the UK during the winter of 2020/21 resulted in an almost complete absence of influenza transmission over that period. This extended period without exposure to influenza viruses is unique and may have resulted in antibody waning and increased population susceptibility. Therefore, there is a possibility of a large resurgence of influenza in the winter of 2021/22. The treatment of influenza in hospitalised patients has progressed little in the last 20 years and there is substantial uncertainty and disagreement about optimal treatment of this patient group.

#### 1.2 Treatment Options

The protocol allows reliable assessment of the effects of multiple different treatments (including re-purposed and novel drugs) on major outcomes in COVID-19, influenza and PIMS-TS. All patients will receive usual care for the participating hospital. The current treatments under evaluation are summarised in Table 1 above with further details provided in sections 2.4-2.6 and in Appendices 1-4 (sections 8.1-8.4).

#### 1.3 Modifications to the number of treatment arms

Other arms can be added if evidence emerges that there are suitable candidate therapeutics. Conversely, in some patient populations, not all trial arms are appropriate (e.g. due to contraindications based on co-morbid conditions or concomitant medication); in some hospitals or countries, not all treatment arms will be available (e.g. due to manufacturing and supply issues); and at some times, not all treatment arms will be active (e.g. due to lack of relevant approvals and contractual agreements). The Trial Steering Committee may elect to pause one or more of the arms in order to increase trial efficiency during a fluctuating epidemic. In any of these situations, randomisation will be between fewer arms. Depending on the availability and suitability of treatments, it may be allowed for participants to be randomised in only one or two parts of the main randomisations.

#### 1.4 Design Considerations

The RECOVERY Protocol describes an overarching trial design to provide reliable evidence on the efficacy of candidate therapies for confirmed COVID-19 and/or influenza infection in hospitalised patients receiving usual standard of care. (Treatments for influenza are only being assessed in the UK.)

In early 2020, when the trial first started, there were no known treatments for COVID-19. The anticipated scale of the epidemic is such that hospitals, and particularly intensive care facilities, may be massively overstretched at some points in time, with around 10% requiring hospitalisation. Similarly, the winter of 2021-22 may pose a similar challenge to hospitals when ongoing COVID-19 cases coincide with a significant number of influenza cases. In such situations, even treatments with only a moderate impact on survival or on hospital resources could be worthwhile. Therefore, the focus of RECOVERY is the impact of candidate treatments on mortality and on the need for hospitalisation or ventilation.

Critically, the trial is designed to minimise the burden on front-line hospital staff working within an overstretched care system during a major epidemic. Eligibility criteria are therefore simple and trial processes (including paperwork) are minimised.

The protocol is deliberately flexible so that it is suitable for a wide range of settings, allowing:

- a broad range of patients to be enrolled in large numbers;
- randomisation between only those treatment arms that are *both* available at the hospital *and* not believed by the enrolling doctor to be contraindicated (e.g. by particular co-morbid conditions or concomitant medications);
- treatment arms to be added or removed according to the emerging evidence; and
- additional substudies may be added to provide more detailed information on side effects or sub-categorisation of patient types but these are not the primary objective and are not required for participation.

In a cohort of 191 hospitalised COVID-19 patients with a completed outcome, the median time from illness onset to discharge was 22.0 days (IQR 18.0-25.0) and the median time to death was 18.5 days (15.0-22.0). Thirty-two patients (17%) required invasive mechanical ventilation and the median time from onset to mechanical ventilation was 14.5 days. Therefore, early endpoint assessment, such as 28 days after randomisation, is likely to provide largely complete outcome data and will permit early assessment of treatment efficacy and safety.[6] For influenza, the average length of hospital stay in the UK is around 9 days, so assessment at 28 days will capture most outcomes.[7]

#### 1.5 Potential for effective treatments to become available

In early 2020, when the trial first started, there were no known treatments for COVID-19. However, over time, effective treatments may become available, typically as the result of reliable information from randomised trials (including from this study). For example, in June 2020, results from the RECOVERY trial showed that dexamethasone 6mg once daily reduces the mortality in COVID-19 patients requiring mechanical ventilation or oxygen. In response, many clinical guidelines now recommend the use of dexamethasone 6mg once daily as standard of care for these types of patients.

The RECOVERY trial randomises eligible participants to usual standard of care for the local hospital alone vs usual standard of care plus one or more additional study treatments. Over time, it is expected that usual standard of care alone will evolve. Thus randomisation will always be relevant to the current clinical situation and the incremental effects of the study treatments will be appropriately assessed.

##### 1.6 Early phase assessments

In the UK, the COVID-19 Therapeutics Advisory Panel (CTAP[a]) may propose that RECOVERY assesses interventions for which additional information is required before they are considered for large-scale assessment of the impact on mortality. Such assessments will be tailored to the uncertainty specific to the intervention and typically be conducted at a subset of sites among a smaller group of participants before the results are reviewed and a decision made whether to include them in the main trial.

#### 2 DESIGN AND PROCEDURES

##### 2.1 Eligibility

Patients are eligible for the study if all of the following are true:

**(i) Hospitalised**

**(ii) a) Viral pneumonia syndrome**

In general, viral pneumonia should be suspected when a patient presents with:

a) typical symptoms (e.g. influenza-like illness with fever and muscle pain, or respiratory illness with cough and shortness of breath); and
b) compatible chest X-ray findings (consolidation or ground-glass shadowing); and
c) alternative causes have been considered unlikely or excluded (e.g. heart failure, bacterial pneumonia).

However, the diagnosis remains a clinical one based on the opinion of the managing doctor.

**(iii) Confirmed SARS-CoV-2 infection (all countries) and/or influenza A or B infection (UK only)**

**(iv) No medical history that might, in the opinion of the attending clinician, put the patient at significant risk if he/she were to participate in the trial**

[a] https://www.gov.uk/government/publications/covid-19-treatments-making-a-proposal-for-clinical-trials/guidance-making-a-proposal-for-covid-19-therapeutics-clinical-trials#uk-covid-19-therapeutics-advisory-panel-uk-ctap

Patients in the UK with SARS-CoV-2 and influenza co-infection are eligible, but would be excluded from certain comparisons (as described in the table on page 1). In addition, if the attending clinician believes that there is a specific contra-indication to one of the active drug treatment arms (see Appendix 2, Appendix 3 [for children], and Appendix 4 for pregnant and breastfeeding women) or that the patient should definitely be receiving one of the active drug treatment arms then that arm will not be available for randomisation for that patient. For patients who lack capacity, an advanced directive or behaviour that clearly indicates that they would not wish to participate in the trial would be considered sufficient reason to exclude them from the trial.

Patients who have been previously recruited into RECOVERY are eligible to be recruited again as long as their previous randomisation was >6 months ago. Patients will not be recruited into the same randomised comparison (e.g. sotrovimab vs. usual care) on more than one occasion, regardless of how far apart they occur.

In some locations, children (aged <18 years) will not be recruited, to comply with local and national regulatory approvals (see Table 1 and Sections 2.4-2.6 and 8.3).

#### 2.2 Consent

Informed consent should be obtained from each patient 16 years and over before enrolment into the study. Due to the poor outcomes in COVID-19 patients who require ventilation (>90% mortality in one cohort[6]), patients who lack capacity to consent due to severe disease (e.g. needs ventilation), and for whom a relative to act as the legally designated representative is not available (in person), randomisation and consequent treatment will proceed with consent provided by a clinician (independent of the trial[b]) who will act as the legally designated representative (if allowed by local regulations).

If they regain capacity, such participants should be provided with information about the trial (ideally prior to discharge, but otherwise as soon as possible thereafter), what their rights are and how to exercise them, but it is not necessary to obtain their written consent[c]. Provision of such information (i.e. the current participant information sheet) will be documented in the medical record.

For children aged <16 years old consent will be sought from their parents or legal guardian. Where possible, children aged between 10-15 years old will also be asked for assent. Children aged ≥16 years old will asked for consent as for adults. Witnessed[d] consent may be obtained over the telephone or web video link if hospital visiting rules or parental infection mean a parent/guardian cannot be physically present.

Information about participants' involvement will be included in routine clinical communications (e.g. discharge summaries) provided to participants (and, in the UK their GPs) . If any other relevant information arises during the trial, this may also be sent to GPs.

---

[b] Independent clinicians may complete study training, but have no other involvement in the trial, e.g. eligibility assessment, or randomisation

[c] Unless required by local regulations. (This is not required in the UK.)

[d] The witness should be impartial i.e. not a member of the research team, but they do not require specific training or knowledge of the trial.

#### 2.3 Baseline information

The following information will be recorded on the web-based form by the attending clinician or delegate:

- Patient details (e.g. name or initials [depending on privacy requirements], NHS/CHI number [UK only] or medical records number, date of birth, sex)
- Clinician details (e.g. name)
- Symptom onset date
- Disease severity as assessed by need for supplemental oxygen, non-invasive ventilation or invasive mechanical ventilation/extracorporeal membrane oxygenation (ECMO)
- Oxygen saturations on air (if available)
- Latest routine measurement of creatinine, C-reactive protein, and D-dimer (if available)
- SARS-CoV-2 test result (and/or influenza test result in UK)
- Major co-morbidity (e.g. heart disease, diabetes, chronic lung disease) and pregnancy (including pregnancy test result in all women of child-bearing potential[e])
- Use of relevant medications (e.g. corticosteroids, anti-virals) and prior vaccination
- Date of hospitalisation
- Contraindication to the study treatment regimens (in the opinion of the attending clinician)
- Name of person completing the form

The person completing the form will then be asked to confirm that they wish to randomise the patient and will then be required to enter their name and e-mail address.

##### 2.3.1 Baseline sample collection (UK only)[f]

###### 2.3.1.1 Participants with COVID-19

Participants with COVID-19 entering sotrovimab, molnupiravir or Paxlovid comparisons should have a serum sample collected **after obtaining consent and prior to randomisation** in which presence of SARS-CoV-2 antigen and antibodies against it may be tested. In addition, a nasal swab should be collected in which the level of SARS-CoV-2 viral RNA (and genotyping for resistance markers) will be measured.

###### 2.3.1.2 Participants with influenza pneumonia

Participants with influenza pneumonia should have a nasal swab collected in which the presence of influenza virus will be measured.

[e] A woman of childbearing potential is defined as a post-menarchal pre-menopausal female capable of becoming pregnant. This includes women on oral, injectable, or mechanical contraception; women who are single; women whose male partners have been vasectomized or whose male partners have received or are utilizing mechanical contraceptive devices. The potential inclusion of any pregnant women should be discussed with a consultant obstetrician (or obstetric physician).
[f] Collection of these samples will continue until the Steering Committee determine (on the basis of data blinded to treatment allocation) that sufficient information is available to assess the effect of treatment on viral load and/or resistance markers.

#### 2.4 Randomised allocation of treatment for COVID-19

In addition to receiving usual care, eligible patients with confirmed SARS-CoV-2 infection will be allocated treatment(s) using a central web-based randomisation service (without stratification or minimisation). A factorial design is used such that eligible patients may be randomised simultaneously to one or more of the study treatment arms (depending on location and infection). The doses in this section are for adults (see Appendix 3 for paediatric dosing).

##### 2.4.1 Randomisation part E

Eligible patients (adult patients ≥18 years old without suspected or confirmed influenza co-infection) and with clinical evidence of hypoxia (i.e. receiving oxygen or with oxygen saturations <92% on room air) may be randomised in a ratio of 1:1 to one of the arms listed below.

- **No additional treatment**[g]

- High-dose corticosteroids: **dexamethasone 20 mg (base) once daily** by mouth, nasogastric tube or intravenous infusion for 5 days follow by **dexamethasone 10 mg (base) once daily** by mouth, nasogastric tube or intravenous infusion for 5 days. [h,i]

##### 2.4.2 Randomisation part F:

Eligible patients (adult patients ≥18 years old) may be randomised in a 1:1 ratio to one of the arms listed below.

- **No additional treatment**

- **Empagliflozin 10 mg once daily** by mouth for 28 days (or until discharge, if earlier). Participants with diabetes allocated empagliflozin should have daily ketone checks while taking the treatment (see Appendix 2 for further details).

##### 2.4.3 Randomisation part J (UK only):

Eligible patients (patients ≥12 years old) may be randomised in a 1:1 ratio to one of the arms listed below.

- **No additional treatment**

- **Sotrovimab 1000 mg in 100 mL 0.9% sodium chloride or 5% dextrose** by intravenous infusion over 1 hour as soon as possible after randomisation.

---

[g] Usual care in hypoxic patients is expected to include low dose (6mg daily) dexamethasone

[h] Treatment should be discontinued at 10 days or on discharge from hospital if sooner. Participants can be given a short 'weaning' course when they complete their study allocation if considered clinically necessary.

[i] Pregnant women should receive either prednisolone (130 mg) orally or hydrocortisone (540 mg in divided doses) intravenously or methylprednisolone (100 mg) intravenously for five days, followed by either prednisolone (65 mg) orally or hydrocortisone (270 mg in divided doses) intravenously or methylprednisolone (50 mg) intravenously for five days.

###### 2.4.4 Randomisation part K:

Eligible patients (patients ≥18 years old) may be randomised in a 1:1 ratio to one of the arms listed below.

- **No additional treatment**

- **Molnupiravir 800 mg twice daily** for 5 days by mouth[j].

###### 2.4.5 Randomisation part L (UK only):

Eligible patients (patients ≥18 years old) may be randomised in a 1:1 ratio to one of the arms listed below.

- **No additional treatment**

- **Paxlovid (nirmatrelvir/ritonavir) 300/100 mg twice daily** for 5 days by mouth[j].

##### 2.5 Randomised allocation of treatment for influenza (UK only)

In addition to receiving usual care, eligible patients with confirmed influenza A or B infection will be allocated treatment(s) using a central web-based randomisation service (without stratification or minimisation). A factorial design is used such that eligible patients may be randomised simultaneously to one or more of the study treatment arms (depending on location and infection). The doses in this section are for adults (see Appendix 3 for paediatric dosing). Study treatments do not need to be continued after discharge from hospital unless otherwise specified.

###### 2.5.1 Randomisation part G: (UK only)

Eligible patients (≥12 years old with or without SARS-CoV-2 co-infection) may be randomised in a ratio of 1:1 to one of the arms listed below.

- **No additional treatment**

- **Baloxavir marboxil 40mg (or 80mg if weight ≥80kg) once daily** by mouth or nasogastic tube to be given on day 1 and day 4[j].

###### 2.5.2 Randomisation part H: (UK only)

Eligible patients (any age, with or without SARS-CoV-2 co-infection) may be randomised in a ratio of 1:1 to one of the arms listed below.

- **No additional treatment**

- **Oseltamivir 75mg twice daily** by mouth or nasogastric tube for five days[j,k].

---

[j] If participant is discharged before course is complete, the participant should be provided with medication to complete the course at home.

[k] Course can be extended to 10 days for immunosuppressed patients at the managing clinician's discretion.

###### 2.5.3 Randomisation part I: (UK only)

Eligible patients (any age without suspected or confirmed SARS-CoV-2 infection) and with clinical evidence of hypoxia (i.e. receiving oxygen or with oxygen saturations <92% on room air) may be randomised in a ratio of 1:1 to one of the arms listed below.

- **No additional treatment**

- Low-dose corticosteroids: **Dexamethasone 6mg once daily given** orally or intravenously for ten days or until discharge (whichever happens earliest)[l]

##### 2.6 Administration of allocated treatment

The details of the allocated study treatments will be displayed on the screen and can be printed or downloaded. The hospital clinicians are responsible for prescription and administration of the allocated treatments. The patient's own doctors are free to modify or stop study treatments if they feel it is in the best interests of the patient without the need for the patient to withdraw from the study (see section 2.9). This study is being conducted within hospitals. Therefore use of medication will be subject to standard medication reviews (typically within 48 hours of enrolment) which will guide modifications to both the study treatment and use of concomitant medication (e.g. in the case of potential drug interactions).

##### 2.7 Collecting follow-up information

The following information will be ascertained at the time of death or discharge or at 28 days after first randomisation (whichever is sooner):

- Vital status (alive / dead, with date and presumed cause of death, if appropriate)
- Hospitalisation status (inpatient / discharged, with date of discharge, if appropriate)
- Use of ventilation (with days of use and type, if appropriate)
- Use of renal dialysis or haemofiltration
- Documented new major cardiac arrhythmia (including atrial and ventricular arrhythmias)
- Major bleeding (defined as intracranial bleeding or bleeding requiring transfusion, endoscopy, surgery, or vasoactive drugs)
- Thrombotic event, defined as either (i) acute pulmonary embolism; (ii) deep vein thrombosis; (iii) ischaemic stroke; (iv) myocardial infarction; or (v) systemic arterial embolism.
- Non-coronavirus / non-influenza infection, categorised by site and putative organism (virus, bacteria, fungus, other)
- Use of any medications included in the RECOVERY trial protocol (including drugs in the same class) or other purported COVID-19 and influenza treatments (e.g. remdesivir, neuraminidase inhibitors)
- Participation in other randomised trials of interventions (vaccines or treatments) for COVID-19 or influenza.
- Metabolic complications: Ketoacidosis; hyperglycaemic hyperosmolar state; hyperglycaemia requiring new use of insulin; severe hypoglycaemia (defined as hypoglycaemia causing reduced conscious level requiring another person to help recover)
- Seizures

---

[l] In pregnancy or breastfeeding women, prednisolone 40 mg administered by mouth (or intravenous hydrocortisone 80 mg twice daily) should be used instead of dexamethasone. For dosing in children see Appendix 3.

- Laboratory results: highest creatinine, alanine (or aspartate) transamine and bilirubin recorded during admission
- Infusion reactions to Sotrovimab
- For pregnant women in UK, ID number in UK Obstetric Surveillance System

Follow-up information is to be collected on all study participants, irrespective of whether or not they complete the scheduled course of allocated study treatment. Study staff will seek follow-up information through various means including medical staff, reviewing information from medical notes, routine healthcare systems, and registries.

For all randomised participants, vital status (alive / dead, with date and presumed cause of death, if appropriate) is to be ascertained at 28 days after first randomisation. This may be achieved through linkage to routine death registration data (e.g. in the UK) or through direct contact with the participant, their relatives, or medical staff and completion of an additional follow-up form. Where available, data from routine healthcare records (including linkage to medical databases held by organisations such as NHS Digital in the UK) will be used to supplement data collected by trial sites. Further details are described in the Definition and Derivation of Baseline Characteristics and Outcomes standard operating procedure.[m]

##### 2.7.1 Follow-up swab samples (UK only)[n]

###### 2.7.1.1 Participants with COVID-19

Participants with COVID-19 in sotrovimab, molnupiravir or Paxlovid comparisons should have a nasal swab collected on days 3 and 5 in which the level of SARS-CoV-2 viral RNA (and genotyping for resistance makers) will be measured.

###### 2.7.1.2 Participants with influenza pneumonia

Participants with influenza pneumonia should have a nasal swab collected on day 5 in which the presence of influenza virus (and genotyping for baloxavir or oseltamivir resistance markers) will be measured.

#### 2.8 Duration of follow-up

All randomised participants are to be followed up until death, discharge from hospital or 28 days after randomisation (whichever is sooner). It is recognised that in the setting of this trial, there may be some variability in exactly how many days after randomisation, information on disease status is collected. This is acceptable and will be taken account of in the analyses and interpretation of results, the principle being that some information about post-randomisation disease status is better than none.

In the UK, longer term (up to 10 years) follow-up will be sought through linkage to electronic healthcare records and medical databases including those held by NHS Digital, Public Health England and equivalent bodies, and to relevant research databases (e.g. UK Biobank, Genomics England). Outside the UK, due to the absence of electronic health data

---

[m] Available at https://www.recoverytrial.net/files/recovery-outcomes-definitions-v3-0.pdf

[n] Collection of these samples will continue until the Steering Committee determine (on the basis of data blinded to treatment allocation) that sufficient information is available to assess the effect of treatment on viral load and/or resistance markers. Participants discharged before day 5 will be asked to take this sample at home and will be provided with instructions and materials to do so.

linkage, additional follow-up will be conducted at 6 months after first randomisation by telephone or in person (at a clinic) in order to collect information on mortality (including date and cause) and re-admission to hospital (including date[s] and primary reason[s]). This information will be captured on a web-based case report form.

##### 2.9 Withdrawal of consent

A decision by a participant (or their parent/guardian) that they no longer wish to continue receiving study treatment should **not** be considered to be a withdrawal of consent for follow-up. However, participants (or their parent/guardian) are free to withdraw consent for some or all aspects of the study at any time if they wish to do so. In accordance with regulatory guidance, de-identified data that have already been collected and incorporated in the study database will continue to be used (and any identifiable data will be destroyed).

For participants who lack capacity, if their legal representative withdraws consent for treatment or methods of follow-up then these activities would cease. If such participants regain capacity and no longer wish to participate then they can withdraw the consent given on their behalf as above.

#### 3 STATISTICAL ANALYSIS

All analyses for reports, presentations and publications will be prepared by the coordinating centre at the Nuffield Department of Population Health, University of Oxford. A more detailed statistical analysis plan will be developed by the investigators and published on the study website whilst still blind to any analyses of aggregated data on study outcomes by treatment allocation.

##### 3.1 Outcomes

###### 3.1.1 Primary and secondary outcomes for evaluation of potential treatments for COVID-19

For each pairwise comparison with the ‘no additional treatment’ arm, the **primary objective** is to provide reliable estimates of the effect of study treatments on all-cause mortality at 28 days after randomisation (with subsidiary analyses of cause of death and of death at various timepoints following discharge).

The **secondary objectives** are to assess the effects of study treatments on (a) duration of hospital stay (time to discharge alive within the first 28 days); and, (b) among patients not on invasive mechanical ventilation at baseline, the composite endpoint of death or need for invasive mechanical ventilation or ECMO.

###### 3.1.2 Primary and secondary outcomes for evaluation of potential treatments for influenza

For each pairwise comparison with the ‘no additional treatment’ arm, the **co-primary objectives** are to provide reliable estimates of the effect of study treatments on (a) all-cause mortality at 28 days after randomisation (with subsidiary analyses of cause of death and of death at various timepoints following discharge) and (b) time to discharge alive from hospital.

Holm's procedure will be used to control the family-wise error rate across these two co-primary outcomes at 5%.[8]

The **secondary objective** is to assess the effects of study treatments on the composite endpoint of death or need for invasive mechanical ventilation or ECMO among patients not on invasive mechanical ventilation at baseline.

##### 3.1.3 Other outcomes for evaluation of all treatments

Other objectives include the assessment of the effects of study treatments on the need for any ventilation (and duration of invasive mechanical ventilation), acute kidney injury and renal replacement therapy, and thrombotic events. Safety outcomes include bleeding, new major cardiac arrhythmias, metabolic complications (ketoacidosis, hyperglycaemic hyperosmolar state, hyperglycaemia requiring new use of insulin, severe hypoglycaemia). Virological outcomes include viral RNA levels in the nasopharynx and the frequency of detection of resistance markers.

Study outcomes will be assessed based on data recorded up to 28 days and up to 6 months after randomisation.

Where available, data from routine healthcare records (including linkage to medical databases held by organisations such as NHS Digital in the UK) and from relevant research studies (such as UK Biobank, Genomics England, ISARIC-4C, the UK Obstetric Surveillance System and PHOSP-COVID) will allow subsidiary analyses of the effect of the study treatments on particular non-fatal events (e.g. ascertained through linkage to Hospital Episode Statistics), the influence of pre-existing major co-morbidity (e.g. diabetes, heart disease, lung disease, hepatic insufficiency, severe depression, severe kidney impairment, immunosuppression), the maternal and infant outcomes in women pregnant at randomisation, and longer-term outcomes as well as in particular sub-categories of patient (e.g. by genotype, pregnancy).

#### 3.2 Methods of analysis

For all outcomes, comparisons will be made between all participants randomised to each treatment and its control, irrespective of whether they received their allocated treatment ("intention-to-treat" analyses).

For time-to-event analyses, each treatment group will be compared with the no additional treatment group using the log-rank test. Kaplan-Meier estimates for the time to event will also be plotted (with associated log-rank p-values). The log-rank 'observed minus expected' statistic (and its variance) will be used to estimate the average event rate ratio (and its confidence interval) for those allocated to each treatment group versus the no additional treatment group. For the primary outcome, participants discharged before 28 days will, in the absence of information to the contrary, be assumed to have survived for 28 days. For binary outcomes where the timing of the event is unknown, the risk ratio and its 95% confidence interval (and associated p-value) will be reported.

Pairwise comparisons within each randomisation will be made between each treatment arm and the no additional treatment arm (reference group) in that particular randomisation. However, since not all treatments may be available or suitable for all patients, those in the no additional treatment arm will only be included in a given comparison if, at the point of their randomisation, they *could* alternatively have been randomised to the active treatment of interest. All p-values will be 2-sided.

Pre-specified subgroup analysis (e.g., level of respiratory support, time since onset of symptoms; sex; age group; ethnicity; use of corticosteroids) will be conducted, with tests for heterogeneity (or trend) performed to assess if the effect in any particular subgroup varies materially from the overall effect. Sensitivity analyses will be conducted among those patients with laboratory confirmed SARS-CoV-2. The effect of each treatment (versus its control) will be assessed in the presence or absence of other relevant treatments the patients may receive either (a) as part of their usual care; or (b) as part of the trial (i.e., other factorial randomisations). Further details will be fully described in the Statistical Analysis Plan.

### 4 DATA AND SAFETY MONITORING

#### 4.1 Recording Suspected Serious Adverse Reactions

The focus is on those events that, based on a single case, are highly likely to be related to the study medication. Examples include anaphylaxis, Stevens-Johnson Syndrome, or bone marrow failure, where there is no other plausible explanation.

Any Serious Adverse Event[o] that is believed with a reasonable probability to be due to one of the study treatments will be considered a Suspected Serious Adverse Reaction (SSAR). In making this assessment, there should be consideration of the probability of an alternative cause (for example, COVID-19 or influenza itself or some other condition preceding randomisation), the timing of the event with respect to study treatment, the response to withdrawal of the study treatment, and (where appropriate) the response to subsequent re-challenge.

All SSARs should be reported by telephone to the Central Coordinating Office and recorded on the study IT system immediately.

#### 4.2 Central assessment and onward reporting of SUSARs

Clinicians at the Central Coordinating Office are responsible for expedited review of reports of SSARs received. Additional information (including the reason for considering it both serious and related, and relevant medical and medication history) will be sought.

The focus of Suspected Unexpected Serious Adverse Reaction (SUSAR) reporting will be on those events that, based on a single case, are highly likely to be related to the study medication. To this end, anticipated events that are either efficacy endpoints, consequences of the underlying disease, or common in the study population will be exempted from expedited reporting. Thus the following events will be exempted from expedited reporting:

---

[o] Serious Adverse Events are defined as those adverse events that result in death; are life-threatening; require in-patient hospitalisation or prolongation of existing hospitalisation; result in persistent or significant disability or incapacity; result in congenital anomaly or birth defect; or are important medical events in the opinion of the responsible investigator (that is, not life-threatening or resulting in hospitalisation, but may jeopardise the participant or require intervention to prevent one or other of the outcomes listed above).

(i) Events which are the consequence of COVID-19 or influenza; and
(ii) Common events which are the consequence of conditions preceding randomisation.

Any SSARs that are not exempt will be reviewed by a Central Coordinating Office clinician and an assessment made of whether the event is "expected" or not (assessed against the relevant Summary of Product Characteristics or Investigator Brochure). Any SSARs that are not expected would be considered a Suspected Unexpected Serious Adverse Reaction (SUSAR).

All confirmed SUSARs will be reported to the Chair of the DMC and to relevant regulatory authorities, ethics committees, and investigators in an expedited manner in accordance with regulatory requirements.

##### 4.3 Recording other Adverse Events

In addition to recording Suspected Serious Adverse Reactions (see section 4.1), information will be collected on all deaths and efforts will be made to ascertain the underlying cause. Other serious or non-serious adverse events will not be recorded unless specified in section 2.7. [p] It is anticipated that for some substudies, more detailed information on adverse events (e.g. through linkage to medical databases) or on other effects of the treatment (e.g. laboratory or radiological features) will be recorded and analysed but this is not a requirement of the core protocol.

##### 4.4 Role of the Data Monitoring Committee (DMC)

During the study, interim analyses of all study data will be supplied in strict confidence to the independent DMC. The DMC will request such analyses at a frequency relevant to the emerging data from this and other studies.

The DMC will independently evaluate these analyses and any other information considered relevant. The DMC will determine if, in their view, the randomised comparisons in the study have provided evidence on mortality that is strong enough (with a range of uncertainty around the results that is narrow enough) to affect national and global treatment strategies. In such a circumstance, the DMC will inform the Trial Steering Committee who will make the results available to the public and amend the trial arms accordingly. Unless this happens, the Trial Steering Committee, Chief Investigator, study staff, investigators, study participants, funders and other partners will remain blind to the interim results until 28 days after the last patient has been randomised for a particular intervention arm (at which point analyses may be conducted comparing that arm with the no additional treatment arm).

The DMC will review the safety and efficacy analyses among children (age <18 years) both separately and combined with the adult data.

[p] Outside the UK, additional serious adverse event information (event description, date of onset, outcome, relatedness to study treatment) will be collected if required by national regulations. This will be collected on a web-based case report form and any forms required by local regulations.

RECOVERY
Randomised Evaluation of COVID-19 Therapy

#### 4.5 Blinding

This is an open-label study. However, while the study is in progress, access to tabular results of study outcomes by allocated treatment allocation will not be available to the research team, patients, or members of the Trial Steering Committee (unless the DMC advises otherwise).

### 5 QUALITY MANAGEMENT

#### 5.1 Quality By Design Principles

In accordance with the principles of Good Clinical Practice and the recommendations and guidelines issued by regulatory agencies, the design, conduct and analysis of this trial is focussed on issues that might have a material impact on the wellbeing and safety of study participants (hospitalised patients with confirmed SARS-CoV-2 or influenza infection) and the reliability of the results that would inform the care for future patients.

The critical factors that influence the ability to deliver these quality objectives are:

- to minimise the burden on busy clinicians working in an overstretched hospital during a major epidemic
- to ensure that suitable patients have access to the trial medication without impacting or delaying other aspects of their emergency care
- to provide information on the study to patients and clinicians in a timely and readily digestible fashion but without impacting adversely on other aspects of the trial or the patient's care
- to allow individual clinicians to use their judgement about whether any of the treatment arms are not suitable for the patient
- to collect comprehensive information on the mortality and disease status

In assessing any risks to patient safety and well-being, a key principle is that of proportionality. Risks associated with participation in the trial must be considered in the context of usual care.

#### 5.2 Training and monitoring

The focus will be on those factors that are critical to quality (i.e. the safety of the participants and the reliability of the trial results). Remedial actions would focus on issues with the potential to have a substantial impact on the safety of the study participants or the reliability of the results.

The study will be conducted in accordance with the principles of International Conference on Harmonisation Guidelines for Good Clinical Research Practice (ICH-GCP) and relevant local, national and international regulations. Any serious breach of GCP in the conduct of the clinical trial will be handled in accordance with regulatory requirements. Prior to initiation of the study at each Local Clinical Centre (LCC), the Central Coordinating Office (CCO) or relevant Regional Coordinating Centre (RCC) will confirm that the LCC has adequate facilities and resources to carry out the study. LCC lead investigators and study staff will be provided with training materials.

In the context of this epidemic, visits to hospital sites are generally not appropriate as they could increase the risks of spreading infection, and in the context of this trial they generally would not influence the reliability of the trial results or the well-being of the participants. In exceptional circumstances, the CCO or RCC may arrange monitoring visits to LCCs as considered appropriate based on perceived training needs and the results of central statistical monitoring of study data.[9,10] The purpose of such visits will be to ensure that the study is being conducted in accordance with the protocol, to help LCC staff to resolve any local problems, and to provide extra training focussed on specific needs. No routine source data verification will take place.

##### 5.3 Data management

LCC clinic staff will use the bespoke study web-based applications for study management and to record participant data (including case report forms) in accordance with the protocol. Data will be held in central databases located at the CCO or on secure cloud servers. In some circumstances (e.g. where there is difficulty accessing the internet or necessary IT equipment), paper case report forms may be required with subsequent data entry by either LCC or CCO staff. Although data entry should be mindful of the desire to maintain integrity and audit trails, in the circumstances of this epidemic, the priority is on the timely entry of data that is sufficient to support reliable analysis and interpretation about treatment effects. CCO staff will be responsible for provision of the relevant web-based applications and for generation of data extracts for analyses.

All data access will be controlled by usernames and passwords, and any changes to data will require the user to enter their username and password. Staff will have access restricted to the functionality and data that are appropriate for their role in the study.

##### 5.4 Source documents and archiving

Source documents for the study constitute the records held in the study main database. These will be retained for at least 25 years from the completion of the study. Identifiable data will be retained only for so long as it is required to maintain linkage with routine data sources (see section 2.8), with the exception of children for whom such data must be stored until they reach 21 years old (due to the statute of limitations). The sponsor and regulatory agencies will have the right to conduct confidential audits of such records in the CCO and LCCs (but should mindful of the workload facing participating hospitals and the infection control requirements during this epidemic).

#### 6 OPERATIONAL AND ADMINISTRATIVE DETAILS

##### 6.1 Sponsor and coordination

The University of Oxford will act as the trial Sponsor. The trial will be coordinated by a Central Coordinating Office (CCO) within the Nuffield Department of Population Health staffed by members of the two registered clinical trials units – the Clinical Trial Service Unit and the National Perinatal Epidemiology Unit Clinical Trials Unit. The CCO will oversee Regional Coordinating Centres which will assist with selection of Local Clinical Centres

(LCCs) within their region and for the administrative support and monitoring of those LCCs. The data will be collected, analysed and published independently of the source of funding.

#### 6.2 Funding

This study is supported by grants to the University of Oxford from UK Research and Innovation/National Institute for Health Research (NIHR) and the Wellcome Trust, and by core funding provided by NIHR Oxford Biomedical Research Centre, the Wellcome Trust, the Bill and Melinda Gates Foundation, UK Foreign, Commonwealth and Development Office, Health Data Research UK, NIHR Health Protection Unit in Emerging and Zoonotic Infections and the Medical Research Council Population Health Research Unit, and NIHR Clinical Trials Unit Support Funding.

#### 6.3 Indemnity

The University has a specialist insurance policy in place which would operate in the event of any participant suffering harm as a result of their involvement in the research (Newline Underwriting Management Ltd, at Lloyd’s of London). In the UK, NHS indemnity operates in respect of the clinical treatment that is provided.

#### 6.4 Local Clinical Centres

The study will be conducted at multiple hospitals (LCCs) within each region. At each LCC, a lead investigator will be responsible for trial activities but much of the work will be carried out by medical staff attending patients with COVID-19 within the hospital and by hospital research nurses, medical students and other staff with appropriate education, training, and experience. Where LCCs plan to recruit children the principal investigator will co-opt support from a local paediatrician and/or neonatologists to oversee the management of children and infants in the trial.

#### 6.5 Supply of study treatments

For licensed treatments (e.g. corticosteroids, oseltamivir) all aspects of treatment supply, storage, and management will be in accordance with standard local policy and practice for prescription medications. Treatments issued to randomised participants will be by prescription. Such study treatments will not be labelled other than as required for routine clinical use. They will be stored alongside other routine medications with no additional monitoring. No accountability records will be kept beyond those used for routine prescriptions.

For unlicensed treatments, manufacture, packaging, labelling and delivery will be the responsibility of the pharmaceutical company and, in the UK, the Department of Health and Social Care. Each LCC will maintain an accountability log and will be responsible for the storage and issue of study treatment. If treatments require storage at a specific temperature, LCCs can use existing temperature-controlled facilities and associated monitoring. Treatment issue to randomised participants will be in accordance with local practice (and may be in line with the processes required for routine prescriptions or compassionate use).

Treatment will be issued to randomised participants by prescription.

#### 6.6 End of trial

The end of the scheduled treatment phase is defined as the date of the last follow-up visit of the last participant. In the UK, it is intended to extend follow-up for a year or more beyond the final study visit through linkage to routine medical records and central medical databases. The end of the study is the date of the final data extraction from NHS Digital (anticipated to be 10 years after the last patient is enrolled).

##### 6.7 Publications and reports

The Trial Steering Committee will be responsible for drafting the main reports from the study and for review of any other reports. In general, papers initiated by the Trial Steering Committee (including the primary manuscript) will be written in the name of the RECOVERY Collaborative Group, with individual investigators named personally at the end of the report (or, to comply with journal requirements, in web-based material posted with the report).

The Trial Steering Committee will also establish a process by which proposals for additional publications (including from independent external researchers) are considered by the Trial Steering Committee. The Trial Steering Committee will facilitate the use of the study data and approval will not be unreasonably withheld. However, the Trial Steering Committee will need to be satisfied that any proposed publication is of high quality, honours the commitments made to the study participants in the consent documentation and ethical approvals, and is compliant with relevant legal and regulatory requirements (e.g. relating to data protection and privacy). The Trial Steering Committee will have the right to review and comment on any draft manuscripts prior to publication.

##### 6.8 Substudies

Proposals for substudies must be approved by the Trial Steering Committee and by the relevant ethics committee and competent authorities (where required) as a substantial amendment or separate study before they begin. In considering such proposals, the Trial Steering Committee will need to be satisfied that the proposed substudy is worthwhile and will not compromise the main study in any way (e.g. by impairing recruitment or the ability of the participating hospitals to provide care to all patients under their care).

### 7 VERSION HISTORY

| Version number | Date | Brief Description of Changes |
|---|---|---|
| 1.0 | 13-Mar-2020 | Initial version |
| 2.0 | 21-Mar-2020 | Addition of hydroxychloroquine. Administrative changes and other clarifications. |
| 3.0 | 07-Apr-2020 | Extension of eligibility to those with suspected COVID-19<br>Addition of azithromycin arm.<br>Addition of inclusion of adults who lack permanently lack capacity.<br>Change to primary outcome from in-hospital death to death within 28 days of randomisation. |
| 4.0 | 14-Apr-2020 | Addition of second randomisation to tocilizumab vs. standard of care among patients with progressive COVID-19. |
| 5.0 | 24-Apr-2020 | Addition of children to study population. |
| 6.0 | 14-May-2020 | Addition of convalescent plasma |
| 7.0 | 18-Jun-2020 | Allowance of randomisation in part B of main randomisation without part A.<br>Removal of hydroxychloroquine and dexamethasone treatment arms. |
| 8.0 | 03-Jul-2020 | Removal of lopinavir-ritonavir<br>Addition of intravenous immunoglobulin arm for children<br>Changes to corticosteroid dosing for children.<br>Addition of baseline serum sample in convalescent plasma randomisation |
| 9.0 | 10-Sep-2020 | Addition of synthetic neutralizing antibodies<br>Additional baseline data collection<br>Addition of countries outside UK |
| 9.1 | 18-Sep-2020 | Addition of information about vaccination of children of pregnant mothers receiving REGN10933+REGN10987 |
| 9.2 [not submitted in UK] | 15-Oct-2020 | Additional information for countries outside UK |
| 10.0 | 26-Oct-2020 | Addition of main randomisation part C<br>General updates to avoid duplication and improve clarity |
| 10.1 | 01-Nov-2020 | Additional information for pregnant women |
| 11.0 | 19-Nov-2020 | Addition of colchicine to main randomisation part A<br>Removal of azithromycin from main randomisation part A<br>Change in randomisation ratio in main randomisation part A from 2:1 to 1:1 |
| 11.1 | 21-Nov-2020 | Clarification of colchicine age thresholds |
| 11.2 [not submitted in UK] | 01-Dec-2020 | Addition of modified aspirin dose if 150mg not available |
| 12.0 | 10-Dec-2020 | Allow second randomisation of children without first randomisation |
| 12.1 | 16-Dec-2020 | Clarification of change in V12.0 |
| 13.0 | 26-Jan-2021 | Addition of baricitinib and anakinra (and change to allocation ratio in second randomisation for children); addition of pregnancy test for women of child-bearing potential (and change to colchicine eligibility);<br>removal of tocilizumab for adults; removal of convalescent plasma and additional assessment of antibody-based therapy; addition of dexamethasone as substitute if methylprednisolone unavailable |
| 14.0 | 15-Feb-2021 | Addition of Early Phase Assessments; the inclusion of dimethyl fumarate for initial early phase assessment; restriction of main randomisation part B to children with COVID-19 pneumonia; modification of barictinib and tocilizumab co-administration guidance |
| 15.0 | 12-Apr-2021 | Removal of aspirin and colchicine; addition of infliximab and high-dose corticosteroids (ex-UK only) |
| 15.1 [not submitted in UK] | 18-May-2021 | Addition of South Africa |

| Version number | Date | Brief Description of Changes |
|---|---|---|
| 16.0 | 05-Jul-2021 | Removal of REGN-COV2 and main randomisation part B<br>Removal of infliximab from main randomisation part E (and associated endemic infection monitoring section)<br>Addition of empagliflozin as main randomisation part F and metabolic outcomes<br>Addition of India, Sri Lanka and Pakistan |
| V16.1 | 08-Jul-2021 | Clarification of design in introduction |
| V17.0 | 06-Aug-2021 | Addition of additional exclusion criteria and safety monitoring for empagliflozin arm<br>Removal of corticosteroids and intravenous immunoglobulin in main randomisation part A (for children) |
| V17.1 | 10-Aug-2021 | Clarification of design for children |
| V18.0 | 13-Oct-2021 | Update to consent section<br>Change in primary outcome and sample size for DMF comparison<br>Clarification of eligibility for PIMS-TS randomisation<br>Removal of 3 month follow-up form for non-UK countries |
| V18.1 | 24-Oct-2021 | Clarification of witnesses for consent of children |
| V19.0 | 12-Nov-2021 | Addition of baloxavir marboxil, oseltamivir, and low-dose corticosteroids as randomised comparisons each vs. usual care alone for patients with influenza (in UK only).<br>Removal of early phase assessment of dimethyl fumarate.<br>Updated statistical analysis section to align with statistical analysis plan and include influenza analyses. |
| V19.1 | 16-Nov-21 | Clarification of baloxavir and weight eligibility |
| V20.0 | 29-Nov-21 | Removal of baricitinib.<br>Extension of COVID-19 high-dose corticosteroid and empagliflozin comparisons to other countries. |
| V21.0 | 17-Dec-21 | Addition of sotrovimab and molnupiravir.<br>Addition of baseline and follow-up samples.<br>Re-randomisation of patients recruited >6 months ago. |
| V21.1 | 19-Dec-21 | Clarifications post-REC review. |
| V22.0 | 19-Jan-22 | Addition of Paxlovid. (Not implemented.) |
| V23.0 | 08-Mar-22 | Clarifications following MHRA review. UKOSS added to section 3.1.3. Extension of molnupiravir to other countries. Removal of tocilizumab/anakinra for PIMS-TS. |
| 23.1 | 15-Mar-22 | Correction of footnotes |

#### Completed comparisons

The last version of the protocol to include the IMP is shown in the table above.

| IMP | Citation |
|---|---|
| Hydroxycholoroquine | New Engl J Med 2020; 383: 2030-40 |
| Dexamethasone (COVID-19) | New Engl J Med 2021; 384: 693-704 |
| Lopinavir-ritonavir | Lancet 2020; 396: 1345-1352 |
| Azithromycin | Lancet 2021; 397: 605-12 |
| Convalescent plasma | Lancet 2021; 397: 2049-59 |
| Tocilizumab | Lancet 2021; 397: 1637-1645 |
| Aspirin | Lancet 2022; 397: 143-151 |
| Colchicine | Lancet Resp Med 2021; 9: 1419-26 |
| REGN-COV2 | Lancet 2022; 399: 665-76 |
| Methylprednisolone (PIMS-TS) | Analysis ongoing |
| Intravenous immunoglobulin (PIMS-TS) | Analysis ongoing |
| Tocilizumab (PIMS-TS) | Follow-up ongoing |
| Anakinra (PIMS-TS) | Follow-up ongoing |
| Dimethyl fumarate | Analysis ongoing |
| Baricitinib | Medrxiv: 10.1101/2022.03.02.22271623v1 |

### 8 APPENDICES

#### 8.1 Appendix 1: Information about the treatment arms

All patients will receive usual care in the participating hospital.

**Corticosteroids:** RECOVERY is assessing the effects of corticosteroids in two different contexts: higher dose *vs* usual care in adults with COVID-19 and hypoxia; and lower dose dexamethasone in adults with influenza and hypoxia (UK only).

Favourable modulation of the immune response is considered one of the possible mechanisms by which corticosteroids might be beneficial in the treatment of severe acute respiratory coronavirus infections, including influenza, COVID-19, SARS and MERS. Common to severe cases of these infections is the presence of hypercytokinemia and the development of acute lung injury or acute respiratory distress syndrome (ARDS).[11-14] Pathologically, diffuse alveolar damage is found in patients who die from these infections.[15]

*Corticosteroids in influenza*
RECOVERY and other randomised trials have now demonstrated the benefit of corticosteroids in hypoxic COVID-19 patients.[16,17] However, the potential role of corticosteroids in severe influenza remains uncertain, with differing practices and controversy. Whilst observational studies report higher mortality associated with the use of corticosteroids in severe influenza, these studies are prone to biases, with a major concern being confounding by indication (the propensity to use corticosteroids only in the more severe patients as a rescue therapy).[18] In practice, use of corticosteroids in severe influenza is variable and widespread.[19] This therapeutic dilemma will only be resolved through an adequately powered randomised trial.

*Corticosteroids in COVID-19*
RECOVERY showed that dexamethasone at a dose of 6mg once daily for ten days or until discharge (whichever happens earliest) provided a significant reduction in mortality. Combining the IL-6 inhibitor tocilizumab with low dose dexamethasone resulted in a further reduction in mortality. This raises the question whether simply increasing the dose of corticosteroid could confer a similar clinical benefit to that of adding tocilizumab, but at substantially lower cost. Of note, even with dexamethasone 6mg and tocilizumab, mortality remained high at 29%. Although other randomised clinical trials in critically ill COVID-19 patients have used higher doses of dexamethasone (20mg once daily for five days followed by 10mg once daily for a further five days) and reported clinical benefit, these doses have not been compared with the lower dose used in RECOVERY. There is, therefore, uncertainty regarding the optimal dose of corticosteroids in moderate to severe COVID-19.Uncertainty remains about whether higher doses of corticosteroids may provide additional benefit in adults with hypoxia hospitalised with COVID-19.

Unlike lower doses, higher doses (>15mg dexamethasone) would completely saturate cytosolic glucocorticoid receptors and have enhanced non-genomic effects. In conditions where rapid control of inflammatory processes are required, short-term, high to very high doses of corticosteroids are used e.g.

- Sepsis dexamethasone 7.5 - 15mg equivalent daily[20]

- ARDS: dexamethasone 20mg for five days followed by 10mg for five days[21]
- Bacterial meningitis: dexamethasone 40mg daily for four days[22]
- Tuberculous Meningitis dexamethasone 0.4mg/kg/day for 7 days then reducing over 8 weeks.[23]
- Rheumatoid arthritis flare: dexamethasone 120mg pulse therapy.[24]
- Community acquired pneumonia: dexamethasone 0.6mg/day for 2 days and methyl prednisolone 200mg /day then 80mg /day for 10 days.[25]

**Empagliflozin:** Sodium glucose co-transporter 2 inhibitors (SGLT-2i) decrease glucose and insulin levels, and shift energy metabolism to an increased reliance on lipid oxidation, with a reduced reliance on glucose, and inhibition of glycolysis.[26] This mechanism may be particularly important in COVID-19, as SARS-CoV-2 may depend on the glycolytic pathway for its replication, stimulating lipogenesis, which appears to be one of the key drivers of cellular damage.[27,28] SGLT-2i rapidly improve endothelial function, possibly because of reduced oxidative stress.[29] SGLT-2i have significant anti-inflammatory effects, reducing levels of C-reactive protein and interleukin-6.[30] Experimental studies have also shown reduced activation of the NLRP3 inflammasome.[31] SGLT-2i increase erythropoiesis resulting in increased haematocrit,[32,33] and together with improved endothelial function[29] may improve oxygen delivery to tissues. Moreover, SGLT-2i result in reduced extracellular volume in patients with fluid overload,[34,35] and appear to reduce pulmonary artery pressure in patients with heart failure rapidly,[36] leading to haemodynamic decongestion. Thus, SGLT-2i may favourably affect multiple processes, including but not limited to energy metabolism, endothelial function, oxidative stress, inflammation and autophagy, which are dysregulated during a major acute illness such as COVID-19. The DARE-19 trial compared dapagliflozin 10 mg with placebo for 30 days among 1250 patients admitted to hospital with COVID-19 who had mild hypoxia ($SpO_2$ ≥94% on ≤5 L/min oxygen) and at least one risk factor (hypertension, type 2 diabetes mellitus, atherosclerotic cardiovascular disease, heart failure or chronic kidney disease).[37] The treatment was well tolerated (11% discontinued prematurely with similar proportion in treatment and placebo group). The hazard ratio for the co-primary outcome of organ failure (non-invasive or invasive ventilation, requirement for cardiovascular support or new/worsened heart failure, doubling of creatinine or dialysis) or death was 0.80 (95% CI 0.58-1.10; 70 *vs* 86 events).[38] Although this trial lacked statistical sensitivity, it supports the rationale for a larger trial.

**Sotrovimab [UK only]:** Sotrovimab (VIR-7831) is a neutralising monoclonal antibody targeting the SARS-CoV-2 spike glycoprotein receptor binding domain. It was identified by screening antibodies from a patient who had been infected during the 2003 SARS-CoV-1 outbreak, and its ability to also neutralise SARS-CoV-2 implies that its binding site is highly conserved, maybe meaning mutational escape will be difficult.[39] The Fc portion of the parent antibody has been modified to extend sotrovimab's half-life to around 49 days. It is given as a single intravenous dose and been well tolerated in clinical studies, although occasional serious hypersensitivity reactions have occurred.

It is licenced in the UK for the treatment of COVID-19 in patients who do not require oxygen and are at high risk of developing severe disease (at a 500 mg dose). The COMET-ICE trial, conducted in 583 such patients, showed that when given within five days of symptom onset it reduced the risk of hospitalisation by 85%, from 7% in the control group to 1% in the sotrovimab group.[40] Evidence in hospitalised patients is limited, and the sotrovimab arm of ACTIV-3 was stopped due to futility after recruiting 344 participants, although no safety concerns were raised.[41] However, by recruiting around 10,000 patients, RECOVERY subsequently showed that another neutralising monoclonal antibody treatment (casirivimab+imdevimab) reduced mortality by 20% in hospitalised patients who were anti-spike antibody negative at baseline.

The Omicron SARS-CoV-2 variant that emerged in late 2021 has multiple spike protein mutations, which have led to its rapid expansion in immune populations. These also appear to cause near complete loss of neutralising activity by the monoclonal antibodies in casirivimab+imdevimab,[42] and reduce the neutralising activity of Sotrovimab about 10-fold.[43,44] Data comparing the peak and day 29 concentrations following 2.4 g casirivimab+imdevimab and 500 mg Sotrovimab demonstrate much lower concentrations of Sotrovimab.[45] These pharmacodynamics and pharmacokinetic considerations underly the selection of a 1000 mg dose in this trial. The published safety of Sotrovimab and higher doses of other anti-spike human monoclonal antibodies (including the 8g dose of casirivimab+imdevimab used in RECOVERY) do not suggest a safety concern with this increased dose.

**Molnupiravir [UK only]:** Molnupiravir is a prodrug of the ribonucloside analogue N-hydroxycytidine (NHC), being rapidly converted into this form in plasma after absorption. NHC is then converted into the active triphosphate form in host cells by endogenous kinases. The SARS-CoV-2 viral RNA polymerase incorporates this into nascent viral RNA, resulting in copying errors that accumulate every replication cycle, ultimately preventing replication by a mechanism known as error catastrophe. This molecular target is conserved between Coronaviruses, and appears to have a high genetic barrier to resistance.[46] Molnupiravir is given orally and has been well tolerated in clinical studies so far, with infrequent reports of gastrointestinal and allergic reactions.

Molnupiravir is licensed in the United Kingdom for the treatment of mild-moderate COVID-19 within 5 days of symptom onset. In the MOVe-OUT trial of 1433 such patients it reduced the risk of hospitalisation or death by 30%, from 9.7% in the placebo group to 6.8% in molnupiravir group.[47] Evidence in hospitalised patients is limited, and the MOVe-IN trial randomised patients 1:1:1:1 to placebo vs. molnupiravir at 3 different doses (200mg, 400mg, 800mg). This study was abandoned after recruiting 304 inpatients as the manufacturer decided it was unlikely to demonstrate clinical benefit, although no safety concerns were raised.[48] However, the study was underpowered to identify moderate but important benefits in hospitalised patients, so a larger trial is needed.

**Paxlovid [UK only]:** Paxlovid is a combination of PF-07321332 (nirmatrelvir) and ritonavir. Nirmatrelvir is a 3-chymotrypsin-like protease inhibitor which inhibits cleavage of polyproteins involved in viral replication.[49] It is packaged with ritonavir which inhibits its CYP3A-dependent metabolism and hence increases the plasma concentration of nirmatrelvir. It is approved in the UK for the treatment of adults with COVID-19 who do not require supplemental oxygen and are at increased risk of progression to severe COVID-19.[50]

Its approval is based on the interim analysis of the EPIC-HR trial in which 2246 participants with COVID-19 (symptom onset ≤5 days previously) were randomised to receive Paxlovid (300/100 mg) or placebo twice daily for 5 days. The primary outcome is the proportion of participants with COVID-19 related hospitalisation or death within 28 days of randomisation.

In the interim analysis, 8/1037 (0.8%) allocated Paxlovid *vs* 66/1046 (6.3%) allocated placebo.[51] In an interim analysis of 774 participants, adverse events were similar between the two groups: 19% among those allocated Paxlovid *vs* 21% among those allocated placebo. Most were mild; only 1.7% *vs* 6.6% were serious and 2.1% *vs* 4.1% led to discontinuation.[52] SARS-CoV-2 main protease polymorphisms associated with reduced sensitivity to nirmatrelvir have been identified.[51] Their frequency and clinical significance is not yet known. Cross-resistance between nirmatrelvir and anti-SARS-CoV-2 monoclonal antibodies, molnupiravir or remdesivir are not expected given their different mechanisms of action.

**Baloxavir marboxil [UK only]:** Baloxavir marboxil is a cap-dependent endonuclease (CEN) inhibitor. CEN is an influenza virus-specific enzyme in the polymerase acidic subunit of the viral RNA polymerase complex. Through its action on CEN, baloxavir inhibits the transcription of influenza virus genomes resulting in inhibition of influenza A and B virus replication. It is approved in the USA, Japan, Australia, Europe, and the United Kingdom for the treatment of uncomplicated influenza and for post-exposure prophylaxis in individuals aged 12 years and older. Baloxavir is given in 2 oral doses (on day 1 and day 4) and is well tolerated, with allergic reactions being the only reported adverse reactions.

Baloxavir is not approved for the treatment of complicated influenza. A phase III placebo-controlled trial of baloxavir in adults hospitalised with severe influenza (Flagstone NCT03684044) did not find a significant reduction in the primary endpoint of time to clinical improvement (personal communication, Roche). However, time to clinical improvement, time to clinical response, influenza related complications, mortality, and time to cessation of viral shedding were all in favour of baloxavir. Fewer adverse events were observed in the baloxavir arm than in the standard of care arm. The Flagstone trial was small, comparing 214 subjects who received baloxavir with 125 who received usual care alone, and a larger study is need to determine whether baloxavir has modest but clinically relevant benefit in patients hospitalised with influenza.

**Oseltamivir [UK only]:**

The neuraminidase inhibitors (oseltamivir and zanamivir) are influenza specific antivirals that have been shown in randomised controlled trials to improve outcomes in uncomplicated influenza and to be effective as post-exposure prophylaxis. They have not, however, been shown to be effective in patients hospitalised with severe influenza. Although observational studies have reported clinical benefit in patients hospitalised with severe influenza, there are no randomised controlled trial data. Consequently, the use of neuraminidase inhibitors in this patient population is variable. A randomised controlled trial of neuraminidase inhibitors in patients hospitalised with severe influenza has been recommended by an expert group convened by the Academy of Medical Sciences and the Wellcome Trust, and most clincians would welcome such a trial.[53,54] The duration of treatment (5 days, or 10 days if the patient is immunosuppressed in the opinion of the managing clinician) is the same as that used in clinical practice and in the Summary of Product Characteristics.

RECOVERY
Randomised Evaluation of COVID-19 Therapy

#### 8.2 Appendix 2: Drug specific contraindications and cautions

**Corticosteroid**

Contraindications:

- Known contra-indication to short-term corticosteroid.
- Patients with suspected or confirmed influenza co-infection are not eligible for the high-dose dexamethasone comparision for COVID-19 (Randomisation part E).
- Patients in the UK with suspected or confirmed SARS-CoV-2 co-infection are not eligible for the low-dose dexamethasone comparison for influenza infection because of the proven benefits of dexamethasone in COVID-19 (Randomisation part I).
- Patients eligible for the Paxlovid comparison (Randomisation part L) will be excluded by the randomisation system from the high-dose dexamethasone comparison for COVID-19 (Randomisation part E) in view of the potential interaction between Paxlovid and dexamethasone.
- Current use of Paxlovid, ritonavir or other potent CYP3A inhibitors.

Cautions:

- Endemic infections may be screened for as required by local practice.
- Other immunomodulatory therapies are not contraindicated, but investigators should consider the total burden of therapy (eg, combining IL-6 receptor antagonist therapy with high-dose dexamethasone).

**Empagliflozin**

Contraindications:

- Type 1 diabetes mellitus (or post-pancreatectomy diabetes)
- Pregnancy and breast-feeding
- History of ketoacidosis
- Other patients with diabetes: blood ketones ≥1.5 mmol/L (or urine ketones ≥2+ if near-patient testing for blood ketones unavailable). Such patients are eligible once their ketosis has resolved.

Cautions:

- Participants with diabetes allocated empagliflozin should have regular checks of blood ketones (or urine ketones if blood ketone testing is unavailable)[q]. Blood ketones should be checked twice daily or urine ketones daily (or if clinical concern). If blood ketones rise ≥1.5 mmol/L (or urine ketones ≥2+), clinicians should:
    - Ensure adequate fluid and calorific intake
    - Consider increasing insulin dose (if on insulin)
    - Inform local diabetes team (if available) and treat ketosis using local protocols
    - Consider discontinuing empagliflozin until ketosis resolves
- Clinicians should consider temporarily discontinuing empagliflozin in participants with diabetes mellitus who cannot maintain oral calorific intake (until nutrition is restored)
- Clinicians should be aware of “euglycaemic ketoacidosis” which occurs with empagliflozin and should check ketones (ideally blood) if this is suspected (e.g. unexplained metabolic acidosis)

[q] These are near-patient tests and no sample will be retained for research purposes.

- Empagliflozin does not cause hypoglycaemia alone, but may do so in combination with insulin or insulin secretagogues. Doses of these other medications may need to be temporarily modified while the participant is taking empagliflozin
- Empagliflozin causes an osmotic diuresis so careful fluid balance assessment is required
- Empagliflozin increases the risk of mycotic genital infections (e.g. candidiasis) which are usually easily treated with topical therapy. It is unclear whether it causes Fournier's gangrene (a very rare genital infection), but clinicians should be aware.

**Sotrovimab**
Contraindications:
- Weight <40kg (if <18 years old; no weight restriction for adults)
- Known hypersensitivity to sotrovimab or the drug product excipients

Cautions: no dose adjustment for kidney or liver function is required.

**Molnupiravir**
Contraindications:
- Age <18 years
- Pregnancy or breast-feeding. Women of child-bearing potential should be advised not to get pregnant while taking molnupiravir or for 4 days after completing the course
- Known hypersensitivity to molnupiravir or its excipients
- Prior treatment with molnupiravir during the index illness

Cautions: no dose adjustment for kidney or liver function is required.

**Paxlovid**
Contraindications:
- Age <18 years
- Severe hepatic impairment (Child-Pugh class C)
- Severe renal impairment (eGFR <30 mL/min/1.73m$^2$)
- First trimester (i.e. first 12 weeks) of pregnancy
- Prior treatment with Paxlovid during the index illness
- Known hypersensitivity to nirmatrelvir (PF-07321332) or ritonavir (including hereditary problems of galactose intolerance, total lactase deficiency or glucose-galactose malabsorption)
- Concomitant therapy with drugs that are highly dependent on CYP3A for clearance and for which elevated plasma concentrations are associated with serious reactions.
  - α1-adrenoreceptor antagonist (afluzosin)
  - Analgesics (pethidine, piroxicam, propoxyphene)
  - Anti-anginal (ranolazine)
  - Anti-arrhythmics (amiodarone, bepridil, dronaderone, encainide, flecainide, propafenone, quinidine)
  - Antibacterials (fusidic acid)
  - Anticancer (neratinib, venetoclax)
  - Anti-gout (colchicine)
  - Antihistamine (astemizole, terfenadine)
  - Antipsychotics (lurasidone, pimozide, clozapine, quietiapine)
  - Ergot derivatives (dihydroergotamine, ergonovine, ergotamine, methylergonovine)

    - Gastrointestinal motility agent (cisapride)
    - Lipid modifying agents (lovastatin, simvastatin, lomitapide)
    - PDE5 inhibitors (avanafil, vardenafil, sildenafil)
    - Sedatives (clorazepate, diazepam, estazolam, flurazepam, triazolam, oral midazolam)
    - High-dose dexamethasone (>6 mg base once daily)
- Concomitant therapy with drugs that are potent CYP3A inducers (which may reduce plasma PF-07321332/ritonavir concentrations):
    - Anticancer (apalutamide)
    - Anticonvulsants (carbamazepine, phenobarbital, phenytoin)
    - Antimycobacterials (rifampicin)
    - Herbal products (St John's Wort)

Cautions:

- Since ritonavir may decrease the efficacy of combined oral contraceptives, women using them should be advised to use effective alternative contraception or an additional barrier method until after one complete menstrual cycle after stopping.
- The necessity of using other drugs metabolised by CYP3A (or which induce or inhibit CYP3A) should be reviewed.[r]
- 
- Patients with moderate renal impairment (eGFR ≥30 <60 mL/min/1.73m$^2$) should receive 150/100 mg twice daily (ie, one PF-07321332 tablet and one ritonavir tablet twice daily). Local pharmacists should remove one PF-07321332 tablet from each dose in the packet provided to the participant (see pharmacy manual at https://www.recoverytrial.net/for-site-staff/pharmacy for further detail).

Managing clinicians may consider if it is appropriate to temporarily withhold contraindicated concomitant medication while receiving Paxlovid or consider alternatives. The risks and benefits of doing so should be explained to the participant. Clear plans should be made about restarting such treatment and – if necessary – any checks that need to be made beforehand. These plans should be communicated to the participant and their general practitioner in the discharge summary.

**Baloxavir Marboxil**

Contraindications:

- Weight <40kg (regardless of age)
- Known hypersensitivity to baloxavir marboxil or the drug product excipients
- Participants who have received baloxavir marboxil for the current influenza infection

**Oseltamivir**

Contraindications:

- Known hypersensitivity to oseltamivir or the drug product excipients
- Participants who have received oseltamivir for the current influenza infection

Cautions:

- Dose should be reduced in presence of renal impairment
    - eGFR ≥30 mL/min/1.73m$^2$: dose as in normal renal function (75 mg twice daily)

[r] A list is available at https://www.covid19-druginteractions.org/. Please note these lists may not be exhaustive.

  - eGFR ≥10 <30 mL/min/1.73m$^2$: 75 mg once daily
  - eGFR <10 mL/min/1.73m$^2$: 75 mg as a single dose on day 1
- Dose should be reduced for adult patients weighing <40 kg to 60 mg twice daily

#### 8.3 Appendix 3: Paediatric dosing information

Children (aged <18 years old) will be recruited in the UK only.

##### Randomisation of children with COVID-19 Pneumonia (Patients <12 years of age will NOT be eligible)

| Arm | Route | Weight | Dose |
|---|---|---|---|
| **No additional treatment** | - | - | - |
| **Sotrovimab** | Intravenous | Children <12 years old excluded | |
| | | <40 kg | Excluded regardless of age |
| | | ≥40 kg | 1000 mg intravenous in 100 mL of 0.9% NaCl or 5% dextrose over 1 hour |

#### Influenza Randomisations

<table>
<tr><th>Arm</th><th>Route</th><th>Weight/Age</th><th>Dose</th></tr>
<tr><td rowspan="3">Oseltamivir<br><br>- 30, 45 and 75 mg capsules<br><br>- Oral suspension [a]</td><td rowspan="3">Oral<br><u>or</u><br>Other enteral routes</td><td>Less than 36 weeks corrected gestational age</td><td>1 mg/kg twice daily for 5 days [b]</td></tr>
<tr><td>0 - 12 months (≥36 weeks corrected gestational age)</td><td><table><tr><th>Weight (kg)</th><th>Dose</th></tr><tr><td><10</td><td>3 mg/kg twice daily for 5 days [b]</td></tr><tr><td>≥ 10</td><td>30 mg twice daily for 5 days [b]</td></tr></table></td></tr>
<tr><td>≥ 1 year</td><td><table><tr><th>Weight (kg)</th><th>Dose</th></tr><tr><td><10</td><td>3 mg/kg twice daily for 5 days [b]</td></tr><tr><td>≥ 10 to 15</td><td>30 mg twice daily for 5 days [b]</td></tr><tr><td>> 15 to 23</td><td>45 mg twice daily for 5 days [b]</td></tr><tr><td>> 23 to 40</td><td>60 mg twice daily for 5 days [b]</td></tr><tr><td>> 40</td><td>75 mg twice daily for 5 days [b]</td></tr></table><br>Those within significant renal impairment (CrCl 10 - 30 mL/min) should receive once daily dosing. Those with CrCl <10 ml/min should receive only a single dose on day 1.</td></tr>
<tr><td>Baloxavir marboxil<br><br>- 20 and 40 mg tablets</td><td>Oral<br><u>or</u><br>Other enteral routes</td><td>≥ 12 years old</td><td><table><tr><th>Weight (kg)</th><th>Dose</th></tr><tr><td><40</td><td>Not eligible</td></tr><tr><td>≥40 < 80</td><td>40 mg on day 1 and day 4</td></tr><tr><td>≥ 80</td><td>80 mg on day 1 and day 4</td></tr></table></td></tr>
<tr><td rowspan="2">Low dose corticosteroids</td><td rowspan="2">Oral<br><u>or</u><br>Other enteral routes<br><u>or</u><br>Intravenous</td><td>Less than 36 weeks corrected gestational age</td><td>Hydrocortisone (IV)<br>0.5 mg/kg every 12 hours for 7 days and then 0.5mg/kg once daily for 3 days</td></tr>
<tr><td>≥0 month (≥36 weeks corrected gestational age)</td><td>Dexamethasone:<br>150 micrograms/kg (as base) once daily (max: 6 mg once daily) for 10 days (or until discharge if sooner)</td></tr>
</table>

[a] Public Health England advises that oseltamivir oral suspension should be reserved for children under the age of 1 year. Children over 1 year of age, those with swallowing difficulties, and those receiving nasogastric oseltamivir, should use capsules which can be opened and mixed into an appropriate sugary liquid.
[b] 10 days if immunocompromised

#### 8.4 Appendix 4: Use of IMPs in pregnant and breastfeeding women

All trial drugs (except empagliflozin, sotrovimab, molnupiravir, Paxlovid and baloxavir) have been used in pregnant women with pre-existing medical disorders where benefits outweigh the risks to fetus or woman, including in the first trimester. The existing data related to each drug is summarised below. The potential inclusion of any pregnant women should be discussed with a consultant obstetrician (or obstetric physician) and all consent discussions should be documented in the medical records.

##### Corticosteroids

Prednisolone or, in women unable to take oral medicine, hydrocortisone or methylprednisolone are recommended instead of dexamethasone treatment in light of accumulating evidence that repeated doses of dexamethasone have deleterious effects on long-term neurodevelopment of the fetus.[55-57] While 90% dexamethasone is transferred transplacentally to the fetus, both hydrocortisone and prednisolone are converted by 11β-hydroxysteroid dehydrogenase to inactive glucocorticoids and considerably less drug is transferred to the fetus. Glucocorticoids can worsen maternal glycaemic control, so blood glucose should be checked and managed appropriately. Otherwise there is no convincing evidence that prednisolone use is associated with increased rates of adverse pregnancy outcomes when taken in the first trimester or later pregnancy.[58] Very low concentrations of prednisolone enter breastmilk. There is a paucity of data about pharmacological use of hydrocortisone, but it is likely that this is also safe when breastfeeding,[58] as also reviewed in the Lactmed database (www.ncbi.nlm.nih.gov/books/NBK501076/). Prednisolone (or hydrocortisone) should be used in breastfeeding women, in preference to dexamethasone.

##### Empagliflozin

Empagliflozin is not recommended for use in pregnant or breastfeeding women. Empagliflozin will only be included in the randomisation of women of child-bearing potential if they have had a negative pregnancy test since admission.

##### Sotrovimab

There are no data from the use of sotrovimab in pregnant women. Since sotrovimab is a human immunoglobulin G animal studies have not been evaluated with respect to reproductive toxicity. No off-target binding was detected in a cross-reactive binding assay using a protein array enriched for human embryofetal proteins. Since sotrovimab is a human immunoglobulin G, it has the potential for placental transfer from the mother to the developing foetus. The potential treatment benefit or risk of placental transfer of sotrovimab to the developing foetus is not known. Sotrovimab may be used during pregnancy where the expected benefit to the mother justifies the risk to the foetus.

There are no data on the excretion of sotrovimab in human milk. The potential treatment benefit or risk to the newborn or infants via breastfeeding is not known. Decisions on whether to breastfeed during treatment or to abstain from sotrovimab therapy should take into account the benefit of breast-feeding for the child and the benefit of therapy for the woman.

##### Molnupiravir

Molnupiravir is not recommended for use in pregnant or breastfeeding women. Molnupiravir will only be included in the randomisation of women of child-bearing potential if they have had a negative pregnancy test since admission.

**Paxlovid**
Preclinical animal reproductive toxicity studies have not identified adverse effects on fetal morphology or embryo-fetal viability in rat or rabbit models with doses of nirmatrelvir up to 12 times the human dose (equivalence based on predicted AUC concentrations). The offspring of pregnant rabbits administered 24 times the equivalent human dose, lower fetal body weights were observed but evidence of maternal toxicity was described (impact on weight gain/food consumption).[52] There is a large amount of published evidence relating to the safety of ritonavir in human pregnancy, collected from antiretroviral and HIV/AIDS pregnancy registries. Overall, these data do not provide compelling evidence that ritonavir use in the first trimester is associated with an increased risk of malformation above the expected background rate of 2-3%. As Paxlovid has not previous been given to pregnant women, women in the first trimester of pregnancy will be excluded from this comparison.

**Baloxavir marboxil**
There are no data from the use of baloxavir marboxil in pregnant women. Animal studies do not indicate direct or indirect harmful effects with respect to reproductive toxicity. Baloxavir treatment may be of particular benefit to pregnant women with influenza, as they are at increased risk of developing severe disease. Preclinical animal models of exposure in pregnancy do not provide evidence of adverse embryo-fetal effects at doses up to five and seven times the human therapeutic dose respectively. The risk of harm from baloxavir in pregnancy is likely to be low given the animal model data, together with the therapeutic target for baloxavir being a virus specific enzyme. It is unknown whether baloxavir marboxil or baloxavir are excreted in human milk, and baloxivir may be considered.

**Oseltamivir**
There are observational data on the use of oseltamivir in pregnant women including >1000 women exposed during the first trimester. These studies found no evidence of adverse embryo-fetal effects. Oseltamivir is currently used in pregnant women. Its use may also be considered in breastfeeding women: it is excreted in breast milk but at low concentrations that would be subtherapeutic dose to the infant.

#### 8.5 Appendix 5: Organisational Structure and Responsibilities

##### Chief Investigator

The Chief Investigator has overall responsibility for:

(i) Design and conduct of the Study in collaboration with the Trial Steering Committee;
(ii) Preparation of the Protocol and subsequent revisions.

##### Trial Steering Committee

The Trial Steering Committee (see below for list of members) is responsible for:

(i) Agreement of the final Protocol and the Data Analysis Plans;
(ii) Reviewing progress of the study and, if necessary, deciding on Protocol changes;
(iii) Review and approval of study publications and substudy proposals;
(iv) Reviewing new studies that may be of relevance.

##### International Steering Committee

The International Steering Committee (see below for list of members) is responsible for:

(i) Reviewing progress of the study in sites outside the UK;
(ii) Review of study publications and substudy proposals;
(iii) Considering potential new therapies to be included in sites outside the UK;
(iv) Assisting RCC in selection of LCCs;
(v) Reviewing new studies that may be of relevance.

##### Data Monitoring Committee

The independent Data Monitoring Committee is responsible for:

(i) Reviewing unblinded interim analyses according to the Protocol;
(ii) Advising the Steering Committee if, in their view, the randomised data provide evidence that may warrant a change in the Protocol (e.g. modification or cessation of one or more of the treatment arms).

##### Central Coordinating Office (CCO)

The CCO is responsible for the overall coordination of the Study, including:

(i) Study planning and organisation of Steering Committee meetings;
(ii) Ensuring necessary regulatory and ethics committee approvals;
(iii) Development of Standard Operating Procedures and computer systems
(iv) Monitoring overall progress of the study;
(v) Provision of study materials to RCCs/LCCs;
(vi) Monitoring and reporting safety information in line with the Protocol and regulatory requirements;
(vii) Dealing with technical, medical and administrative queries from LCCs.

**Regional Coordinating Centre (RCC)**
The RCCs are responsible for:

(i) Ensuring necessary regulatory and ethics committee approvals;
(ii) Provision of study materials to LCCs;
(iii) Dealing with technical, medical and administrative queries from LCCs.

**Local Clinical Centres (LCC)**
The LCC lead investigator and LCC clinic staff are responsible for:

(i) Obtaining all relevant local permissions (assisted by the CCO);
(ii) All trial activities at the LCC, including appropriate training and supervision for clinical staff;
(iii) Conducting trial procedures at the LCC in line with all relevant local policies and procedures;
(iv) Dealing with enquiries from participants and others.

#### STEERING COMMITTEE

(Major organisational and policy decisions, and scientific advice; blinded to treatment allocation)

| | |
|---|---|
| Chief Investigator | Peter Horby |
| Deputy Chief Investigator | Martin Landray |
| Clinical Trial Unit Lead | Richard Haynes |
| Co-investigators | Kenneth Baillie (Scotland Lead), Maya Buch, Saul Faust, Thomas Jaki, Katie Jeffery, Edmund Juszczak, Marian Knight, Wei Shen Lim, Marion Mafham, Alan Montgomery, Aparna Mukherjee, Andrew Mumford, Kathy Rowan, Guy Thwaites, Jeremy Day |

#### International Committees

| | |
|---|---|
| *Asia* | |
| Chair | Do Van Dung |
| Regional Lead Investigators | Guy Thwaites, Jeremy Day |
| Independent members: | Vietnam : Nguyen Ngo Quang, Prof. Binh<br>Indonesia: Erlina Burhan, Bachti Alisjahbana<br>Nepal: Janak Koirala, Sudha Basnet |
| Other members: | Evelyne Kestelyn, Buddha Basnyat, Pradip Gyanwali, Raph Hamers, Peter Horby |
| *Africa* | |
| Chair | TBC |
| Independent members: | Ghana: TBD<br>South Africa: TBD |
| Other members: | John Amuasi, Peter Horby, Jeremy Nel |

#### DATA MONITORING COMMITTEE

(Interim analyses and response to specific concerns)

| | |
|---|---|
| Chair | Peter Sandercock |
| Members | Janet Darbyshire, David DeMets, Robert Fowler,<br>David Lalloo, Mohammed Munavvar, Adilia Warris, Janet Wittes |
| Statisticians (non-voting) | Jonathan Emberson, Natalie Staplin |

### 9 REFERENCES

1. Zhu N, Zhang D, Wang W, et al. A Novel Coronavirus from Patients with Pneumonia in China, 2019. N Engl J Med 2020;382:727-33.
2. Shi R, Shan C, Duan X, et al. A human neutralizing antibody targets the receptor-binding site of SARS-CoV-2. Nature 2020;584:120-4.
3. Huang C, Wang Y, Li X, et al. Clinical features of patients infected with 2019 novel coronavirus in Wuhan, China. Lancet 2020;395:497-506.
4. Wang D, Hu B, Hu C, et al. Clinical Characteristics of 138 Hospitalized Patients With 2019 Novel Coronavirus-Infected Pneumonia in Wuhan, China. JAMA 2020.
5. Whittaker E, Bamford A, Kenny J, et al. Clinical Characteristics of 58 Children With a Pediatric Inflammatory Multisystem Syndrome Temporally Associated With SARS-CoV-2. JAMA 2020.
6. Zhou F, Yu T, Du R, et al. Clinical course and risk factors for mortality of adult inpatients with COVID-19 in Wuhan, China: a retrospective cohort study. Lancet 2020;395:1054-62.
7. Moss JWE, Davidson C, Mattock R, Gibbons I, Mealing S, Carroll S. Quantifying the direct secondary health care cost of seasonal influenza in England. BMC public health 2020;20:1464.
8. Holm S. A Simple Sequentially Rejective Multiple Test Procedure. Scandinavian Journal of Statistics 1979;6:65-70.
9. Venet D, Doffagne E, Burzykowski T, et al. A statistical approach to central monitoring of data quality in clinical trials. Clin Trials 2012;9:705-13.
10. Oversight of Clinical Investigations--A Risk-Based Approach to Monitoring. 2013. (Accessed 18 August 2017, at https://www.fda.gov/downloads/Drugs/GuidanceComplianceRegulatoryInformation/Guidances/UCM269919.pdf.)
11. Lau SKP, Lau CCY, Chan KH, et al. Delayed induction of proinflammatory cytokines and suppression of innate antiviral response by the novel Middle East respiratory syndrome coronavirus: implications for pathogenesis and treatment. J Gen Virol 2013;94:2679-90.
12. de Jong MD, Simmons CP, Thanh TT, et al. Fatal outcome of human influenza A (H5N1) is associated with high viral load and hypercytokinemia. Nat Med 2006;12:1203-7.
13. Liu Q, Zhou YH, Yang ZQ. The cytokine storm of severe influenza and development of immunomodulatory therapy. Cell Mol Immunol 2016;13:3-10.
14. Short KR, Veeris R, Leijten LM, et al. Proinflammatory Cytokine Responses in Extra-Respiratory Tissues During Severe Influenza. J Infect Dis 2017;216:829-33.
15. Xu Z, Shi L, Wang Y, et al. Pathological findings of COVID-19 associated with acute respiratory distress syndrome. Lancet Respir Med 2020;8:420-2.
16. RECOVERY Collaborative Group, Horby P, Lim WS, et al. Dexamethasone in Hospitalized Patients with Covid-19. N Engl J Med 2021;384:693-704.
17. W. H. O. Rapid Evidence Appraisal for COVID-19 Therapies Working Group, Sterne JAC, Murthy S, et al. Association Between Administration of Systemic Corticosteroids and Mortality Among Critically Ill Patients With COVID-19: A Meta-analysis. JAMA 2020;324:1330-41.
18. Lansbury LE, Rodrigo C, Leonardi-Bee J, Nguyen-Van-Tam J, Shen Lim W. Corticosteroids as Adjunctive Therapy in the Treatment of Influenza: An Updated Cochrane Systematic Review and Meta-analysis. Crit Care Med 2020;48:e98-e106.
19. Hui DS, Lee N, Chan PK, Beigel JH. The role of adjuvant immunomodulatory agents for treatment of severe influenza. Antiviral Res 2018;150:202-16.
20. Rochwerg B, Oczkowski SJ, Siemieniuk RAC, et al. Corticosteroids in Sepsis: An Updated Systematic Review and Meta-Analysis. Crit Care Med 2018;46:1411-20.
21. Villar J, Ferrando C, Martinez D, et al. Dexamethasone treatment for the acute respiratory distress syndrome: a multicentre, randomised controlled trial. Lancet Respir Med 2020;8:267-76.
22. Glimaker M, Brink M, Naucler P, Sjolin J. Betamethasone and dexamethasone in adult community-acquired bacterial meningitis: a quality registry study from 1995 to 2014. Clin Microbiol Infect 2016;22:814 e1-e7.
23. Thwaites GE, Nguyen DB, Nguyen HD, et al. Dexamethasone for the treatment of tuberculous meningitis in adolescents and adults. N Engl J Med 2004;351:1741-51.
24. Sadra V, Khabbazi A, Kolahi S, Hajialiloo M, Ghojazadeh M. Randomized double-blind study of the effect of dexamethasone and methylprednisolone pulse in the control of rheumatoid arthritis flare-up: a preliminary study. Int J Rheum Dis 2014;17:389-93.
25. van Woensel JB, van Aalderen WM, de Weerd W, et al. Dexamethasone for treatment of patients mechanically ventilated for lower respiratory tract infection caused by respiratory syncytial virus. Thorax 2003;58:383-7.

26. Daniele G, Xiong J, Solis-Herrera C, et al. Dapagliflozin Enhances Fat Oxidation and Ketone Production in Patients With Type 2 Diabetes. Diabetes Care 2016;39:2036-41.
27. Codo AC, Davanzo GG, Monteiro LB, et al. Elevated Glucose Levels Favor SARS-CoV-2 Infection and Monocyte Response through a HIF-1α/Glycolysis-Dependent Axis. Cell Metab 2020;32:437-46.e5.
28. Icard P, Lincet H, Wu Z, et al. The key role of Warburg effect in SARS-CoV-2 replication and associated inflammatory response. Biochimie 2021;180:169-77.
29. Solini A, Giannini L, Seghieri M, et al. Dapagliflozin acutely improves endothelial dysfunction, reduces aortic stiffness and renal resistive index in type 2 diabetic patients: a pilot study. Cardiovasc Diabetol 2017;16:138.
30. Bonnet F, Scheen AJ. Effects of SGLT2 inhibitors on systemic and tissue low-grade inflammation: The potential contribution to diabetes complications and cardiovascular disease. Diabetes & metabolism 2018;44:457-64.
31. Kim SR, Lee SG, Kim SH, et al. SGLT2 inhibition modulates NLRP3 inflammasome activity via ketones and insulin in diabetes with cardiovascular disease. Nat Commun 2020;11:2127.
32. Lambers Heerspink HJ, de Zeeuw D, Wie L, Leslie B, List J. Dapagliflozin a glucose-regulating drug with diuretic properties in subjects with type 2 diabetes. Diabetes, obesity & metabolism 2013;15:853-62.
33. Ghanim H, Abuaysheh S, Hejna J, et al. Dapagliflozin Suppresses Hepcidin And Increases Erythropoiesis. The Journal of clinical endocrinology and metabolism 2020;105.
34. Ohara K, Masuda T, Morinari M, et al. The extracellular volume status predicts body fluid response to SGLT2 inhibitor dapagliflozin in diabetic kidney disease. Diabetol Metab Syndr 2020;12:37.
35. Griffin M, Rao VS, Ivey-Miranda J, et al. Empagliflozin in Heart Failure: Diuretic and Cardiorenal Effects. Circulation 2020;142:1028-39.
36. Mullens W, Martens P, Forouzan O, et al. Effects of dapagliflozin on congestion assessed by remote pulmonary artery pressure monitoring. ESC Heart Fail 2020;7:2071-3.
37. Kosiborod M, Berwanger O, Koch GG, et al. Effects of dapagliflozin on prevention of major clinical events and recovery in patients with respiratory failure because of COVID-19: Design and rationale for the DARE-19 study. Diabetes, obesity & metabolism 2021;23:886-96.
38. Dapagliflozin in Respiratory Failure in Patients With COVID-19 - DARE-19. 2021. (Accessed 09-Jun-2021, at https://www.acc.org/Latest-in-Cardiology/Clinical-Trials/2021/05/14/02/40/DARE-19.)
39. Pinto D, Park YJ, Beltramello M, et al. Cross-neutralization of SARS-CoV-2 by a human monoclonal SARS-CoV antibody. Nature 2020;583:290-5.
40. Gupta A, Gonzalez-Rojas Y, Juarez E, et al. Early Treatment for Covid-19 with SARS-CoV-2 Neutralizing Antibody Sotrovimab. N Engl J Med 2021;385:1941-50.
41. NIH-Sponsored ACTIV-3 Clinical Trial Closes Enrollment into Two Sub-Studies. 2021. at https://www.nih.gov/news-events/news-releases/nih-sponsored-activ-3-clinical-trial-closes-enrollment-into-two-sub-studies.)
42. Wilhelm A, Widera M, Grikscheit K, et al. Reduced Neutralization of SARS-CoV-2 Omicron Variant by Vaccine Sera and monoclonal antibodies. MedRxiv 2021.
43. Cathcart AL, Havenar-Daughton C, Lempp FA, et al. The dual function monoclonal antibodies VIR-7831 and VIR-7832 demonstrate potent in vitro and in vivo activity against SARS-CoV-2. BioRxiv 2021.
44. Cao YC, Wang Y, Jian F, et al. B.1.1.529 escapes the majority of SARS-CoV-2 neutralizing antibodies of diverse epitopes. BioRxiv 2021.
45. GlaxoSmithKline. Xevudy - summary of product characteristics2021.
46. Agostini ML, Pruijssers AJ, Chappell JD, et al. Small-Molecule Antiviral beta-d-N (4)-Hydroxycytidine Inhibits a Proofreading-Intact Coronavirus with a High Genetic Barrier to Resistance. J Virol 2019;93.
47. Merck announces results from MOVe-OUT Study. 2021. at https://www.merck.com/news/merck-and-ridgeback-biotherapeutics-provide-update-on-results-from-move-out-study-of-molnupiravir-an-investigational-oral-antiviral-medicine-in-at-risk-adults-with-mild-to-moderate-covid-19/.)
48. Merck progress update. 2021. at https://www.merck.com/news/merck-and-ridgeback-biotherapeutics-provide-update-on-progress-of-clinical-development-program-for-molnupiravir-an-investigational-oral-therapeutic-for-the-treatment-of-mild-to-moderate-covid-19/.)
49. Owen DR, Allerton CMN, Anderson AS, et al. An oral SARS-CoV-2 Mpro inhibitor clinical candidate for the treatment of COVID-19. Science 2021;374:1586-93.
50. Regulatory approval of Paxlovid. 2021. (Accessed 13-Jan-2022, at https://www.gov.uk/government/publications/regulatory-approval-of-paxlovid.)
51. Food and Drug Administration. Fact sheet for healthcare providers: emergency use authorisation for Paxlovid. 2021.
52. Pfizer. Summary of Product Characteristics for Paxlovid. 2021.

53. Bradbury N, Nguyen-Van-Tam J, Lim WS. Clinicians' attitude towards a placebo-controlled randomised clinical trial investigating the effect of neuraminidase inhibitors in adults hospitalised with influenza. BMC Health Services Research 2018;18:311.
54. Academy of Medical Sciences, Wellcome Trust. Use of Neuraminidase Inhibitors in Influenza. 2015.
55. Tam EW, Chau V, Ferriero DM, et al. Preterm cerebellar growth impairment after postnatal exposure to glucocorticoids. Sci Transl Med 2011;3:105ra.
56. Newnham JP, Jobe AH. Should we be prescribing repeated courses of antenatal corticosteroids? Semin Fetal Neonatal Med 2009;14:157-63.
57. Chang YP. Evidence for adverse effect of perinatal glucocorticoid use on the developing brain. Korean J Pediatr 2014;57:101-9.
58. Flint J, Panchal S, Hurrell A, et al. BSR and BHPR guideline on prescribing drugs in pregnancy and breastfeeding-Part II: analgesics and other drugs used in rheumatology practice. Rheumatology (Oxford) 2016;55:1698-702.

### 10 CONTACT DETAILS

Website: www.recoverytrial.net
(copies of this protocol and related forms and information can be downloaded)

**RECOVERY Central Coordinating Office:**
Richard Doll Building, Old Road Campus, Roosevelt Drive, Oxford OX3 7LF
United Kingdom

**RECOVERY Vietnam:**
Oxford University Clinical Research Unit, Centre for Tropical Medicine, 764 Vo Van Kiet, District 5, Ho Chi Minh City, Vietnam


**RECOVERY Indonesia:**
Eijkman Oxford Clinical Research Unit (EOCRU), Eijkman Institute for Molecular Biology
Jl. P. Diponegoro No. 69, Jakarta-Indonesia 10430

**RECOVERY Nepal:**
Clinical Trial Unit, Oxford University Clinical Research Unit-Nepal, Patan Academy of Health Sciences, Kathmandu, Nepal

**RECOVERY Ghana:**
Kumasi Center for Collaborative Research in Tropical Medicine
KNUST, Southend Asuogya Road, GPS: AK-312-1059, Kumasi, Ghana

**RECOVERY South Africa:**
Wits Health Consortium, 31 Princess of Wales Terrace, Parktown, Johannesburg, South Africa

**RECOVERY Sri Lanka & Pakistan:**
National Intensive Care Surveillance - M.O.R.U, 2nd Floor, YMBA Building, Borella, Colombo 08, Sri Lanka

**RECOVERY India:**
Indian Council of Medical Research, Division of Epidemiology and Communicable Diseases, Ramalingaswami Bhavan, Ansari Nagar, ICMR-110029

#### To RANDOMISE a patient, visit:

---

RECOVERY
Randomised Evaluation of COVID-19 Therapy

Website: www.recoverytrial.net

---

#### Appendix 2: RECOVERY Trial Statistical Analysis Plan V3.2

### RECOVERY

Randomised Evaluation of COVID-19 Therapy

### Statistical Analysis Plan

**Version 3.2**

**Date: 17 December 2021**

Aligned with protocol version: 18.1, 29 October 2021

IRAS no: 281712
REC ref: EE/20/0101
ISRCTN: 50189673
EudraCT: 2020-001113-21

Nuffield Department of
POPULATION HEALTH

UNIVERSITY OF OXFORD

#### Abbreviations

| | |
|---|---|
| ADaM | Analysis Data Model |
| AE | Adverse event |
| CDISC | The Clinical Data Interchange Standards Consortium |
| CI | Confidence interval |
| COVID | Coronavirus-induced disease |
| CPAP | Continuous Positive Airway Pressure |
| CRP | C-reactive protein |
| DMC | Data Monitoring Committee |
| ECMO | Extra Corporeal Membrane Oxygenation |
| eCRF | Electronic case report form |
| ICD | International Classification of Diseases |
| ICNARC | Intensive Care National Audit and Research Centre |
| ITT | Intention to treat |
| MedDRA | Medical Dictionary for Regulatory Activities |
| OPCS-4 | National Health Service OPCS Classification of Interventions and Procedures version 4 |
| SARS | Severe acute respiratory syndrome |
| SARS-CoV-2 | Severe acute respiratory syndrome coronavirus 2 |
| $S/F_{94}$ ratio | Ratio of peripheral oxygen saturation to fractional inspired oxygen concentration when peripheral oxygen saturation at or below 94% |
| SSAR | Suspected serious adverse reaction |
| SUSAR | Suspected unexpected serious adverse reaction |
| TSC | Trial Steering Committee |

#### List of authors and reviewers (up to and including SAP version 1.1)

##### *Authors*

Dr Louise Linsell, Lead Trial Statistician, Nuffield Department of Population Health (NDPH), University of Oxford

Jennifer Bell, Trial Statistician, NDPH, University of Oxford

##### *Reviewers*

Professor Jonathan Emberson, Data Monitoring Committee (DMC) Statistician, NDPH, University of Oxford (prior to unblinded interim analysis of trial outcomes)

Professor Richard Haynes, Clinical Coordinator, NDPH, University of Oxford

Professor Peter Horby, Chief Investigator (CI), Nuffield Department of Medicine, University of Oxford

Professor Thomas Jaki, TSC Member, Department of Mathematics and Statistics, Lancaster University

Associate Professor Edmund Juszczak, TSC Member, NDPH, University of Oxford (until 6 July 2020)

Professor Martin Landray, Deputy CI, NDPH, University of Oxford

Professor Alan Montgomery, TSC Member, Nottingham Clinical Trials Unit, University of Nottingham

Dr Natalie Staplin, DMC Statistician, NDPH, University of Oxford (prior to unblinded interim analysis of trial outcomes)

#### List of authors and reviewers (version 2.0 onwards)

Professor Edmund Juszczak, TSC Member (University of Nottingham from 6 July 2020)

Professor Alan Montgomery (University of Nottingham), TSC Member

Professor Thomas Jaki (University of Cambridge) co-investigator and TSC Member

Enti Spata, Trial Statistician, NDPH, University of Oxford

Professor Richard Haynes, Clinical Coordinator, NDPH, University of Oxford

Professor Martin Landray, Deputy CI, NDPH, University of Oxford

Professor Peter Horby, CI, Nuffield Department of Medicine, University of Oxford

#### Roles and responsibilities

##### *Trial Statisticians*

*Until 30th September 2020:* Dr Louise Linsell and Jennifer Bell (NDPH, University of Oxford)

Role: To develop the statistical analysis plan (blinded to trial allocation) and conduct the final comparative analyses for Lopinavir-Ritonavir¸ Corticosteroid (dexamethasone) and Hydroxychloroquine (main randomisation part A).

*From 1st October 2020:* Enti Spata (NDPH, University of Oxford)

Role: To develop the statistical analysis plan (blinded to trial allocation) and conduct the final comparative analyses for all other treatment arms.

##### *Data Monitoring Committee (DMC) Statisticians*

Professor Jonathan Emberson and Dr Natalie Staplin (NDPH, University of Oxford)

Role: To conduct regular interim analyses for the DMC. Contribution restricted up until unblinded to trial allocation.

##### *Statisticians on the Trial Steering Committee (TSC)*

Professor Edmund Juszczak (University of Nottingham), Professor Alan Montgomery (University of Nottingham), and Professor Thomas Jaki (University of Cambridge)

Role: Major organisational and policy decisions, and scientific advice; blinded to treatment allocation.

##### *Trial IT systems & Programmers*

Andy King, David Murray, Richard Welsh (NDPH, University of Oxford)

Role: To generate and prepare reports monitoring the randomisation schedule. To supply data snapshots for interim and final analysis. Responsibility for randomisation system, clinical databases and related activities.

Bob Goodenough (NDPH, University of Oxford)

Role: Validation of IT systems

Dr Will Stevens, Karl Wallendszus (NDPH, University of Oxford)

Role: To produce analysis-ready datasets according to CDISC standards.

### 1 INTRODUCTION

This document details the proposed presentation and analysis for the main paper(s) reporting results from the multicentre randomised controlled trial RECOVERY (ISRCTN50189673) to investigate multiple treatments on major outcomes in inpatients for COVID-19 (clinically suspected or laboratory confirmed).

The results reported in these papers will follow the strategy set out here, which adheres to the guidelines for the content of a statistical analysis plan (SAP).[1] Any subsequent analyses of a more exploratory nature will not be bound by this strategy.

Suggestions for subsequent analyses by oversight committees, journal editors or referees, will be considered carefully in line with the principles of this analysis plan.

Any deviations from the statistical analysis plan will be described and justified in the final report. The analysis will be carried out by identified, appropriately qualified and experienced statisticians, who will ensure the integrity of the data during their processing.

This SAP is based on multiple versions of the protocol. All regulatory documents can be found in the RECOVERY trial directory: https://www.recoverytrial.net/for-site-staff/site-set-up-1/regulatory-documents.

SAP versions 1.0 & 1.1 applied to the first three principal comparisons (hydroxychloroquine, dexamethasone, and lopinavir-ritonavir versus no additional treatment respectively), for which data matured in the first UK wave of the pandemic. However, due to its later introduction, enrolment of patients in the azithromycin arm was much slower. Over time, factorial randomisations and a second randomisation have been added, introducing new treatment arms including convalescent plasma, tocilizumab, synthetic neutralizing antibodies, and aspirin. Version 2.0 of the SAP was produced in response to these changes, combined with the fact that use of corticosteroids (one of the original treatment arms) is now the usual standard of care for many patients.

SAP version 3.0 now includes the following revisions:

- **REGN-COV2:** Specification of analysis method (see appendix).
- **Early phase assessments:** Additional analyses for treatments undergoing early phase assessment (introduced in protocol version 14.0; modified in SAP version 3.1); see section 9.
- **6 month follow-up:** Analyses based on information available up to 6 months after randomisation (modified in SAP version 3.1); see section 10.

The primary outcome for children will be the duration of hospitalisation (and death is an extremely rare event). The analyses of data from children will be specified in a separate Statistical Analysis Plan.

### 2 BACKGROUND INFORMATION

#### 2.1 Rationale

In early 2020, as the protocol was being developed, there were no approved treatments for COVID-19. The aim of the trial is to provide reliable evidence on the efficacy of candidate therapies (including re-purposed and novel drugs) for suspected or confirmed COVID-19 infection on major outcomes in hospitalised adult patients receiving standard care.

#### 2.2 Objectives of the trial

##### 2.2.1 *Primary objective*

To provide reliable estimates of the effect of study treatments on all-cause mortality within 28 days of the relevant randomisation.

##### 2.2.2 *Secondary objectives*

To investigate the effect of study treatments on the duration of hospital stay and on the combined endpoint of use of invasive mechanical ventilation (including Extra Corporal Membrane Oxygenation [ECMO]) or death.

#### 2.3 Trial design

This is a multi-centre, multi-arm, adaptive, open label, randomised controlled trial with three possible stages of randomisation, as described below. The trial is designed with streamlined processes in order to facilitate rapid large-scale recruitment with minimal data collection.

#### 2.4 Eligibility

##### 2.4.1 *Inclusion criteria*

Patients are eligible for the trial if all of the following are true:

- Hospitalised
- SARS-Cov-2 infection (clinically suspected or laboratory confirmed)
- No medical history that might, in the opinion of the attending clinician, put the patient at significant risk if they were to participate in the trial.

##### 2.4.2 *Exclusion criteria*

If one or more of the active drug treatments is not available at the hospital or is believed, by the attending clinician, to be contraindicated (or definitely indicated) for the specific patient, then this fact will be recorded via the web-based form prior to randomisation; random allocation will then be between the remaining arms.

#### 2.5 Treatments

All patients will receive standard management for the participating hospital. The main randomisation will be between the following treatment arms (although not all arms may be available at any one time). The doses listed are for adults; paediatric dosing is described in the protocol.

##### 2.5.1 *Main randomisation part A:*

- **No additional treatment**
- **Lopinavir 400mg-Ritonavir 100mg** by mouth (or nasogastric tube) every 12 hours for 10 days. [Introduced in protocol version 1.0; **enrolment closed** 29 June 2020]
- **Corticosteroid** in the form of dexamethasone**,** administered as an oral liquid or intravenous preparation 6 mg once daily for 10 days. In pregnancy, prednisolone 40 mg administered by mouth (or intravenous hydrocortisone 80 mg twice daily) should be used instead. [Introduced in protocol version 1.0; **enrolment closed to adults** 8 June 2020]
- **Hydroxychloroquine** by mouth for 10 days (4 doses in first 24 hours and 1 dose every 12 hours for 9 days). [Introduced in protocol version 2.0; **enrolment closed** 5 June 2020]
- **Azithromycin 500mg** by mouth (or nasogastric tube) or intravenously once daily for a total of 10 days. [Introduced in protocol version 3.0; **enrolment closed** 27 November 2020]
- **Colchicine** by mouth for 10 days (1.5 mg in first 12 hours then 0.5 mg twice daily). [Introduced in protocol version 12.0; **enrolment closed** 5 March 2021.]
- **Dimethyl fumarate** 120 mg every 12 hours for 4 doses followed by 240 mg every 12 hours by mouth for 8 days (10 days in total). [Introduced in protocol version 14.0; enrolment ongoing.] **Undergoing Early Phase Assessment**

##### 2.5.2 *Main randomisation part B:*

In a factorial design, eligible patients may be randomised to the arms below. The doses listed are for adults; paediatric dosing is described in the protocol.

- **No additional treatment**
- **Convalescent plasma** Single unit of ABO compatible convalescent plasma (275mls ± 75 mls) intravenous per day on study days 1 (as soon as possible after randomisation) and 2 (with a minimum of 12-hour interval between 1st and 2nd units). ABO identical plasma is preferred if available. The second transfusion should not be given if patient has a suspected serious adverse reaction during or after the first transfusion. [Introduced in protocol version 6.0; **enrolment closed** 15 January 2021]
- **Synthetic neutralising antibodies** (REGN-COV2; adults and children aged ≥12 years only - children who weigh <40kg will also not be eligible for this treatment). A single dose of REGN10933 + REGN10987 8 g (4 g of each monoclonal antibody) in 250ml 0.9% saline infused intravenously over 60 minutes ± 15 minutes as soon as possible after randomisation. [Introduced in protocol version 9.1; **enrolment closed** 22 May 2021]

##### 2.5.3 *Main randomisation part C:*

In a factorial design, eligible patients may be randomised to the arms below. The dose listed is for adults; children are excluded from this comparison.

- **No additional treatment**
- **Aspirin** 150 mg by mouth (or nasogastric tube) or per rectum once daily until discharge. [Introduced in protocol version 10.1; **enrolment closed** 21 March 2021]

##### 2.5.4 *Main randomisation part D:*

In a factorial design, eligible patients may be randomised to the arms below. The dose listed is for adults; children <2 years old or with PIMS-TS are excluded from this comparison.

- **No additional treatment**
- **Baricitinib** 4 mg by mouth (or nasogastric tube) once daily for 10 days. [Introduced in protocol version 13.0; **enrolment ongoing**]

##### 2.5.5 *Main randomisation part E:*

In a factorial design, eligible patients may be randomised to the arms below. The dose listed is for adults; children <18 years old are excluded from this comparison.

- **No additional treatment**
- **High-dose corticosteroids** dexamethasone 20 mg once daily for 5 days, followed by dexamethasone 10 mg once daily for 5 days. [Introduced in protocol version 13.0; **enrolment ongoing**]

##### 2.5.6 *Main randomisation part F:*

In a factorial design, eligible patients may be randomised to the arms below. The dose listed is for adults; children <18 years old are excluded from this comparison.

- **No additional treatment**
- **Empagliflozin** 10 mg once daily for 28 days. [Introduced in protocol version 16.1; **enrolment ongoing**]

##### 2.5.7 *Second randomisation for adults with progressive COVID-19*

Patients enrolled in the main RECOVERY trial and with clinical evidence of a hyper-inflammatory state may be considered for a second randomisation if they meet the following criteria:

- Randomised into the main RECOVERY trial no more than 21 days ago
- Clinical evidence of progressive COVID-19:
  - oxygen saturation <92% on room air or requiring oxygen; and

- C-reactive protein (CRP) ≥75 mg/L

- No medical history that might, in the opinion of the attending clinician, put the patient at significant risk if they were to participate in this aspect of the RECOVERY trial

Eligible participants may be randomised between the following treatment arms:

- **No additional treatment**
- **Tocilizumab** by intravenous infusion with the dose determined by body weight. [Introduced in protocol version 4.0; **enrolment closed** 24 January 2021]

#### 2.6 Definitions of primary and secondary outcomes

Outcomes will be assessed at 28 days and then 6 months after the relevant randomisation. Analysis of longer-term outcomes collected beyond this will be described in a separate Statistical Analysis Plan.

##### 2.6.1 *Primary outcome*

Mortality (all-cause)

##### 2.6.2 *Secondary clinical outcomes*

- Time to discharge from hospital
- Use of invasive mechanical ventilation (including Extra Corporal Membrane Oxygenation [ECMO]) or death (among patients not on invasive mechanical ventilation or ECMO at time of randomisation)

##### 2.6.3 *Subsidiary clinical outcomes*

- Use of ventilation (overall and by type) among patients not on ventilation (of any type) at time of randomisation
- Duration of invasive mechanical ventilation among patients on invasive mechanical ventilation at time of randomisation (defined as time to successful cessation of invasive mechanical ventilation: see section 5.1.7)
- Use of renal dialysis or haemofiltration (among patients not on renal dialysis or haemofiltration at time of randomisation)
- Thrombotic events (overall and by type; introduced in Protocol version 10.1)

##### 2.6.4 *Safety outcomes*

- Cause-specific mortality (COVID-19, other infection, cardiac, stroke, other vascular, cancer, other medical, external, unknown cause)
- Major cardiac arrhythmia (recorded on follow-up forms completed from 12 May 2020 onwards)
- Major bleeding (overall and by type; introduced in Protocol version 10.1)
- Early safety of antibody-based therapy (sudden worsening in respiratory status; severe allergic reaction; temperature >39°C or ≥2°C rise since randomisation; sudden

hypotension; clinical haemolysis; and thrombotic events within the first 72 hours; Main randomization phase B only)

- Non-coronavirus infection (overall and by site and putative organism [virus, bacteria, fungus, other]; introduced in Protocol version 14.0)

###### 2.6.5 *Detailed derivation of outcomes*

The detailed derivation of outcomes included in statistical analysis will be described separately in a data derivation document and included in the Study Data Reviewer's Guide.

##### 2.7 Hypothesis framework

For each of the primary, secondary and subsidiary outcomes, the null hypothesis will be that there is no true difference in effect between any of the treatment arms.

##### 2.8 Sample size

The larger the number randomised, the more accurate the results will be, but the numbers that can be randomised will depend critically on how large the epidemic becomes. If substantial numbers are hospitalised in the participating centres then it may be possible to randomise several thousand with moderate disease and a few thousand with severe disease. Some indicative sample sizes and projected recruitment will be estimated using emerging data for several different scenarios. Sample size and recruitment will be monitored by the TSC throughout the trial.

##### 2.9 Randomisation

Eligible patients will be randomised using a 24/7 secure central web-based randomisation system, developed and hosted within NDPH, University of Oxford. Users of the system will have no insight into the next allocation, given that simple randomisation is being used. If a patient is randomised inadvertently more than once during the same hospital admission, the first allocation will be used.

The implementation of the randomisation procedure will be monitored by the Senior Trials Programmer, and the TSC notified if an error in the randomisation process is identified.

###### 2.9.1 *Main randomisation part A*

Simple randomisation will be used to allocate participants to one of the following treatment arms (in addition to usual care), which is subject to change:

- No additional treatment
- Lopinavir-Ritonavir [Introduced in protocol version 1.0; **enrolment closed** 29 June 2020]
- Corticosteroid [Introduced in protocol version 1.0; **enrolment closed to adults** 8 June 2020]
- Hydroxychloroquine [Introduced in protocol version 2.0; **enrolment closed** 5 June 2020]
- Azithromycin [Introduced in protocol version 3.0; **enrolment closed** 27 November 2020]
- Colchicine [Introduced in protocol version 11.1; **enrolment closed** 5 March 2021]

- Dimethyl fumarate [Introduced in protocol version 14.0; **enrolment ongoing**]

The randomisation programme will allocated patients in a ratio of 2:1 between the no additional treatment arm and each of the other arms that are not contra-indicated and are available when multiple arms were included in the protocol. Hence if all 4 active treatment arms are available, then the randomisation will be in the ratio 2:1:1:1:1. If one or more of the active drug treatments is not available at the hospital or is believed, by the attending clinician, to be contraindicated (or definitely indicated) for the specific patient, then this fact will be recorded via the web-based form prior to randomisation; random allocation will then be between the remaining arms (in a 2:1:1:1, 2:1:1 or 2:1 ratio). Since the closure of the azithromycin comparison, all comparisons in part A have used a 1:1 ratio.

##### 2.9.2 *Main randomisation part B*

In a factorial design, eligible patients will be randomised simultaneously using simple randomisation with allocation ratio 1:1:1 to one of the following arms, which is subject to change:

- No additional treatment
- Convalescent plasma [Introduced in protocol version 6.0; **enrolment closed** 15 January 2021]
- Synthetic neutralising antibodies [Introduced in protocol version 9.1; **enrolment closed** 22 May 2021]

If the active treatment is not available at the hospital, the patient does not consent to receive convalescent plasma, or is believed, by the attending clinician, to be contraindicated for the specific patient, then this fact will be recorded via the web-based form and the patient will be excluded from the relevant arm in Randomisation part B.

##### 2.9.3 *Main randomisation part C*

In a factorial design, eligible patients will be randomised simultaneously using simple randomisation with allocation ratio 1:1 to one of the following arms, which is subject to change:

- No additional treatment
- Aspirin [Introduced in protocol version 10.1; **enrolment closed** 21 March 2021]

##### 2.9.4 *Main randomisation part D*

In a factorial design, eligible patients will be randomised simultaneously using simple randomisation with allocation ratio 1:1 to one of the following arms, which is subject to change:

- No additional treatment
- Baricitinib [Introduced in protocol version 13.0; **enrolment ongoing**]

##### 2.9.5 *Main randomisation part E*

In a factorial design, eligible patients will be randomised simultaneously using simple randomisation with allocation ratio 1:1 to one of the following arms, which is subject to change:

- No additional treatment
- High-dose corticosteroids [Introduced in protocol version 15.0; **enrolment ongoing**]

##### 2.9.6 *Main randomisation part F*

In a factorial design, eligible patients will be randomised simultaneously using simple randomisation with allocation ratio 1:1 to one of the following arms, which is subject to change:

- No additional treatment
- Empagliflozin [Introduced in protocol version 16.1; **enrolment ongoing**]

Note: From protocol version 7.0 onwards, randomisation is permitted in part B of main randomisation without randomisation in part A. From protocol version 10.1 onwards, randomisation is permitted in any combination of parts A to F.

##### 2.9.7 *Second randomisation for adults with progressive COVID-19*

Eligible participants will be randomised using simple randomisation with an allocation ratio 1:1 between the following arms, which is subject to change:

- No additional treatment
- Tocilizumab [Introduced in protocol version 4.0; **enrolment closed** 24 January 2021]

#### 2.10 Blinding

This is an open-label study. However, while the study is in progress, access to tabular results of study outcomes by treatment allocation will not be available to the research team, CIs, trial statisticians, clinical teams, or members of the TSC (unless the DMC advises otherwise). The DMC and DMC statisticians will be unblinded.

#### 2.11 Data collection schedule

Baseline and outcome information will be collected on trial-specific electronic case report forms (eCRFs) and entered into a web-based IT system by a member of the hospital or research staff. Follow-up information will be collected on all study participants, irrespective of whether they complete the scheduled course of allocated study treatment. Study staff will seek follow-up information through various means, including routine healthcare systems and registries.

All randomised participants will be followed up until death or 6 months post-randomisation to the main trial (whichever is sooner). NHS Digital and equivalent organisations in the devolved nations will supply data fields relevant to trial baseline and outcome measures to NDPH, University of Oxford on a regular basis, for participants enrolled into the trial. This will be combined with the trial-specific data collected via the web-based IT system and adjudicated internally.

Longer term (up to 10 years) follow-up will be sought through linkage to electronic healthcare records and medical databases including those held by NHS Digital, Public Health England and equivalent bodies, and to relevant research databases (e.g. UK Biobank, Genomics England).

##### 2.12 Data monitoring

During the study all study data will be supplied in strict confidence to the independent DMC for independent assessment and evaluation. The DMC will request such analyses at a frequency relevant to the emerging data from this and other studies.

The DMC has been requested to determine if, in their view, the randomised comparisons in the study have provided evidence on mortality that is strong enough (with a range of uncertainty around the results that is narrow enough) to affect national and global treatment strategies. Hence, multiple reviews by the Data Monitoring Committee have no material impact on the final analysis. In such a circumstance, the DMC will inform the TSC who will make the results available to the public and amend the trial arms accordingly.

##### 2.13 Trial reporting

The trial will be reported according to the principles of the CONSORT statements.[2, 3, 4] The exact composition of the trial publication(s) depends on the size of the epidemic, the availability of drugs, and the findings from the various pairwise comparative analyses (with the no additional treatment arm) in the main trial.

#### 3 ANALYSIS POPULATIONS

##### 3.1 Population definitions

The intention to treat (ITT) population will be all participants randomised, irrespective of treatment received. This ITT population will be used for analysis of efficacy and safety data.
For interim analyses, baseline data will be reported for all participants with data available and outcome data will be reported for all participants who have died, been discharged from hospital, or reached day 28 after the first randomisation.

#### 4 DESCRIPTIVE ANALYSES

##### 4.1 Participant throughput

The flow of participants through the trial will be summarised for each separate pairwise comparison using a CONSORT diagram. The flow diagram will show the contribution of participants from each of the paths (from each of the parts of the main randomisation and from the second randomisation), where applicable. The flow diagrams will describe the numbers of participants randomly allocated, who received allocation, withdrew consent, and included in the ITT analysis population. The flow diagrams for arms in the main randomisation will also report the number of participants who underwent the second randomisation.

##### 4.2 Baseline comparability of randomised groups

The following characteristics will be described separately for patients randomised to each main comparison (for each separate pairwise comparison of active treatment with the no additional treatment arm), and separately for the first and second randomisation.

###### 4.2.1 Main randomisation (parts A, B and C)

- Age at randomisation
- Sex
- Ethnicity
- Region (UK, non-UK)
- Time since COVID-19 symptoms onset
- Time since hospitalisation
- Current respiratory support
- Comorbidities (diabetes, heart disease, chronic lung disease, tuberculosis, human immunodeficiency virus, severe liver disease, severe kidney impairment)
- SARS-Cov-2 test result
- If female, known to be pregnant
- Use of systemic corticosteroid (including those allocated to corticosteroid in part A)
- Use of other relevant treatments (e.g. remdesivir, interleukin-6 antagonist, monoclonal anti-SARS-CoV-2 neutralising antibody)
- For part B only, anti-SARS-CoV-2 antibody concentration
- For treatment comparisons introduced in protocol v9.1 onwards:
    - C-reactive protein
    - Estimated glomerular filtration rate (calculated using the CKD-EPI formula)
    - D-dimer

###### 4.2.2 Second randomisation

In addition to the above:

- Current respiratory support
- Latest oxygen saturation measurement
- Latest C-reactive protein
- Latest ferritin
- Latest estimated glomerular filtration rate (calculated using the CKD-EPI formula)
- Allocation in main randomisation parts A, B, C, D and E
- Interval between first and second randomisation

The number and percentage will be presented for binary and categorical variables. The mean and standard deviation or the median and the interquartile range will be presented for continuous variables.

##### 4.3 Completeness of follow-up

All reasonable efforts will be taken to minimise loss to follow-up, which is expected to be minimal as data collection for primary and secondary outcomes using trial-specific eCRFs is combined with linkage to routine clinical data on study outcomes from NHS Digital, ICNARC, and similar organisations in the devolved nations.

The number and percentage of participants with follow-up information at day 28 and at 6 months after the relevant randomisation will be reported. Data will be shown for each of the following: all-cause mortality, hospital discharge status, ventilation status, and will be shown for each randomised group for the main and second randomisation separately.

#### 4.4 Adherence to treatment

The number and proportion of patients who did not receive the treatment they were allocated to will be reported. If any other trial treatment options were known to be received, instead of or in addition to, the allocated treatment during the 28-day follow-up period after the first randomisation, these will be collected and reported. Details on the number of days (or doses) of treatment received will be reported for all trial treatments received where available.

### 5 COMPARATIVE ANALYSES

For all outcomes, the primary analysis will be performed on the intention to treat (ITT) population at 28 days after randomisation. (Additional details specific to the comparison of REGN-COV2 vs. usual care are provided in Appendix A.An ITT analysis of all outcomes at 6 months post-randomisation will also be conducted.

Pairwise comparisons will be made between each treatment arm and the no additional treatment arm (reference group) in that particular randomisation (main randomisation part A, main randomisation part B, main randomisation part C, main randomisation part D, main randomisation part E and second randomisation). Since not all treatments may be available or suitable for all patients, those in the no additional treatment arm will only be included in a given comparison if, at the point of their randomisation, they *could* alternatively have been randomised to the active treatment of interest (i.e. the active treatment was available at the time and it was not contra-indicated). The same applies to treatment arms added at a later stage; they will only be compared to those patients recruited concurrently.

#### 5.1 Main randomisation part A

##### 5.1.1 *Primary outcome*

Mortality (all-cause) will be summarised with counts and percentages by randomised comparison group. A time-to-event analysis will be conducted using the log-rank test, with the p-value reported. Kaplan-Meier estimates for the time to event will also be plotted (with associated log-rank p-values). The log-rank 'observed minus expected' statistic (and its variance) will be used to calculate the one-step estimate of the event rate ratio and confidence interval for each treatment group versus the no additional treatment group.[5] For the primary outcome, discharge alive before the relevant time period (28 days after randomisation) will be assumed as absence of the event (unless there is additional data confirming otherwise).

##### 5.1.2 *Secondary outcomes*

##### 5.1.3 *Time to discharge alive from hospital*

A time-to-event analysis will be used to compare each treatment group with the no additional treatment group using the log-rank test. As described for the primary outcome, the rate ratio and its confidence interval will be estimated from the log-rank observed minus expected statistic and its variance, and Kaplan-Meier curves will be drawn. Patients who die in hospital will be censored after 28 days after randomisation. This gives an unbiased estimate of the recovery rate and comparable estimates to the competing risks approach in the absence of other censoring (which is expected to be very minimal).[6]

##### 5.1.4 *Use of invasive mechanical ventilation (including ECMO) or death*

Counts and percentages will be presented by randomised group and the risk ratio will be calculated for each pairwise comparison with the no additional treatment arm, with confidence intervals and p-values reported. The absolute risk difference will also be presented with confidence intervals. Each component of this composite outcome will also be summarised. Patients who were already on invasive mechanical ventilation or ECMO at randomisation will be excluded from these analyses.

##### 5.1.5 *Subsidiary clinical outcomes*

##### 5.1.6 *Use of ventilation (overall and by type)*

Counts and percentages will be presented by randomised group for patients who received any assisted ventilation, together with risk ratios and confidence intervals for each pairwise comparison with the no additional treatment arm. The number of patients receiving the two main types of ventilation will also be reported: non-invasive ventilation (including CPAP, other non-invasive ventilation or high-flow nasal oxygen), and invasive mechanical ventilation (including ECMO). Patients who were already receiving ventilation[a] at randomisation will be excluded from these analyses.

##### 5.1.7 *Duration of invasive mechanical ventilation (time to successful cessation of invasive mechanical ventilation)*

Successful cessation of invasive mechanical ventilation will be defined as removal of invasive mechanical ventilation within (and survival to) 28 days after randomisation. A time-to-event analysis will be used to compare each treatment group with the no additional treatment group using the log-rank test, as described above. The rate ratio and its confidence interval will be estimated from the log-rank observed minus expected statistic and its variance, and Kaplan-Meier curves will be drawn. Patients who die within 28 days of randomisation will be censored *after* 28 days after randomisation. Patients who were not already on invasive mechanical ventilation or ECMO at randomisation will be excluded from these analyses.

##### 5.1.8 *Use of renal dialysis or haemofiltration*

Counts and percentages will be presented by randomised group and the risk ratio will be calculated for each pairwise comparison with the no additional treatment arm, with

[a] Participants recruited to the main randomisation prior to protocol version 9.1 who were already receiving oxygen at randomisation will also be excluded from these analyses (since it is not possible to distinguish those who were already receiving non-invasive ventilation).

confidence intervals and p-values reported. The absolute risk difference will also be presented with confidence intervals. Patients who were already on renal dialysis or haemofiltration at randomisation will be excluded from these analyses.

##### 5.1.9 *Thrombotic event*

Counts and percentages will be presented by randomised group. The absolute risk differences will also be presented with confidence intervals. Type of thrombotic event will also be described: (i) acute pulmonary embolism; (ii) deep vein thrombosis; (iii) ischaemic stroke, (iv) myocardial infarction; (v) systemic arterial embolism; and (vi) all sites combined.

#### 5.2 Main randomisation part B

In the factorial design, the main effects of treatments evaluated in part B will be presented and tested across all arms in main randomisation parts A, C, D, E and F combined, as described in 5.1. (Assessments of whether the effects of treatments in part B vary depending on other randomised treatments are described in section 5.10).

#### 5.3 Main randomisation part C

In the factorial design, the main effects of treatments evaluated in part C will be presented and tested across all arms in main randomisation parts A, B, D, E and F combined, as described in 5.1. (Assessments of whether the effects of treatments in part C vary depending on other randomised treatments are described in section 5.10).

#### 5.4 Main randomisation part D

In the factorial design, the main effects of treatments evaluated in part D will be presented and tested across all arms in main randomisation parts A, B, C, E and F combined, as described in 5.1. (Assessments of whether the effects of treatments in part D vary depending on other randomised treatments are described in section 5.10).

#### 5.5 Main randomisation part E

In the factorial design, the main effects of treatments evaluated in part E will be presented and tested across all arms in main randomisation parts A, B, C, D and F combined, as described in 5.1. (Assessments of whether the effects of treatments in part E vary depending on other randomised treatments are described in section 5.10).

#### 5.6 Main randomisation part F

In the factorial design, the main effects of treatments evaluated in part F will be presented and tested across all arms in main randomisation parts A to E combined, as described in 5.1. (Assessments of whether the effects of treatments in part F vary depending on other randomised treatments are described in section 5.10).

#### 5.7 Second randomisation

Evaluation of treatment effects in the main randomisation and the second randomisation will be conducted independently, as described in 5.1.

#### 5.8 Pre-specified subgroup analyses

Pre-specified subgroup analyses will be conducted for the main randomisation (parts A, B, C, D, E and F) and the second randomisation, for the following outcomes:

- Mortality (all-cause)
- Time to discharge from hospital
- Use of invasive mechanical ventilation (including ECMO) or death

Tests for heterogeneity (or tests for trend for 3 or more ordered groups) will be conducted to assess whether there is any good evidence that the effects in particular subgroups differ materially from the overall effect seen in all patients combined. Results will be presented on forest plots as event rate ratios, or risk ratios, with confidence intervals. The following subgroups will be examined based on information at randomisation:

- Age (<70; 70-79; 80+ years)
- Sex (Male; Female)
- Ethnicity (White; Black, Asian or Minority Ethnic)
- Region (UK, non-UK)
- Time since illness onset (≤7 days; >7 days)
- Requirement for respiratory support
    - For main randomisation: None; Oxygen only; Non-invasive ventilation; Invasive mechanical ventilation (including ECMO)[b]
    - For second randomisation: No ventilator support (including no or low-flow oxygen); Non-invasive ventilation (including CPAP, other non-invasive ventilation, or high-flow nasal oxygen), Invasive mechanical ventilation (including ECMO)
- Use of systemic corticosteroid (including dexamethasone)
- For part B only: Recipient anti-SARS-CoV-2 antibody concentration at randomisation ($<8 \times 10^6$ units; $\geq 8 \times 10^6$ units[c]). (This will be the key subgroup for the REGN-COV2 comparison.)

#### 5.9 Sensitivity analyses

Sensitivity analyses of the primary and secondary outcomes will be conducted among those patients with a positive test for SARS-COV-2 (i.e. confirmed cases).

#### 5.10 Other exploratory analyses

In addition, exploratory analyses will be conducted to test for interactions between treatments allocated in each of the different randomisations, provided that doing so does not lead to premature unblinding of results for ongoing comparators.

[b] Participants recruited before protocol V9.1 who were receiving oxygen would be presented in a fifth subgroup but not included in the test for trend

Non-randomised exploratory analyses will be used to explore the likely influence of different levels of convalescent plasma antibody concentrations on the efficacy of convalescent plasma.

Additional analyses will set the results for children (<18 years) and pregnant women in the context of the overall results.

#### 5.11 Adjustment for baseline characteristics

The main analyses described above will be unadjusted for baseline characteristics. However, if there are any important imbalances between the randomised groups in key baseline pre-specified subgroups (see section 5.4) or allocation in the orthogonal components of the main randomisation, where applicable, emphasis will be placed on analyses that are adjusted for the relevant baseline characteristic(s). This will be done using Cox regression for the estimation of adjusted hazard ratios and a log-binomial regression model for the estimation of adjusted risk ratios.

#### 5.12 Significance levels and adjustment of p-values for multiplicity

Evaluation of the primary trial (main randomisation) and secondary randomisation will be conducted independently, and no adjustment be made for these. Formal adjustment will not be made for multiple treatment comparisons, the testing of secondary and subsidiary outcomes, or subgroup analyses (with one exception; see Appendix A). However, due allowance for multiple testing will be made in the interpretation of the results: the larger the number of events on which a comparison is based and the more extreme the P-value after any allowance has been made for the nature of the particular comparison (i.e. primary or secondary; pre-specified or exploratory), the more reliable the comparison and, hence, the more definite any finding will be considered. 95% confidence intervals will be presented for the main comparisons.

#### 5.13 Statistical software employed

The statistical software SAS version 9.4 and R Studio 3.6.2 (or later) for Windows will be used for the interim and final analyses.

#### 5.14 Data standards and coding terminology

Datasets for analysis will be prepared using CDISC standards for SDTM and ADaM. Wherever possible, clinical outcomes (which may be obtained in a variety of standards, including ICD10 and OPCS-4) will be coded using MedDRA version 20.1.

### 6 SAFETY DATA

Suspected serious adverse reactions (SSARs) and suspected unexpected serious adverse reactions (SUSARs) will be listed by trial allocation.

For each of the following, counts and percentages will be presented by randomised group. Where possible, the absolute risk differences will also be presented with confidence intervals:

#### 6.1 Cause-specific mortality

Cause-specific mortality (COVID-19, other infection, cardiac, stroke, other vascular, cancer, other medical, external, unknown cause) will be analysed in a similar manner to the primary outcome.

#### 6.2 Major cardiac arrhythmia

Type of arrhythmia will also be described: (i) atrial flutter or fibrillation; (ii) supraventricular tachycardia; (iii) ventricular tachycardia; (iv) ventricular fibrillation; (v) atrioventricular block requiring intervention, with subtotals for (i)-(ii) and (iii)-(iv).

#### 6.3 Major bleeding

Type of bleeding will also be described: (i) intracranial bleeding; (ii) gastro-intestinal bleeding; (iii) other bleeding site, and (iv) all sites combined.

#### 6.4 Early safety of anti-coronavirus antibody-based therapy

Additional safety data will be collected in a subset of patients randomised to part B: (i) sudden worsening in respiratory status; (ii) severe allergic reaction; (iii) temperature >39°C or ≥2°C rise since randomisation; (iv) sudden hypotension; (v) clinical haemolysis; and (vi) thrombotic event.

#### 6.5 Other infections

Other infections occurring after randomisation will be described. These will be classified primarily by site (pneumonia, urinary tract, biliary, other intra-abdominal, blood stream, skin, other). Information on putative organism (other virus, bacterial, fungal, other and unknown) is also collected.

#### 6.6 Metabolic complications

Incidence of the following metabolic complications after randomisation will be described:

- Ketoacidosis (defined as combination of ketosis [blood ketones ≥1.5 mmol/L or urine ketones ≥2+] and acidosis [venous bicarbonate <15 mmol/L)
- Hyperglycaemic hyperosmolar state
- Other hyperglycaemia requiring new use of insulin
- Severe hypoglycaemia (causing reduced conscious level requiring another person to help recover)

### 7 ADDITIONAL POST-HOC EXPLORATORY ANALYSIS

Any post-hoc analysis requested by the oversight committees, a journal editor or referees will be labelled explicitly as such. Any further future analyses not specified in the analysis protocol will be exploratory in nature and will be documented in a separate statistical analysis plan.

### 8 DIFFERENCES FROM PROTOCOL

The testing of multiple treatment arms will not formally be adjusted for, but given the number of comparisons, due allowance will be made in their interpretation. Formal methods of adjustment for multiplicity were not adopted because of treatment arms being added over time (including the factorial convalescent plasma comparison), unequal recruitment into each arm, and the ultimate number of treatments under evaluation not being known in advance.

This analysis plan will be updated prior to unblinding of the 6-month follow-up results. Additional analyses may be specified, e.g. to explore the impact of randomised treatment allocation on hospital re-admission for COVID-19.

### 9 EARLY PHASE ASSESSMENTS

The following approach is required for the evaluation of treatments indicated as undergoing Early Phase Assessment in the protocol (introduced in Protocol version 14.0):

#### 9.1 Definitions of clinical outcomes

##### 9.1.1 *Primary outcome*

- WHO ordinal scale on day 5

##### 9.1.2 *Secondary clinical outcomes*

- Time to sustained improvement (i.e., value better than baseline value persisting for >1 day) by at least one category on the WHO ordinal scale from baseline
- $S/F_{94}$ ratio at day 5
- Time to discharge from hospital
- Improvement in clinical status at day 10
- Blood C-reactive protein at day 5

##### 9.1.3 *Subsidiary clinical outcomes*

- All other subsidiary outcomes as described above (section 2.6.3)

##### 9.1.4 *Safety outcomes*

- Flushing (incidence, severity)
- Gastrointestinal symptoms (incidence, severity)
- Reasons for stopping study treatment
- Transaminitis (ALT >3x upper limit of normal)
- Acute kidney injury (creatinine >1.5x value entered at randomisation)
- All other subsidiary outcomes as described above (section 2.6.4)

#### 9.2 Baseline comparability of randomised groups

Unless otherwise specified, analyses will follow the plan described above (section 4). In addition, the following characteristics will be described:

- Oxygen saturation measurement on air (if available)
- $S/F_{94}$ ratio
- WHO Ordinal Scale
- All other characteristics as described above (section 4.2)

#### 9.3 Comparative analysis

Unless otherwise specified, comparative analyses will follow the plan described above (section 5). In addition,

##### 9.3.1 *Primary outcome*

The primary comparison will involve an "intention to treat" analysis among all participants randomised between the active arm and its control of the effect of the active treatment on WHO scale at day 5, adjusted for baseline score. A proportional odds model will be used to assess the common odds ratio of better outcome for each pairwise comparison with the no additional treatment arm.[8] In addition, a sensitivity analysis to the proportional odds model using Howard's method will be performed if the proportional odds assumption is not satisfied.[9]

##### 9.3.2 *Secondary outcomes*

###### *9.3.2.1 Time to sustained improvement by at least one category on the WHO ordinal scale from baseline*

A time-to-event analysis will be used to compare each treatment group with the no additional treatment group using the log-rank test (restricted to the first 10 days of the trial as the WHO score is not collected after this). The rate ratio and its confidence interval will be estimated from the log-rank observed minus expected statistic and its variance, and Kaplan-Meier curves will be drawn.

###### *9.3.2.2 Improvement in clinical status at day 10*

Counts and percentages will be presented by randomised group for patients with an improvement of at least one category on the WHO ordinal scale from baseline, together with odds ratios and confidence intervals for each pairwise comparison with the no additional treatment arm.

###### *9.3.2.3 Blood C-reactive protein at day 5*

Geometric mean C-reactive protein at day 5 will be compared between treatment arms. Estimates will be obtained from analysis of covariance (ANCOVA) for the log transformed CRP values after adjustment for each participant's baseline value. Approximate standard errors for the geometric means will be calculated from the confidence intervals. Missing CRP values will be handled as described in section 9.3.2.5.

###### *9.3.2.4 $S/F_{94}$ ratio at day 5*

Mean $S/F_{94}$ ratio at day 5 will be compared between treatment arms. Estimates will be obtained from analysis of covariance (ANCOVA) after adjustment for each participant's baseline $S/F_{94}$ ratio. Missing $S/F_{94}$ ratio values will be handled as described in section 9.3.2.5

###### *9.3.2.5 Imputation of missing data*

All analyses will be done according to the intention-to-treat principle and, hence, missing secondary outcome data will be imputed. For each of the continuous outcomes (e.g., CRP, $S/F_{94}$ ratio) missing post-randomisation results will be imputed using multiple imputation, using 20 imputed data sets, with results across imputations being combined using the methods of Rubin.[10] The imputation procedure will take into consideration each participant's key baseline characteristics (listed in section 5.8), treatment allocation and any intermediate follow-up values of the biomarker, where available. For $S/F_{94}$ ratio, WHO ordinal scale values on days 3 and 5 will also be used in the imputation procedure. For patients who are discharged from hospital and for whom it is not possible to measure $S/F_{94}$ ratio at day 5, a value of 4.76[d] will be imputed. The results from these analyses will be compared with those from equivalent "complete-case" analyses, but primary emphasis will be placed on the results after multiple imputation. All multiple imputation analyses will be implemented using the multiple imputation procedure in SAS version 9.4 (SAS Institute, Cary NC), using the expectation-maximization algorithm (which assumes a multivariate normal distribution) to impute values.

[d] 4.76 = 1.0/0.21 (ie, the value of healthy lungs which provide 100% saturations when breathing 21% oxygen)

For any continuous variables with missing baseline values, the mean among those with observed values will be imputed.

###### 9.3.3 *Safety outcomes*

Counts and percentages will be presented by randomised group. The absolute risk differences will also be presented with confidence intervals for each of the following:

- Flushing (incidence, severity)
- Gastrointestinal symptoms (incidence, severity)
- Reasons for stopping study treatment
- Transaminitis (ALT >3x upper limit of normal)
- Acute kidney injury (creatinine >1.5x value entered at randomisation)

### 10 6-MONTH ASSESSMENTS

This section details the proposed analysis of the clinical outcomes 6 months after initial randomisation in the RECOVERY trial.

#### 10.1 Trial outcomes

Unless otherwise specified, primary, secondary, subsidiary, and safety outcomes are as specified earlier in this document. Subgroup analyses will be conducted in the same subgroups as used in the 28 day outcome publications.

##### 10.1.1 *Changes to definition of clinical outcomes*

###### *10.1.1.1 Use of ventilation*

For the secondary and subsidiary clinical outcomes, use of ventilation includes ventilation occurring during index admission, or where the participant is readmitted. (Elective admissions will be excluded since ventilation recorded during such admissions are likely to be related to elective surgery rather than complications of COVID-19.)

###### *10.1.1.2 Use of renal dialysis or haemofiltration*

Use of renal dialysis or haemofiltration at any point during the 6 months following randomisation is included.

##### 10.1.2 *Additional exploratory analyses*

###### *10.1.2.1 Hospital recorded diagnoses*

For UK participants, diagnoses recorded in hospital datasets after randomisation are identified where they are the primary diagnoses relating to a period in inpatient care. These diagnoses are classified according to whether they are recorded during a planned or emergency (including transfers) admission.

Diagnoses will be tabulated by the categories defined for analysis of cause specific mortality (see below and section 2.6.4). Additional subcategories will be considered (see table below) with any categories containing a small number of events (e.g. fewer than 10) combined with other relevant categories.

Table: Sub-categories of hospital recorded diagnosis considered

| | | |
|---|---|---|
| COVID-19 | | |
| Other infection | Skin soft tissue | Bacterial/fungal/viral/TB/other/unspecified |
| | Abdominal | Bacterial/fungal/viral/TB/other/unspecified |
| | Respiratory | Bacterial/fungal/viral/TB/other/unspecified |
| | Bone and joint | Bacterial/fungal/viral/TB/other/unspecified |
| | Urinary | Bacterial/fungal/viral/TB/other/unspecified |
| | Bloodstream | Bacterial/fungal/viral/TB/other/unspecified |
| | Other | Bacterial/fungal/viral/TB/other/unspecified |
| | Unspecified | Bacterial/fungal/viral/TB/other/unspecified |

| | | |
|---|---|---|
| Cardiovascular | Cardiac<br>Stroke<br>Other vascular | MI/other CHD/Heart failure/other cardiac<br>Haemorrhagic/ischaemic/unknown<br>Arterial thrombo-embolism/venous thromboembolism/other vascular |
| Other | Cancer<br>Diabetes<br>Extra-cranial bleed or perforation<br>Liver<br>Renal<br>Respiratory (not infection)<br>Other medical cause | <br><br>GI/other |
| External | | |
| Unknown | | |

The start date for each diagnosis extracted from the hospitalisation dataset will be used for time-to-event analyses.

Data on admissions to hospital after randomisation will be collected in a complementary way for participants outside the UK.

###### *10.1.2.2 Total duration of critical and hospital in-patient care*

In order to assess the total burden of care for the participant and the health system, the following will be extracted from the routine healthcare data and presented as mean (SD) duration in days:

- Total duration (in days) of hospital in-patient care during the 6 months after randomisation
- Total duration (in days) of critical care during the 6 months after randomisation
- Total number of admissions categorised by planned vs emergency (including transfers)

#### 10.2 Censoring and analysis

For the 6 month analyses, participants will be censored at the earliest of death, withdrawal of consent or on study day 184 (where day of randomisation is study day 1).

By 6 months, nearly all participants have either died or been discharged alive, allowing the full effects of the trial treatments on the index admission (i.e. the admission in which the participant was randomised) to be assessed.

### 11 REFERENCES

#### 11.1 Trial documents

Study protocol, case report forms, training materials, and statistical analysis plan are published on the trial website.

#### 11.2 Other references

1. Gamble C, Krishan A, Stocken D, Lewis S, Juszczak E, Doré C, Williamson PR, Altman DG, Montgomery A, Lim P, Berlin J, Senn S, Day S, Barbachano Y, Loder E. Guidelines for the Content of Statistical Analysis Plans in Clinical Trials. JAMA 2017;318(23):2337-2343.

2. Schulz KF, Altman DG, Moher D for the CONSORT Group. CONSORT 2010 Statement: updated guidelines for reporting parallel group randomised trials. BMJ 2010;340:698-702.

3. Juszczak E, Altman DG, Hopewell S, Schulz KF. Reporting of multi-arm parallel-group randomized trials: extension of the CONSORT 2010 statement. JAMA 2019;321(16):1610-1620.

4. Dimairo M, Pallmann P, Wason J, Todd S, Jaki T, Julious SA, Mander AP, Weir CJ, Koenig F, Walton MK, Nicholl JP, Coates E, Biggs K, Hamasaki T, Proschan MA, Scott JA, Ando Y, Hind D, Altman DG; ACE Consensus Group. The Adaptive designs CONSORT Extension (ACE) statement: a checklist with explanation and elaboration guideline for reporting randomised trials that use an adaptive design. BMJ. 2020 Jun 17;369:m115. doi: 10.1136/bmj.m115. PMID: 32554564; PMCID: PMC7298567.

5. Peto R, Pike MC, Armitage P, Breslow NE, Cox DR, Howard SV, Mantel N, McPherson K, Peto J, Smith PG. Design and analysis of randomized clinical trials requiring prolonged observation of each patient. Part II: analysis and examples. Br J Cancer 1977;35:1-39.

6. Betensky RA and Schoenfeld DA. Nonparametric Estimation in a Cure Model with Random Cure Times. Biometrics 2001;57:282-286.

7. The National SARS-CoV-2 Serology Assay Evaluation Group. Performance characteristics of five immunoassays for SARS-CoV-2: a head-to-head benchmark comparison. Lancet Infect Dis 2020; 20: 1390-1400

8. McCullagh P. Regression models for ordinal data (with discussion). J R Statist Soc B. 1980;42:109–142.

9. Howard G, Waller JL, Voeks JH, Howard VJ, Jauch EC, Lees KR, et al. A simple, assumption-free, and clinically interpretable approach for analysis of modified Rankin outcomes. Stroke. 2012;43:664–669

10. Rubin D. Multiple imputation for non-response in surveys. New York: John Wiley; 1987.

### 12 APPENDIX A: ANALYSES OF REGN-COV2

#### 12.1 Background & rationale

The RECOVERY trial is testing multiple interventions in a broad population of patients hospitalised with COVID-19. The protocol and statistical analysis plan outline the methods that are to be used in the analysis of these interventions and, to date, the same approach has been appropriate for all completed comparisons. However, it is important that the statistical analysis plan be informed by the best available information about the treatment being tested[1] and the pathophysiology of the disease.

Relevant new information about the effects of REGN-COV2 have emerged since it was added to the trial in September 2020.

REGN-COV2 is a mixture of two synthetic monoclonal antibodies which bind to the receptor binding domain of the SARS-CoV-2 spike protein and neutralise the virus.[2] Recently-published trials of REGN-COV2 in ambulatory patients (i.e. those recently diagnosed in the community) have demonstrated that it has larger effects on viral load among people who are "seronegative" at the time of randomisation (i.e. they do not have detectable antibodies of their own against SARS-CoV-2), and seropositive patients derive little or no benefit (in terms of reduction in viral load) from REGN-COV2, compared to placebo.[3] Participant serostatus therefore is a potentially key modifier of the effect of REGN-COV2 that may be observed in RECOVERY.

All participants entering the REGN-COV2 comparison in RECOVERY are asked to provide a serum sample which is sent to a central laboratory at the University of Oxford, where antibodies against SARS-CoV-2 are measured using a validated assay. Previous assessments of this assay alongside commercially available assays shows excellent performance at discriminating prior SARS-CoV-2 infection with sensitivity and specificity above 98%.[4]

Earlier versions of the statistical analysis plan recognised the importance of the seronegative subgroup, but review of the emerging literature and regulatory guidance[5] has led to a change in approach to these analyses. The revised analysis plan for the REGN-COV2 comparison explicitly tests the hypothesis that any benefit of REGN-COV2 on the primary outcome may be wholly or largely restricted to patients who are seronegative at the time of randomisation with little or no benefit among those who are seropositive at that point.

For the avoidance of doubt, all decisions about this modification to the analytical plan were made before recruitment was complete and before any members of the trial steering committee (who are responsible for drafting and approving the SAP) or investigators had access to any unblinded analyses of clinical outcome data for the REGN-COV2 comparison. No members of the independent Data Monitoring Committee (who are the only individuals who can review interim unblinded analyses) were involved in this change.

#### 12.2 Analytical plan

The primary outcome and secondary outcomes remain unchanged. For each outcome, rate ratios and 95% confidence intervals will be calculated separately for participants who are seronegative, seropositive, or with unknown status as well as for the whole trial population. A test for heterogeneity between seronegative and seropositive participants will be presented. The results will be interpreted based on the totality of the evidence.

For the purposes of any regulatory submission: Because any beneficial effect of REGN-COV2 is hypothesised to be larger among seronegative participants (and may be negligible in seropositive participants), the primary outcome will first be assessed among participants who are known to be seronegative at randomisation. If the null hypothesis is rejected in the seronegative group at 2-tailed p=0.05, then the primary outcome will be assessed among the whole population (i.e. seronegative, seropositive, and those with unknown status combined). Otherwise, no further hypothesis testing will be performed.

A similar approach will be taken for each of the two pre-specified secondary outcomes (discharge alive within 28 days and, among patients not on invasive mechanical ventilation at baseline, the use of invasive mechanical ventilation or death) if both primary hypotheses are rejected. Hypothesis testing will first be conducted among the participants who are known to be seronegative at randomisation and, if the null hypothesis is rejected at 2-tailed p=0.025, then will be assessed among the whole population (see Table).

**Table: Hierarchical Testing Order**

| Hierarchy Number | Type of Outcome | Outcome | Analysis Population | Significance level, α (2-sided) |
|---|---|---|---|---|
| 1. | Primary | Mortality (all-cause), 28 days after randomisation | Seronegative at randomisation | 0.05 |
| 2. | Primary | Mortality (all-cause), 28 days after randomisation | All participants randomised | 0.05 |
| 3.* | Secondary | Time to discharge alive from hospital, within 28 days after randomisation | Seronegative at randomisation | 0.025 |
| 4. | Secondary | Time to discharge alive from hospital, within 28 days after randomisation | All participants randomised | 0.025 |
| 3.* | Secondary | Use of invasive mechanical ventilation (including ECMO) or death | Seronegative and not on invasive mechanical ventilation at randomisation | 0.025 |
| 4. | Secondary | Use of invasive mechanical ventilation (including ECMO) or death | All participants randomised not on invasive mechanical ventilation at randomisation | 0.025 |

* These will be performed simultaneously. Testing will only proceed to the respective overall population if the null hypothesis is rejected in the seronegative group at the specified level of statistical significance.

#### 12.3 References

1. Food and Drug Administration. E9 Statistical Principles for Clinical Trials. 1998.
2. Hansen J, Baum A, Pascal KE, et al. Studies in humanized mice and convalescent humans yield a SARS-CoV-2 antibody cocktail. Science 2020;369:1010-4.
3. Weinreich DM, Sivapalasingam S, Norton T, et al. REGN-COV2, a Neutralizing Antibody Cocktail, in Outpatients with Covid-19. N Engl J Med 2021;384:238-51.

4. National S-C-SAEG. Performance characteristics of five immunoassays for SARS-CoV-2: a head-to-head benchmark comparison. Lancet Infect Dis 2020;20:1390-400.
5. Food and Drug Administration. Enrichment strategies for clinical trials to support determination of effectiveness of human drugs and biological products - guidance for industry. 2019.

### 13 APPROVAL

| | | |
|---|---|---|
| Trial Statistician | Name: Mr Enti Spata | |
| | Signature: | Date: |
| Chief Investigator | Name: Professor Peter Horby | |
| | Signature: | Date: |
| Deputy Chief Investigator | Name: Professor Martin Landray | |
| | Signature: | Date: |
| Steering Committee Statistician | Name: Professor Edmund Juszczak | |
| | Signature: | Date: |
| Steering Committee Statistician | Name: Professor Alan Montgomery | |
| | Signature: | Date: |
| Steering Committee Statistician | Name: Professor Thomas Jaki | |
| | Signature: | Date: |

#### 14 DOCUMENT HISTORY

| Version | Date | Edited by | Comments/Justification | Timing in relation to unblinded interim monitoring | Timing in relation to unblinding of Trial Statisticians |
|---|---|---|---|---|---|
| 0.1 | 20/03/20 | LL/JB | First draft. | Prior | Prior |
| 0.2 | 01/04/20 | LL/JB | Comments and amendments from Martin Landray, Jonathan Emberson & Natalie Staplin. Also aligned with updated protocol and CRFs. | Prior | Prior |
| 0.3 | 01/04/20 | EJ/LL | Further edits and comments. | Prior | Prior |
| 0.4 | 07/04/20 | JB/EJ/LL | Following statistics group meeting on 02/04/20. | Prior | Prior |
| 0.5 | 22/04/20 | JB/LL/EJ | Following statistics group meeting on 09/04/20 and further protocol update. | After | Prior |
| 0.6 | 24/04/20 | LL | Following statistics group meeting on 23/04/20. | After | Prior |
| 0.7 | 10/05/20 | LL | Protocol update. | After | Prior |
| 0.8 | 15/05/20 | LL | Following statistics group meeting on 15/05/20. | After | Prior |
| 0.9 | 27/05/20 | LL | Further comments from TSC members prior to interim analysis on 28/05/20. | After | Prior |
| 1.0 | 09/06/20 | LL | Revised following the stopping of the hydroxychloroquine arm, and prior to the trial statisticians receiving unblinded data for this arm. | After | Prior |
| 1.1 | 21/06/20 | LL/JB/RH | Additional clarification of ventilation denominators. Adjustment for any imbalances of subgroup characteristics between treatment arms at randomisation. Clarification of analysis of composite outcome. Removal of 'Unknown' ethnicity subgroup. Addition of section 5.5 Adjustment for baseline characteristics. | After | After unblinding of hydroxychloroquine and dexamethasone arms. |

| Version | Date | Edited by | Comments/Justification | Timing in relation to unblinded interim monitoring | Timing in relation to unblinding of Trial Statisticians |
|---|---|---|---|---|---|
| 2.0 | 04/11/20 | EJ/ES | Revised to reflect changes in protocol, including introduction of factorial randomisations and new arms, including convalescent plasma, tocilizumab, synthetic neutralizing antibodies (REGN-COV2, and aspirin. | Prior to interim analysis of aspirin arm<br><br>After interim analyses of all other arms | After unblinding of 28-day results for hydroxychloroquine, lopinavir-ritonavir, and dexamethasone arms.<br><br>Prior to unblinding of any other arms |
| 2.1 | 02/12/20 | ES | Addition of colchicine. Modification of definition of recipient antibody concentration subgroup. | Prior to interim analyses including antibody results or of colchicine arm. | After unblinding of 28-day results for hydroxychloroquine, lopinavir-ritonavir, and dexamethasone arms.<br><br>Prior to unblinding of any other arms |
| 2.2 | 27/01/21 | ES | Clarification of non-invasive ventilation-related subgroups. Addition of baricitinib. | Prior to interim analyses of baricitinib arm. | After unblinding of 28-day results for hydroxychloroquine, lopinavir-ritonavir, azithromycin and dexamethasone arms (and primary outcome in overall population in convalescent plasma arm).<br><br>Prior to unblinding of any other arms |

| Version | Date | Edited by | Comments/Justification | Timing in relation to unblinded interim monitoring | Timing in relation to unblinding of Trial Statisticians |
|---|---|---|---|---|---|
| 3.0 | 15/05/21 | ES | Specification of method for REGN-COV2 comparison (appendix A).<br>Addition of early phase assessment of dimethyl fumarate.<br>Addition of infliximab and high-dose corticosteroids. | Prior to interim analyses of infliximab or high-dose steroids. | After unblinding of 28-day results for hydroxychloroquine, lopinavir-ritonavir, azithromycin, dexamethasone, colchicine and convalescent plasma arms.<br><br>Prior to unblinding of any other arms. |
| 3.1 | 29/10/21 | RH | Modification of early phase assessments to align with protocol V18.1<br>Modification of 6 months analysis section. | Prior to early phase assessment s or 6 month analyses. | Prior to unblinding of dimethyl fumarate or 6 month outcome data. |
| 3.2 | 17/12/21 | RH | Update to early phase assessments. | Prior to 6 month analyses | Prior to unblinding of dimethyl fumarate |

#### Appendix 3: Definition and Derivation of Baseline Characteristics and Outcomes

RECOVERY
Randomised Evaluation of COVID-19 Therapy

### Definition and Derivation of Baseline Characteristics and Outcomes

#### Contents

1 Version ... 2
2 Scope ... 2
3 Abbreviations ... 3
4 Data sources ... 3
4.1 Electronic case report forms ... 3
4.1.1 Main randomisation ... 3
4.1.2 Second randomisation ... 4
4.1.3 Convalescent plasma safety eCRF ... 5
4.1.4 Follow-up ... 5
4.1.5 Non-UK sites ... 5
4.2 Registries and NHS datasets ... 6
4.2.1 Hospital admissions datasets ... 6
4.2.2 PEDW contains data relating to admissions to NHS hospitals in Wales. The data fields used in the RECOVERY trial are equivalent to those used in SUSAPC and HESAPC Mortality datasets ... 7
4.2.3 COVID specific datasets ... 7
4.2.4 Intensive Care Datasets ... 8
4.2.5 Disease specific registries ... 9
5 Baseline characteristics ... 9
5.1.1 Baseline corticosteroid use ... 9
5.2 Additional baseline characteristics ... 10
6 Outcomes ... 10
6.1 All-cause mortality ... 10
6.1.1 Sources ... 10
6.1.2 Discrepancies ... 11
6.2 Cause-specific mortality ... 11
6.3 Time to discharge ... 11
6.3.1 Sources ... 12
6.3.2 Discrepancies ... 12
6.4 Use and duration of ventilation ... 12
6.4.1 Sources ... 12

13 Appendix 5: Example hospital recorded diagnoses showing extraction of start and end dates 23

### 1 Version

| Date | Version | Comments |
|---|---|---|
| 06-Jun-2020 | 0.1 | Initial version |
| 08-Jun-2020 | 0.2 | Minor updates |
| 09-Jun-2020 | 1.0 | First released version |
| 11-Dec-2020 | 2.0 | Update to sections 6.4 (use of assisted ventilation) and 6.6 (use of renal replacement therapy) |
| 06-Jan-2020 | 3.0 | Update to clarify the derivation of outcomes and baseline data for the second randomisation and define complete follow-up |
| 14-April-2022 | 4.0 | Updates to frequency of dataset transfers and additional datasets. Addition of section 8relating to 6-month outcomes. Addition of appendix 4 to provide detail on discharge outcome |

### 2 Scope

This document describes the definition and derivation of the primary, secondary and other outcomes of the RECOVERY trial for the published trial analyses. It should be read alongside the study protocol which defines the study outcomes briefly, and the Statistical Analysis Plan (SAP) which describes the statistical methods used to analyse these outcomes. The SAP refers to this document (see Section 2.6.4 Detailed derivation of outcomes) which provides detail on how the outcomes are defined, captured and derived.

Most outcomes have more than one potential source which improves completeness of capture but also will inevitably identify discrepancies between different sources. This document describes the principles for how such discrepancies are resolved; the rules for this were developed blind to results. Further details of the methods are described in the RECOVERY trial internal operating procedure for identifying data discrepancies.

### 3 Abbreviations

| | |
|---|---|
| ADDE | Annual District Death Extract |
| CCDS | Critical Care Dataset |
| CHESS | COVID-19 Hospitalisation in England Surveillance System |
| CPAP | Continuous Positive Airway Pressure |
| CRP | C-reactive protein |
| ECMO | Extra-corporeal membrane oxygenation |
| eCRF | Electronic Case Report Form |
| FCE | Finished Consultant Episode |
| FU | Follow-up |
| HESAPC | Hospital Episode Statistics Admitted Patient Care |
| HFNO | High-flow nasal oxygen |
| ICD-10 | International Classification of Diseases $10^{th}$ edition |
| ICNARC | Intensive Care National Audit and Research Centre |
| IMV | Invasive mechanical ventilation |
| NHSCR | NHS Central Register (Scotland) |
| NIV | Non-invasive ventilation |
| NRS | National Records of Scotland |
| ONS | Office for National Statistics (ONS) |
| OPCS-4 | Office of Population Censuses Surveys Classification of Surgical Operations and Procedures 4th revision |
| PDS | Patient Demographic Service |
| PEDW | Patient Episode Database for Wales |
| RRT | Renal replacement therapy |
| PHE | Public Health England |
| SAP | Statistical Analysis Plan |
| SICSAG | Scottish Intensive Care Society Audit Group |
| SMR | Scottish Morbidity Record |
| SUSAPC | Secondary Use Service Admitted Patient Care |
| UKRR | UK Renal Registry |
| WDSD | Welsh Demographic Service |
| WRRS | Welsh Results Reporting Service |

### 4 Data sources

#### 4.1 Electronic case report forms

##### 4.1.1 Main randomisation

The Randomisation eCRF is completed by hospital staff after patients (or a legal representative) have given consent to participate in the trial. It collects the following participant information:

- Identifiers
    - First name, family name

  - NHS number
  - Date of birth
  - Sex (male/female/unknown)
- Inclusion criteria
  - COVID-19 symptom onset date
  - Date of hospitalisation
- Details of acute illness
  - Requirement for oxygen[1]
  - Requirement for ventilatory support (none, continuous positive airway pressure, non-invasive ventilation, high-flow nasal oxygen, invasive mechanical ventilation (IMV) or extra-corporeal membrane oxygenation) (ECMO)
  - Latest oxygen saturation
  - Latest C-reactive protein, creatinine and D-dimer measurement (if available)
- Comorbidities
  - Diabetes
  - Heart disease
  - Chronic lung disease
  - Tuberculosis
  - HIV
  - Severe chronic liver disease
  - Severe kidney impairment (eGFR <30 mL/min/1.73$m^2$ or on dialysis)
  - Long QT syndrome
  - Pregnancy
- Current treatment
  - Macrolide antibiotics
  - Aspirin or other antiplatelet therapy
  - Warfarin or direct oral anticoagulant
  - Venous thromboembolism prophylaxis (standard or increased dose due to COVID-19)
  - Remdesivir
  - Systemic corticosteroids
- Other
  - Weight (children only)

###### 4.1.2 Second randomisation

The Second Randomisation eCRF is completed by hospital staff when they wish to randomise participants between tocilizumab or standard care alone if they fulfil the protocol-defined oxygenation and inflammation criteria. It collects the following participant information:

[1] NHS England advice published on 9 April 2020 stated that the usual oxygen target saturation for prescribed oxygen should change from 94-98% to 92-96% in the first instance. Hospitals may further reduce this to 90-94% if clinically appropriate according to prevailing oxygen demands. https://www.england.nhs.uk/coronavirus/wp-content/uploads/sites/52/2020/04/C0256-specialty-guide-oxygen-therapy-and-coronavirus-9-april-2020.pdf. Guidance on admission to hospital was similar in Scotland. https://www.nhsggc.org.uk/media/259232/covid-19_gps_national_supporting_guidance_for_scottish_general_practice.pdf although hospital guidelines in Scotland did not specify a target oxygen saturation.

- Inclusion criteria
    - Requirement for oxygen
    - Current level of ventilation support (none/CPAP/NIV/HFNO/IMV/ECMO)
    - Latest CRP
- Other information
    - Latest ferritin and creatinine

###### 4.1.3 Convalescent plasma safety eCRF

This eCRF is completed by hospital staff as soon as possible after 72 hours post-main randomisation for participants who entered the convalescent plasma comparison. It collects the following information:

- Adherence to convalescent plasma allocation (number of units received, whether any were stopped early)
- Adverse events
    - Sudden worsening of respiratory status
    - Severe allergic reaction
    - Temperature ≥39C (or rise ≥2C above baseline)
    - Sudden hypotension
    - Clinical haemolysis
    - Thrombotic event

###### 4.1.4 Follow-up

The FU eCRF is completed by hospital staff at the earliest of (i) discharge from acute care (see Section 6.3 below), (ii) death, or (iii) 28 days after the main randomisation. It collects the following information from date of randomisation onwards:

- Adherence to randomised allocation, and receipt of other study treatments or relevant therapies (and number of days of treatment)
- Vital status and underlying cause of death (COVID, other infection, cardiovascular, other; if other, a free text description is collected)
- Date of discharge
- Requirement for assisted ventilation (CPAP, NIV, HFNO, IMV, ECMO) and number of days of assisted ventilation and IMV/ECMO separately
- Occurrence of major cardiac arrhythmia (atrial flutter/fibrillation, supraventricular tachycardia, ventricular tachycardia [including torsades de pointes], ventricular fibrillation or bradycardia requiring intervention) (from 12 May 2020)
- Occurrence of thrombotic event (pulmonary embolism; deep-vein thrombosis; ischaemic stroke; myocardial infarction; systemic arterial embolism; other) (from 6 November 2020)
- Occurrence of clinically-significant bleeding i.e. intracranial or requiring intervention (blood transfusion; surgery; endoscopy; vasoactive drug or blood transfusion), by site (intra-cranial; gastrointestinal; other) (from 6 November 2020)
- Requirement for renal replacement therapy and peak creatinine after randomisation
- Other infections after randomisation (by site and by organism type)
- Metabolic complications (ketoacidosis, hyperglycaemia, hypoglycaemia)

###### 4.1.5 Non-UK sites

Whereas in the UK participants will be followed by linkage with routinely collected data (see Section 4.2) for up to 10 years after randomisation, in other countries this is not possible. Sites will be asked to complete an additional case report form for participants discharged alive from hospital at 28 days after randomisation to confirm vital status (and date and cause of death if relevant).

#### 4.2 Registries and NHS datasets

##### 4.2.1 Hospital admissions datasets

###### *4.2.1.1 Secondary Use Service Admitted Patient Care*

The SUSAPC dataset is a repository of data hosted by NHS Digital that relates to in-patient care provided in England, which aims to enable reporting and analyses to support the NHS in the delivery of healthcare services. These data are submitted on a regular basis by NHS hospital trusts and at pre-arranged dates during the year. Submissions are consolidated, validated and cleaned and then incorporated into the HESAPC dataset. Data may be incomplete in places and is not quality assured to the same extent as HES, but is available more rapidly.

In the SUSAPC dataset, each record contains data relating to a continuous period of care under one consultant known as a Finished Consultant Episode (FCE). FCEs can be grouped together to form 'Spells'. Each spell is a continuous periods of inpatient care within one hospital. Each FCE contains data about the patient (e.g. sex, ethnicity), the specialty providing the care (e.g. cardiology), ICD-10 diagnostic and OPCS-4 procedure codes, along with dates for each procedure and details about the admission and discharge and other data.

For the main RECOVERY analyses the following data are used;

- Admission method (which indicates whether the admission was emergency or elective and whether it involved a transfer from another healthcare provider)
- Admission source (used to identify transfers between hospitals)
- Ethnicity
- Sex
- Date of admission and discharge
- Start and end date of the FCE
- Discharge method and destination (which may indicate death of participant)
- Diagnoses recorded during FCE (ICD-10 coded)
- Procedures performed during FCE (OPCS-4 coded) and corresponding dates

Linked SUSAPC data are imported to the RECOVERY trial database approximately monthly.

###### *4.2.1.2 Hospital Episode Statistics Admitted Patient Care*

HESAPC contains data relating to admissions to NHS hospitals in England and is produced from the SUSAPC following a number of cleaning and validation steps. For participants in England, HESAPC is available for the 5 year period prior to enrolment in the study. For the main RECOVERY analyses these data are used to identify prior medical conditions on the basis of recorded ICD-10 and OPCS-4 codes (excluding the admission during which the patient was randomised). For the analysis of 6-month outcomes, these data are used to identify the Hospital Recorded Diagnoses (see section 8). Linked HESAPC data are imported to the RECOVERY trial database quarterly.

###### *4.2.1.3 NHS Central Register Scottish Morbidity Record One*

The NHSCR SMR01 data set holds episode level data on hospital inpatient and day case discharges from acute specialities from hospitals in Scotland. The data fields used in the RECOVERY trial are equivalent to those used in SUSAPC and HESAPC. Linked NHSCR-SMR01 data are imported approximately monthly.

###### *4.2.1.4 Patient Episode Data Wales*

##### 4.2.2 PEDW contains data relating to admissions to NHS hospitals in Wales. The data fields used in the RECOVERY trial are equivalent to those used in SUSAPC and HESAPC Mortality datasets

###### *4.2.2.1 Patient Demographic Service*

The PDS is the electronic database of NHS patient details such as name, address, date of birth and NHS Number for patients in England. For RECOVERY it is used to provide information on fact and date of death. It provides both 'informal' notifications of death (which occur when a health care provider is informed of their patients death and records the reported date of death in their electronic data systems) and 'formal' notifications of death (which are provided by the Office for National Statistics).

###### *4.2.2.2 Office for National Statistics Mortality data*

The ONS mortality data contains information related to a person's death taken from the death certificate for all deaths registered in England and Wales. The following data are provided

- The underlying cause of death
- Contributory causes of death
- Other conditions recorded on the death certificate but not contributing to death
- Whether a post-mortem took place

Clinical data are recorded using ICD-10 codes. Linked ONS mortality data are imported into the RECOVERY trial via a quarterly extract from NHS Digital.

###### *4.2.2.3 Welsh Demographic Service*

WDS data are the electronic database of NHS patient details for patients in Wales and are similar to PDS (4.2.2), providing fact and date of death (including formal or informal notifications). Linked data for RECOVERY participants recruited via sites in Wales will be available for future analysis.

###### *4.2.2.4 National Records of Scotland Mortality Data*

The NRS mortality data contain information related to a person's death taken from the death certificate for all deaths registered in Scotland. The data provided includes the date of death and the underlying and contributory causes of death coded in ICD-10. Linked data are imported into the RECOVERY trial database approximately monthly.

##### 4.2.3 COVID specific datasets

###### *4.2.3.1 Public Health England Second Generation Surveillance data*

The SGSS is an application that captures, stores and manages routine laboratory surveillance data on infectious diseases and antimicrobial resistance from laboratories across England. Once the reports have been loaded into SGSS, each record is subject to a number of validation processes, and local LIMS codes are translated to SGSS codes to standardise the data for analysis. The data is stored in a central database within PHE and details of tests indicating SAR-CoV-2 have been made available to NHS Digital for dissemination for a limited time period. For each test, the following data are available

- Date the sample was collected
- Date the result was reported
- Organism identified (only SARS-CoV-2)

Linked PHE SGSS data are imported into the RECOVERY trial approximately monthly.

###### *4.2.3.2 Public Health Scotland COVID-19 laboratory antigen test positive list*
The Electronic Communication of Surveillance in Scotland (ECOSS) collects routine laboratory surveillance data on infectious diseases from laboratories in Scotland. The data provided to RECOVERY is limited to SARS-CoV-2 results along with the date of the sample and result.

###### *4.2.3.3 Welsh Results Reporting Service Pathology Data*
The WRRS contains all Pathology Test Results for Wales in a single database. Tests indicating a positive SAR-CoV-2 antigen linked to the trial participants are obtained.

###### *4.2.3.4 COVID-19 Hospitalisation in England Surveillance System*
PHE has established the COVID-19 Hospitalisation in England Surveillance System (CHESS), which collects epidemiological data (demographics, risk factors, clinical information on severity, and outcome) on COVID-19 infection in patients requiring hospitalisation and ICU/HDU level care. This dataset has been made available to NHS Digital for dissemination for a limited time period. For RECOVERY the following information is used;

- Date of ICU/HDU admission and discharge
- Use of respiratory support during the admission (including oxygen via cannulae or mask, high flow nasal oxygen, non-invasive ventilation, invasive mechanical ventilation and ECMO)
- Complications during the admission (including viral pneumonia, secondary bacterial pneumonia, ARDS, unknown, and other co-infections)

The CHESS dataset is imported into the RECOVERY trial approximately monthly.

###### *4.2.3.5 GPES Data for Pandemic Planning and Research (COVID-19) (GDPPR)*
GDPPR data is available for RECOVERY participants in England. Data includes patient demographic information and coded medical information (mainly in SNOMED codes).

###### 4.2.4 Intensive Care Datasets

###### *4.2.4.1 Intensive Care National Audit and Research Centre*
The ICNARC Case Mix Programme is the national clinical audit covering all NHS adult, general intensive care and combined intensive care/high dependency units in England, Wales and Northern Ireland, plus some additional specialist and non-NHS critical care units. Data are collected about the first 24 hours in ICU/HDU and at discharge from the ICU/HDU with a further data collection point after discharge from hospital. For RECOVERY, the following data recorded at discharge from ICU/HDU are used:

- Date of admission to and discharge from ICU/HDU
- Use of Advanced Respiratory Support (ARS), Basic Respiratory Support (BRS) or Renal Support during the admission
- The number of days of ARS, BRS or Renal Support during the admission
- Date of death (if relevant)

Linked ICNARC data is requested for hospitals recruiting to RECOVRY and are imported approximately monthly.

###### *4.2.4.2 Scottish Intensive Care Society Audit Group*
SICSAG collects data from all general adult Intensive Care Units, Combined Units and the majority of High Dependency Units in Scotland using the WardWatcher system. The following data are used in the RECOVERY trial:

- Date of admission and discharge from ICU/HDU

- Used of mechanical ventilation via endotracheal tube or tracheostomy and use of haemofiltration for each day of during admission

Linked SICSAG data are imported into the RECOVERY trial approximately monthly.

###### *4.2.4.3 Critical Care dataset*

In England and Wales much of the key data collected by ICNARC is also available in the CCDS from NHS Digital or the SAIL datalink Wales. However, both the ICNARC and CCDS data can be subject to different delays during collection, consolidation and dissemination and therefore either source may be incomplete at any one time-point. Both sources are therefore combined to provide information about ICU/HDU care for participants in England and Wales.

##### 4.2.5 Disease specific registries

###### *4.2.5.1 UK Renal Registry*

The UK Renal Registry collates data from renal units and hospital laboratories in all four nations in the UK. Linked data relating to laboratory tests for patients who trigger a hospital laboratory “acute kidney injury alert” are available for a subset of patients. Data relating to the provision of care for end stage kidney disease discuss is provided to RECOVERY on an annual basis.

### 5 Baseline characteristics

Baseline characteristics for the trial cohort are obtained from the first randomisation eCRF for the main randomisation comparisons. For the second randomisation comparisons, the baseline data are obtained either from the second randomisation form directly (e.g. baseline use of respiratory support) or from a calculation based on the first randomisation form data and the number of days between the first and second randomisation forms (e.g. days since symptom onset).

Where fields are missing, they may be supplemented by data from the linked health care data. Generally corrections to the randomisation eCRF data are not made. Exceptions to this would include key participant identifiers (Date of birth, NHS or CHI number) or cases where information is missing. For example, if a site later report that the date of birth was entered incorrectly, this would be confirmed with the site (recorded in the trial data query system) and updated (with appropriate audit trail).

##### 5.1.1 Baseline corticosteroid use

Baseline steroid use is determined as follows:

- Baseline steroid use = yes if allocated dexamethasone in main randomisation OR responded ‘yes’ to baseline steroid question on main randomisation form (OR [for tocilizumab comparison only] responded ‘yes’ to baseline steroid question on second randomisation form
- Otherwise, Baseline steroid use = no if answered ‘no’ to steroid question on main OR [for tocilizumab comparison only] second randomisation forms
- Otherwise, Baseline steroid use = not asked if recruited prior to June 18th[2]
- Otherwise, Baseline steroid use = unknown

[2] From 18th June onwards a question on baseline systemic corticosteroid use was added to the main randomisation form following the release of the dexamethasone comparison results.

For the purposes of analysis, baseline steroid use = no and not asked will be combined for subgroup analyses. Participants with baseline steroid use = unknown will be exluded from subgroup analysis, but the number in this subgroup provided in a footnote.

#### 5.2 Additional baseline characteristics

Some baseline characteristics that are not collected on the randomisation eCRF may be extracted from registry data or other sources. These include:

- Ethnicity by Office for National Statistics 2001 census categories (White, BAME [Mixed, Asian or Asian British, Black or Black British, Other Ethnic Groups], Unknown) from linked health care records. Ethnic groups characterised using SNOMED codes within the GDPPR data are mapped to these categories. Where ethnicity records are discrepant between individual episodes in HES/SMR01/PEDW, the most frequently recorded code is used. Within the GDPPR dataset ethnicity is recorded in two places, the ethnic field in the patient table and the presence of a relevant SNOMED code in the journals table. The most recent code in the journals table is used, where available, otherwise the code from the patient table is used. Where there is discrepancy between the best estimate from GDPPR and HES/SMR01/PEDW existsthe GDPPR code is used. Where neither are available the most frequent fode in the SUS data is used. Individual SNOMED codes are categorised as defined with the SNOMED hierarchy and ethnicity categoriese according to the UK department of health categories.[3]
- Confirmed SARS-CoV-2 diagnostic test from linked health care records. A positive SARS-CoV-2 with a test date within 28 days of the date of first randomisation is considered as confirmed SARS-CoV-2. In the absence of such data for a participant, the data from the randomisation eCRF may be used.
- Comorbidity score: It is possible to calculate comorbidity and frailty scores (e.g. Charlston Comorbidity Score) from prior linked hospital admissions data and this will be done for future exploratory analyses (not specified in the trial SAP).
- Prior End Stage Kidney Disease (see section 6.6)
- Risk: The risk of death by 28 days can be modelled using available baseline characteristics (in the overall trial population) and a risk score derived. Participants will be divided into thirds based on this score (such that each third has approximately the same number of deaths), with the tertiles rounded to clinically-relevant values. For the main trial analyses the groups will defined as risk of death by 28 days of <30%; ≥30 ≤45%; and >45%.

### 6 Outcomes

#### 6.1 All-cause mortality

The primary outcome is all-cause mortality at 28 days after randomisation. All-cause mortality will also be assessed at 6 months and other later time points.

##### 6.1.1 Sources

Information on death may come from the following sources:

- FU eCRF (for deaths within first 28 days after randomisation)
- PDS (for participants in England)
- PDS Wales ((or participants in Wales)
- SUSAPC (for participants in England)

[3] https://www.ethnicity-facts-figures.service.gov.uk/style-guide/ethnic-groups

- SMR01 (for participants in Scotland)
- PEDW (for participants in Wales)
- ONS mortality data (for participants in England and Wales)
- NRS mortality data (for participants in Scotland)

In general, the primary source will be considered ONS (which includes formal death notification within PDS) and NRS mortality data as these are the official national death registries.

##### 6.1.2 Discrepancies

###### *6.1.2.1 Fact of death*

The ONS and NRS mortality data will be considered the defining source for fact of death. In order to allow rapid analysis of results, other sources (e.g. informal death notification via PDS, report of death on the FU eCRF, report of death from SUSAPC) are used for DMC and interim analyses. Cases where these reports are not later substantiated by ONS or NRS are individually reviewed and are not considered as deaths, unless a suitable explanation exists.

###### *6.1.2.2 Date of death*

The ONS and NRS data will be considered the defining source for date of death. In order to allow rapid analysis of data, other sources may be used. Where data sources are discrepant the following hierarchy is applied;

- ONS/NRS (most reliable for date of death), then
- Linked hospital admissions data, then
- FU eCRF , then
- PDS informal death notification (least reliable for date of death)

#### 6.2 Cause-specific mortality

The cause of death for the 28 day analysis will be the underlying cause of death as provided by ONS. The causes of death will be categorised as follows:

- Non-vascular death
    - Death from infection
        - Death from COVID-19
        - Death from other infection
    - Death from cancer
    - Death from other medical causes
    - External deaths
- Vascular death
    - Cardiac death
    - Stroke death
    - Other vascular death
- Unknown death

The ICD-10 codes contributing to these categories are available to download from the RECOVERY website.

#### 6.3 Time to discharge

Time to discharge (which is a more accurate term for duration of admission because only the period from randomisation onwards is relevant) is defined as the number of days a participant remained in hospital for acute care after randomisation. Discharge excludes transfer to another acute hospital, but might include transfer to community hospital for rehabilitation or a hospice for end-of-life care.

###### 6.3.1 Sources

Information on date of discharge may come from the following sources:

- FU eCRF
- SUSAPC (for participants in England)
- PEDW (for participants in Wales)
- SMR01 (for participants in Scotland)

The participant is considered to have been discharged from hospital if there is a discharge date recorded with a discharge method and destination which do not indicate that the participant died or was transferred (see appendix 4). In addition there must be no other admission with an admission date up to 4 days before or 1 day after the discharge date where either the method or source of the admission recorded suggest transfer from another hospital (see appendix 4). The first date of discharge which fulfils these criteria after first or second randomisation is used to determine time to discharge.

###### 6.3.2 Discrepancies

Linked hospital admissions data will be used if date of discharge is discrepant with FU eCRF data. If no linked hospital admissions data are available and the FU eCRF indicates discharge without a date, the date of completion for the FU eCRF will be used.

##### 6.4 Use and duration of ventilation

Assisted ventilation can be broadly divided into

i. Invasive mechanical ventilation (IMV) which includes ECMO (a secondary outcome in combination with all-cause mortality)
ii. Non-invasive ventilation which includes CPAP, NIV and HFNO (which are included in the subsidiary outcomes)

Information on non-invasive ventilation was collected because at the time the trial was designed there were concerns that the availability of mechanical ventilators would be insufficient to meet demand, so some patients would be treated with non-invasive ventilation when in other circumstances they would have received invasive mechanical ventilation. In reality this situation did not occur, so the emphasis of the analyses (and efforts to resolve discrepancies) is on invasive mechanical ventilation.

###### 6.4.1 Sources

Information on ventilation may come from the following sources:

- FU eCRF
- SUSAPC/SMR01/PEDW
- ICNARC
- SICSAG
- CHESS
- CCDS

However, the coding of ventilation is different in each source.

###### 6.4.2 Fact of assisted ventilation

A participant is considered to have received IMV/ECMO if use of these treatments was recorded on the FU eCRF; if a relevant procedure code was recorded in

SUSAPC/SMR01/PEDW within 28 days of randomisation (Appendix 1); if days of advanced respiratory support (ARS) in the ICNARC/CCDS data were considered to fall between randomisation and 28 days (see section 6.4.3) or if the daily SICSAG record indicated that the participant was receiving respiratory support via an endotracheal tube or tracheostomy.

A participant is considered to have received non-invasive ventilation if the site recorded 'yes' to the question 'did the participant receive assisted ventilation' or 'yes' to any of the individual types of non-invasive ventilation (CPAP, BIPAP, HFNO) on the FU eCRF; if a relevant procedure code was recorded in SUSAPC/SMR01/PEDW within 28 days of randomisation (Appendix 2) or if use of HFNO or NIV was recorded in CHESS when the admission and discharge date were both between randomisation and 28 days.

##### 6.4.3 Duration of invasive mechanical ventilation

The data from the critical care datasets (ICNARC, CCDS and SICSAG) are considered the primary source of the duration of IMV. Within ICNARC/CCDS, ARS is considered to be equivalent to IMV, however only the dates of admission and discharge from ICU/HDU and the number of days of ARS are provided. The days of ARS within each critical care episode are assumed to be continuous. The days of ARS were assumed to include randomisation if the participant was recorded as receiving IMV at baseline on the first or second randomisation eCRF as appropriate. Otherwise, the days of ARS are assumed to start from admission to critical care, occur at the mid-point of the critical care admission or end on discharge from critical care depending on the level of care recorded on admission and discharge and, in some cases, the destination on discharge (Appendix 2). Using these assumptions, the information from both ICNARC and the CCDS were used to identify whether IMV was received on each of the 28 days following randomisation. The SICSAG daily record indicated use of IMV on each day.

If no relevant information on IMV is received from ICNARC/CCDS/SICSAG, then the duration of IMV was obtained from the FU eCRF. Cessation of mechanical ventilation is deemed successful if it occurs within (and the participant survives until) 28 days after randomisation.

#### 6.5 Major cardiac arrhythmia

Major cardiac arrhythmias are defined as either:

i. Atrial flutter or fibrillation
ii. Supraventricular tachycardia
iii. Ventricular tachycardia (including torsades de pointes)
iv. Ventricular fibrillation
v. Significant bradycardia (requiring intervention)

##### 6.5.1 Sources

Information on cardiac arrhythmias is collected on the FU eCRF (but only for those eCRFs completed from 12 May 2020 onwards when these outcomes were added).

#### 6.6 Renal replacement therapy

Renal replacement therapy (RRT) includes haemodialysis, haemofiltration (and their combination) and peritoneal dialysis. (Kidney transplantation is not relevant in this case.) Individuals receiving RRT at baseline are identified as follows;

- Patients already receiving renal replacement for End Stage Kidney Disease at baseline are identified using linked hospitalisation data (appendix 3).

- From the ICNARC/CCDS data, the combination of the number of Renal Support Days and the start and end date of a critical episode may imply that they must have been receiving renal support at randomisation.
- The SICSAG daily record indicates that Renal Support was received on the day of, or on the day before randomisation.
- A procedure code in SUS/SMR01/PEDW indicating dialysis or haemofiltration with a date within the 3 days prior to first or second randomisation as appropriate (appendix 1).
- (When available) A record of prior RRT (without documented recovery) from the UK Renal Registry

##### 6.6.1 Sources

- FU eCRF
- Linked hospitalisation data (SUSAPC, HES, PEDW, SMR01)
- ICNARC
- SICSAG
- UKRR

##### 6.6.2 Discrepancies

Use of RRT is collected on the FU eCRF. Use of RRT is also identified within the linked hospitalisation data from relevant OPCS-4 codes (Appendix 1). Use of RRT in the ICNARC/CCDS is identified from the recording of Renal Support days where the both the date of admission to and discharge from critical care fall between randomisation and 28 days. The SICSAG daily record indicates RRT if Renal Support is recorded on any day between randomisation and 28 days.

Further information on renal outcomes may become available from the UK Renal Registry data.

### 7 Competeness of Follow-up

For the 28 day analysis, follow-up information is considered to be complete if a FU eCRF has been completed, or data has been received from a hospital admissions dataset (SUSAPC, PEDW or SMR01) which includes data from the admission during which the participant was randomised.

### 8 Analysis of outcomes at 6-months

#### 8.1 Collection of outcomes at 6-months in the UK

In the UK, outcome collection after the initial 28-day follow-up is undertaken by linkage to the routine healthcare datasets, with no further eCRF completion by the site staff. Unless indicated below, the outcomes analysed at 6-months are derived in the same way as for the main trial analyses described in section 6.

##### 8.1.1 Use of ventilation

For the analysis of outcomes at 6-months, use of ventilation is defined in the same way as described in section 6.4. However, periods of ventilation during an elective (i.e. planned) admission following the index admission are excluded, since such procedures are likely to be related to elective surgery rather than complications of COVID-19. Dates of subsequent admissions are obtained from HESAPC and categorised into elective admission or non-elective admission (including emergency admissions and transfers) on the basis of recorded the admission method (see Appendix 4).

##### 8.1.2 Hospital recorded diagnosis

Diagnoses recorded as the primary reason for a period of in-hospital care are extracted from HESAPC, SMR01 and PEDW. Diagnostic codes are restricted to the first diagnostic position and ICD-10 codes in other positions are not considered. ICD-10 codes within the same block (e.g. I25.1 and I25.2) are considered to relate to the same hospital recorded diagnosis. For each hospital spell the first ICD-10 code recorded within the relevant block is extracted along with a start and end date. The start date is defined as the start of the first episode in which an ICD-10 code in the relevant block is recorded within that spell. The end date is defined as the end of the episode in which an ICD-10 code in the relevant block is recorded within that spell. Examples showing how the dates are extracted are shown in Appendix 5.

Diagnoses for which the first record in that spell is in an episode which started after randomisation are considered to be post-randomisation. Only post-randomisation diagnoses are to be used for the analyses.

Caution should be applied when considering absolute event rates derived from the hospital recorded diagnosis. As can be seen from example 1 and 3 in Appendix 5, more than one hospital recorded diagnoses could be derived from one clinical event, where ICD-10 codes from different blocks are used to record the same clinical event in subsequent episodes. While this is unlikely to result in bias when assessing the proportional effects of treatment, the absolute number of hospital recorded diagnoses should not be interpreted as the absolute number of serious adverse events.

##### 8.1.3 Total duration of critical and hospital in-patient care

Total duration of hospital in-patient care during the 6-months after randomisation is derived from HESAPC based on admission and discharge dates. This is categorised separately by elective vs non-elective (including transfers) as defined in Appendix 4. The total duration of critical care during the 6-months after randomisation is derived from the dates of admission to and discharge from critical care in ICNARC, SUSCCDS, PEDWCCDS and SICSAG. If a period of critical care exists in any of these datasets it will contribute days to this outcome.

#### 8.2 Collection of 6-month outcomes outside the UK

Sites will complete a case report form at 6 months after randomisation to capture information on vital status, use of ventilation and any admissions to hospital.

### 9 Appendix 1: OPCS-4 and ICD-10 codes used to identify assisted ventilation and other outcomes in the linked hospitalisation data

| Outcome | code | Code type | Description |
|---|---|---|---|
| Use of CPAP | E85.6 | OPCS | Continuous positive airway pressure |
| Use of NIV | E85.2 | OPCS | Non-invasive ventilation NEC |
| Use IMV | E85.1 | OPCS | Invasive ventilation |
| Use of ECMO | X58.1 | OPCS | Extracorporeal membrane oxygenation |
| Use of RRT | X40.1 | OPCS | Renal dialysis |
| | X40.3 | OPCS | Haemodialysis NEC |
| | X40.4 | OPCS | Haemofiltration |

(OPCS and ICD-10 codes used to identify serious arrhythmia and other non-fatal outcomes to be added at a later date.)

### 10 Appendix: 2: Rules for determining start/end of advanced respiratory support days in the critical care datasets

Information is available in ICNARC/CCDS on

- The start and end date of the critical care episode
- The level of care at admission to the unit
- The level of care at discharge from the unit
- The reason for discharge from the unit
- The number of days of Advance Respiratory Support (ARS) received during the episode

The table below defines the rules for deciding whether the days on ARS in an ICNARC/CCDS episode should count from admission onwards (A), before discharge (D) or at the midpoint between admission and discharge (M)

| | | Level of care at admission to the unit | | | | |
|---|---|---|---|---|---|---|
| | | 0 | 1 | 2 | 3 | blank |
| Level of care at discharge from the unit | 0 | M | M | M | A | A |
| | 1 | M | M | M | A | A |
| | 2 | M | M | M | A | A |
| | 3 | D | D | D | A | D |
| | blank | * | * | * | A | A |

* If the reason for discharge from the unit is 'comparable critical care' or 'more-specialist critical care' then D, otherwise M.

The following definitions are taken from the ICNARC data collection manual Version 3.1 (29 June 2009).

Level 3 – indicated by one or more of the following:

- admissions receiving advanced respiratory monitoring and support due to an acute illness
- admissions receiving monitoring and support for two or more organ system dysfunctions (excluding gastrointestinal support) due to an acute illness
- admissions solely receiving basic respiratory monitoring and support and basic cardiovascular monitoring and support due to an acute illness only meet Level 2

Level 2 – indicated by one or more of the following:

- admissions receiving monitoring and support for one organ system dysfunction (excluding gastrointestinal support) due to an acute illness
- admissions solely receiving advanced respiratory monitoring and support due to an acute illness meet Level 3
- admissions solely receiving basic respiratory and basic cardiovascular monitoring and support due to an acute illness meet Level 2
- admissions receiving pre-surgical optimisation including invasive monitoring and treatment to improve organ system function
- admissions receiving extended post-surgical care either because of the procedure and/or the condition of the admission
- admissions stepping down to Level 2 from Level 3 care

Level 1 – indicated by one or more of the following:

- admission recently discharged from a higher level of care
- admissions receiving a greater degree of observation, monitoring, intervention(s), clinical input or advice than Level 0 care
- admissions receiving critical care outreach service support fulfilling the medium-score group, or higher, as defined by NICE Guidelines 50

Level 0 – indicated by the following:

- admissions in hospital and receiving normal ward care

### 11 Appendix 3: Definition of prior RRT for End Stage Renal Disease

A previously validated algorithm was adapted to identify people requiring dialysis for ESRD from the prior HES/SMR01/PEDW.

Individuals who met the criteria for Rules 2-4 during a hospital admission prior to the admission during which they were randomised were considered to have prior ESRD provided they did not meet the criteria for Rule 1 after meeting the other criteria.

**Rule 1: Kidney Transplantation**

Occurrence of any incident kidney transplant code (with no removal within 90 days), or a prevalent kidney transplant code with no removal having occurred prior to the record.

**Rule 2: Peritoneal maintenance dialysis**

Occurrence of any admission with a peritoneal dialysis code (without diagnosis of acute kidney injury).

**Rule 3: Definite maintenance dialysis**

Occurrence of a dialysis code in a patient who has had:

(a) a diagnostic code for ESRD any time prior to, or within 365 days; or

(b) the insertion of an AV fistula or graft any time prior to, or within 365 days.

**Rule 4: Probable maintenance dialysis**

The occurrence of at least two episodes containing a dialysis code, with at least 90 days between the start of the first recorded dialysis, and the start of any subsequent dialysis (without agnosis of acute kidney injury).

#### Relevant ICD-10 and OPCS-4 codes for Rules 1-4 above

| Group | Category | ICD-10 | OPCS-4 | Description |
|---|---|---|---|---|
| Diagnosis | Acute kidney injury | N17 | | Acute renal failure |
| Diagnosis | End-stage renal disease | N18.0 | | End-stage renal disease |
| Diagnosis | End-stage renal disease | N18.5 | | Chronic kidney disease, stage 5 |
| Diagnosis | End-stage renal disease | Q60.1 | | Renal agenesis, bilateral |
| Dialysis | Dialysis | E85.3 | | Secondary systemic amyloidosis (dialysis related) |
| Dialysis | Dialysis | Y60.2 | | Unintentional cut, puncture, perforation or haemorrhage during surgical and medical care; during kidney dialysis….. |
| Dialysis | Dialysis | Y61.2 | | Foreign object accidentally left in body during surgical and medical care; during kidney dialysis or other perfusion |
| Dialysis | Dialysis | Y62.2 | | Failure of sterile precautions during surgical and medical care; during kidney dialysis or other perfusion |
| Dialysis | Dialysis | Y84.1 | | Other medical procedures as the cause of abnormal reaction of the patient, or of later complication; kidney dialysis |
| Dialysis | Dialysis | Z99.2 | | Dependence on enabling machines and devices, not elsewhere classified; dependence on renal dialysis |
| Dialysis | Dialysis | | X40.1 | Renal dialysis |
| Dialysis | Haemodialysis | T82.4 | | Mechanical complication of vascular dialysis catheter |
| Dialysis | Haemodialysis | Z49.1 | | Care involving dialysis; extracorporeal dialysis |
| Dialysis | Haemodialysis | | X40.3 | Haemodialysis NEC |
| Dialysis | Haemodialysis | | X40.4 | Haemofiltration |
| Dialysis | Insertion of AVF or graft | | L74.1 | Insertion of arteriovenous prosthesis |
| Dialysis | Insertion of AVF or graft | | L74.2 | Creation of arteriovenous fistula NEC |
| Dialysis | Insertion of AVF or graft | | L74.6 | Creation of graft fistula for dialysis |
| Dialysis | Insertion of AVF or graft | | L74.8 | Other specified arteriovenous shunt |
| Dialysis | Insertion of AVF or graft | | L74.9 | Unspecified arteriovenous shunt |
| Dialysis | Insertion of PD catheter | | X41.1 | Insertion of ambulatory peritoneal dialysis catheter |
| Dialysis | Peritoneal dialysis | Z49.2 | | Care involving dialysis; other dialysis |
| Dialysis | Peritoneal dialysis | | X40.2 | Peritoneal dialysis NEC |
| Dialysis | Peritoneal dialysis | | X40.5 | Automated peritoneal dialysis |
| Dialysis | Peritoneal dialysis | | X40.6 | Continuous ambulatory peritoneal dialysis |
| Dialysis | Tunnelled line insertion | | L91.5 | Insertion of tunnelled venous catheter |
| Transplantation | Incident kidney transplant | | M01.2 | Allotransplantation of kidney from live donor |
| Transplantation | Incident kidney transplant | | M01.3 | Allotransplantation of kidney from cadaver NEC |
| Transplantation | Incident kidney transplant | | M01.4 | Allotransplantation of kidney from cadaver heart beating |
| Transplantation | Incident kidney transplant | | M01.5 | Allotransplantation of kidney from cadaver heart non-beating |
| Transplantation | Incident kidney transplant | | M01.8 | Other specified transplantation of kidne |
| Transplantation | Incident kidney transplant | | M01.9 | Unspecified transplantation of kidney |
| Transplantation | Prevalent kidney transplant | N16.5 | | Renal tubulo-interstitial disorders in transplant rejection |
| Transplantation | Prevalent kidney transplant | T86.1 | | Kidney transplant failure and rejection |
| Transplantation | Prevalent kidney transplant | Z94.0 | | Kidney transplant status |
| Transplantation | Prevalent kidney transplant | | M08.4 | Exploration of transplanted kidney |
| Transplantation | Prevalent kidney transplant | | M17.4 | Post-transplantation of kidney examination - recipient |
| Transplantation | Prevalent kidney transplant | | M17.8 | Other specified interventions associated with transplantation of kidney |
| Transplantation | Prevalent kidney transplant | | M17.9 | Unspecified interventions associated with transplantation of kidney |
| Transplantation | Removal of kidney transplant | | M02.6 | Excision of rejected transplanted kidney |

### 12 Appendix 4: Definitions of discharge and of elective/planned admissions

***Definition of discharge used for the time to discharge outcome (see section 6.3)***

| Dataset | Criteria | Definition |
|---|---|---|
| PEDW | Discharge method not died or tranfer | Discharge method not 4 or 8, and<br>Discharge destination not 49, 51, 52, 53, 55, 56, 57, 79, 87, 98 |
| | No other admission up to 4 days before or 1 day after discharge which suggests transfer | Admission source 51 or 87, or<br>Admission method 2B, 81 or 28 |
| HES/SUS | Discharge not died or tranfer | Discharge method not 4 or 8, and<br>Discharge destination not 49, 50, 51, 52, 53, 79, 87 or 98 |
| | No other admission up to 4 days before or 1 day after discharge which suggests transfer | Admission source 51 or 87, or<br>Admission method 2B, 81 or 28 |
| SMR01 | Discharge not died or tranfer | Discharge type not 40-43, and<br>Discharge type is 10, 11, 18, 19, 70, 20-23, 28, 29 |
| | No other admission up to 4 days before or 1 day after discharge which suggests transfer | Admission type 18, 30, 36, 38, 39, 40 |

***Definition of planned / elective admissions used for the 6-months outcomes (see section 8.1.1)***

| Dataset | Admission type | Definitions |
|---|---|---|
| PEDW | Planned | If admission method NOT (21 or 22 or 23 or 24 or 25 or 27 or 28 or 81) |
| HES/SUS | Planned | IF admission method NOT (21 or 22 or 23 or 24 or 25 or 28 or 81 or 2A or 2B or 2C or 2D) |
| SMR01 | Planned | IF admission type NOT (18 or 20 or 21 or 22 or 30 or 31 or 32 or 33 or 34 or 35 or 36 or 38 or 39) |

### 13 Appendix 5: Example hospital recorded diagnoses showing extraction of start and end dates

Table: Four example HESAPC spells each containing three episodes

| | Example 1 | Example 2 | Example 3 | Example 4 |
|---|---|---|---|---|
| Episode 1<br>Episode start date 01/02/2021<br>Episode end date 02/02/2021 | R07.4 Chest pain unspecified | I219 Acute myocardial infarction, unspecified | J18.0 Bronchopneumonia unspecified | N17.9 Acute renal failure unspecified |
| Episode 2<br>Episode start date 02/02/2021<br>Episode end date 05/02/2021 | I21.4 Acute subendocardial myocardial infarction | I210 Acute transmural myocardial infarction of anterior wall | J15.9 Bacterial pneumonia unspecified | I26.0 Pulmonary embolism with mention of acute cor pulmonale |
| Episode 3<br>Episode start date 05/02/2021<br>Episode end date 08/02/2021 | A04.7 Enterocolitis due to Clostridium difficile | I210 Acute transmural myocardial infarction of anterior wall | J15.2 Pneumonia due to staphylococcus | N17.9 Acute renal failure unspecified |

The hospital recorded diagnoses and relevant dates which would be extracted from these examples are as follows:

Example 1:

- R07.4 Start date 01/02/2021 End date 02/02/2021
- I21.4 Start date 02/02/2021 End date 05/02/2021
- A04.7 Start date 05/02/2021 End date 08/02/2021

Example 2:

- I219 Start date 01/02/2021 End date 08/02/2021

Example 3:

- J18.0 Start date 01/02/2021 End date 02/02/2021
- J15.9 Start date 02/02/2021 End date 08/02/2021

Example 4:

- N17.9 Start date 01/02/2021 End date 08/02/2021
- I26.0 Start date 02/02/2021 End date 05/02/2021
